# Genomic reconstruction of the SARS-CoV-2 epidemic in England

**DOI:** 10.1101/2021.05.22.21257633

**Authors:** Harald S. Vöhringer, Theo Sanderson, Matthew Sinnott, Nicola De Maio, Thuy Nguyen, Richard Goater, Frank Schwach, Ian Harrison, Joel Hellewell, Cristina Ariani, Sonia Gonçalves, David Jackson, Ian Johnston, Alexander W. Jung, Callum Saint, John Sillitoe, Maria Suciu, Nick Goldman, Jasmina Panovska-Griffiths, The Wellcome Sanger Institute Covid-19 Surveillance Team, The COVID-19 Genomics UK (COG-UK) Consortium, Ewan Birney, Erik Volz, Sebastian Funk, Dominic Kwiatkowski, Meera Chand, Inigo Martincorena, Jeffrey C. Barrett, Moritz Gerstung

**Affiliations:** European Molecular Biology Laboratory, European Bioinformatics Institute EMBL-EBI, Hinxton, UK; Joint Biosecurity Center JBC; Wellcome Sanger Institute, Hinxton, UK; The Francis Crick Institute, London, UK; Public Health England PHE; London School of Hygiene & Tropical Medicine, London, UK; Big Data Institute, University of Oxford, UK; Imperial College, London, UK; German Cancer Research Centre dkfz, Heidelberg, Germany

## Abstract

The evolution of the SARS-CoV-2 pandemic continuously produces new variants, which warrant timely epidemiological characterisation. Here we use the dense genomic surveillance generated by the COVID-19 Genomics UK Consortium to reconstruct the dynamics of 71 different lineages in each of 315 English local authorities between September 2020 and June 2021. This analysis reveals a series of sub-epidemics that peaked in the early autumn of 2020, followed by a jump in transmissibility of the B.1.1.7/Alpha lineage. Alpha grew when other lineages declined during the second national lockdown and regionally tiered restrictions between November and December 2020. A third more stringent national lockdown suppressed Alpha and eliminated nearly all other lineages in early 2021. However, a series of variants (mostly containing the spike E484K mutation) defied these trends and persisted at moderately increasing proportions. Accounting for sustained introductions, however, indicates that their transmissibility is unlikely to have exceeded that of Alpha. Finally, B.1.617.2/Delta was repeatedly introduced to England and grew rapidly in the early summer of 2021, constituting approximately 98% of sampled SARS-CoV-2 genomes on June 26.

## Main

The SARS-CoV-2 virus accumulates approximately 24 point mutations per year, or 0.3 per viral generation ^1–3^. Tracking mutations across successive virus generations enables researchers to follow transmission clusters, define distinct viral lineages and model their behaviour. Most of these mutations appear to be evolutionarily neutral, but as the SARS-CoV-2 epidemic swept around the world in the spring of 2020, it became apparent that the virus is continuing to adapt to its human host. An initial sign was the emergence and global spread of the spike protein variant D614G in the second quarter of 2020. Epidemiological analyses estimated that this mutation, which defines the B.1 lineage, confers a 20% transmissibility advantage over the original A lineage isolated in Wuhan, China ^4^.

A broad range of lineages have been defined since, which can be used to track SARS-CoV-2 transmission across the globe ^5,6^. For example, B.1.177/EU-1, emerged in Spain in early summer of 2020 and spread across Europe through travel ^7^. Subsequently, four variants of concern (VOC) have been identified by the WHO and other public health authorities: The B.1.351/Beta lineage was discovered in South Africa^8^, where it spread rapidly in late 2020. The B.1.1.7/Alpha lineage was first observed in Kent in September 2020 ^9^ from where it swept through the United Kingdom and large parts of the world due to a 50-60% ^10–13^ increase in transmissibility. P.1/Gamma originated in Brazil ^14,15^ and has spread throughout South America. Most recently, B.1.617.2/Delta was associated with a large surge of COVID-19 in India in April 2021 and subsequently around the world.

### Spatiotemporal genomic epidemiology of SARS-CoV-2 lineages in England

In the United Kingdom, by late June 2021 the COVID-19 Genomics UK Consortium (COG-UK) has sequenced close to 600,000 viral samples. These data have enabled detailed reconstruction of the dynamics of the first wave of the epidemic in the UK between February and August 2020 ^16^. Here, we leverage a subset of those data: genomic surveillance generated by the Wellcome Sanger Institute Covid-19 Surveillance Team as part of COG-UK, to characterise the growth rates and geographic spread of different SARS-CoV-2 lineages and reconstruct how newly emerging variants changed the course of the epidemic. We will discuss the key events of the reconstructed epidemic in chronological order.

Our data covers England between September 1, 2020 and June 26, 2021 encompassing three epidemic waves and two national lockdowns (**Figure 1a**). In this time period, we sequenced 281,178 viral genomes, corresponding to an average of 7.2% (281,178/3,894,234) of all positive tests from PCR testing for the wider population outside the National Health Service (Pillar 2), ranging from 5% in the winter of 2020 to 38% in the early summer of 2021, and filtered to remove cases associated with international travel (**Methods**; **Extended Data Figure 1a,b**). Overall a total of 328 SARS-CoV-2 lineages were identified using the PANGO lineage definition ^5^. As some of these lineages were only rarely and intermittently detected, we collapsed these based on the underlying phylogenetic tree into a set of 71 lineages such that each resulting lineage constituted at least 100 genomes, unless the lineage has been designated a VOC, variant under investigation (VUI) or variant in monitoring by Public Health England^17^ (**Figure 1b-d, Supplementary Table 1, 2**).

**Figure 1.**
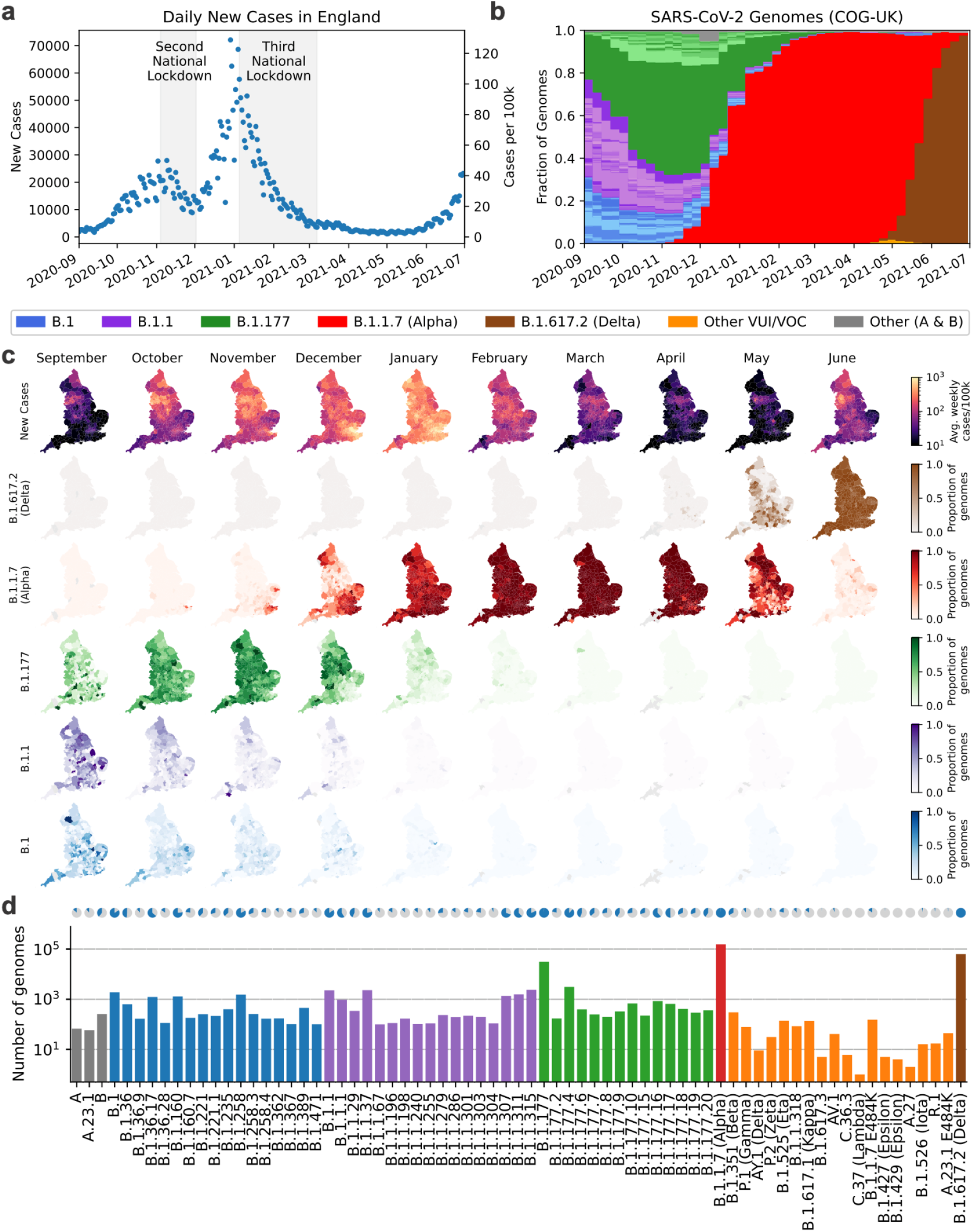
SARS-CoV-2 surveillance sequencing in England between September 2020 and June 2021. **a**. Positive Pillar 2 SARS-CoV-2 tests in England. **b**. Relative frequency of 328 different PANGO lineages, representing approximately 7.2% of tests shown in **a. c**. Positive tests (top row) and frequency of 4 major lineages across 315 English lower tier local authorities. **d**. Absolute frequency of sequenced genomes mapped to 71 PANGO lineages. Blue areas in the pie charts are proportional to the fraction of LTLAs where a given lineage was observed.

These data reveal a diversity of lineages in the fall of 2020 followed by sweeps of the Alpha and Delta variants (**Figure 1b, Supplementary Table 2, 3**). **Figure 1c** shows the geographic distribution of cases and of different lineages, studied at the level of 315 English Lower Tier Local Authorities (LTLAs), administrative regions with approximately 100,000-200,000 inhabitants.

### Modeling the dynamics of SARS-CoV-2 lineages

We developed a Bayesian statistical model that tracks the fraction of genomes from different lineages in each LTLA in each week and fits the daily total number of positive Pillar 2 tests (**Extended Data Figure 2**; **Methods**). The multivariate logistic regression model is conceptually similar to previous approaches in its estimation of relative growth rates^10,11^ and accounts for differences in the epidemiological dynamics between LTLAs, and allows for the introduction of new lineages (**Figure 2a-c**). Despite the sampling noise in a given week, the fitted proportions recapitulate the observed proportions of genomes as revealed by 35 example LTLAs covering the geography of England (**Figure 2b,c**, **Supplementary Note 1,2**). The quality of fit is confirmed by different probabilistic model selection criteria (**Extended Data Figure 3**) and also evident at the aggregated regional level (**Extended Data Figure 4**).

**Figure 2.**
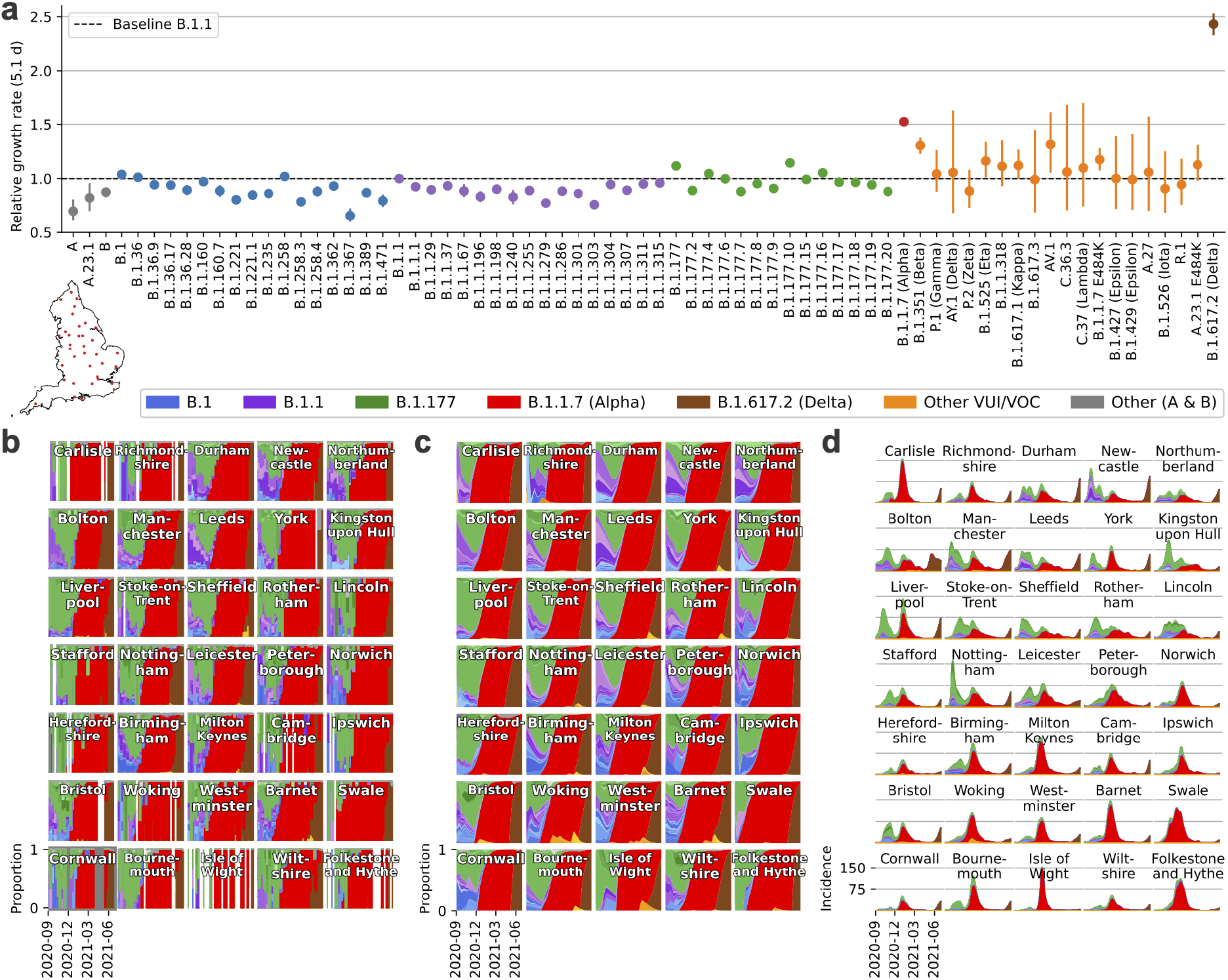
Spatiotemporal model of 71 SARS-CoV-2 lineages in 315 English LTLAs between September 2020 and June 2021. **a**. Average growth rates for 71 lineages. **b**. Lineage specific relative frequency for 35 selected LTLAs, arranged by longitude and latitude to geographically cover England. **c**. Fitted lineage-specific relative frequency for the same LTLAs as **b. d**. Fitted lineage-specific incidence for the same LTLAs as in **b**.

While the relative growth rate of each lineage is identical, the fitted patterns of viral proportions in each LTLA are dynamic due to the timing and rate of introduction of each lineage. The model also calculates total and lineage-specific local incidences and time-dependent *R*_*t*_ values by negative binomial spline fitting of the number of daily positive PCR tests (**Methods; Figure 2d**; **Extended Data Figure 2c**). Together, this enables a quantitative reconstruction of different periods of the epidemic.

### Multiple sub-epidemics and expansion of B.1.177 in the autumn of 2020

The autumn of 2020 was characterised by a surge of cases, concentrated in the north of England, which peaked in November 2020 triggering a second national lockdown (**Figure 1a,c**). This second wave initially featured B.1 and B.1.1 sublineages, which were slightly more prevalent in the south and north of England, respectively (**Figure 2b,c**). Yet the proportion of B.1.177 and its geographically diverse sublineages steadily increased across LTLAs from around 25% at the beginning of September to 65% at the end of October. This corresponds to a growth rate between 8% (growth per 5.1d; 95% CI: 7-9) and 12% (95% CI: 11-13) greater than that of B.1 or B.1.1. The trend of B.1.177 expansion relative to B.1 persisted throughout January (**Extended Data Figure 5a**) and involved a number of monophyletic sublineages that arose in the UK and similar patterns in Denmark^18^ (**Extended Data Figure 5b**). Such behavior cannot easily be explained by international travel, which was the major factor in B.1.177’s initial spread throughout Europe in the summer of 2020 ^7^. However, the biological explanation for this growth advantage is unclear as the characteristic A222V spike variant is not believed to confer a growth advantage ^7^.

### Alpha-specific growth during restrictions from November 2020 to March 2021

The subsequent third wave from December 2020 to February 2021 was almost exclusively driven by Alpha/B.1.1.7 as described previously ^10,11,19^. The rapid sweep of Alpha was due to an estimated transmissibility advantage of 1.52 compared to B.1.1 (growth per 5.1d; 95% CI 1.50-1.55; **Figure 2a**), assuming an unchanged generation interval distribution ^20^. The growth advantage is thought to stem from spike mutations facilitating ACE2 receptor binding (N501Y)^21,22^ and furin cleavage (P681H)^23^. Alpha grew during a period of restrictions, which were insufficient to contain a much more transmissible variant (**Figure 3a**).

**Figure 3.**
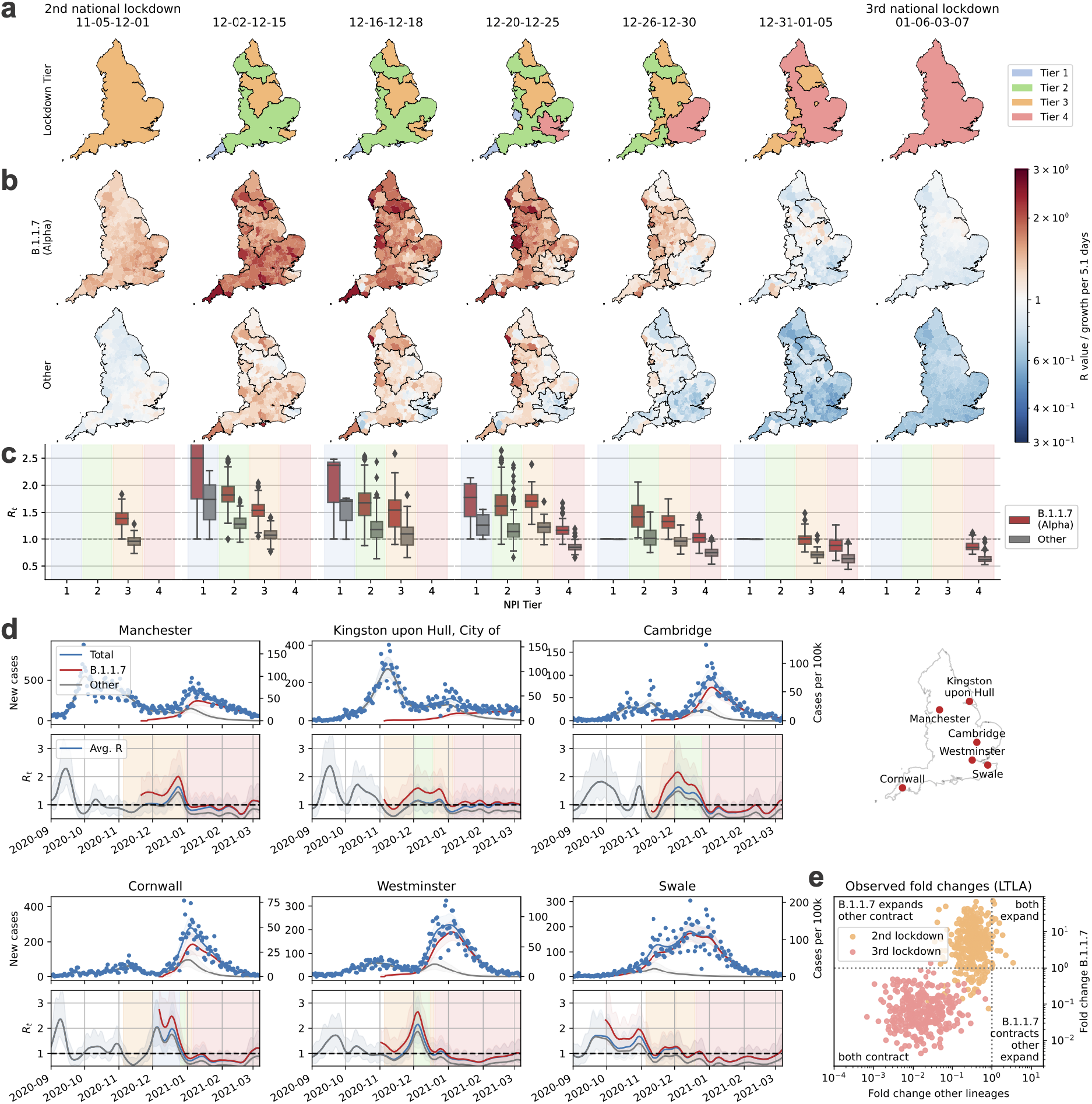
Growth of B.1.1.7/Alpha and other lineages in relation to lockdown restrictions between November 2020 and March 2021. **a**. Maps and dates of national and regional restrictions in England. Second national lockdown: closed hospitality businesses, contacts ≤ 2 outdoors only, open schools, reasonable excuse needed for leaving home^64^. Tier 1: private indoor gatherings ≤ 6 persons. Tier 2: as tier 1, restricted hospitality services, gatherings ≤ 6 in public outdoor places. Tier 3: as tier 2, most hospitality businesses closed. Tier 4: as tier 3, single outdoor contact. Third national lockdown: Closed schools with the exception of key workers. **b**. Local lineage-specific *R*_*t*_ values for Alpha and the average *R*_*t*_ value (growth per 5.1d) of all other lineages in the same periods. **c**. Boxplots of *R*_*t*_ values shown in **b**, boxes show quartiles, whiskers extend to 1.5x the inter quartile range. **d**. Total and lineage-specific incidence (top) and *R*_*t*_ values (bottom) for 6 selected LTLAs during the period of restrictions. **e**. Crude lineage-specific fold changes (odds ratios) for Alpha and other lineages across the second (orange) and third national lockdown (red).

The second national lockdown from November 5 to December 1, 2021 successfully reduced total cases, but this masked a lineage-specific rise (*R*_*t*_ > 1; defined as growth per 5.1d) of Alpha and simultaneous decline of other hitherto dominant lineages (*R*_*t*_ < 1) in 78% (246/315) of LTLAs (**Figure 3b,c**)^24^. This pattern of Alpha-specific growth during lockdown is supported by a model-agnostic analysis of raw case numbers and proportions of Alpha genomes (**Figure 3e**).

Three levels of regionally-tiered restrictions were introduced in December 2020 ^25^ (**Figure 3a**). The areas under different tiers of restrictions visibly and quantitatively coincide with the resulting local *R*_*t*_ values, with greater *R*_*t*_ values in areas with lower restrictions (**Figure 3a-c**). The reopening caused a surge of cases across all tiers with *R*_*t*_ > 1, which is also evident in selected time series (**Figure 3d**). As Alpha cases surged, more areas were placed under tier 3 and a higher tier 4 was introduced. Nevertheless, Alpha continued to grow (*R*_*t*_ > 1) in most areas, presumably driven by increased social interaction over Christmas (**Figure 3c**).

Following the peak of 72,088 daily cases on December 29 (**Figure 1a**), a third national lockdown was announced on January 4 (**Figure 3a**). The lockdown and increasing immunity derived from infection and increasing vaccination^26^ led to a sustained contraction of the epidemic to approximately 5,500 daily cases by March 8, when restrictions began to be lifted by reopening schools (further steps of easing occurred on April 12 and May 17). In contrast to the second national lockdown 93% (296/315) of LTLAs exhibited a contraction of both Alpha and other lineages (**Figure 3e)**.

### Elimination of SARS-CoV-2 lineages from January to April 2021

The lineage-specific rates of decline during the third national lockdown and throughout March 2021 resulted in large differences in lineage-specific incidence (**Figure 4a**). Cases of Alpha contracted nationally from a peak of around 50,000 daily new cases to around 2,739 (CI: 2,666-2,806) on April 1. At the same time B.1.177, the most prevalent lineage in November 2020 fell to only about 6 (95% CI 4-10) detected cases per day. Moreover, the incidence of most other lineages present in the autumn of 2020 was well below 1 after April 2021, implying that the majority of them have been eliminated. The number of observed distinct PANGO lineages declined from a peak of 137 to only 22 in the first week of April 2021 (**Figure 4b**). While this may in part be attributed to how PANGO lineages were defined, we note that the period of contraction did not replenish the genetic diversity lost due to the selective sweep by Alpha (**Extended Data Figure 6**).

**Figure 4.**
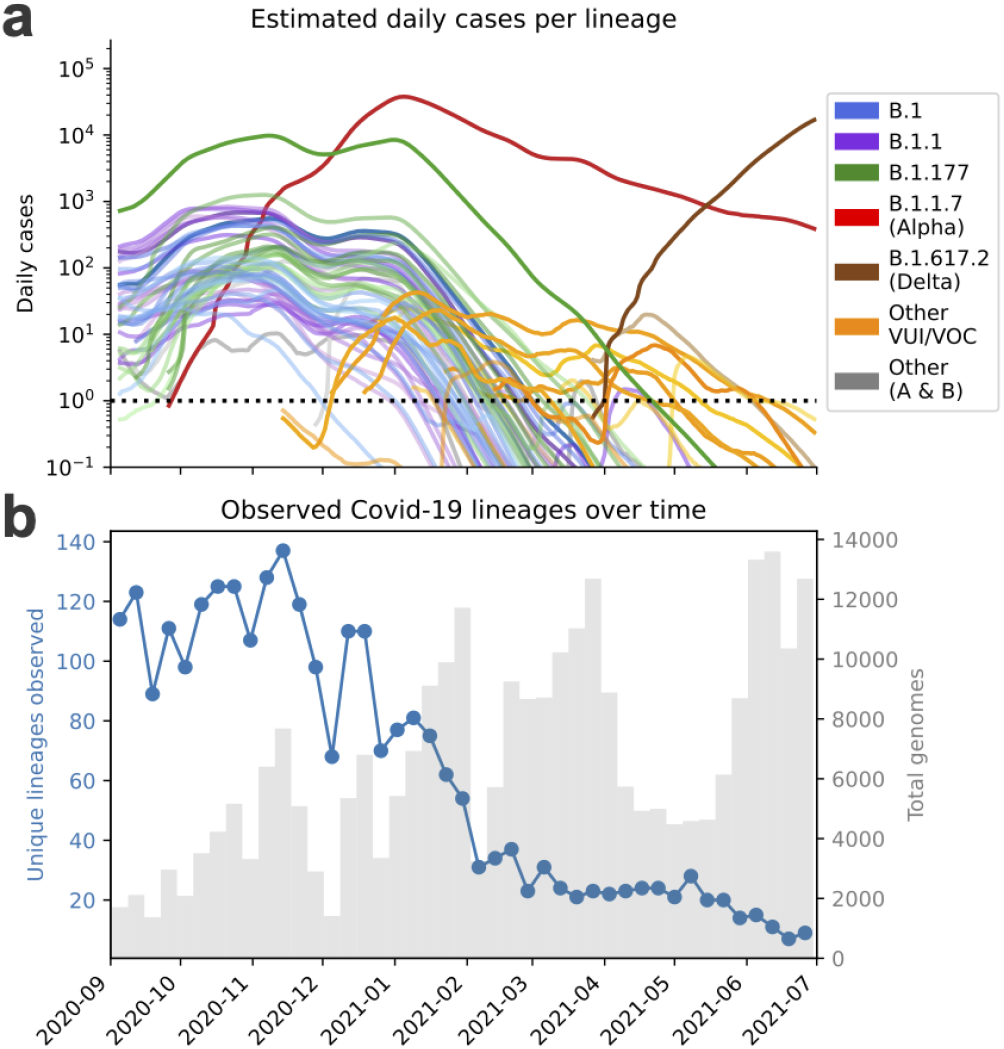
Elimination of SARS-CoV-2 lineages during spring 2021. **a** modelled lineage-specific incidence in England. Colors resemble major lineages as indicated and shadings thereof indicate sublineages. **b**. Observed number of PANGO lineages per week.

### Emergence of refractory variants with E484K mutations between December 2020 and May 2021

At the same time during which many formerly dominant SARS-CoV-2 lineages were eliminated, a number of new variants were imported or emerged (**Figure 4a**). These include the VOCs B.1.351/Beta, P.1/Gamma, which carry the spike variant N501Y also found in B.1.1.7/Alpha and a similar pair of mutations (K417N/T, E484K) each shown to reduce the binding affinity of antibodies from vaccine derived or convalescent sera ^21,27–30^. The ability to escape from prior immunity is consistent with the epidemiology of Beta in South Africa ^8^ and especially the surge of Gamma in Manaus ^15^. The VUIs B.1.525/Eta, B.1.1.318 and P.2/Zeta also harbour E484K spike mutations as per their lineage definition, and sublineages of Alpha and A.23.1 acquired E484K were found in England (**Figure 5a,b**).

**Figure 5.**
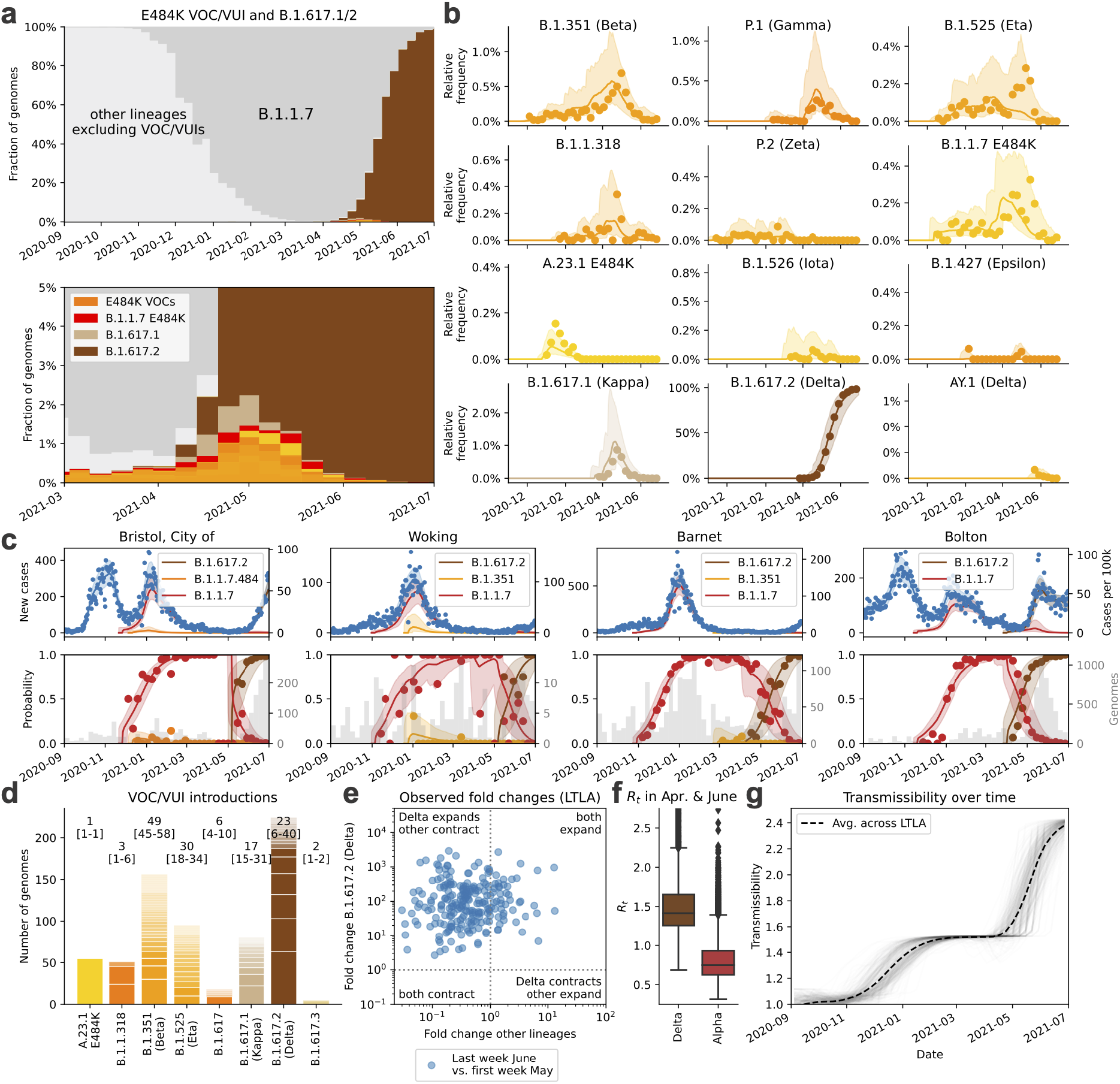
Dynamics of VOC and VUIs between January and June 2021. **a**. Observed relative frequency of other lineages (light grey), Alpha/B.1.1.7 (dark grey), VOC/VUIs (orange), and Delta/B.1.617.2 (brown). **b**. Observed and modelled relative frequency of VOC/VUIs in England. **c**. Total and relative lineage-specific incidence in four selected LTLAs. **d**. Estimated UK VOC/VUI clade numbers (numbers in square parentheses represent minimum and maximum numbers) and sizes. **e**. Crude growth rates (odds ratios) of Delta and Alpha between April and June 2021, as in **Fig. 3e. f**., Boxplots of lineage-specific *R*_*t*_ values in the same period, as in **Fig. 3d. g**. Changes of the average transmissibility across 315 LTLAs during the study period.

The proportion of these E484K containing VOCs and VUIs was consistently 0.3-0.4% from January to early April 2021. A transient rise especially of the Beta and Gamma variants was observed in May 2021 (**Figure 5a,b**). Yet the dynamics were largely stochastic and characterised by a series of individual and localised outbreaks, possibly curtailed by local surge testing efforts against Beta and Gamma variants (**Figure 5c**). Consistent with this transient nature of the outbreaks, the estimated growth rates of these variants were typically lower than Alpha (**Figure 2a**).

Sustained imports from international travel were a critical driving mechanism behind the observed number of non-Alpha cases. A detailed phylogeographic analysis establishing the most parsimonious sets of monophyletic and exclusively domestic clades, which can be interpreted as individual introductions, confirms that A.23.1 with E484K (1 clade) is likely to have been of domestic origin as no genomes of the same clade were observed internationally (**Figure 5d**; **Extended Data Figure 7; Methods**). The estimated number of introductions was lowest for B.1.1.318 (3 introductions, range 1-6), and highest for Beta (49; range 45-58) and Eta (30; range 18-34). While our data explicitly exclude genomes sampled directly from travellers, these repeated introductions make it clear that the true growth rate due to transmission is lower than the observed increase in the number of surveillance genomes.

### Rapid rise of Delta from April to June 2021

The B.1.617.1/Kappa and B.1.617.2/Delta lineages, first detected in India in 2020, began to appear in English surveillance samples in March 2021. Unlike other VOCs, Delta/Kappa do not contain N501Y or E484K mutations, but their L452R mutation may reduce antibody recognition^28^ and P681R enhances furin cleavage^31^, similar to Alpha’s P681H. The frequency of Delta, which harbours further spike mutations of unknown significance, increased rapidly and reached levels of 98% (12,474/12,689) on June 26, 2021 (**Figure 5a,b**). While initially constrained to a small number of large local clusters, such as in Bolton, in May 2021 (**Figure 5c**), Delta has been detected in all LTLAs by June 26 (**Figure 1c**). The sweep of Delta occurred at a rate around 59% (growth per 5.1d, CI 53-66) higher than Alpha with minor regional variation (**Figure 2a, Extended Data Figure 4e**).

The rapid rise of Delta contrasts to Kappa, which grew more slowly despite being introduced at a similar time and into a similar demographic background (**Figure 2a, Figure 5b**). This is also evident in the phylogeographic analysis (based on data as of May 1): Delta’s 224 genomes derive from larger clades (23 introductions, range 6-40; ∼10 genomes for every introduction) compared to the 80 genomes of Kappa (17 introductions, range 15-31, ∼3-4 genomes per introduction) and also other VOCs/VUIs (**Figure 5d; Extended Data Figure 8**). The AY.1 lineage, derived from Delta and containing an additional K417N mutation, appeared only transiently (**Figure 5b**).

Delta’s sustained domestic growth and international spread^32^ relative to the Alpha lineage are first evidence of a biological growth advantage. Causes appear to be a combination of increased transmissibility and immune evasion. Evidence for higher transmissibility are the high rates of spread in younger, unvaccinated age groups, reports of elevated secondary attack rates^17^ and a higher viral load^33^. Further, vaccine efficacy against infection by Delta is diminished, depending on the type of vaccine ^34,35^ and reinfection is more frequent^36^, both supported by experimental work demonstrating reduced antibody neutralisation of Delta by vaccine derived and convalescent sera ^37,38^.

The higher growth rate of Delta, combined with gradual reopening and proceeding vaccination, repeated the dichotomous pattern of lineage-specific decline and growth, however now with declining Alpha (*R*_*t*_ < 1) and growing Delta (*R*_*t*_ > 1; **Figure 5e,f**). Overall, we estimate that the spread of more transmissible variants between August 2020 and the early summer of 2021, increased the average growth rate of circulating SARS-CoV-2 in England by a factor of 2.39 (95% CI 2.25-2.42) (**Figure 5g**). Thus previously effective interventions may prove insufficient to contain newly emerging and more transmissible variants.

## Discussion

Here we reconstructed the SARS-CoV-2 epidemic in England from September 2020 to June 2021 in unprecedented genomic, spatial and temporal detail thanks to dense genomic surveillance data. Identifying lineages which consistently grew faster than others in each local authority – and thus at the same time, under the same restrictions and in a comparable population – pinpointed a series of variants with elevated transmissibility, in broad agreement with other reports ^10,11,13,15,32^. We note our precise growth rate estimates have a number of limitations. The growth rates of novel and thus rare variants is stochastic due to introductions and local outbreaks. Transmission depends both on the viral variant and the immunity of the host population, which changed from less than 20% to over 90% in the study period^39^. This will influence the growth rates of VOCs/VUIs with immune evasion capabilities over time. The effect of immunity is currently not modelled, but may become more important in the future as SARS-CoV-2 becomes endemic. Further technical considerations are discussed at the end of the **Methods** section.

The third and fourth waves in England were each caused by more transmissible variants, which outgrew restrictions sufficient to suppress previous variants. During the second national lockdown, Alpha grew despite falling numbers for other lineages and, similarly, Delta took hold in April and May when cases of Alpha were falling (**Figure 4a**). The fact that such growth was initially masked by the falling cases of dominant lineages highlights the need of dense genomic surveillance and rapid analysis in order to devise optimal and timely control strategies. Such surveillance should ideally be global, as even though Delta was associated with a large wave of cases in India, its transmissibility remained unclear at the time due to a lack of systematic genomic surveillance data.

The 2.4-fold increase in growth rate during the study period as a result of new variants is also likely to have consequences for the future course of the pandemic. If this increase in growth rate was explained solely by higher transmissibility it would raise the basic reproduction number *R*_*0*_ from a value of around 2.5-3 in the spring of 2020^40^ the range of 6-7 for Delta. This is likely to spur new waves of the epidemic in countries which have so far been able to control the epidemic despite low vaccination rates and may exacerbate the situation elsewhere. Even though the exact herd immunity threshold depends on contact patterns and the distribution of immunity across age groups ^41,42^, it is worth considering that Delta may increase the threshold to values around 0.85. Given current estimates of vaccine efficacy ^34,35,43^ this would require nearly 100% vaccination coverage. Even though more than 90% of adults had antibodies against SARS-CoV-2^39^ and close to 70% had received two doses of vaccination, England saw rising Delta variant cases in the first weeks of July 2021. It can thus be expected that other countries with high vaccination coverage are also likely to experience rising cases when restrictions are lifted.

SARS-CoV-2 is likely to continue its evolutionary adaptation process to humans^44^. Thus far variants with considerably higher transmissibility have had strongest positive selection, and swept through England during the 10 months of this investigation. But the possibility that an increasingly immune population may now select for variants with better immune escape highlights the need for continued systematic, and ideally global, genomic surveillance of the virus.

## Methods

### Pillar 2 SARS-CoV-2 testing data

Publicly available daily SARS-CoV-2 test result data from testing for the wider population outside the National Health Service (Pillar 2 newCasesBySpecimenDate) was downloaded from https://coronavirus.data.gov.uk/ spanning the date range from 2020-09-01 to 2021-06-30 for 315 English lower tier local authorities (downloaded on 2021-07-20). These data are mostly positive PCR tests, with about 4% of results from lateral flow tests without PCR confirmation. In this dataset, the City of London is merged with Hackney, and Isles of Scilly are merged with Cornwall due to their small number of inhabitants, thereby reducing the number of English LTLAs from 317 to 315. Population data for each LTLA was downloaded from the Office of National Statistics, https://www.ons.gov.uk/peoplepopulationandcommunity/populationandmigration/populationestimates/datasets/populationestimatesforukenglandandwalesscotlandandnorthernireland.

### SARS-CoV-2 surveillance sequencing

281,178 (Sep-June) were collected as part of random surveillance of positive tests of residents of England from four Pillar 2 Lighthouse Labs. The samples were collected between 2020-09-01 and 2021-06-26. A random selection of samples was taken, after excluding those known to be taken during quarantine of recent travellers, and samples from targeted and local surge testing efforts. The available metadata made this selection imperfect, but these samples should be an approximately random selection of infections in England during this time period, and the large sample size makes our subsequent inferences robust.

We amplified RNA extracts from these tests with C_t_ < 30 using the ARTIC amplicon protocol, https://www.protocols.io/workspaces/coguk/publications. We sequenced 384-sample pools on Illumina NovaSeq, and produced consensus fasta sequences according to the ARTIC nextflow processing pipeline https://github.com/connor-lab/ncov2019-artic-nf. Lineage assignments were made using Pangolin ^5^, according to the latest lineage definitions at the time, except for B.1.617, which we re-analysed after the designation of sub-lineages B.1.617.1, .2 and .3. Lineage prevalence was computed from 281,178 genome sequences. The genomes were mapped to the same 315 English LTLAs for testing data described above. Mapping was performed from outer postcodes to LTLA, which can introduce some misassignment to neighbouring LTLAs. Furthermore, lineages in each LTLA were aggregated to counts per week for a total of 43 weeks, defined beginning on Sunday and ending on Saturday.

Lastly, the complete set of 328 SARS-CoV-2 PANGO lineages was collapsed into lineages using the underlying phylogenetic tree, such that each resulting lineage constituted at least 100 genomes each during the study period with the exception of VOCs or selected VUIs, which were included regardless.

### Spatio-temporal genomic surveillance model

A hierarchical Bayesian model was used to fit local incidence data in a given day in each local authority and jointly estimate the relative historic prevalence and transmission parameters. In the following denotes time and is measured in days. We use the convention that bold uppercase variables such as ***B*** are matrix-variate, usually a combination of region and lineage. Bold lowercase variables ***b*** are vector-variate.

#### Motivation

Suppose that ***x′***(*t*) = (***b*** + *r*(*t*)) · ***x***(*t*) describes the ODE for the viral dynamics for a set of *l* different lineages. Here *r*(*t*) is a scalar time-dependent logarithmic growth rate thought to reflect lineage-independent transmission determinants, which changes over time in response to behavior, NPIs and immunity. This reflects a scenario where the lineages only differ in terms of the intensity of transmission, but not the inter generation time distribution. The ODE is solved by 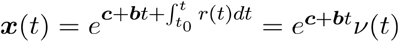. The term *ν*(*t*) contributes the same factor to each lineage and therefore drops from the relative proportions of lineages ***p*** (*t*) = ***x***(*t*)/ **Σ*x***(*t*) ∝ *e*^***c***+***b****t*^.

In the given model the lineage prevalence ***p***(*t*) follows a multinomial logistic-linear trajectory. Moreover the total incidence factorises into *μ*(*t*) = *ν*(*t*) Σ*e*^***c***+***b****t*^, which provides a basis to separately estimate the total incidence *μ*(*t*) from Pillar 2 test data and lineage-specific prevalence ***p***(*t*) from genomic surveillance data (which is taken from a varying proportion of positive tests). Exploiting the equations above one can subsequently calculate lineage-specific estimates by multiplying *μ*(*t*) with the respective genomic proportions ***p***(*t*).

#### Incidence

In the following we describe a flexible semi-parametric model of the incidence. Let ***μ*** (*t*) be the expected daily number of positive Pillar 2 tests and the population size in each of 315 LTLAs. Denote ***λ***(*t*) =log***μ***(*t*) − log***s*** the logarithmic daily incidence per capita at time in each of the 315 LTLAs.

Suppose *f ′* (*t*) is the daily growth rate of the epidemic, i.e., the number of new infections caused by the number of people infected at time. As new cases are only noticed and tested after a delay with distribution *u*, the resulting number of cases *f* * (*t*) will be given by the convolution

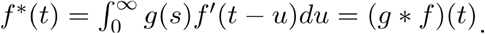

The time from infection to test is given by the incubation time plus the largely unknown distribution of the time from symptoms to test, which in England was required to take place within 5d of symptom onset. To account for these factors the log normal incubation time distribution from ^45^ is scaled by the equivalent of changing the mean by 2d. The convolution shifts cases approximately 6d into the future and also spreads them out according to the width of (**Extended Data Figure 2a**).

In order to parametrise the short and longer term changes of the logarithmic incidence ***λ***(*t*), we use a combination of weekly and *k* − *h* monthly cubic basis splines ***f***(*t*) = (***f***_1_(*t*), …, ***f***_*k*_(*t*)). The knots of the weekly splines uniformly tile the observation period except for the last 6 weeks.

Each spline basis function is convolved with the time to test distribution g,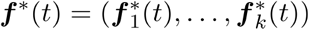 as outlined above and used to fit the logarithmic incidence. The derivatives of the original basis ***f*** ′(*t*) are used to calculate the underlying growth rates and *R*_*t*_ values, as shown further below. The convolved spline basis ***f****\**(*t*) is used to fit the per capita incidence in each LTLA as (**Extended Data Figure 2b**):

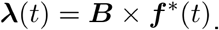

This implies that fitting the incidence function for each of the *m* local authorities is achieved by a suitable choice of coefficients ***B*** ∈ ℝ^*m*×*k*^, that is one coefficient for each spline function for each of the LTLAs. The parameters ***B*** have a univariate Normal prior distribution each, which reads for LTLA *i* and spline *j*:

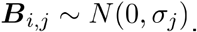

The standard deviation of the prior regularises the amplitude of the splines and is chosen σ_*j*_ = 0.2 as for weekly splines and σ_*j*_ = 1 for monthly splines. This choice was found to reduce the overall variance resulting from the high number of weekly splines, meant to capture rapid changes in growth rates, but which can lead to instabilities particularly at the end of the time series, when not all effects of changes in growth rates are observed yet. The less regularised monthly splines reflect trends on the scale of several weeks and are therefore subject to less noise.

Lastly, we introduce a term accounting for periodic differences in weekly testing patterns (there are typically 30% lower specimens taken on weekends; **Figure 1A**).

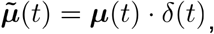

Where the scalar *δ*(*t*) = *δ*(t − *i* ×7) ∀*i* ∈ ℕ and prior distribution *δ*(*t*) ∼ LogNormal(0,1) for *t* = 1, …,6 and *δ*(0) = 1.

The total incidence was fitted to the observed number of positive daily tests ***X*** by a negative binomial with a dispersion *ω* = 10. The overdispersion buffers against non-Poissonian uncorrelated fluctuations in the number of daily tests.

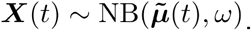

The equation above assumes that all elements of ***X***(*t*) are independent, conditional on 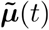.

#### Growth rates and *R*_*t*_ values

A convenient consequence of the spline basis of log ***μ*** = ***λ***, is that the delay-adjusted daily growth rate of the local epidemic, simplifies to:

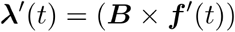

where 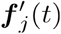 represents the first derivative of the *j* th cubic spline basis function.

In order to express the daily growth rate as an approximate reproductive number *R*_*t*_, one needs to consider the distribution of the inter generation time, which is assumed to be Gamma distributed with mean 6.3 days (*α* =2.29, *β* =0.36) ^45^. The *R*_*t*_ value can be expressed as a Laplace transform of the inter generation time distribution ^46^. Effectively, this shortens the relative time period because the exponential dynamics put disproportionally more weight on stochastically early transmissions over late ones. For reasons of simplicity and being mindful also of the uncertainties of the intergeneration time distribution, we approximate *R*_*t*_ values by multiplying the logarithmic growth rates with a value of 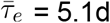, which was found to be a reasonable approximation to the convolution required to calculate *R*_*t*_ values (denoted here by the lower case symbol ***ρ***(*t*)in line with our convention for vector-variate symbols and to avoid confusion with the epidemiological growth rate *r*_*t*_),

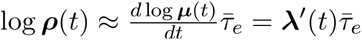

Hence the overall growth rate scaled to an effective inter generation time of 5.1d can be readily derived from the derivatives of the spline basis and the corresponding coefficients. The values derived from the approach are in very close agreement with those of the method of ^47^, but shifted according to the typical delay from infection to test (**Extended Data Figure 2b**).

#### Genomic prevalence

The dynamics of the relative frequency ***p***(*t*) of each lineage was modelled using a logistic-linear model in each LTLA, as motivated earlier. Define the logistic prevalence of each lineage in each LTLA as ***L***(*t*) = logit ***p***(*t*). This is modelled using the piecewise linear expression

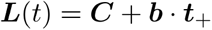

where *b* may be interpreted as a lineage specific growth advantage and ***C*** as an offset term of dimension (LTLA x lineages). Time ***t***_+_ is measured since introduction ***t***_0_ and defined as

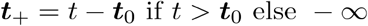

and accounts for the fact that lineages can be entirely absent prior to a stochastically distributed time period preceding their first observation. This is because in the absence of such a term the absence of a lineage prior to the point of observation can only be explained by higher growth rate compared to the preceding lineages, which may not necessarily be the case. As the exact time of introduction is generally unknown a stochastic three week period 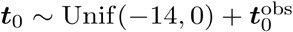 of prior to the first observation 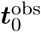 was chosen.

As the inverse logit transformation projects onto the *l* − 1 dimensional simplex *S*_*l*−1_ and thus loses one degree of freedom, B.1.177 was set as a baseline with

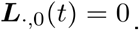

The offset parameters ***C*** are modelled across LTLAs as independently distributed multivariate Normal random variables with a lineage specific mean and covariance Σ = 10 · *I*_*l*−1_, where *I*_*l*−1_ denotes a (*l* − 1) × (*l* − 1) identity matrix. The lineage specific parameters growth rate ***b*** and average offset ***c*** are modelled using IID Normal prior distributions

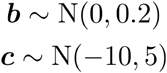

The time-dependent relative prevalence ***p***(*t*) of SARS-CoV2 lineages was fitted to the number of weekly genomes ***Y***(*t*) in each LTLA by a Dirichlet-Multinomial distribution with expectation 𝔼 **[*Y*(*t*)] ≈ *p*(*t*) · *G*(*t*)** where ***G*(*t*)** are the total number of genomes sequenced from each LTLA in each week. For LTLA *i* this is defined as:

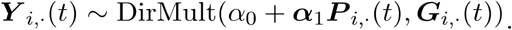

The scalar parameter *α*_0_ = 0.01 can be interpreted as a weak prior with expectation 1/*n*, which makes the model less sensitive to the introduction of single new lineages, which can otherwise exert a very strong effect. Further, the array ***α***_1_ = cases/2 increases the variance to account for the fact that, especially at high sequencing coverage (genomes ≈ cases), cases and thus genomes are likely to be correlated and overdispersed as they may derive from a single transmission event. Other choices such as *α*_1_ = 1000, which make the model converge to a standard Multinomial, leave the conclusions qualitatively unchanged. This model aspect is illustrated in **Extended Data Figure 2c**.

#### Lineage-specific incidence and growth rates

From the two definitions above it follows that the lineage specific incidence is given by multiplying the total incidence in each LTLA ***μ***(*t*) with the corresponding lineage frequency estimate ***P***(*t*) for lineage *j* at each time point

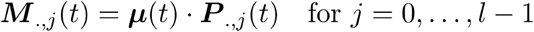

Further corresponding lineage-specific *R*_*t*_ values ***R***(*t*) in each LTLA can be calculated from the lineage agnostic average *R*_*t*_ value ***ρ***(*t*) and the lineage proportions ***P***(*t*) as

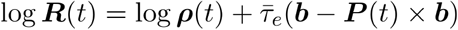

By adding the log growth rate fold changes ***b*** and subtracting the average log growth rate change ***P***(*t*) × ***b***. It follows that 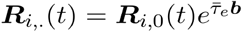, where ***R***_*i*,0_(*t*) is the *R*_*t*_ value of the reference lineage *j =* 0 (for which ***b***_0_ = 0) in LTLA *i*. It follows that all other lineage-specific the *R*_*t*_ values are proportional to this baseline at any given point in time with factor 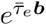.

#### Inference

The model was implemented in numpyro ^48,49^ and fitted using stochastic variational inference ^50^. Guide functions were multivariate normal distributions for each row (corresponding to an LTLA) of ***B, C*** to preserve the correlations across lineages and time as well as for (***b, c***) to also model correlations between growth rates and typical introduction.

### Phylogeographic analyses

To infer VOC introduction events into the UK and corresponding clade sizes, we investigated VOC genome sequences from GISAID https://www.gisaid.org/ available from any country. We downloaded multiple sequence alignments of genome sequences with release dates 17-04-2021 (for the analysis of lineages A.23.1, B.1.1.318, B.1.351, B.1.525) and 05-05-2021 (for the analysis of B.1.617 sublineages). We then extracted a sub-alignment from each lineage (following the 01-04-2021 version of PANGOlin for the 17-04-2021 alignment and the 23-04-2021 version of PANGOlin for the 05-05-2021 alignment), and, for each sub-alignment, we inferred a phylogeny via maximum likelihood using FastTree2 version 2.1.11 ^51^ with default options and GTR substitution model ^52^.

On each VOC/VUI phylogeny we inferred the minimum and maximum number of introductions of the considered SARS-CoV-2 lineage into the UK compatible with a parsimonious migration history of the ancestors of the considered samples; we also measure clade sizes for one specific example parsimonious migration history. We only count introduction events into the UK that result in at least one descendant from the set of UK samples that we consider in this work for our hierarchical Bayesian model; similarly, we measure clade sizes by the number of UK samples considered here included in such clades. Multiple occurrences of identical sequences were counted as separate cases, since this helps us identify rapid SARS-CoV-2 spread.

When using parsimony, we only consider migration histories along a phylogenetic tree that are parsimonious in terms of the number of migration events from and to the UK (in practice we collapse all the non-UK locations into a single one). Also, since SARS-CoV-2 phylogenies present substantial numbers of polytomies, that is, phylogenetic nodes where the tree topology cannot be reconstructed due to lack of mutation events on certain branches, we developed a tailored dynamic programming approach to efficiently integrate over all possible splits of polytomies and over all possible parsimonious migration histories. The idea of this method is somewhat similar to typical Bayesian phylogeographic inference (e.g. ^53^) in that it allows us to at least in part integrate over phylogenetic uncertainty and uncertainty in migration history; however, it also represents a very simplified version of these analyses, more so than ^16^, as it considers most of the phylogenetic tree as fixed, ignores sampling times, and uses parsimony instead of a likelihood-based approach. Parsimony is expected to represent a good approximation in the context of SARS-CoV-2, due to the shortness (both in time and substitutions) of the phylogenetic branches considered ^54,55^. The main advantage of our approach is that, thanks to the dynamic programming implementation, it is more computationally efficient than Bayesian alternatives, as the most computationally demanding step is the inference of the maximum likelihood phylogenetic tree. This allows us to infer plausible ranges for numbers of introduction events for large datasets and to quickly update our analyses as new sequences become available. The other advantage of this approach is that it allows us to easily customize the analysis and to focus on inferred UK introductions that result in at least one UK surveillance sample, while still making use of non-surveillance UK samples to inform the inferred phylogenetic tree and migration history. Note that possible biases due to uneven sequencing rates across the world (e.g. ^54^) apply to our approach as well as other popular phylogeographic methods.

Our approach works by traversing the maximum likelihood tree starting from the terminal nodes and ending at the root (postorder traversal). Here, we define a “UK clade” as a maximal subtree of the total phylogeny for which all terminal nodes are from the UK, all internal nodes are inferred to be from the UK, and at least one terminal node is a UK surveillance sample; the size of a UK clade is defined as the number of UK surveillance samples in it. At each node, using values already calculated for all children nodes (possibly more than two children in the case of a multifurcation), we calculate the following quantities: i) the maximum and minimum number of possible descendant UK clades of the current node, over the space of possible parsimonious migration histories, and conditional on the current node being UK or non-UK; ii) the number of migration events compatible with a parsimonious migration history in the subtree below the current node, and conditional on the current node being UK or non-UK; iii) the size so far of the UK clade the current node is part of, conditional on it being UK; iv) A sample of UK clade sizes for the subtree below the node. To calculate these quantities, for each internal node, and conditional on each possible node state (UK or non-UK), we consider the possible scenarios of having 0 of 1 migration event between the internal node and its children nodes (migration histories with more than 1 migration event between the node and its children are surely not parsimonious in our analysis and can be ignored).

To confirm the results of our analyses based on parsimony, we have also used the new Bayesian phylogenetic approach Thorney BEAST^16^ (https://beast.community/thorney_beast) for VOCs for which it was computationally feasible, that is, excluding B.1.351. For each VOC, we used in Thorney BEAST the same topology inferred with FastTree2 as for our parsimony analysis; in addition, we used treetime^56^ 0.8.2 to estimate a timed tree and branch divergences for use in Thorney BEAST. We used a 2-state (“UK” and “non-UK”) mugration model^53^ of migration to infer introductions into the UK, but again, only counted, from the posterior sample trees, UK clades with at least one UK surveillance sample. We used a Skygrid^57^ tree coalescent prior with 6 time intervals. The comparison of parsimony and Bayesian estimates is shown in **Extended Data Figure 8d**.

#### ONS infection survey analysis

Data from the cross sectional infection survey was downloaded from https://www.ons.gov.uk/peoplepopulationandcommunity/healthandsocialcare/conditionsanddiseases/bulletins/coronaviruscovid19infectionsurveypilot/30april2021.

Comparison of ONS incidence estimates with hospitalisation, case and death rates was conducted by estimating infection trajectories separately from observed cases, hospitalisations and deaths ^58,59^, convolving them with estimated PCR detection curves ^60^, and dividing the resulting PCR prevalence estimates by the estimated prevalence from the ONS Community Infection Survey at the midpoints of the 2-week intervals over which prevalence was reported in the survey.

#### Limitations

A main limitation of the model is that the underlying transmission dynamics are deterministic and stochastic growth dynamics are only accounted for in terms of (uncorrelated) overdispersion. For that reason the estimated growth rates may not accurately reflect the viral transmissibility, especially a low prevalence. While the logistic growth assumption is a consistent estimator of the average transmission dynamics, individual outbreaks may deviate from these dynamics and therefore provide unreliable estimates. It is therefore important to assess whether consistent growth patterns in multiple independent areas are observed.

In its current form the model only accounts for a single introduction event per LTLA. While this problem is in part alleviated by the high spatial resolution, which spreads introductions across 315 LTLAs, it is important to investigate whether sustained introductions inflate the observed growth rates, as in the case of the Delta variant or other VOCs and VUIs. This can be achieved by a more detailed phylogeographic assessment and the assessment of monophyletic sublineages.

Furthermore there is no explicit transmission modelled from one LTLA to another. As each introduction is therefore modelled separately, this makes the model conservative in ascertaining elevated transmission as single observed cases across different LTLAs can be explained by their introduction.

The inferred growth rates also cannot identify a particular mechanism which may be caused by higher viral load, longer infectivity or greater susceptibility. Lineages could potentially differ by their inter-generation time, which would lead to a non linear scaling. Here we did not find convincing evidence in incidence data for such effects. in contrast to previous reports ^24^. However, contact tracing data indicates that the inter-generation time may be shortening for more transmissible lineages such as Delta^33,61^.

Also lineages, such as Beta, Gamma or Delta differ in their ability to evade prior immunity. As immunity changes over time, this might lead to a differential growth advantage over time. It is therefore advisable to assess whether a growth advantage is constant over periods in which immunity changes considerably.

A further limitation underlies the nature of lineage definition and assignment. The PANGO lineage definition^5^ assigns lineages to geographic clusters, which have by definition expanded, which can induce a certain survivor bias, often followed by winner’s curse. Another issue results from the fact that very recent variants may not be classified as a lineage despite having grown, which can inflate the growth rate of ancestral lineages over sublineages.

As the total incidence is modelled based on the total number of positive PCR tests it may be influenced by testing capacity with the total number of tests having approximately tripled between September 2020 and March 2021. This can potentially lead to a time trend in recorded cases and thus baseline *R*_*t*_ values if the access to testing changed, e.g. by too few available tests during high incidence, or changes to the eligibility to test with fewer symptoms intermittently. Generally, the observed incidence was in good agreement with representative cross-sectional estimates from the Office of National Statistics ^62,63^, except for a period of peak incidence from late December 2020 to January 2021 (**Extended Data Figure 1d**). Values after March 8, 2021 need to be interpreted with caution as pillar 2 PCR testing was supplemented by lateral flow devices, which increased the number of daily tests to more than 1.5 million.

The modelled curves are smoothed over intervals of approximately 7 days using cubic splines, creating a possibility that later time points influence the period of investigation and cause a certain waviness of the *R*_*t*_ value pattern. An alternative parameterization using piecewise linear basis functions per week (i.e., constant *R*_*t*_ values per week) leaves the overall conclusions and extracted parameters broadly unchanged.

## Data Availability

PCR test data are publicly available at https://coronavirus.data.gov.uk/. SARS-CoV-2 genome data and geolocations can be obtained under controlled access from https://www.cogconsortium.uk/data/. A filtered, privacy conserving version of the data set is publicly available at https://covid19.sanger.ac.uk/downloads. The data and a version of the analysis with fewer lineages can be interactively explored at https://covid19.sanger.ac.uk.

https://www.cogconsortium.uk/data/

https://covid19.sanger.ac.uk/downloads

https://covid19.sanger.ac.uk

## Code availability

Code for spatio-temporal modeling of different viral lineage is available at https://github.com/gerstung-lab/genomicsurveillance and as a PyPI package (genomicsurveillance). This phylogeographic model has been implemented in python scripts, and the code is available from https://github.com/NicolaDM/phylogeographySARS-CoV-2. Code for ONS infection survey analysis is available at https://github.com/jhellewell14/ons_severity_estimates.

## Acknowledgements

COG-UK is supported by funding from the Medical Research Council (MRC) part of UK Research & Innovation (UKRI), the National Institute of Health Research (NIHR) and Genome Research Limited, operating as the Wellcome Sanger Institute. We would like to thank our colleagues at EMBL-EBI, the Wellcome Sanger Institute and from COG-UK for stimulating discussions and helpful comments on this manuscript. HSV and MG are supported by a grant from the Department of Health and Social Care. AWJ, EB and MG are beneficiaries from grant NNF17OC0027594 from the Novo Nordisk Foundation. TS is supported by grant 210918/Z/18/Z, and JH and SF by grant 210758/Z/18/Z from the Wellcome Trust. HSV, NDM, AWJ, NG, EB and MG are supported by EMBL. We would like to thank Elias Allara (Cambridge) and Georgia Whitton (Sanger) for providing outer postcode to LTLA mappings, Rupert Beale for comments and John McCrone for setting up Thorney Beast analysis. We thank all the contributors who submitted genome sequences to GISAID. Acknowledgement tables for individual sequences are deposited at https://github.com/NicolaDM/phylogeographySARS-CoV-2.

## Ethical approval

This study was done as part of surveillance for COVID-19 under the auspices of Section 251 of the National Health Service Act 2006. It therefore did not require individual patient consent or ethical approval. The COVID-19 Genomics UK (COG-UK) study protocol was approved by the Public Health England Research Ethics Governance Group.

## Author contributions

HSV and MG developed the analysis code, which HSV implemented with input from AWJ. HSV created most Figures. MS analysed, annotated and aggregated viral genome data. NDM conducted phylogeographic analyses supervised by NG. TS, RG, MS, and HSV developed the interactive spatiotemporal viewer. TN, FS, IH, RA, CA, SG, DJ, IJ, CS, JS, TS, MS analysed genomic surveillance data under supervision of DK, MC, IM and JCB. JH and SF analysed ONS data and helped with epidemiological modeling and data interpretation. EV analysed growth rates and helped with data interpretation. EB and JPG supervised HSV and helped with data interpretation. JCB and MG supervised the analysis with advice from IM. MG, HSV, MS, NDM, TS, IM and JCB wrote the manuscript with input from all co-authors.

## Extended Data Figures

**Extended Data Figure 1, related to Figure 1.**
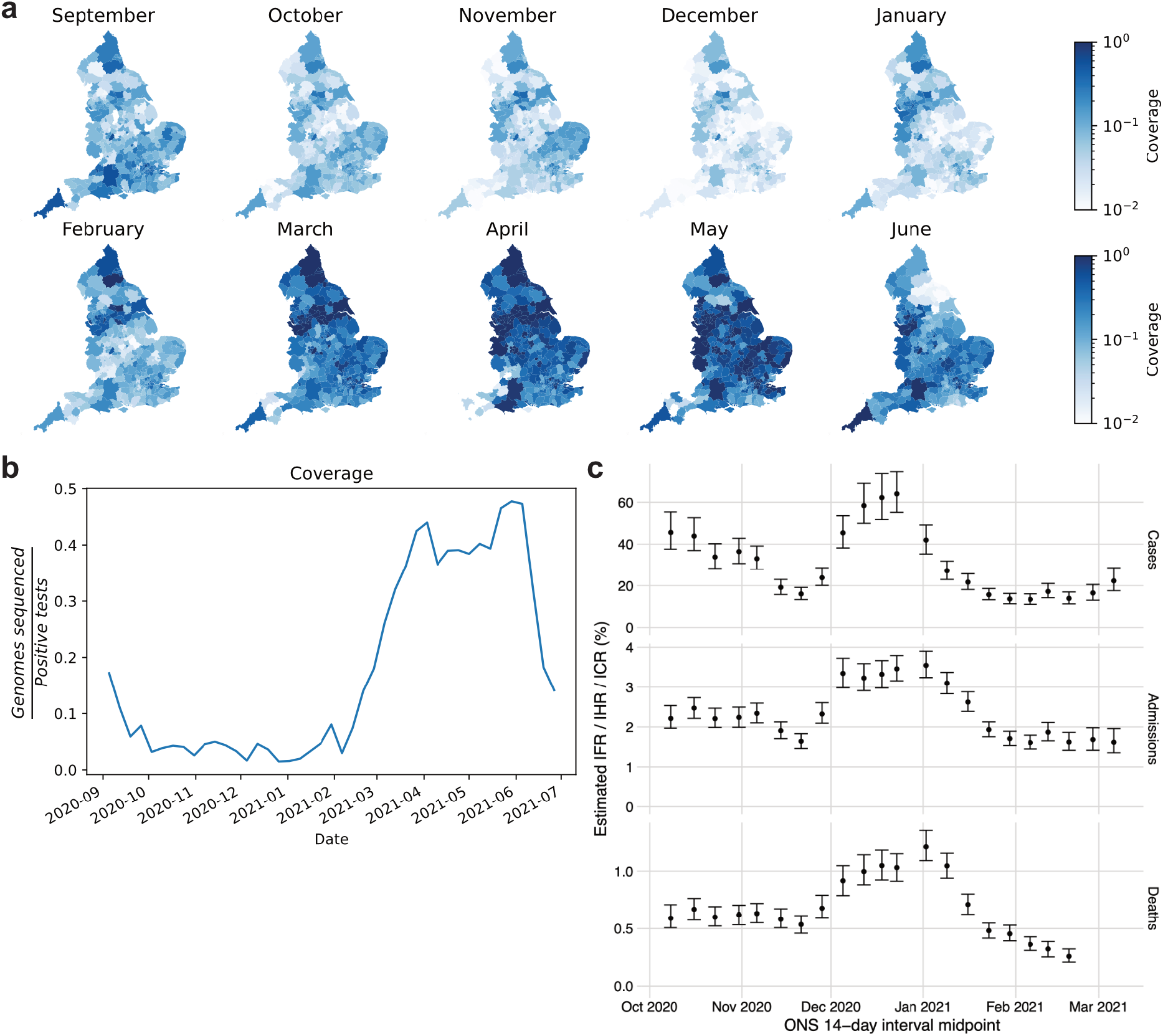
SARS-CoV-2 surveillance sequencing in England between September 2020 and June 2021. **a**. Local monthly coverage across 315 LTLAs. **b**. Weekly coverage of genomic surveillance sequencing. **c**. Hospitalisation, case and infection fatality rates relative to ONS prevalence.

**Extended Data Figure 2:**
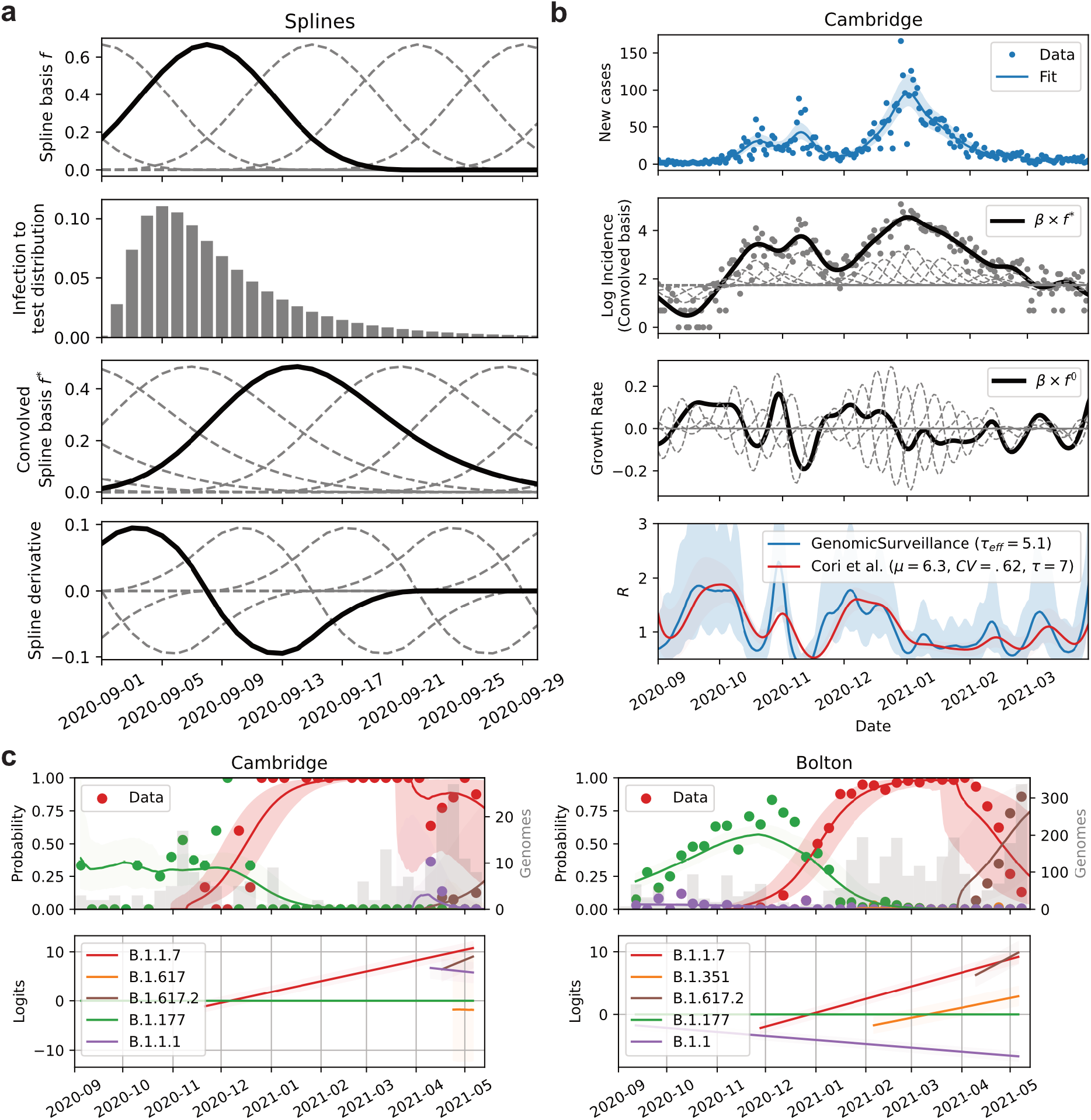
Genomic surveillance model of total incidence and lineage-specific frequencies. **a**. Cubic basis splines (top row) are convolved with the infection to test distribution (row 2 and 3) and used to fit the log incidence in a LTLA and its corresponding derivatives (growth rates; bottom row). **b**. Example incidence (top row), logarithmic incidence with individual convolved basis functions (dashed lines, row 2), growth rate with individual spline basis derivatives (dashed lines, row 3) and resulting (case) reproduction numbers (growth rate per 5.1d) from our approach (GenomicSurveillance) and estimates by EpiEstim ^47^, shifted by 10d to approximate a case reproduction number. **c**. The relative frequencies of 62 different lineages are modelled using piecewise multinomial logistic regression. The linear logits are modelled to jump stochastically within 21d prior to first observation to account for the effects of new introductions. Shown are the logits of 5 selected lineages in two different LTLAs.

**Extended Data Figure 3:**
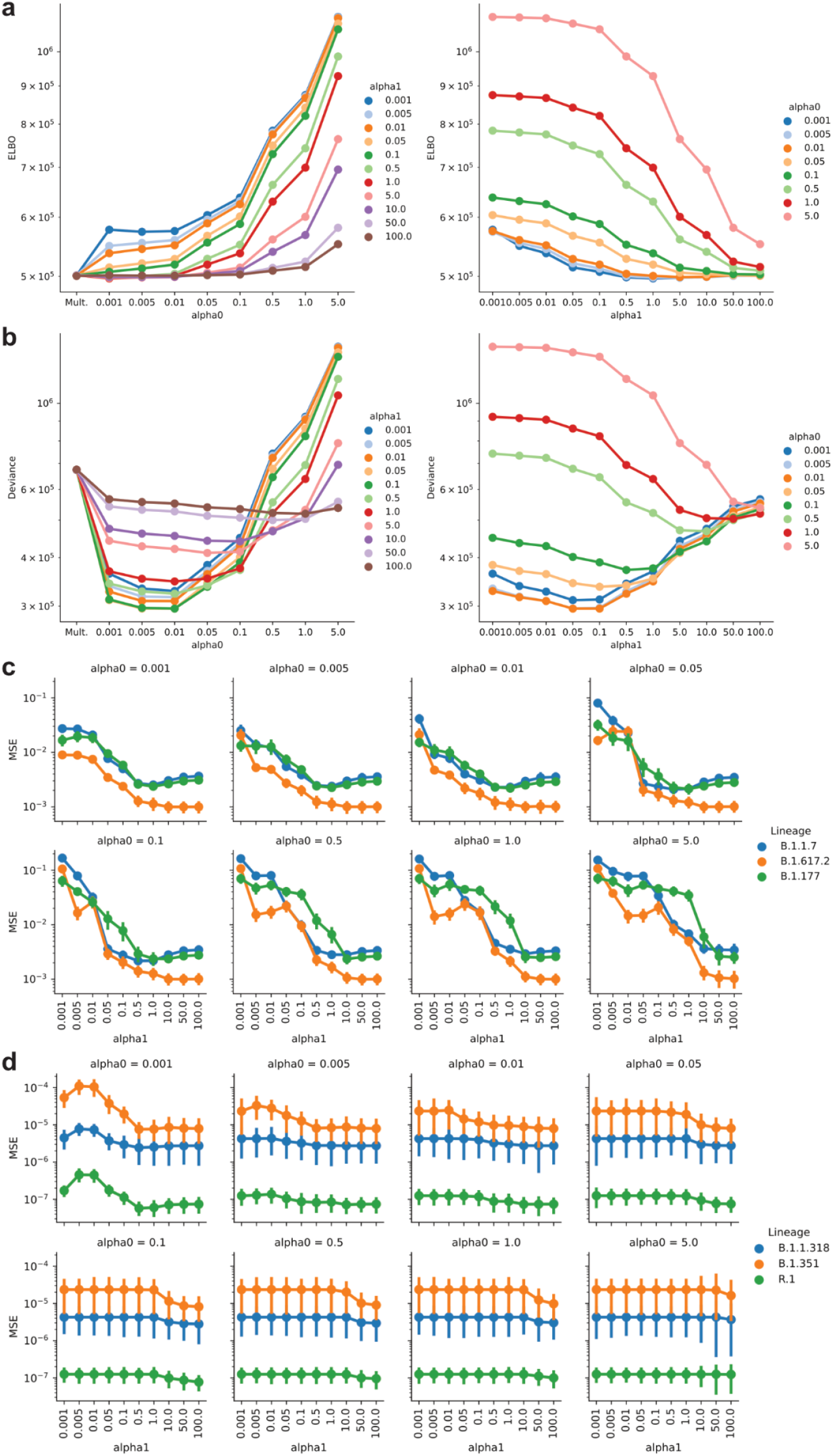
Genomic surveillance model selection. **a**. Model loss in terms of the ELBO objective function and the model hyperparameters alpha0 and alpha1 (see **Methods**). **b**. Model deviance (calculated as -2 x log pointwise predictive density) with respect to the model hyperparameters alpha0 and alpha1 (see **Methods**). **c**. Mean squared error (MSE) of modelled weekly proportions of highly prevalent lineages with respect to the model parameters alpha0 and alpha1 (see **Methods**). **d**. Same as in **c**, but for lineages exhibiting low frequencies (VOCs).

**Extended Data Figure 4.**
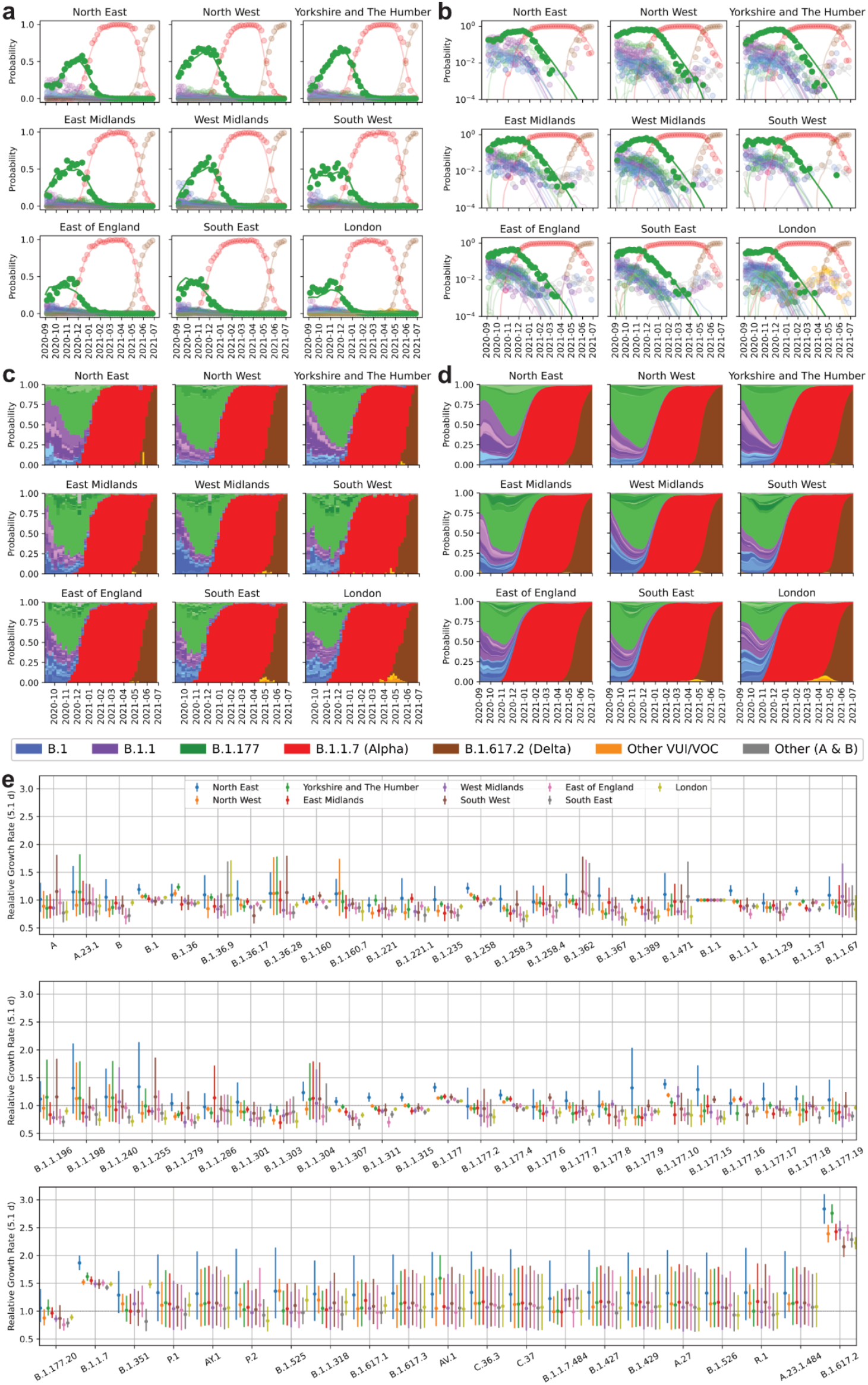
Spatiotemporal model of 71 SARS-CoV-2 lineages in 315 English LTLAs between September 2020 and June 2021. **a**. Regional lineage specific relative frequency of lineages contributing more than 50 genomes during the time period shown. Dots denote observed data, lines the fits aggregated to each region. **b**. Same as **a**, but on a log scale. **c**. Same data as in a, shown as stacked bar charts. Colors resemble major lineages as indicated and shadings thereof indicate sublineages. **d**. Same fits as in **a**, shown as stacked segments. **e**. Average growth rates for 71 SARS-Cov2 lineages estimated in different regions in England.

**Extended Data Figure 5.**
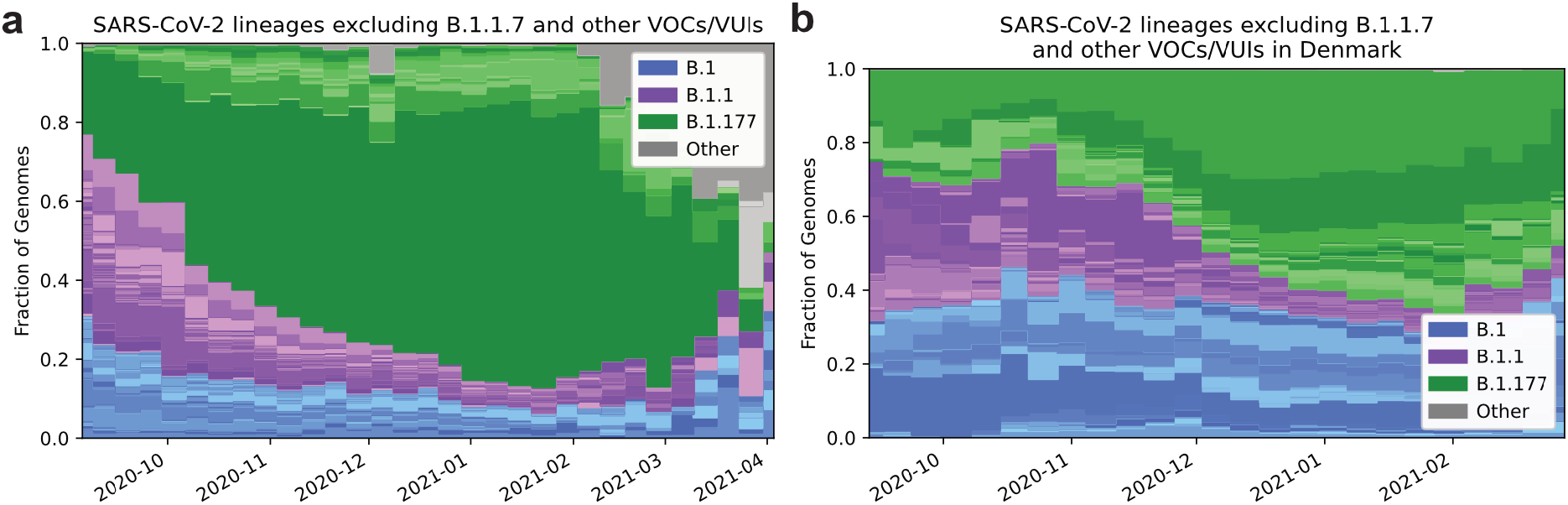
Relative growth of B.1.177. **a**. Lineage-specific relative frequency data in England, excluding B.1.1.7 and other VOCs/VUIs (Category Other includes: A, A.18, A.20, A.23, A.25, A.27, A.28, B, B.29, B.40, None). Colors resemble major lineages as indicated and shadings thereof indicate sublineages. **b**. Lineage-specific relative frequency data in Denmark, excluding B.1.1.7 and other VOCs/VUIs. Colors resemble major lineages as indicated and shadings thereof indicate sublineages.

**Extended Data Figure 6.**
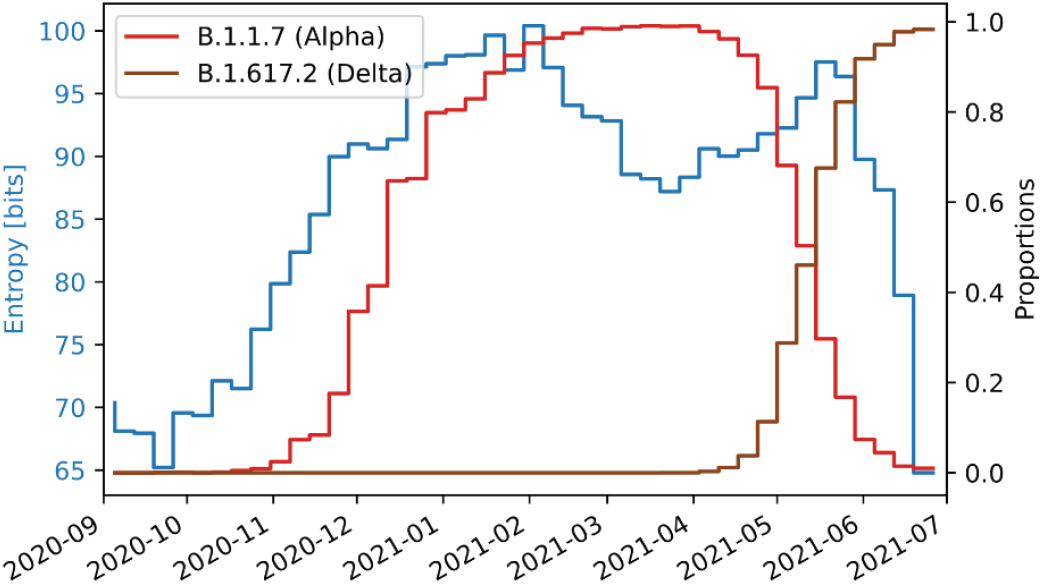
Genomic diversity of the SARS-CoV-2 epidemic. Shown is the entropy (blue), total number of observed Pango lineages (grey, divided by 4), as well as the proportion of B.1.1.7 (orange, right axis). The sweep of B.1.1.7 causes an intermittent decline of genomic diversity as measured by the entropy.

**Extended Data Figure 7.**
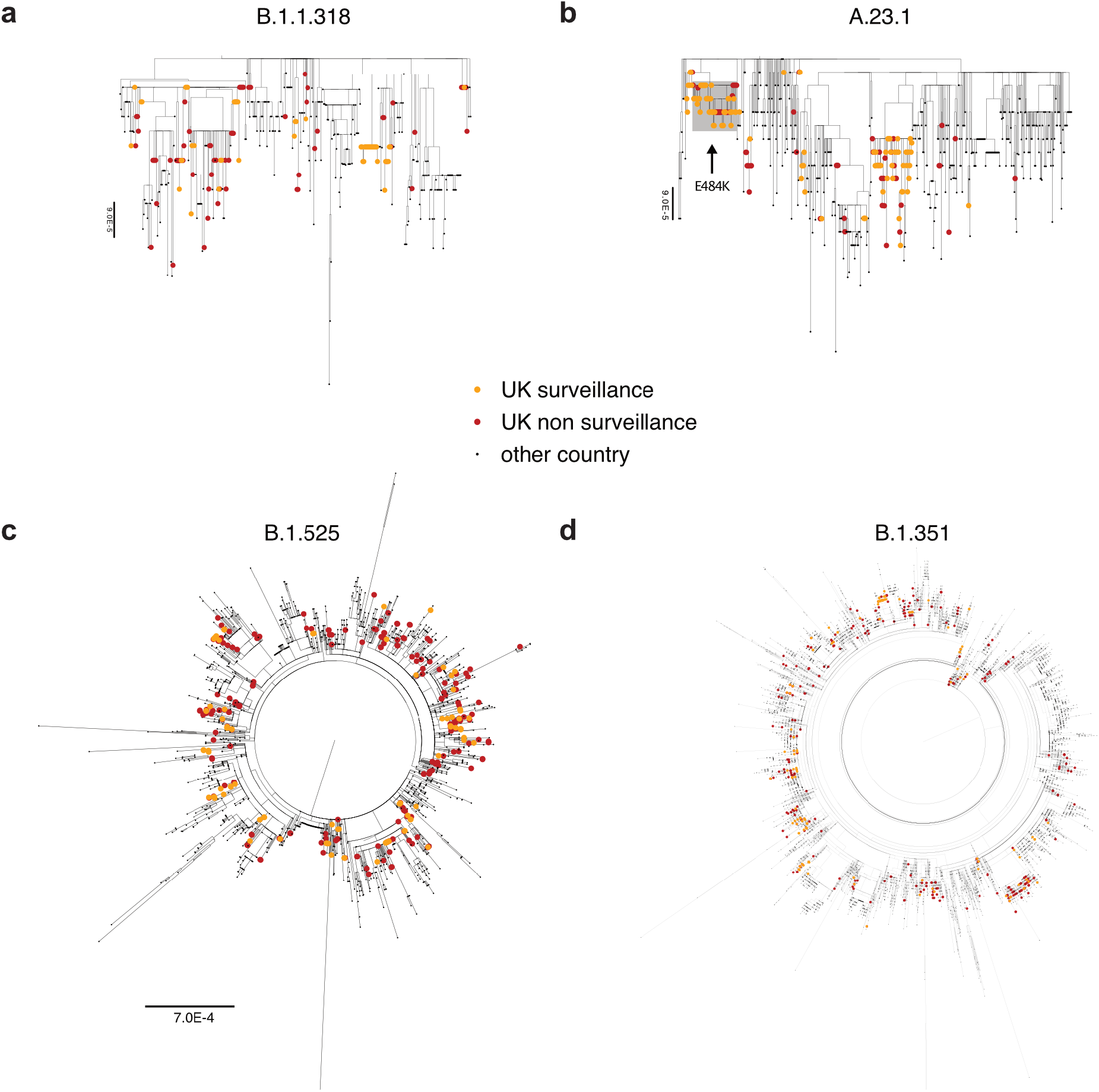
Global phylogenetic trees of selected VOCs/VUIs. English surveillance and other (targeted and quarantine) samples are highlighted respectively orange and red.

**Extended Data Figure 8.**
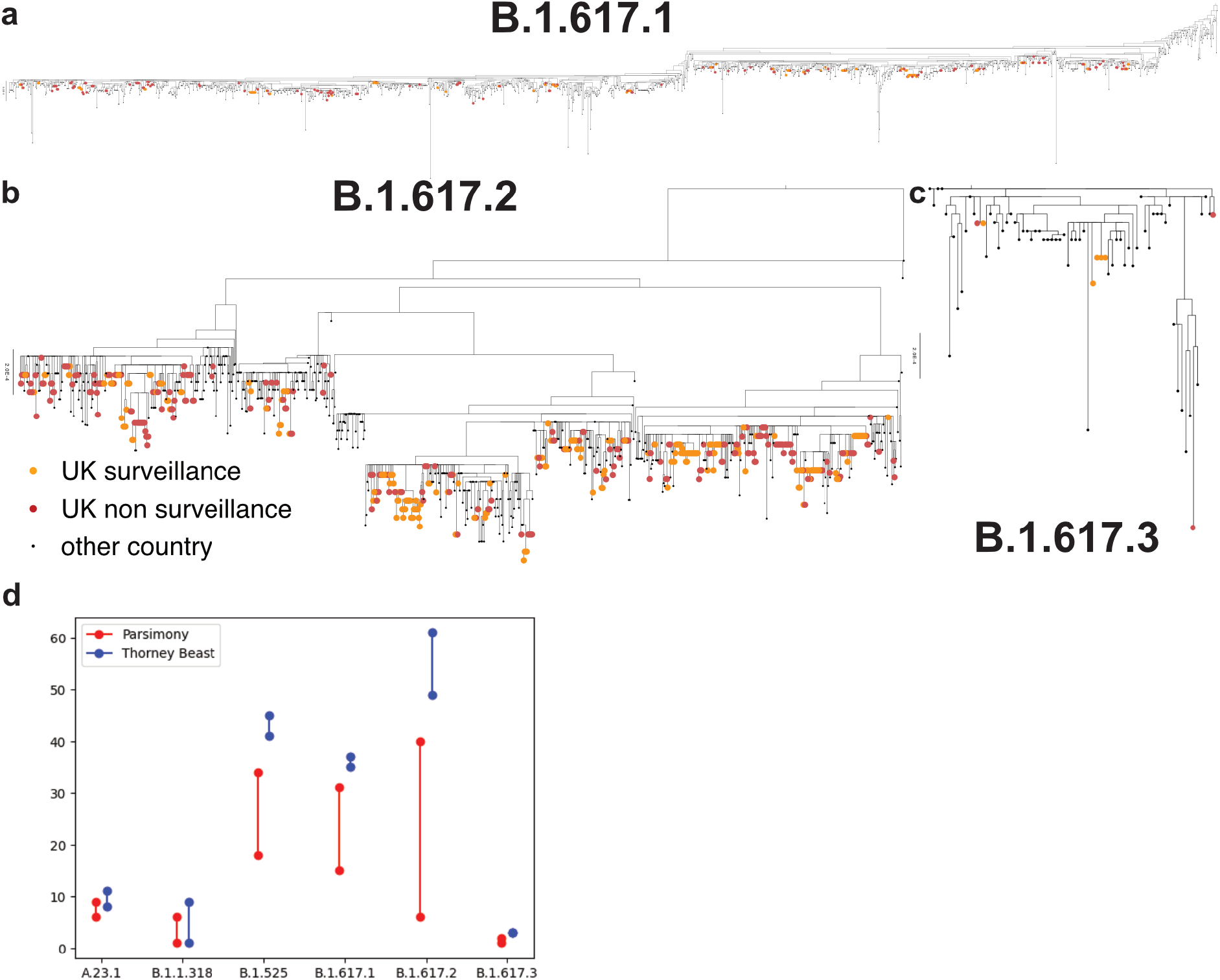
Global phylogenetic trees of B.1.617 sublineages. **a, b and c**. English surveillance and other (targeted and quarantine) samples are highlighted respectively orange and red. The trees of B.1.617.1 and B.1.617.2 are rooted. **d**. Number of UK introductions inferred by parsimony (minimum and maximum numbers) and by Thorney BEAST (95% posterior CI) for each VOC.

