## Supplementary Notes for "Genomic reconstruction of the SARS-CoV-2 epidemic in England"

**Abstract**

The evolution of the SARS-CoV-2 pandemic continuously produces new variants, which warrant timely epidemiological characterisation. Here we use the dense genomic surveillance generated by the COVID-19 Genomics UK Consortium to reconstruct the dynamics of 7162 different lineages in each of 315 English local authorities between September 2020 and June 2021. This analysis reveals a series of sub-epidemics that peaked in the early autumn of 2020, followed by a jump in transmissibility of the B.1.1.7/Alpha lineage. Alpha grew when other lineages declined during the second national lockdown and regionally tiered restrictions between November and December 2020. A third more stringent national lockdown eventually suppressed Alpha and eliminated nearly all other lineages in early 2021. However, a series of variants (mostly containing the spike E484K mutation) defied these trends and persisted at moderately increasing proportions. Accounting for sustained introductions, however, indicates that their transmissibility is unlikely to have exceeded that of Alpha. Finally, B.1.617.2/Delta was repeatedly introduced to England and grew rapidly in the early summer of 2021, constituting approximately 98% of sampled SARS-CoV-2 genomes on June 27.

### Contents

1 Supplementary Note 1 3

2 Supplementary Note 2 11

3 Supplementary Note 3 92

### 1 Supplementary Note 1

Supplementary Note 1 Fig. 1: **Observed proportions of sequenced SARS-Cov2 lineages in 315 LTLA in England.** The titles in each tile refer to LTLA look up codes (Supplementary Note 3) and colours indicate major SARS-CoV-2 lineages (blue: B.1, purple: B.1.1, green: B.1.177, red: B.1.1.7 (Alpha), orange: other VOC/VUI, brown: B.1.617.2 (Delta) and grey: Other (A & B)).

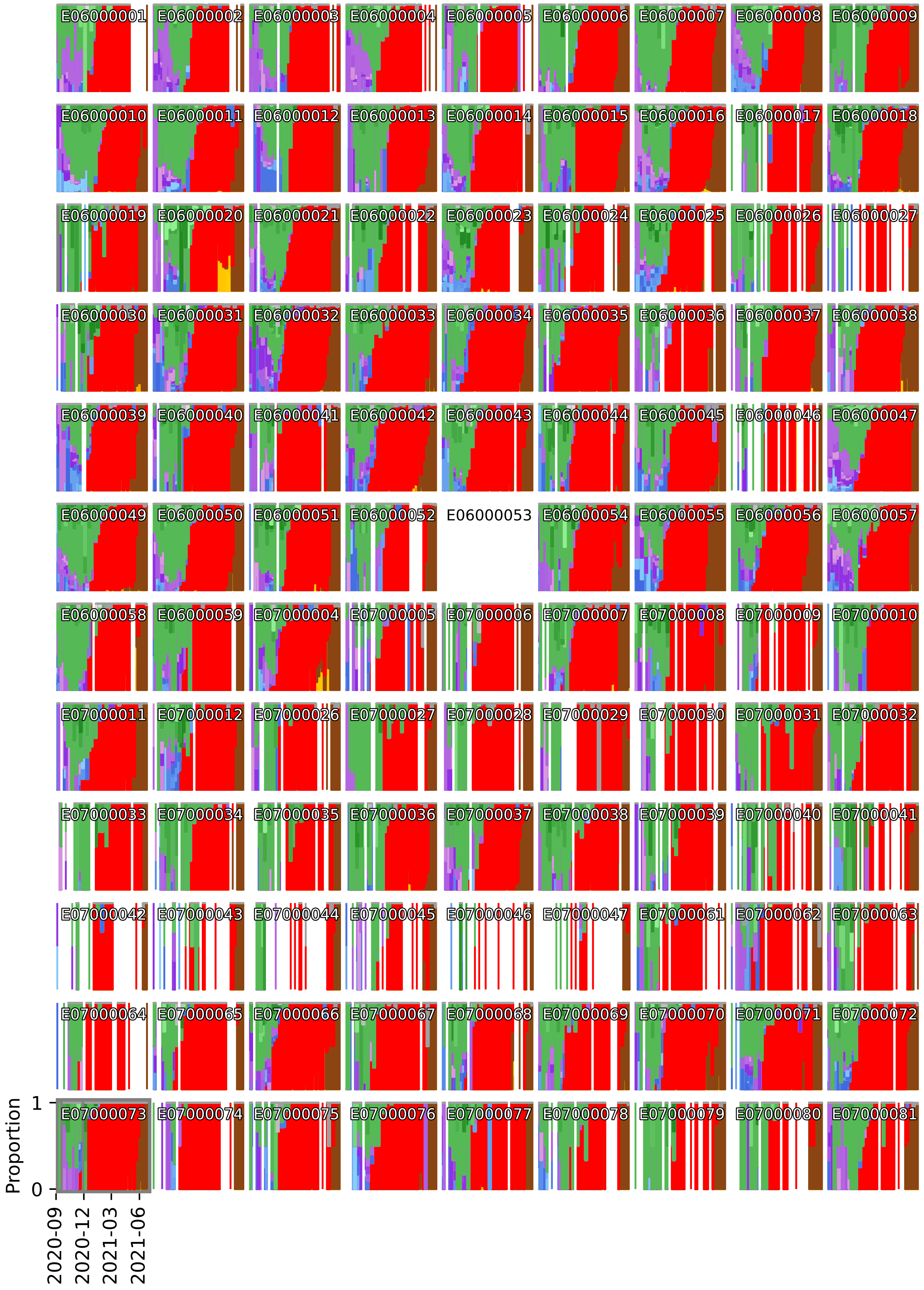
E06000001
E06000002
E06000003
E06000004
E06000005
E06000006
E06000007
E06000008
E06000009
E06000010
E06000011
E06000012
E06000013
E06000014
E06000015
E06000016
E06000017
E06000018
E06000019
E06000020
E06000021
E06000022
E06000023
E06000024
E06000025
E06000026
E06000027
E06000030
E06000031
E06000032
E06000033
E06000034
E06000035
E06000036
E06000037
E06000038
E06000039
E06000040
E06000041
E06000042
E06000043
E06000044
E06000045
E06000046
E06000047
E06000049
E06000050
E06000051
E06000052
E06000053
E06000054
E06000055
E06000056
E06000057
E06000058
E06000059
E07000004
E07000005
E07000006
E07000007
E07000008
E07000009
E07000010
E07000011
E07000012
E07000026
E07000027
E07000028
E07000029
E07000030
E07000031
E07000032
E07000033
E07000034
E07000035
E07000036
E07000037
E07000038
E07000039
E07000040
E07000041
E07000042
E07000043
E07000044
E07000045
E07000046
E07000047
E07000061
E07000062
E07000063
E07000064
E07000065
E07000066
E07000067
E07000068
E07000069
E07000070
E07000071
E07000072
E07000073
E07000074
E07000075
E07000076
E07000077
E07000078
E07000079
E07000080
E07000081
Proportion
1
0
2020-09
2020-12
2021-03
2021-06

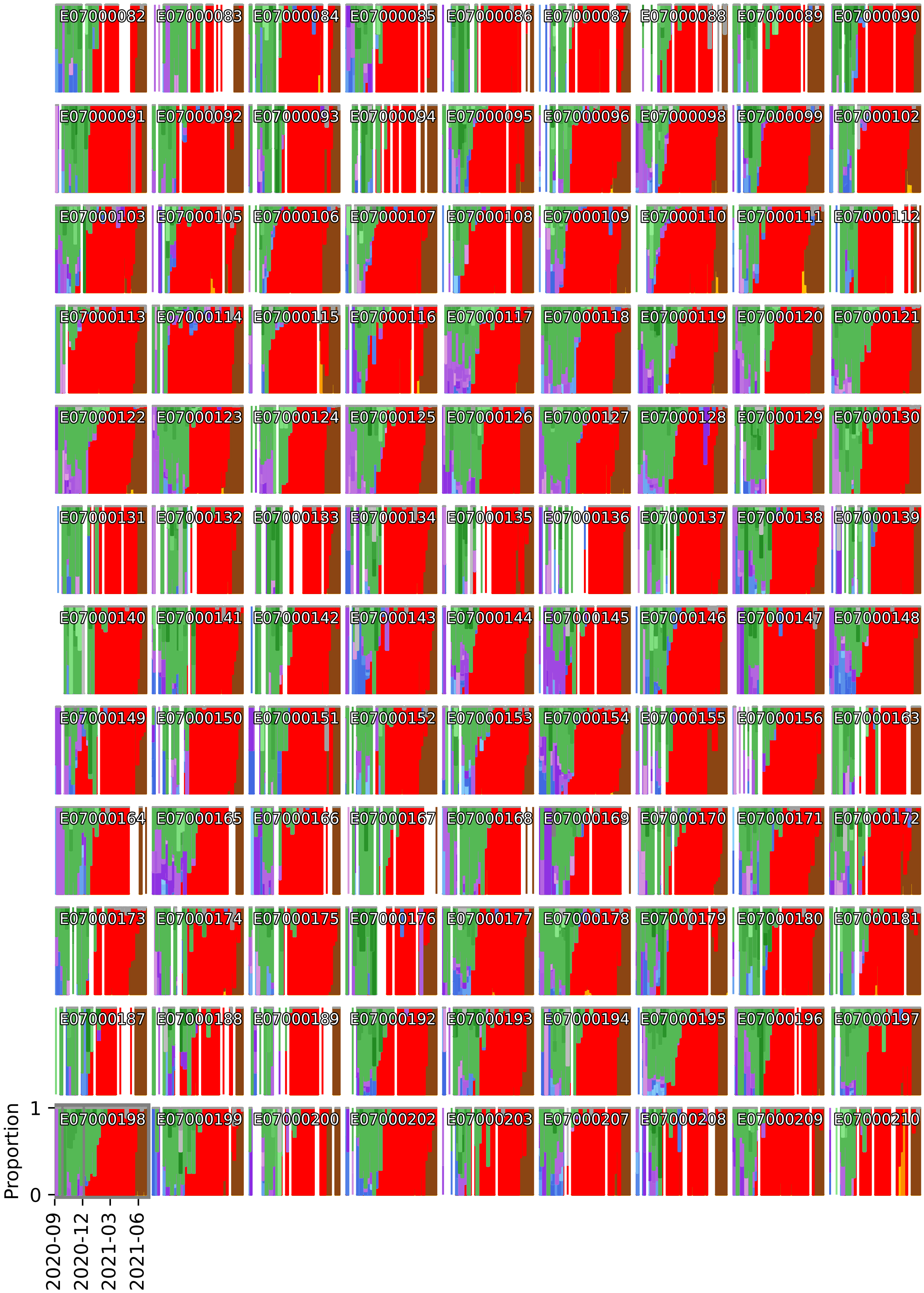
E07000082
E07000083
E07000084
E07000085
E07000086
E07000087
E07000088
E07000089
E07000090
E07000091
E07000092
E07000093
E07000094
E07000095
E07000096
E07000098
E07000099
E07000102
E07000103
E07000105
E07000106
E07000107
E07000108
E07000109
E07000110
E07000111
E07000112
E07000113
E07000114
E07000115
E07000116
E07000117
E07000118
E07000119
E07000120
E07000121
E07000122
E07000123
E07000124
E07000125
E07000126
E07000127
E07000128
E07000129
E07000130
E07000131
E07000132
E07000133
E07000134
E07000135
E07000136
E07000137
E07000138
E07000139
E07000140
E07000141
E07000142
E07000143
E07000144
E07000145
E07000146
E07000147
E07000148
E07000149
E07000150
E07000151
E07000152
E07000153
E07000154
E07000155
E07000156
E07000163
E07000164
E07000165
E07000166
E07000167
E07000168
E07000169
E07000170
E07000171
E07000172
E07000173
E07000174
E07000175
E07000176
E07000177
E07000178
E07000179
E07000180
E07000181
E07000187
E07000188
E07000189
E07000192
E07000193
E07000194
E07000195
E07000196
E07000197
E07000198
E07000199
E07000200
E07000202
E07000203
E07000207
E07000208
E07000209
E07000210
Proportion
1
0
2020-09
2020-12
2021-03
2021-06

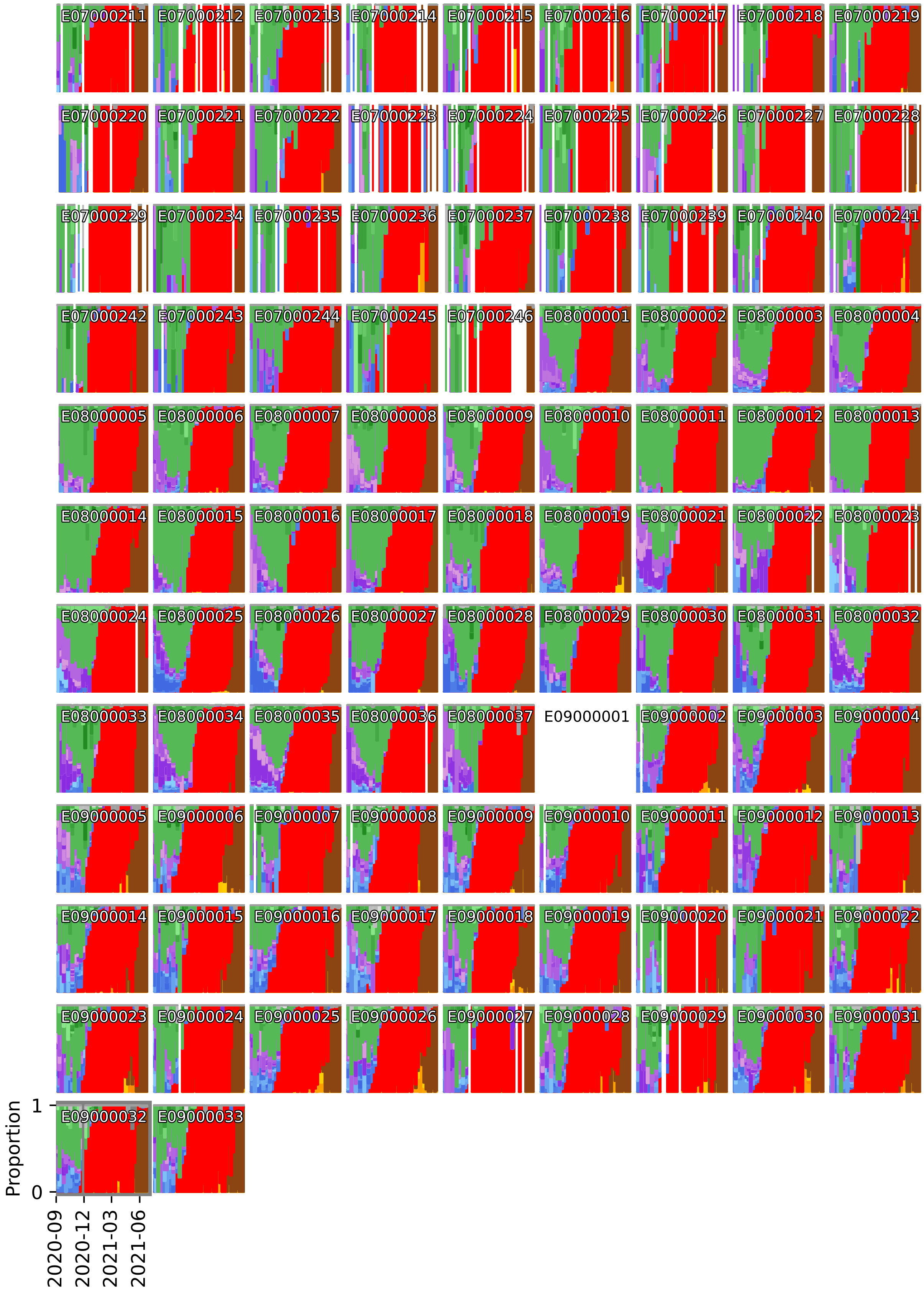
E07000211
E07000212
E07000213
E07000214
E07000215
E07000216
E07000217
E07000218
E07000219
E07000220
E07000221
E07000222
E07000223
E07000224
E07000225
E07000226
E07000227
E07000228
E07000229
E07000234
E07000235
E07000236
E07000237
E07000238
E07000239
E07000240
E07000241
E07000242
E07000243
E07000244
E07000245
E07000246
E08000001
E08000002
E08000003
E08000004
E08000005
E08000006
E08000007
E08000008
E08000009
E08000010
E08000011
E08000012
E08000013
E08000014
E08000015
E08000016
E08000017
E08000018
E08000019
E08000021
E08000022
E08000023
E08000024
E08000025
E08000026
E08000027
E08000028
E08000029
E08000030
E08000031
E08000032
E08000033
E08000034
E08000035
E08000036
E08000037
E09000001
E09000002
E09000003
E09000004
E09000005
E09000006
E09000007
E09000008
E09000009
E09000010
E09000011
E09000012
E09000013
E09000014
E09000015
E09000016
E09000017
E09000018
E09000019
E09000020
E09000021
E09000022
E09000023
E09000024
E09000025
E09000026
E09000027
E09000028
E09000029
E09000030
E09000031
E09000032
E09000033
Proportion
1
0
2020-09
2020-12
2021-03
2021-06

Supplementary Note 1 Fig. 2: **Modelled proportions of sequenced SARS-Cov2 lineages in 315 LTLA in England.** The titles in each tile refer to LTLA look up codes (Supplementary Note 3) and colours indicate major SARS-CoV-2 lineages (blue: B.1, purple: B.1.1, green: B.1.177, red: B.1.1.7 (Alpha), orange: other VOC/VUI, brown: B.1.617.2 (Delta) and grey: Other (A & B)).

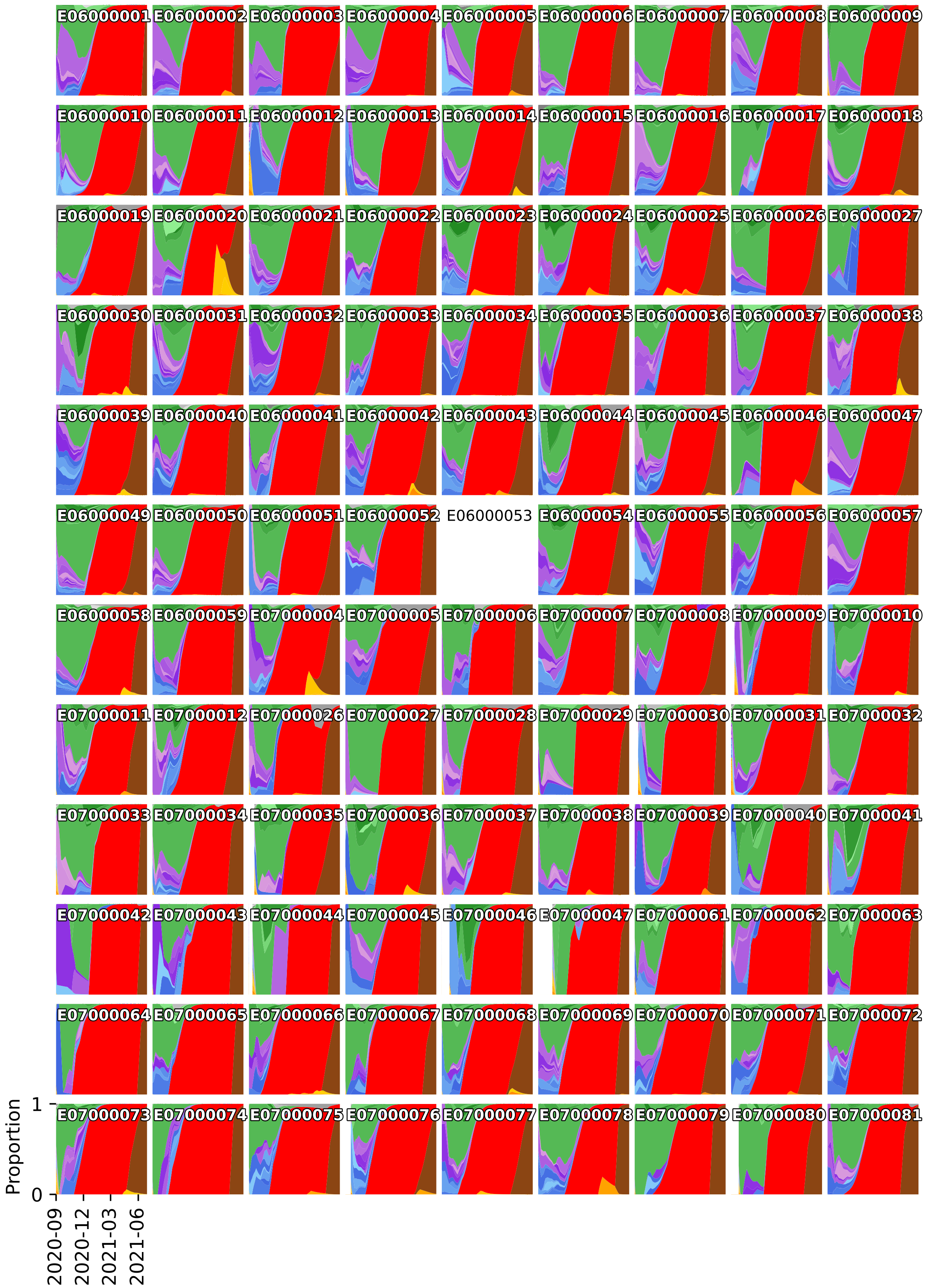
E06000001 E06000002 E06000003 E06000004 E06000005 E06000006 E06000007 E06000008 E06000009
E06000010 E06000011 E06000012 E06000013 E06000014 E06000015 E06000016 E06000017 E06000018
E06000019 E06000020 E06000021 E06000022 E06000023 E06000024 E06000025 E06000026 E06000027
E06000030 E06000031 E06000032 E06000033 E06000034 E06000035 E06000036 E06000037 E06000038
E06000039 E06000040 E06000041 E06000042 E06000043 E06000044 E06000045 E06000046 E06000047
E06000049 E06000050 E06000051 E06000052 E06000053 E06000054 E06000055 E06000056 E06000057
E06000058 E06000059 E07000004 E07000005 E07000006 E07000007 E07000008 E07000009 E07000010
E07000011 E07000012 E07000026 E07000027 E07000028 E07000029 E07000030 E07000031 E07000032
E07000033 E07000034 E07000035 E07000036 E07000037 E07000038 E07000039 E07000040 E07000041
E07000042 E07000043 E07000044 E07000045 E07000046 E07000047 E07000061 E07000062 E07000063
E07000064 E07000065 E07000066 E07000067 E07000068 E07000069 E07000070 E07000071 E07000072
E07000073 E07000074 E07000075 E07000076 E07000077 E07000078 E07000079 E07000080 E07000081
Proportion
1
0
2020-09
2020-12
2021-03
2021-06

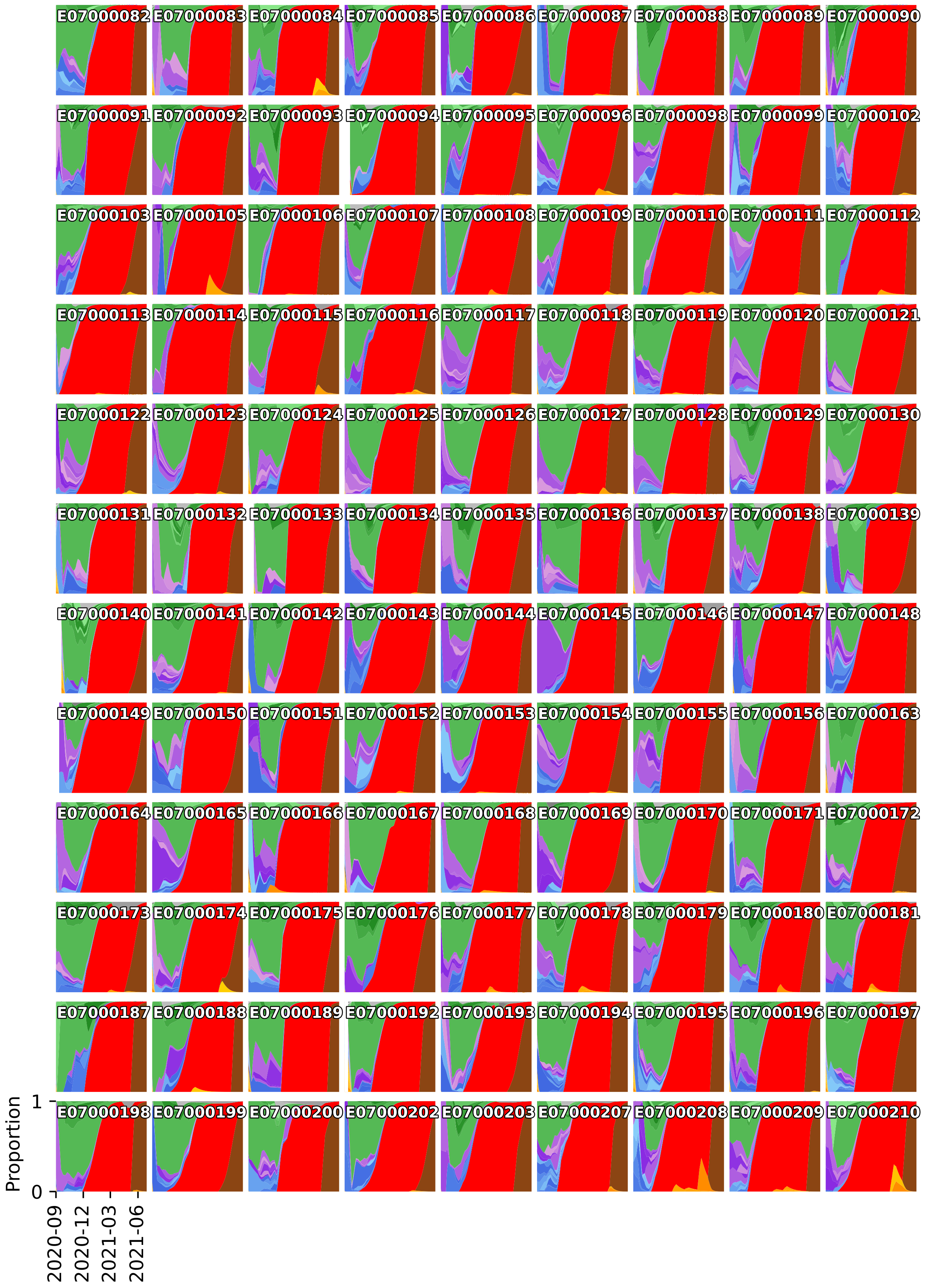
E07000082
E07000083
E07000084
E07000085
E07000086
E07000087
E07000088
E07000089
E07000090
E07000091
E07000092
E07000093
E07000094
E07000095
E07000096
E07000098
E07000099
E07000102
E07000103
E07000105
E07000106
E07000107
E07000108
E07000109
E07000110
E07000111
E07000112
E07000113
E07000114
E07000115
E07000116
E07000117
E07000118
E07000119
E07000120
E07000121
E07000122
E07000123
E07000124
E07000125
E07000126
E07000127
E07000128
E07000129
E07000130
E07000131
E07000132
E07000133
E07000134
E07000135
E07000136
E07000137
E07000138
E07000139
E07000140
E07000141
E07000142
E07000143
E07000144
E07000145
E07000146
E07000147
E07000148
E07000149
E07000150
E07000151
E07000152
E07000153
E07000154
E07000155
E07000156
E07000163
E07000164
E07000165
E07000166
E07000167
E07000168
E07000169
E07000170
E07000171
E07000172
E07000173
E07000174
E07000175
E07000176
E07000177
E07000178
E07000179
E07000180
E07000181
E07000187
E07000188
E07000189
E07000192
E07000193
E07000194
E07000195
E07000196
E07000197
E07000198
E07000199
E07000200
E07000202
E07000203
E07000207
E07000208
E07000209
E07000210
Proportion
1
0
2020-09
2020-12
2021-03
2021-06

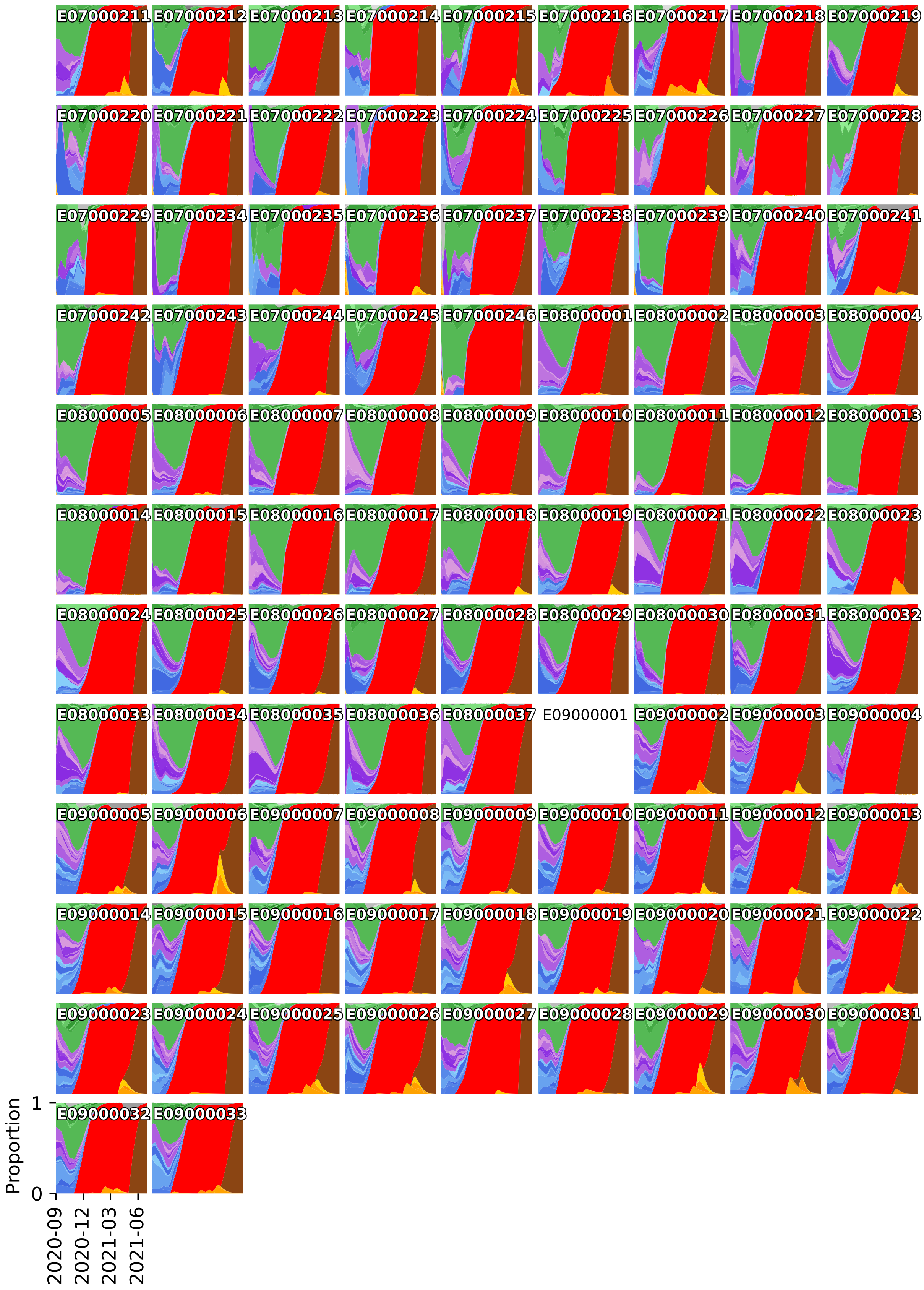
E07000211
E07000212
E07000213
E07000214
E07000215
E07000216
E07000217
E07000218
E07000219
E07000220
E07000221
E07000222
E07000223
E07000224
E07000225
E07000226
E07000227
E07000228
E07000229
E07000234
E07000235
E07000236
E07000237
E07000238
E07000239
E07000240
E07000241
E07000242
E07000243
E07000244
E07000245
E07000246
E08000001
E08000002
E08000003
E08000004
E08000005
E08000006
E08000007
E08000008
E08000009
E08000010
E08000011
E08000012
E08000013
E08000014
E08000015
E08000016
E08000017
E08000018
E08000019
E08000021
E08000022
E08000023
E08000024
E08000025
E08000026
E08000027
E08000028
E08000029
E08000030
E08000031
E08000032
E08000033
E08000034
E08000035
E08000036
E08000037
E09000001
E09000002
E09000003
E09000004
E09000005
E09000006
E09000007
E09000008
E09000009
E09000010
E09000011
E09000012
E09000013
E09000014
E09000015
E09000016
E09000017
E09000018
E09000019
E09000020
E09000021
E09000022
E09000023
E09000024
E09000025
E09000026
E09000027
E09000028
E09000029
E09000030
E09000031
E09000032
E09000033
Proportion
1
0
2020-09
2020-12
2021-03
2021-06

### 2 Supplementary Note 2

Supplementary Note 2 Fig. 1: **Spatiotemporal model of 71 SARS-CoV-2 lineages in 315 English LTLAs between September 2020 and June 2021.** Upper panel: Total and relative lineage-specific incidence of B.1.177, B.1.1.7 (Alpha), B.1.617.2 (Delta) and prevalent VUI/VOC (if present). Middle panel: Modelled proportions of B.1.177, B.1.1.7 (Alpha), B.1.617.2 (Delta) and prevalent VUI/VOC (if present). Bottom panel: Local lineage-specific $R_t$ values for B.1.177, B.1.1.7 (Alpha), B.1.617.2 (Delta) and prevalent VUI/VOC (if present) and the average $R_t$ value (growth per 5.1d) of all other lineages in the same periods. In a small number of LTLAs (Redcar and Cleveland, Southampton, County Durham, Cheshire West and Chester, Cornwall, Bournemouth, Erewash, Stockport) we noted very large $R_t$ values at around the time of the introduction of the delta variant. These are more likely reflections of stochastic introduction events at times of very low incidence, rather than sustained transmission.

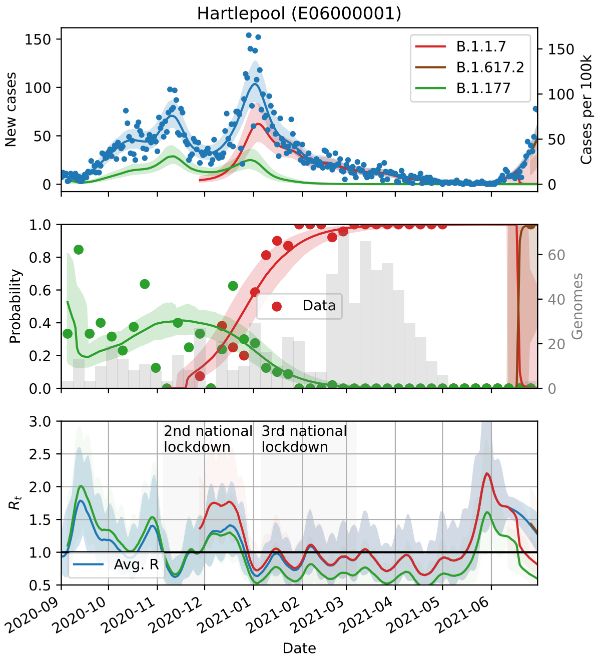
Hartlepool (E06000001)
B.1.1.7
B.1.617.2
B.1.177
New cases
Cases per 100k
Data
Probability
Genomes
2nd national lockdown
3rd national lockdown
Avg. R
$R_t$
Date

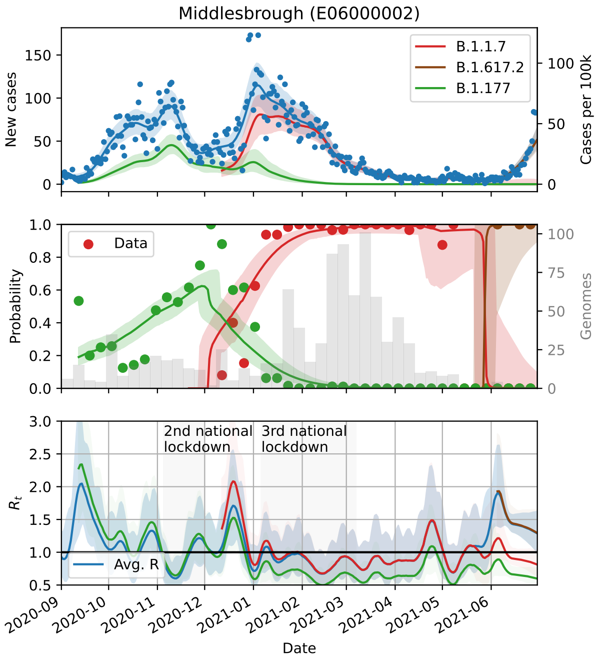
Middlesbrough (E06000002)
B.1.1.7
B.1.617.2
B.1.177
New cases
Cases per 100k
Data
Probability
Genomes
2nd national lockdown
3rd national lockdown
Avg. R
$R_t$
Date

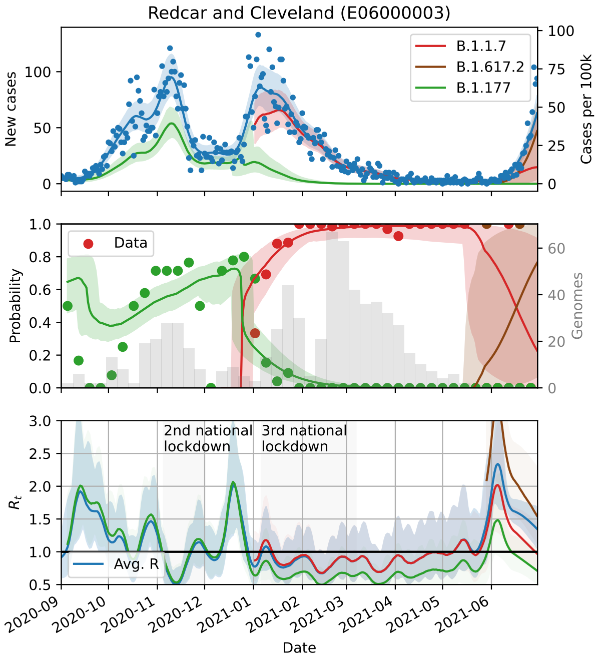
Redcar and Cleveland (E06000003)
B.1.1.7
B.1.617.2
B.1.177
New cases
Cases per 100k
Data
Probability
Genomes
2nd national lockdown
3rd national lockdown
Avg. R
$R_t$
Date

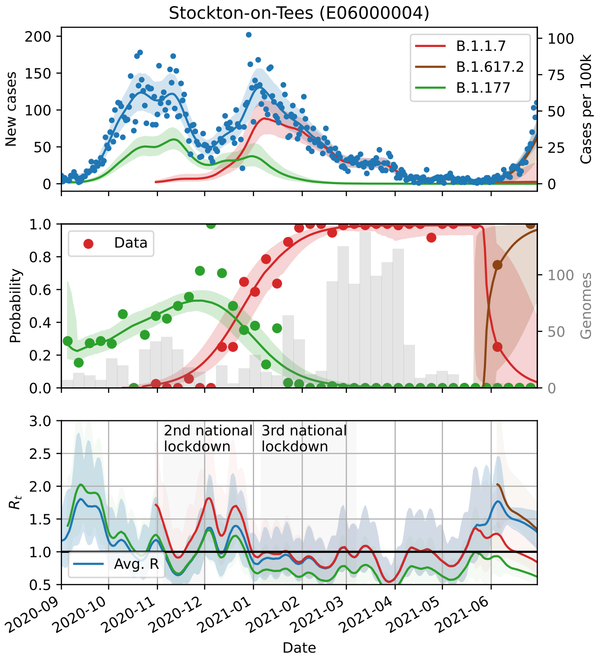
Stockton-on-Tees (E06000004)
B.1.1.7
B.1.617.2
B.1.177
New cases
Cases per 100k
Data
Probability
Genomes
2nd national lockdown
3rd national lockdown
Avg. R
$R_t$
Date

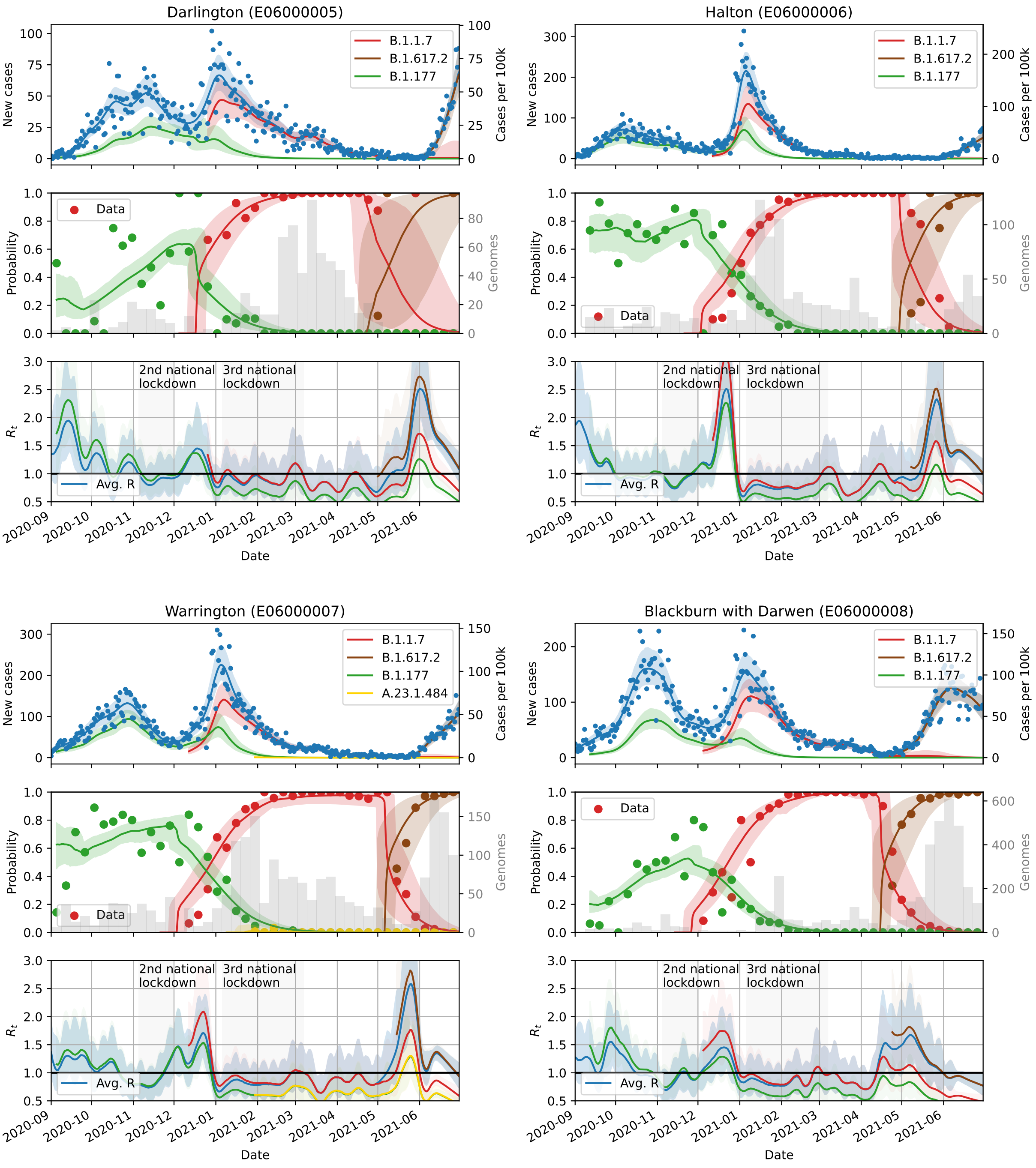
Darlington (E06000005)
Halton (E06000006)
Warrington (E06000007)
Blackburn with Darwen (E06000008)
B.1.1.7
B.1.617.2
B.1.177
A.23.1.484
Data
Avg. R
New cases
Cases per 100k
Probability
Genomes
$R_t$
2nd national lockdown
3rd national lockdown
2020-09
2020-10
2020-11
2020-12
2021-01
2021-02
2021-03
2021-04
2021-05
2021-06
Date

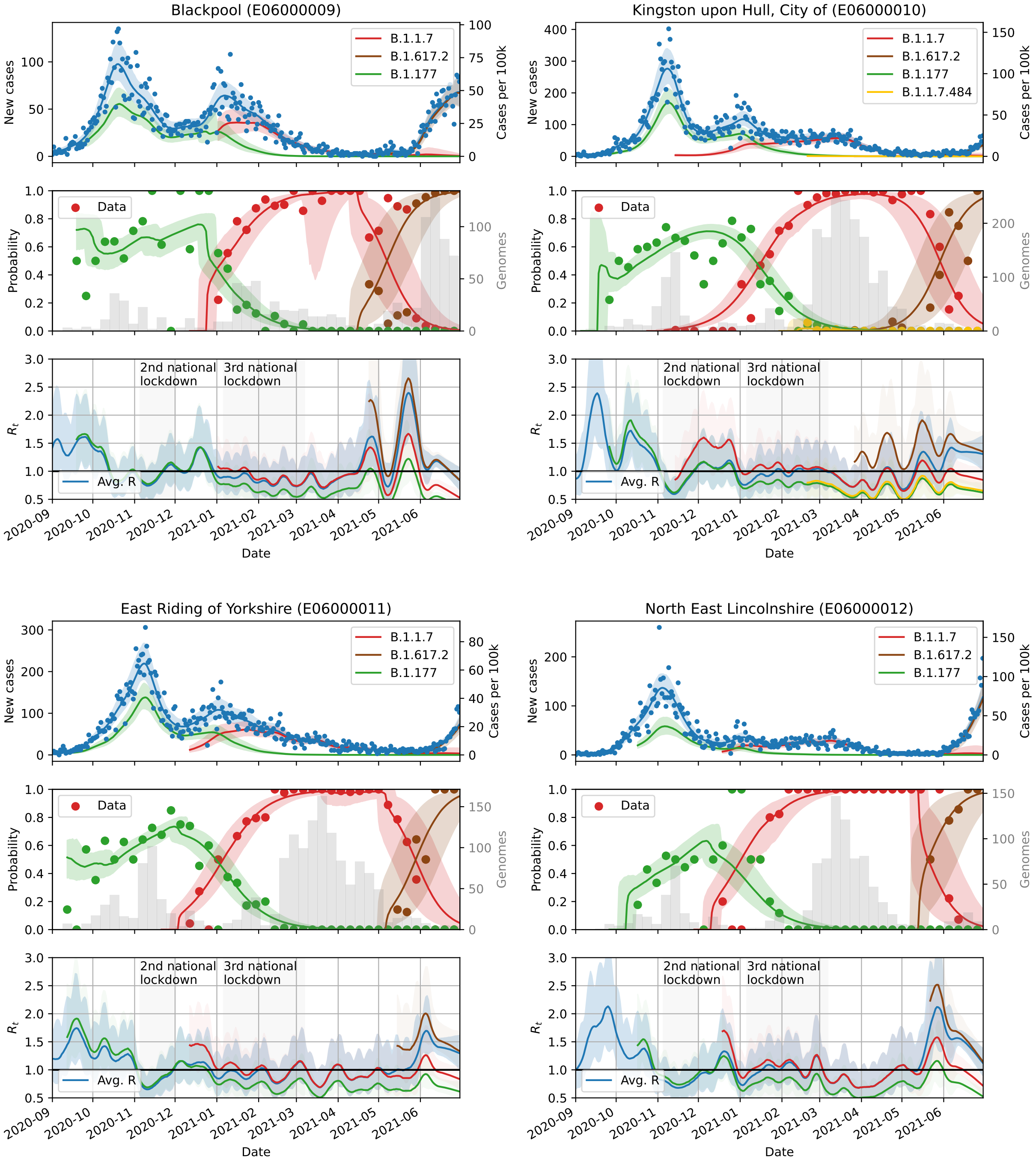
Blackpool (E06000009)
Kingston upon Hull, City of (E06000010)
East Riding of Yorkshire (E06000011)
North East Lincolnshire (E06000012)
B.1.1.7
B.1.617.2
B.1.177
B.1.1.7.484
New cases
Cases per 100k
Data
Probability
Genomes
2nd national lockdown
3rd national lockdown
Avg. R
$R_t$
2020-09
2020-10
2020-11
2020-12
2021-01
2021-02
2021-03
2021-04
2021-05
2021-06
Date

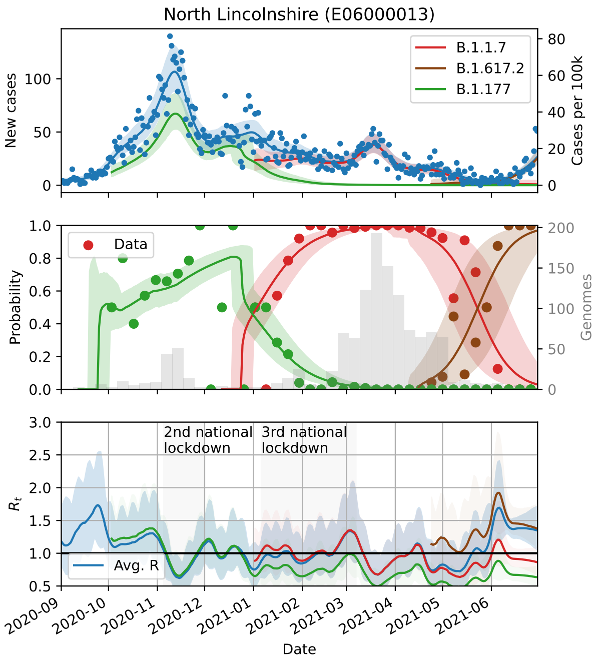
North Lincolnshire (E06000013)
B.1.1.7
B.1.617.2
B.1.177
New cases
Cases per 100k
Data
Probability
Genomes
2nd national lockdown
3rd national lockdown
Avg. R
$R_t$
Date

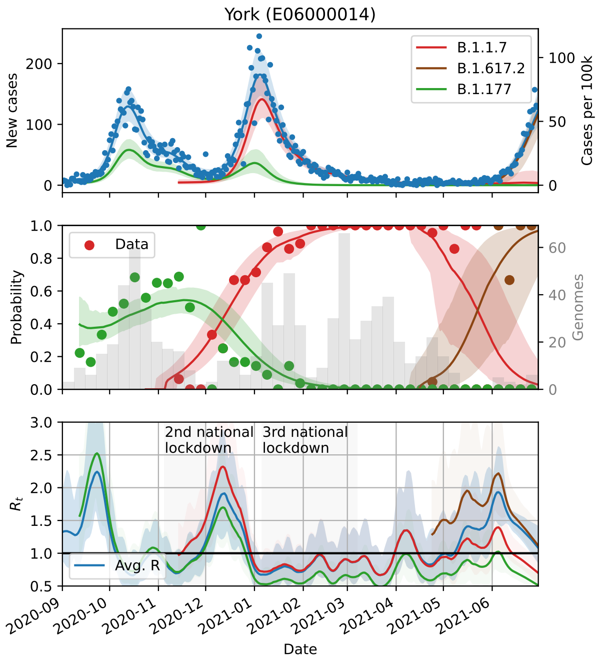
York (E06000014)
B.1.1.7
B.1.617.2
B.1.177
New cases
Cases per 100k
Data
Probability
Genomes
2nd national lockdown
3rd national lockdown
Avg. R
$R_t$
Date

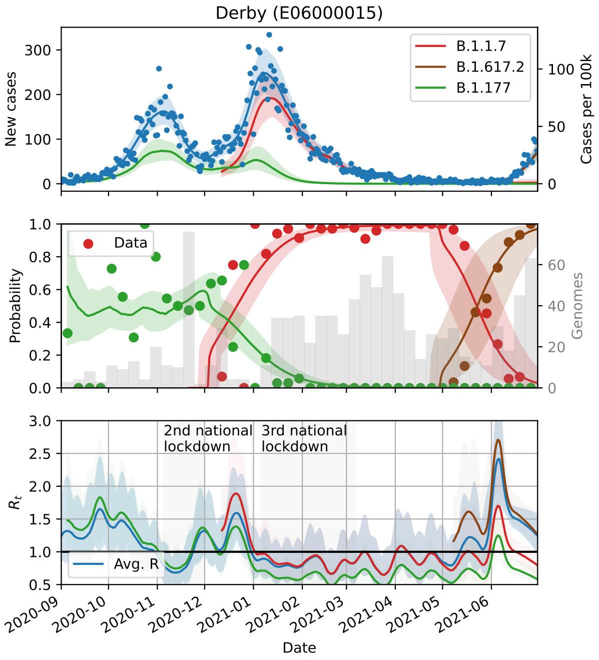
Derby (E06000015)
B.1.1.7
B.1.617.2
B.1.177
New cases
Cases per 100k
Data
Probability
Genomes
2nd national lockdown
3rd national lockdown
Avg. R
$R_t$
Date

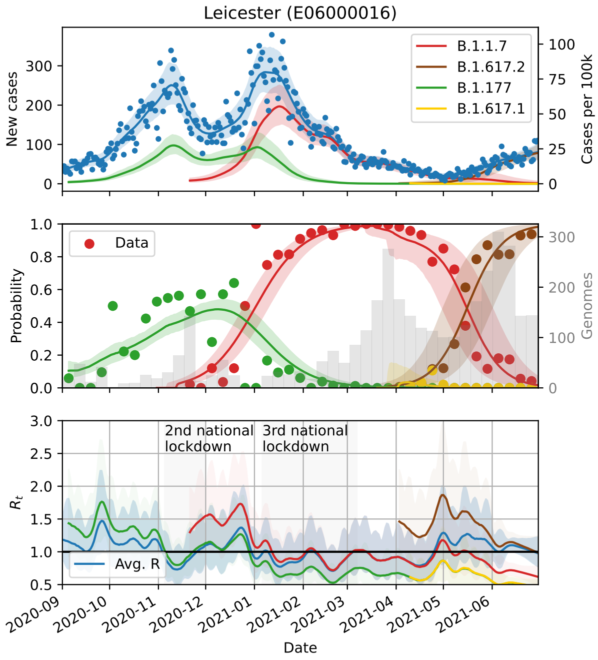
Leicester (E06000016)
B.1.1.7
B.1.617.2
B.1.177
B.1.617.1
New cases
Cases per 100k
Data
Probability
Genomes
2nd national lockdown
3rd national lockdown
Avg. R
$R_t$
Date

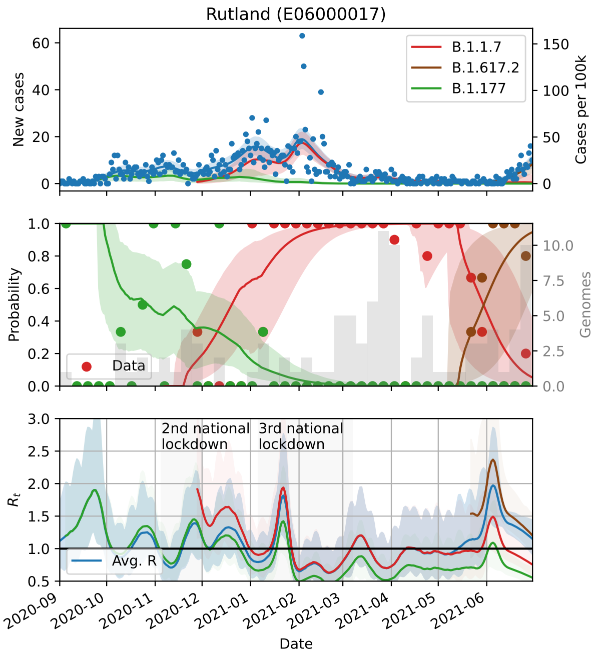
Rutland (E06000017)
New cases
Cases per 100k
B.1.1.7
B.1.617.2
B.1.177
Probability
Genomes
Data
2nd national lockdown
3rd national lockdown
$R_t$
Avg. R
Date

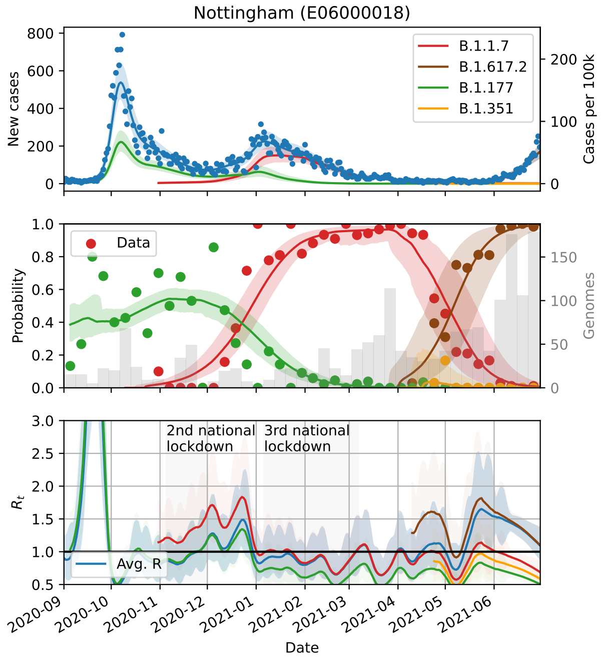
Nottingham (E06000018)
New cases
Cases per 100k
B.1.1.7
B.1.617.2
B.1.177
B.1.351
Probability
Genomes
Data
2nd national lockdown
3rd national lockdown
$R_t$
Avg. R
Date

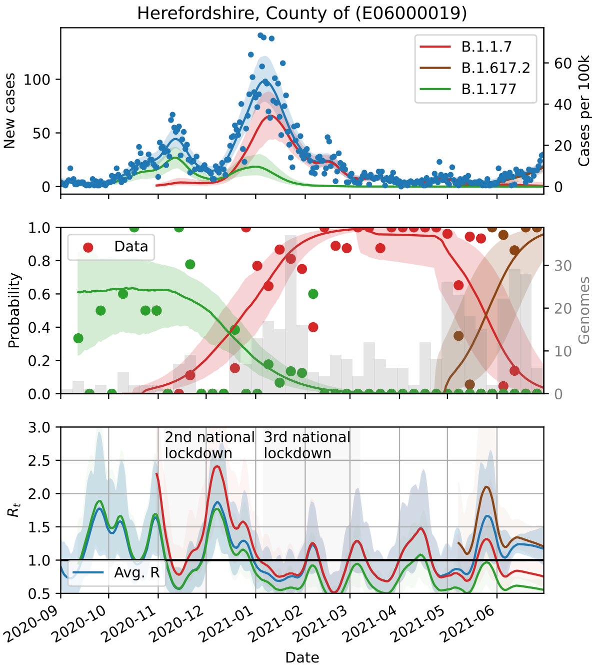
Herefordshire, County of (E06000019)
New cases
Cases per 100k
B.1.1.7
B.1.617.2
B.1.177
Probability
Genomes
Data
2nd national lockdown
3rd national lockdown
$R_t$
Avg. R
Date

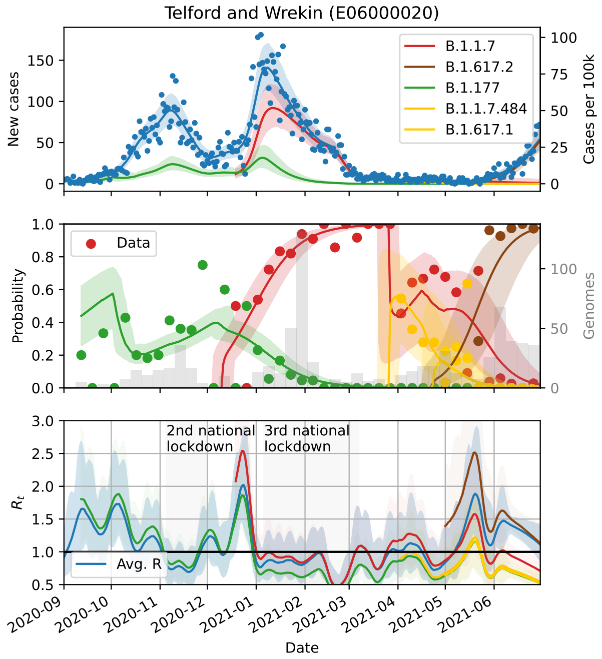
Telford and Wrekin (E06000020)
New cases
Cases per 100k
B.1.1.7
B.1.617.2
B.1.177
B.1.1.7.484
B.1.617.1
Probability
Genomes
Data
2nd national lockdown
3rd national lockdown
$R_t$
Avg. R
Date

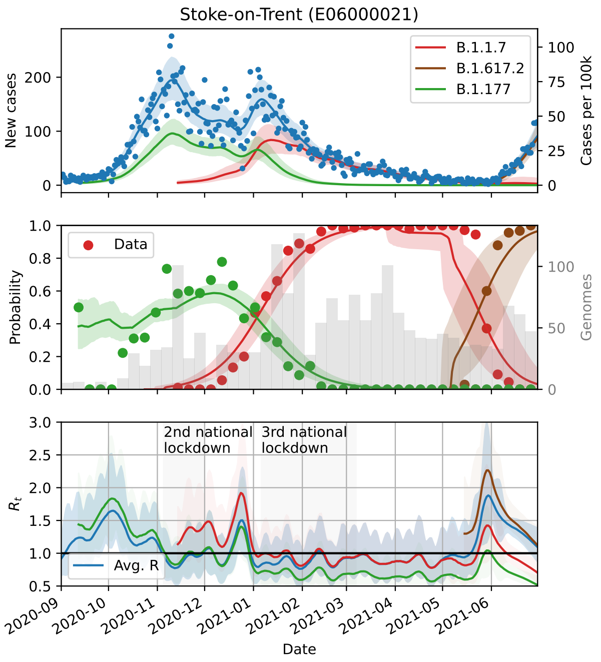
Stoke-on-Trent (E06000021)
New cases
Cases per 100k
B.1.1.7
B.1.617.2
B.1.177
Data
Probability
Genomes
2nd national lockdown
3rd national lockdown
$R_t$
Avg. R
Date

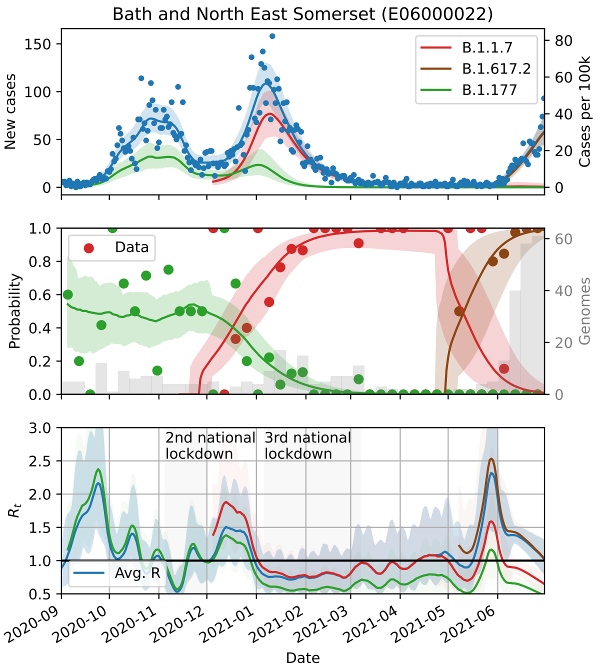
Bath and North East Somerset (E06000022)
New cases
Cases per 100k
B.1.1.7
B.1.617.2
B.1.177
Data
Probability
Genomes
2nd national lockdown
3rd national lockdown
$R_t$
Avg. R
Date

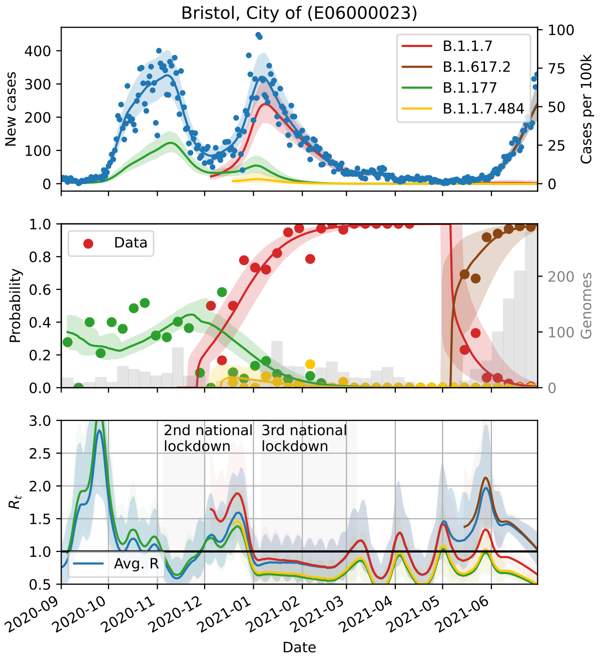
Bristol, City of (E06000023)
New cases
Cases per 100k
B.1.1.7
B.1.617.2
B.1.177
B.1.1.7.484
Data
Probability
Genomes
2nd national lockdown
3rd national lockdown
$R_t$
Avg. R
Date

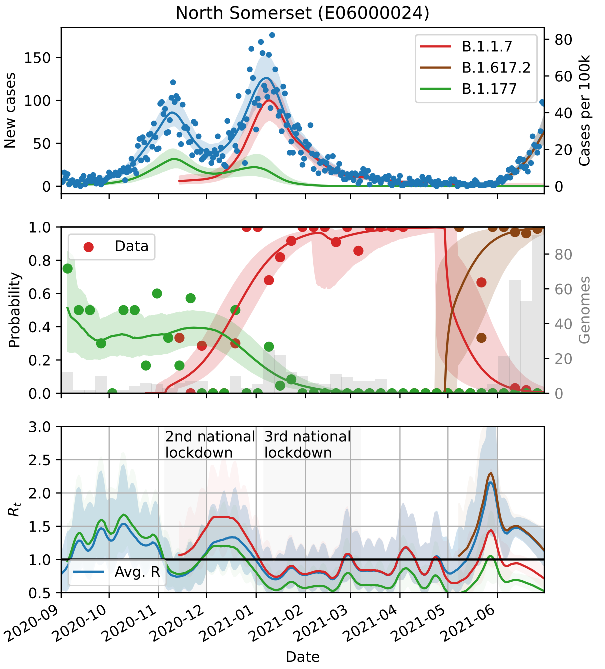
North Somerset (E06000024)
New cases
Cases per 100k
B.1.1.7
B.1.617.2
B.1.177
Data
Probability
Genomes
2nd national lockdown
3rd national lockdown
$R_t$
Avg. R
Date

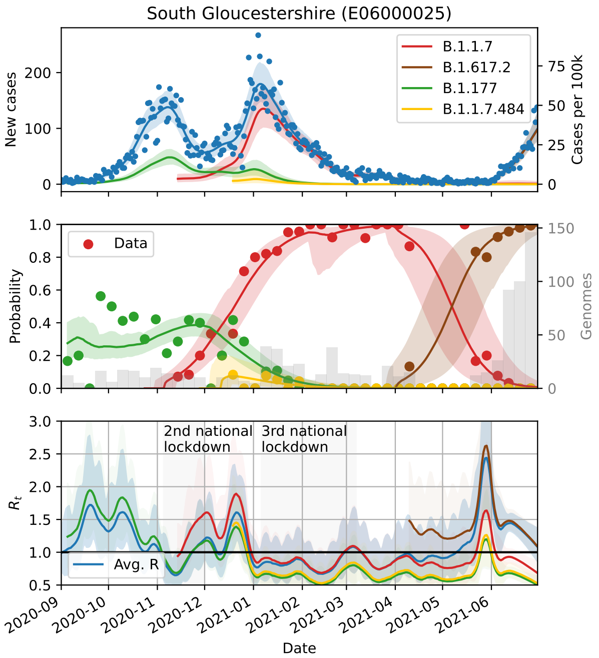
South Gloucestershire (E06000025)
B.1.1.7
B.1.617.2
B.1.177
B.1.1.7.484
New cases
Cases per 100k
Data
Probability
Genomes
2nd national lockdown
3rd national lockdown
Avg. R
$R_t$
Date

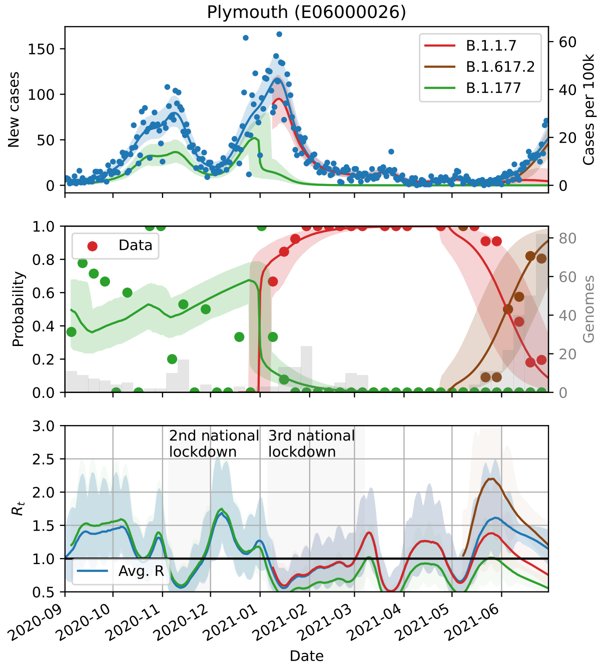
Plymouth (E06000026)
B.1.1.7
B.1.617.2
B.1.177
New cases
Cases per 100k
Data
Probability
Genomes
2nd national lockdown
3rd national lockdown
Avg. R
$R_t$
Date

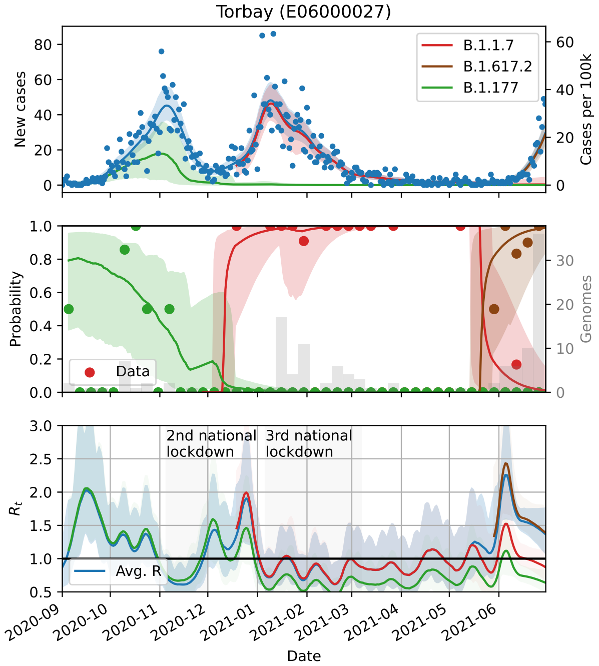
Torbay (E06000027)
B.1.1.7
B.1.617.2
B.1.177
New cases
Cases per 100k
Data
Probability
Genomes
2nd national lockdown
3rd national lockdown
Avg. R
$R_t$
Date

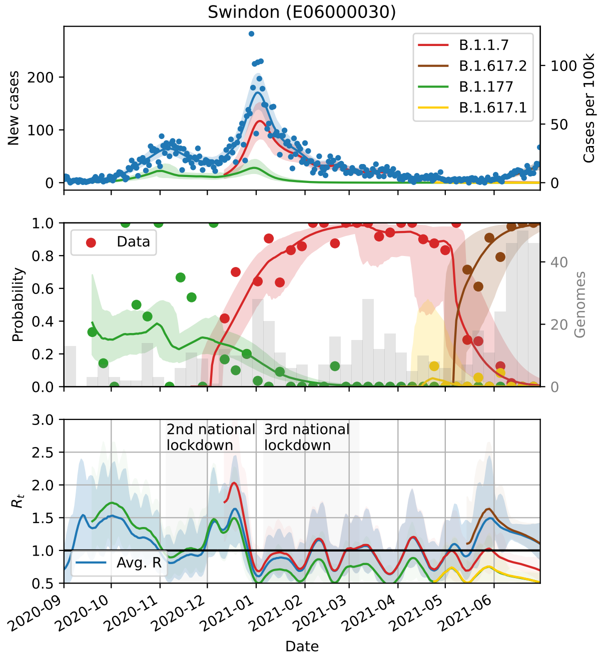
Swindon (E06000030)
B.1.1.7
B.1.617.2
B.1.177
B.1.617.1
New cases
Cases per 100k
Data
Probability
Genomes
2nd national lockdown
3rd national lockdown
Avg. R
$R_t$
Date

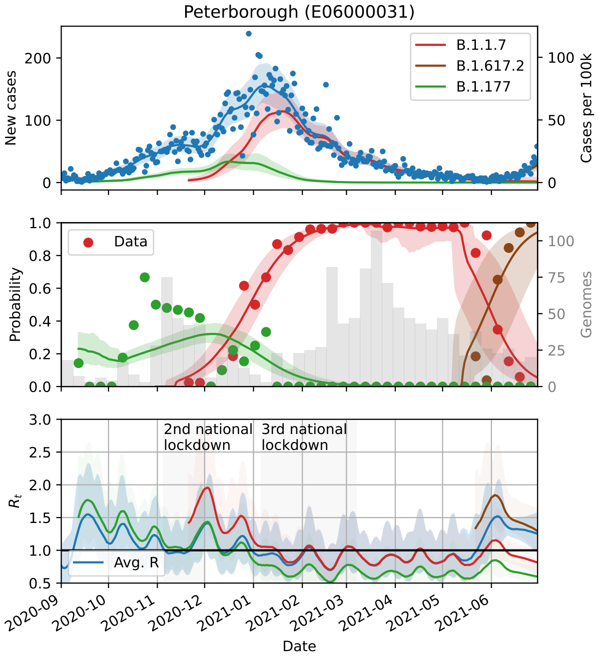
Peterborough (E06000031)
B.1.1.7
B.1.617.2
B.1.177
New cases
Cases per 100k
Data
Probability
Genomes
2nd national lockdown
3rd national lockdown
$R_t$
Avg. R
2020-09
2020-10
2020-11
2020-12
2021-01
2021-02
2021-03
2021-04
2021-05
2021-06
Date

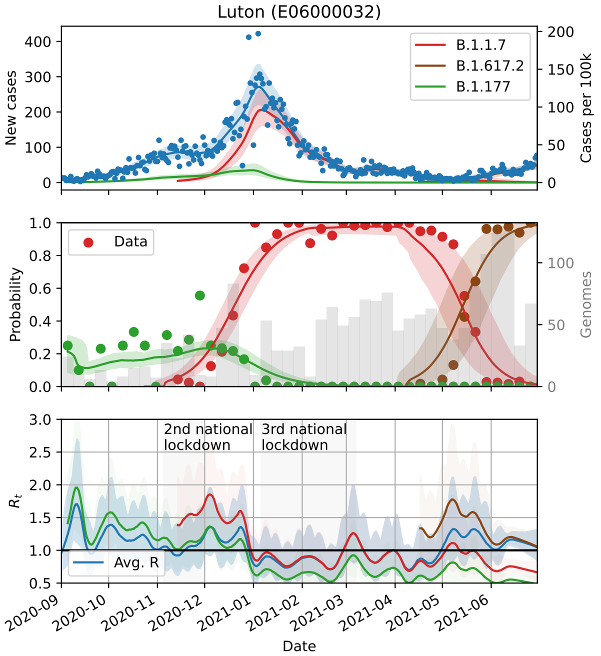
Luton (E06000032)
B.1.1.7
B.1.617.2
B.1.177
New cases
Cases per 100k
Data
Probability
Genomes
2nd national lockdown
3rd national lockdown
$R_t$
Avg. R
2020-09
2020-10
2020-11
2020-12
2021-01
2021-02
2021-03
2021-04
2021-05
2021-06
Date

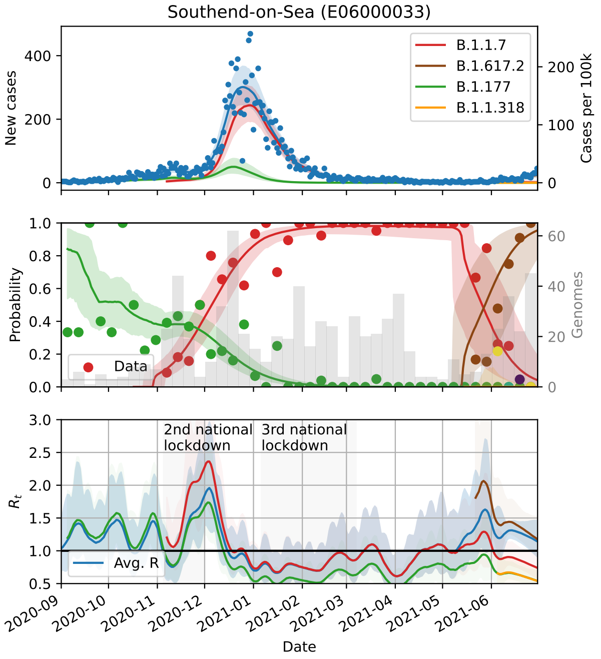
Southend-on-Sea (E06000033)
B.1.1.7
B.1.617.2
B.1.177
B.1.1.318
New cases
Cases per 100k
Data
Probability
Genomes
2nd national lockdown
3rd national lockdown
$R_t$
Avg. R
2020-09
2020-10
2020-11
2020-12
2021-01
2021-02
2021-03
2021-04
2021-05
2021-06
Date

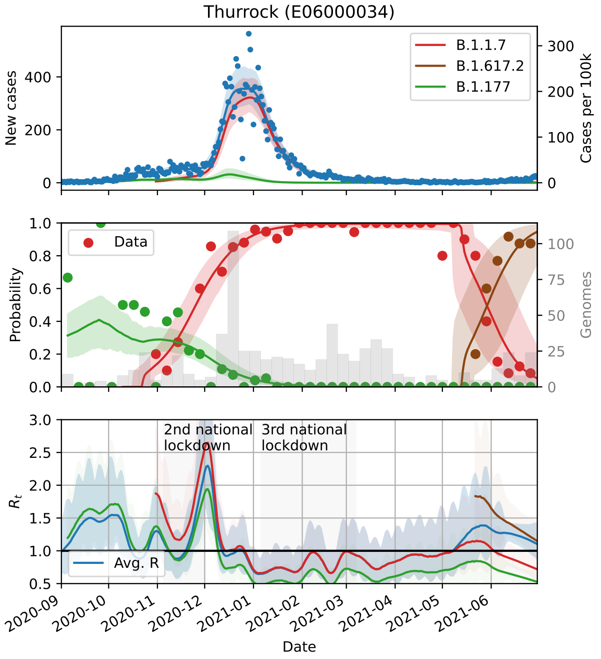
Thurrock (E06000034)
B.1.1.7
B.1.617.2
B.1.177
New cases
Cases per 100k
Data
Probability
Genomes
2nd national lockdown
3rd national lockdown
$R_t$
Avg. R
2020-09
2020-10
2020-11
2020-12
2021-01
2021-02
2021-03
2021-04
2021-05
2021-06
Date

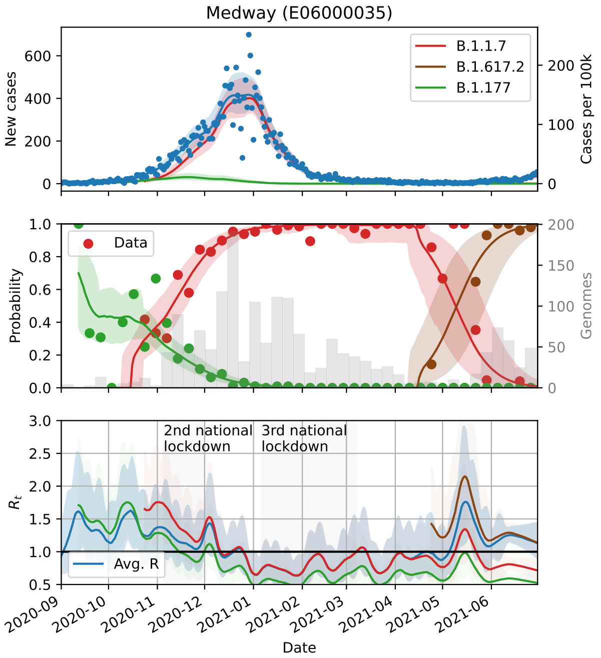
Medway (E06000035)
New cases
Cases per 100k
B.1.1.7
B.1.617.2
B.1.177
Probability
Genomes
Data
$R_t$
2nd national lockdown
3rd national lockdown
Avg. R
Date

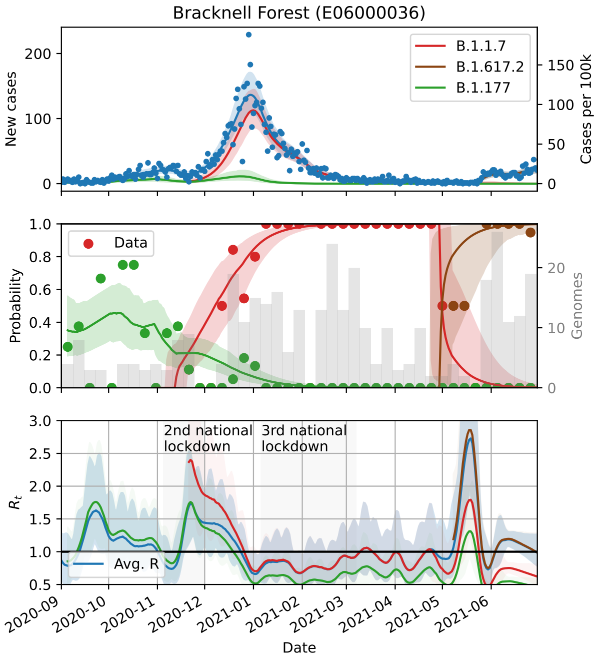
Bracknell Forest (E06000036)
New cases
Cases per 100k
B.1.1.7
B.1.617.2
B.1.177
Probability
Genomes
Data
$R_t$
2nd national lockdown
3rd national lockdown
Avg. R
Date

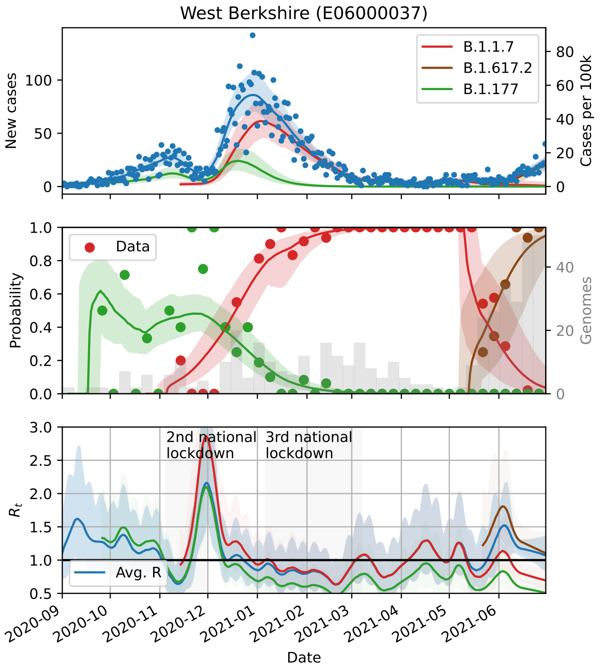
West Berkshire (E06000037)
New cases
Cases per 100k
B.1.1.7
B.1.617.2
B.1.177
Probability
Genomes
Data
$R_t$
2nd national lockdown
3rd national lockdown
Avg. R
Date

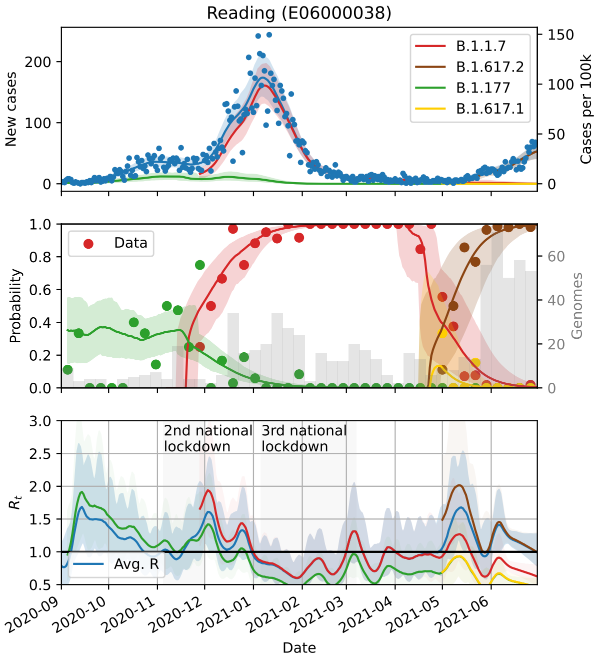
Reading (E06000038)
New cases
Cases per 100k
B.1.1.7
B.1.617.2
B.1.177
B.1.617.1
Probability
Genomes
Data
$R_t$
2nd national lockdown
3rd national lockdown
Avg. R
Date

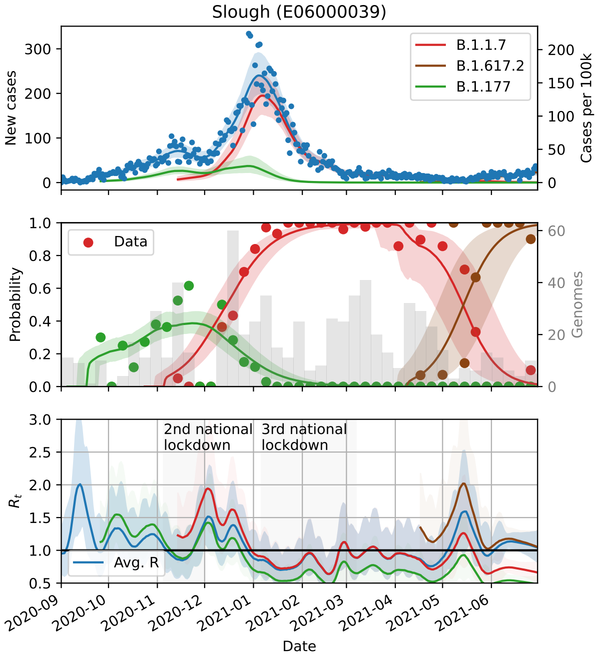
Slough (E06000039)
New cases
Cases per 100k
B.1.1.7
B.1.617.2
B.1.177
Data
Probability
Genomes
2nd national lockdown
3rd national lockdown
$R_t$
Avg. R
2020-09
2020-10
2020-11
2020-12
2021-01
2021-02
2021-03
2021-04
2021-05
2021-06
Date

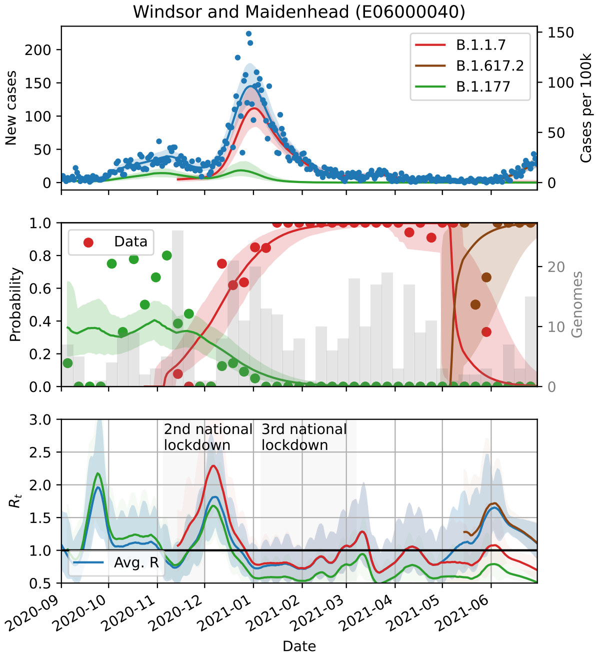
Windsor and Maidenhead (E06000040)
New cases
Cases per 100k
B.1.1.7
B.1.617.2
B.1.177
Data
Probability
Genomes
2nd national lockdown
3rd national lockdown
$R_t$
Avg. R
2020-09
2020-10
2020-11
2020-12
2021-01
2021-02
2021-03
2021-04
2021-05
2021-06
Date

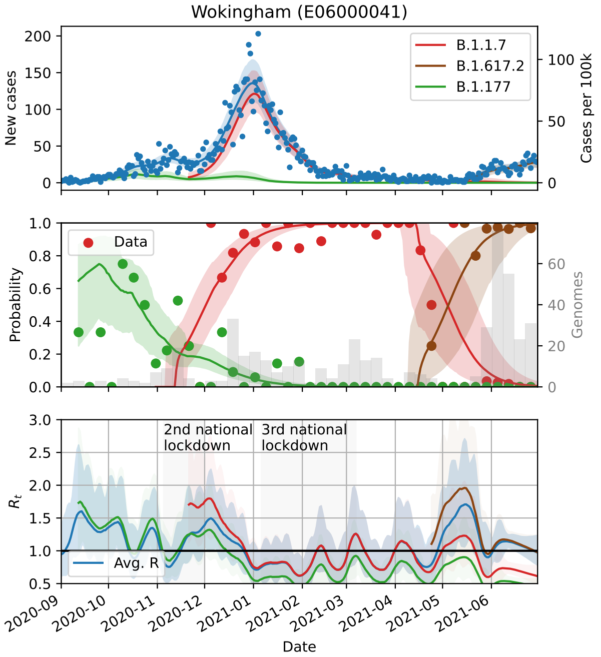
Wokingham (E06000041)
New cases
Cases per 100k
B.1.1.7
B.1.617.2
B.1.177
Data
Probability
Genomes
2nd national lockdown
3rd national lockdown
$R_t$
Avg. R
2020-09
2020-10
2020-11
2020-12
2021-01
2021-02
2021-03
2021-04
2021-05
2021-06
Date

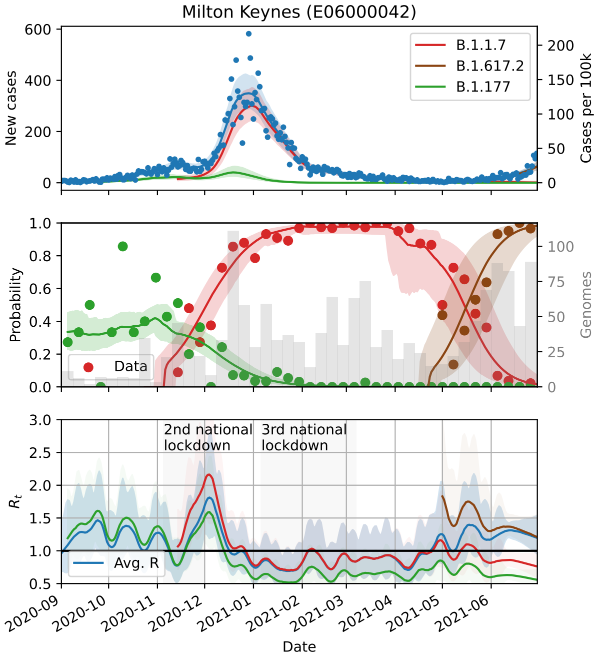
Milton Keynes (E06000042)
New cases
Cases per 100k
B.1.1.7
B.1.617.2
B.1.177
Data
Probability
Genomes
2nd national lockdown
3rd national lockdown
$R_t$
Avg. R
2020-09
2020-10
2020-11
2020-12
2021-01
2021-02
2021-03
2021-04
2021-05
2021-06
Date

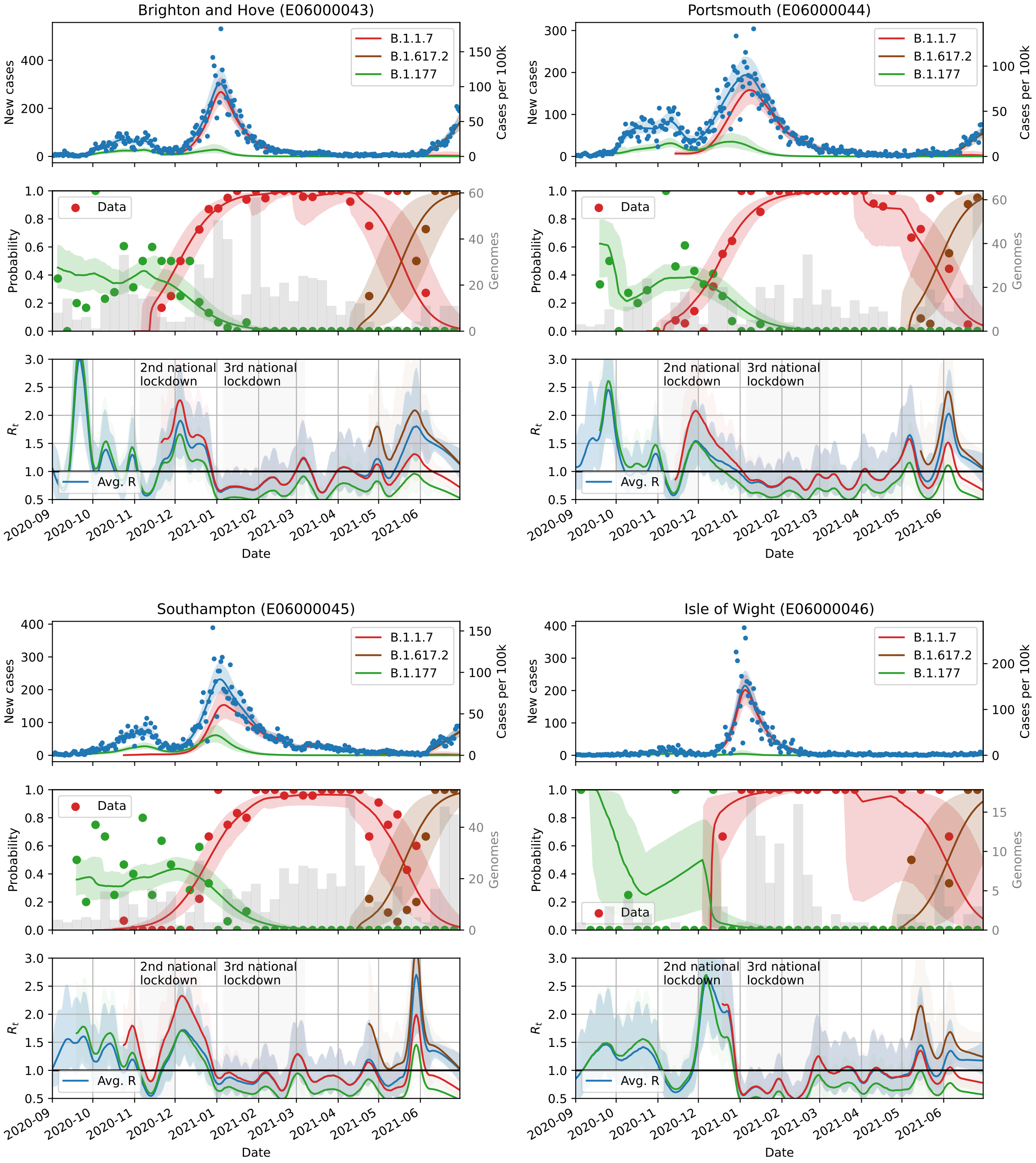

Brighton and Hove (E06000043)
Portsmouth (E06000044)
Southampton (E06000045)
Isle of Wight (E06000046)
B.1.1.7
B.1.617.2
B.1.177
New cases
Cases per 100k
Probability
Genomes
Data
$R_t$
Avg. R
2nd national lockdown
3rd national lockdown
Date

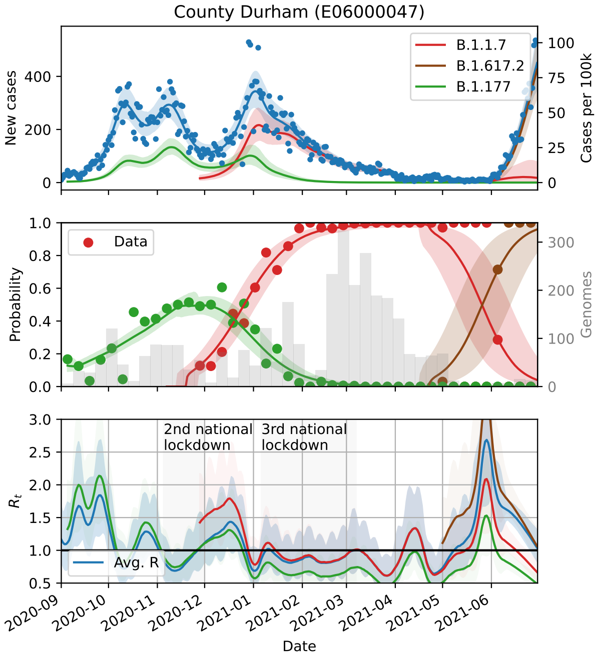
County Durham (E06000047)
New cases
Cases per 100k
B.1.1.7
B.1.617.2
B.1.177
Data
Probability
Genomes
2nd national lockdown
3rd national lockdown
Avg. R
$R_t$
Date

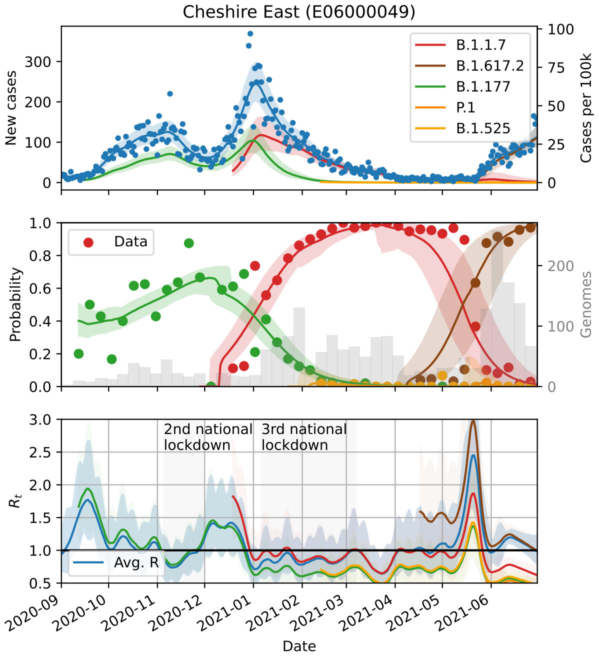
Cheshire East (E06000049)
New cases
Cases per 100k
B.1.1.7
B.1.617.2
B.1.177
P.1
B.1.525
Data
Probability
Genomes
2nd national lockdown
3rd national lockdown
Avg. R
$R_t$
Date

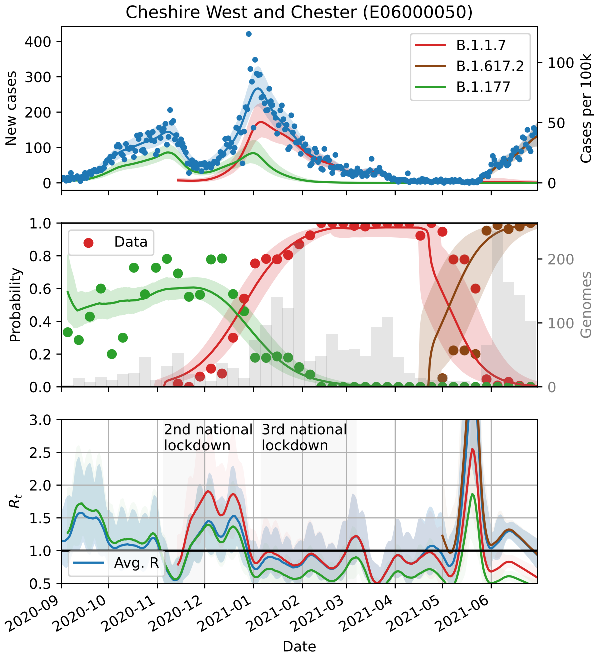
Cheshire West and Chester (E06000050)
New cases
Cases per 100k
B.1.1.7
B.1.617.2
B.1.177
Data
Probability
Genomes
2nd national lockdown
3rd national lockdown
Avg. R
$R_t$
Date

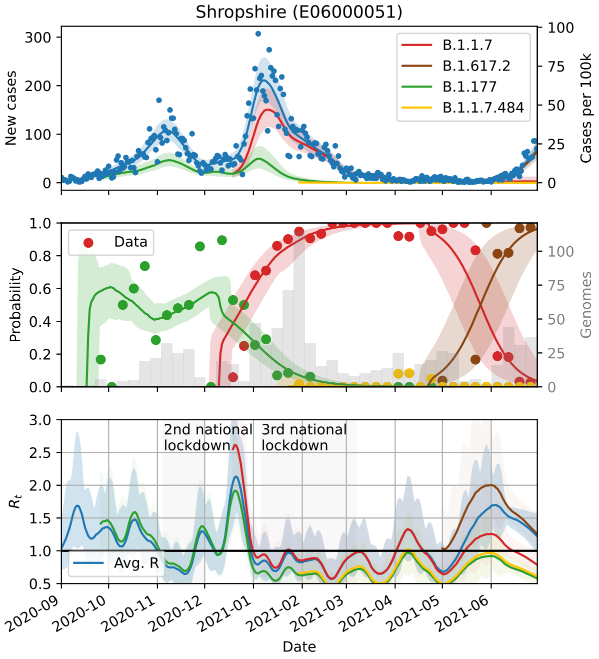
Shropshire (E06000051)
New cases
Cases per 100k
B.1.1.7
B.1.617.2
B.1.177
B.1.1.7.484
Data
Probability
Genomes
2nd national lockdown
3rd national lockdown
Avg. R
$R_t$
Date

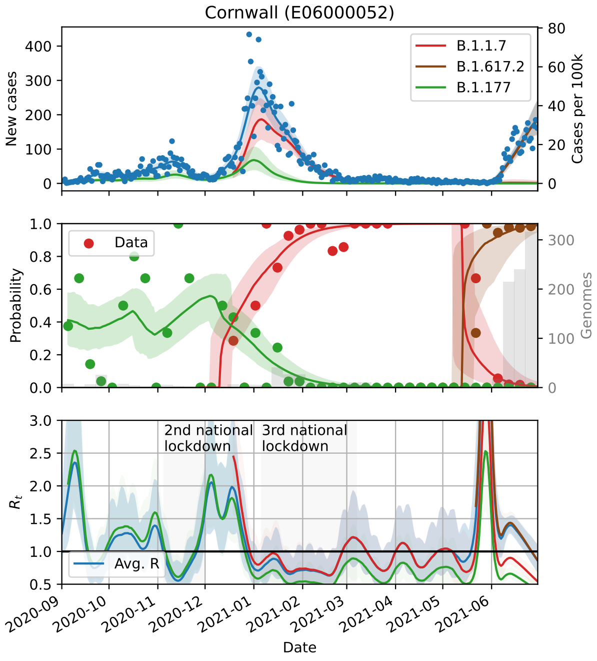
Cornwall (E06000052)
New cases
Cases per 100k
B.1.1.7
B.1.617.2
B.1.177
Probability
Data
Genomes
2nd national lockdown
3rd national lockdown
$R_t$
Avg. R
Date

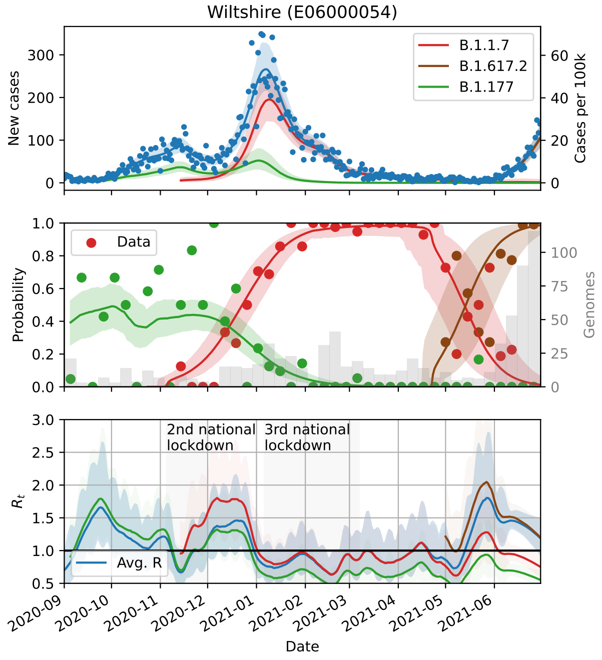
Wiltshire (E06000054)
New cases
Cases per 100k
B.1.1.7
B.1.617.2
B.1.177
Probability
Data
Genomes
2nd national lockdown
3rd national lockdown
$R_t$
Avg. R
Date

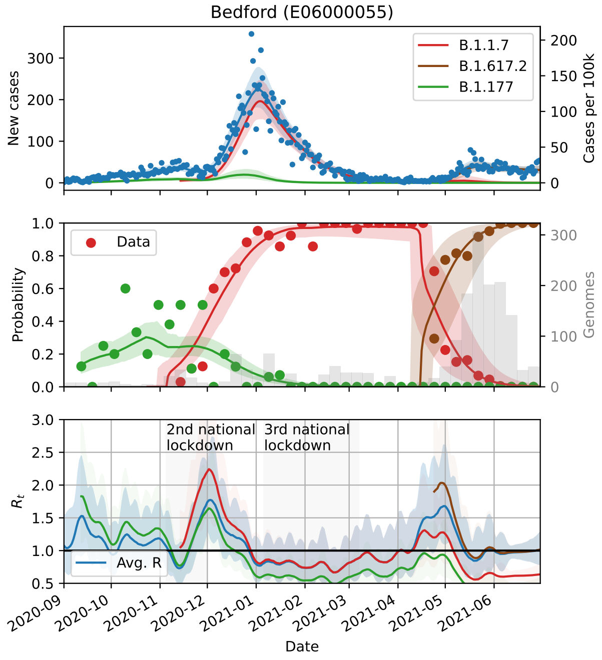
Bedford (E06000055)
New cases
Cases per 100k
B.1.1.7
B.1.617.2
B.1.177
Probability
Data
Genomes
2nd national lockdown
3rd national lockdown
$R_t$
Avg. R
Date

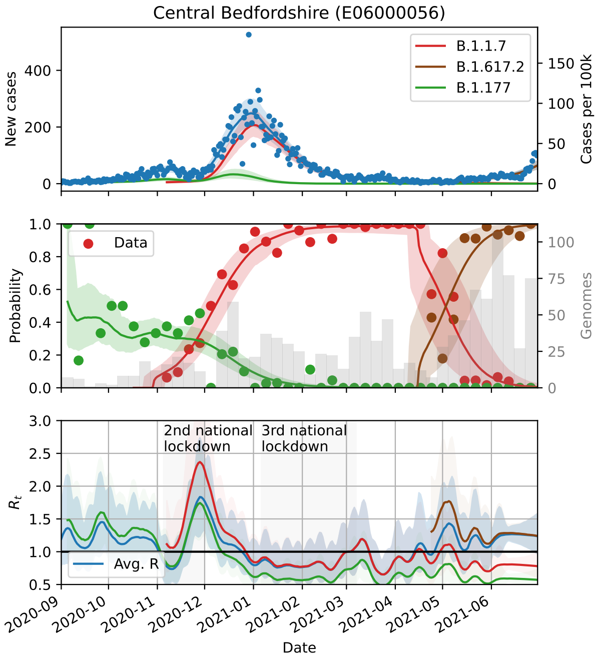
Central Bedfordshire (E06000056)
New cases
Cases per 100k
B.1.1.7
B.1.617.2
B.1.177
Probability
Genomes
Data
$R_t$
2nd national lockdown
3rd national lockdown
Avg. R
Date

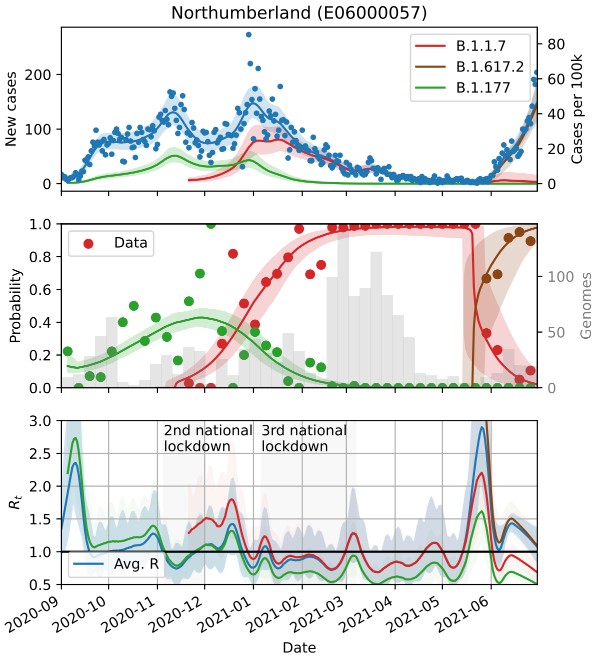
Northumberland (E06000057)
New cases
Cases per 100k
B.1.1.7
B.1.617.2
B.1.177
Probability
Genomes
Data
$R_t$
2nd national lockdown
3rd national lockdown
Avg. R
Date

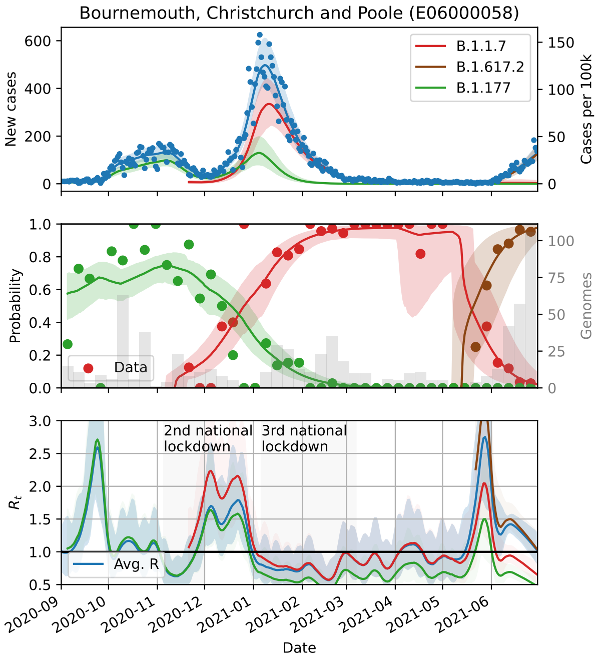
Bournemouth, Christchurch and Poole (E06000058)
New cases
Cases per 100k
B.1.1.7
B.1.617.2
B.1.177
Probability
Genomes
Data
$R_t$
2nd national lockdown
3rd national lockdown
Avg. R
Date

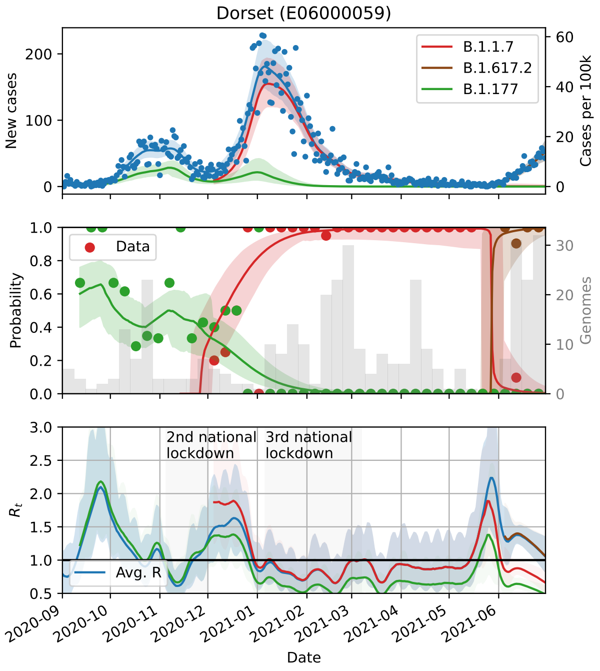
Dorset (E06000059)
New cases
Cases per 100k
B.1.1.7
B.1.617.2
B.1.177
Probability
Genomes
Data
$R_t$
2nd national lockdown
3rd national lockdown
Avg. R
Date

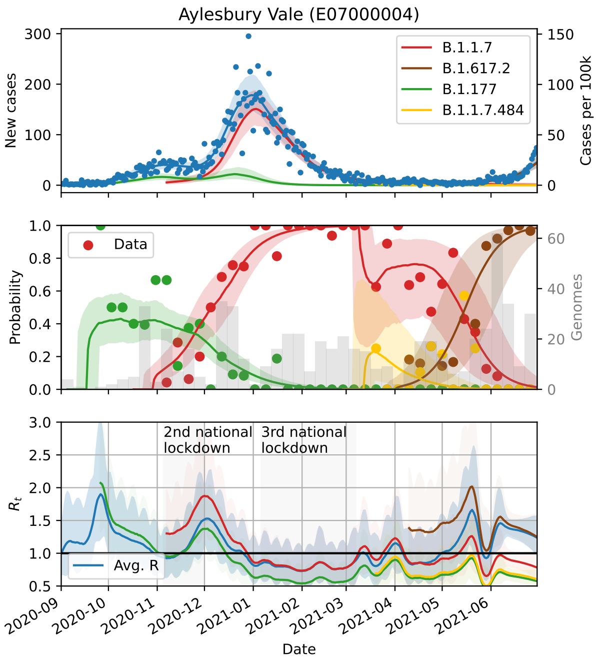
Aylesbury Vale (E07000004)
New cases
Cases per 100k
B.1.1.7
B.1.617.2
B.1.177
B.1.1.7.484
Probability
Genomes
Data
$R_t$
2nd national lockdown
3rd national lockdown
Avg. R
Date

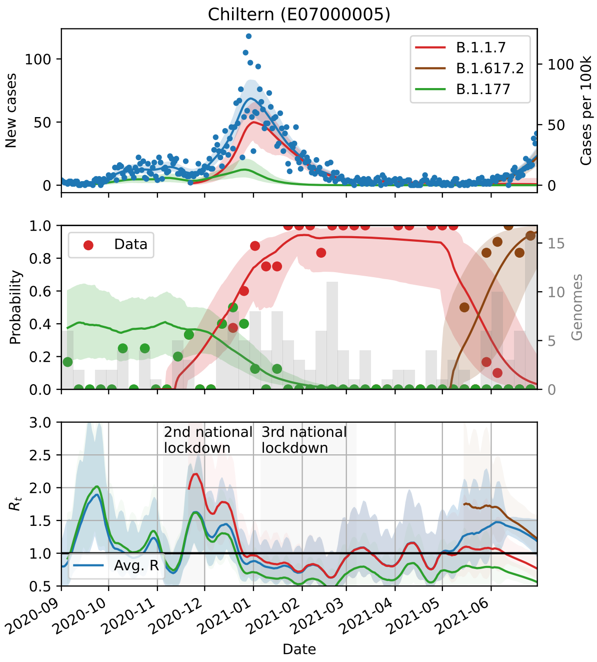
Chiltern (E07000005)
New cases
Cases per 100k
B.1.1.7
B.1.617.2
B.1.177
Probability
Genomes
Data
$R_t$
2nd national lockdown
3rd national lockdown
Avg. R
Date

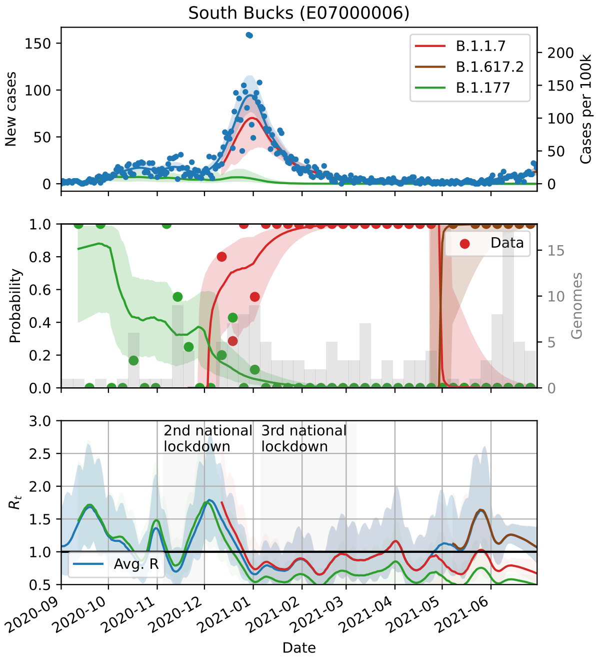
South Bucks (E07000006)
New cases
Cases per 100k
B.1.1.7
B.1.617.2
B.1.177
Probability
Genomes
Data
$R_t$
2nd national lockdown
3rd national lockdown
Avg. R
Date

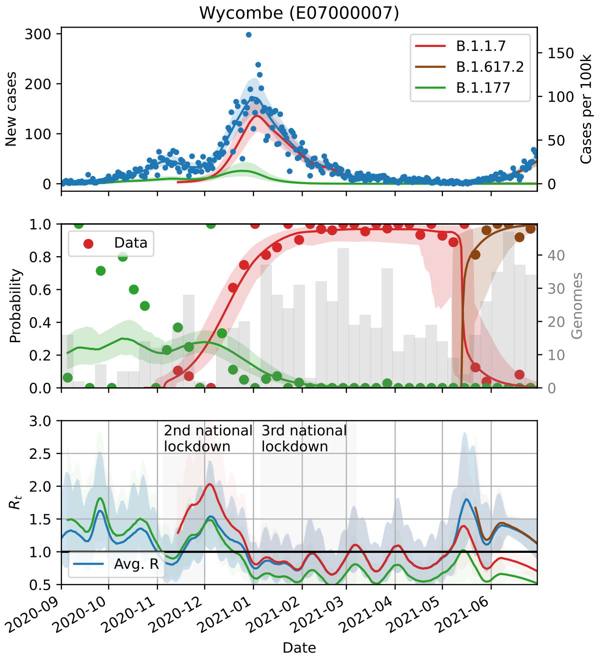
Wycombe (E07000007)
New cases
Cases per 100k
B.1.1.7
B.1.617.2
B.1.177
Probability
Genomes
Data
$R_t$
2nd national lockdown
3rd national lockdown
Avg. R
Date

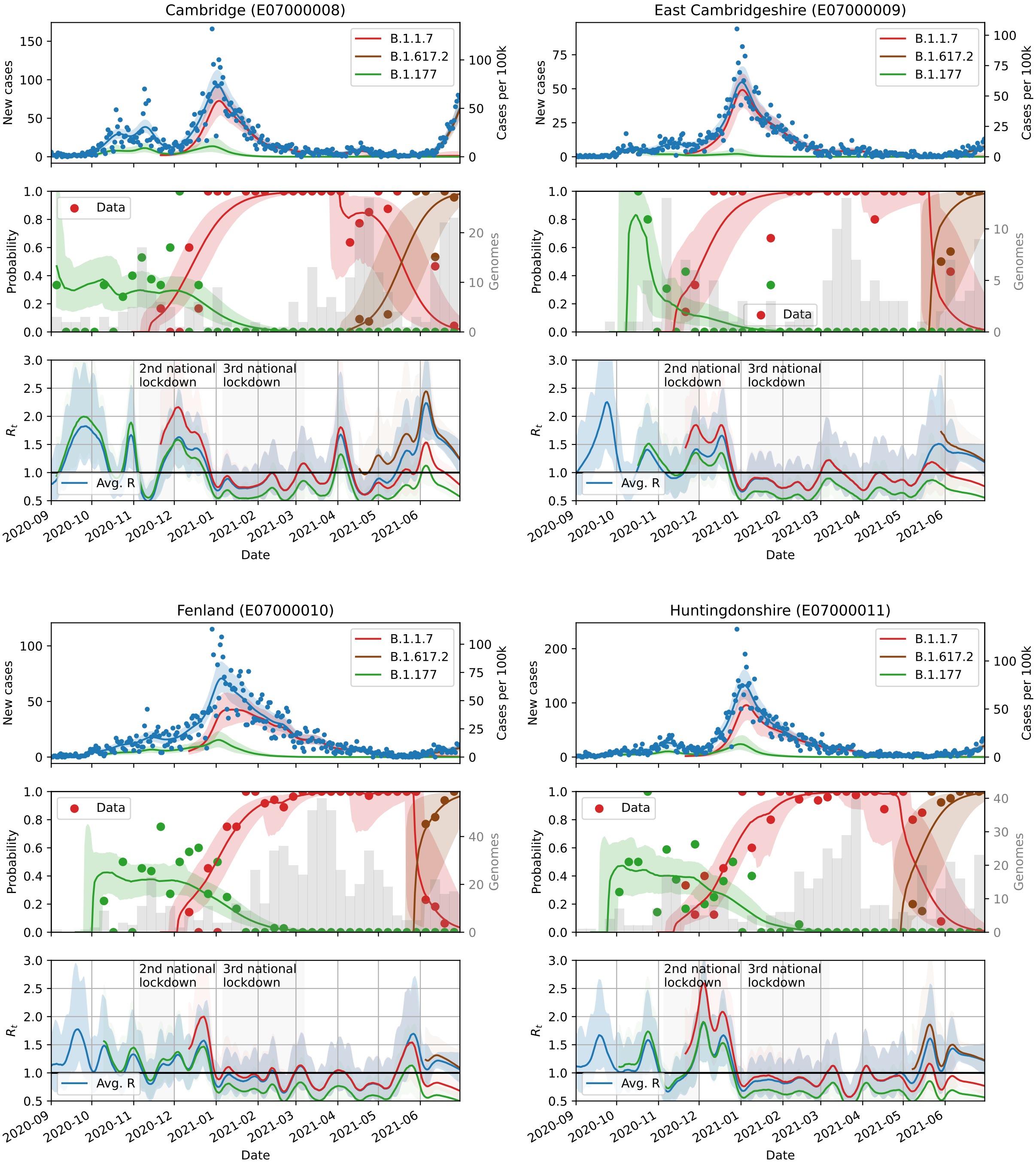
Cambridge (E07000008)
East Cambridgeshire (E07000009)
Fenland (E07000010)
Huntingdonshire (E07000011)
B.1.1.7
B.1.617.2
B.1.177
Data
Avg. R
New cases
Cases per 100k
Probability
Genomes
$R_t$
2nd national lockdown
3rd national lockdown
Date
2020-09
2020-10
2020-11
2020-12
2021-01
2021-02
2021-03
2021-04
2021-05
2021-06

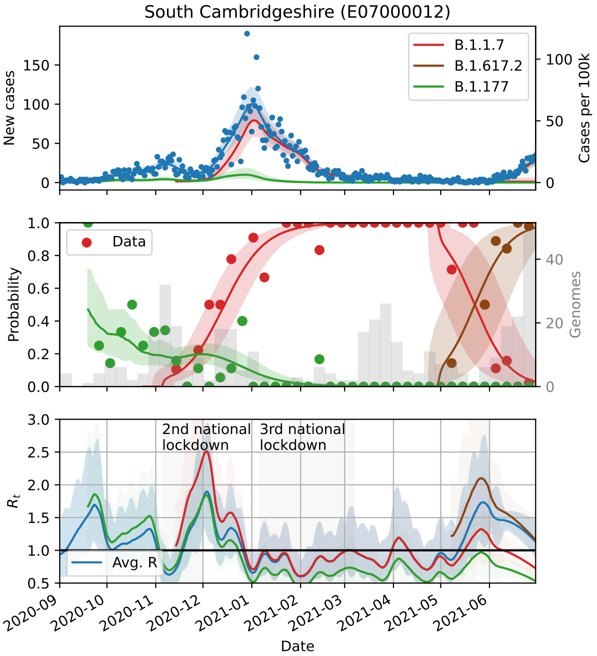
South Cambridgeshire (E07000012)
B.1.1.7
B.1.617.2
B.1.177
New cases
Cases per 100k
Data
Probability
Genomes
2nd national lockdown
3rd national lockdown
$R_t$
Avg. R
Date

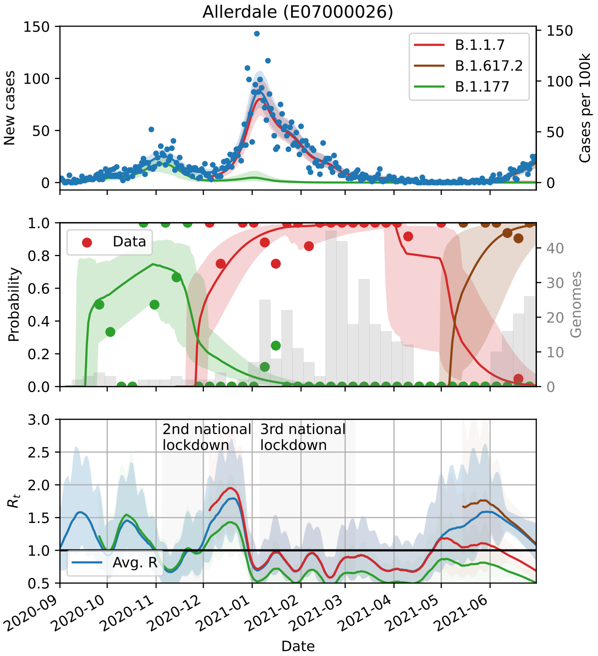
Allerdale (E07000026)
B.1.1.7
B.1.617.2
B.1.177
New cases
Cases per 100k
Data
Probability
Genomes
2nd national lockdown
3rd national lockdown
$R_t$
Avg. R
Date

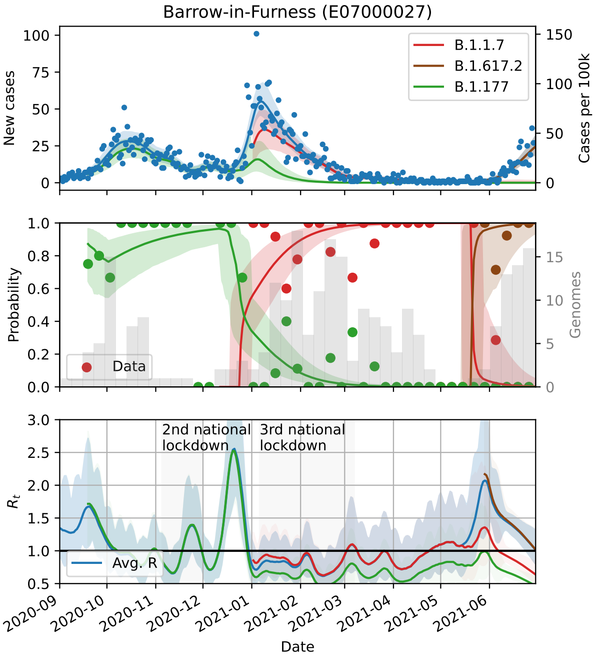
Barrow-in-Furness (E07000027)
B.1.1.7
B.1.617.2
B.1.177
New cases
Cases per 100k
Data
Probability
Genomes
2nd national lockdown
3rd national lockdown
$R_t$
Avg. R
Date

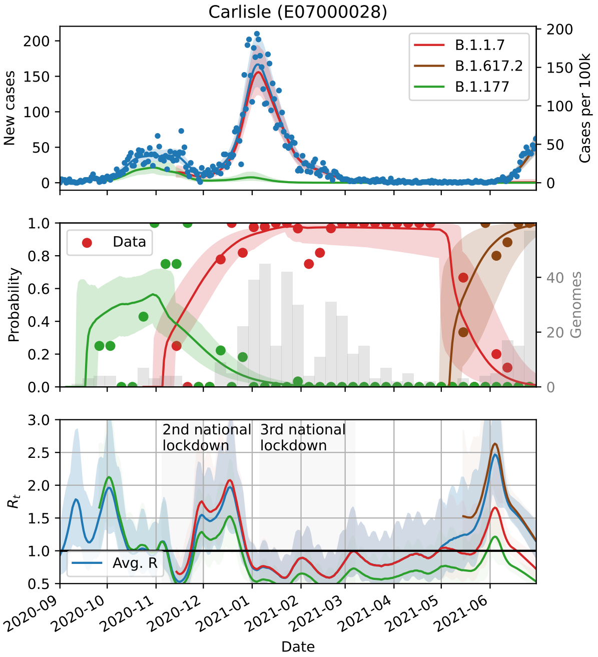
Carlisle (E07000028)
B.1.1.7
B.1.617.2
B.1.177
New cases
Cases per 100k
Data
Probability
Genomes
2nd national lockdown
3rd national lockdown
$R_t$
Avg. R
Date

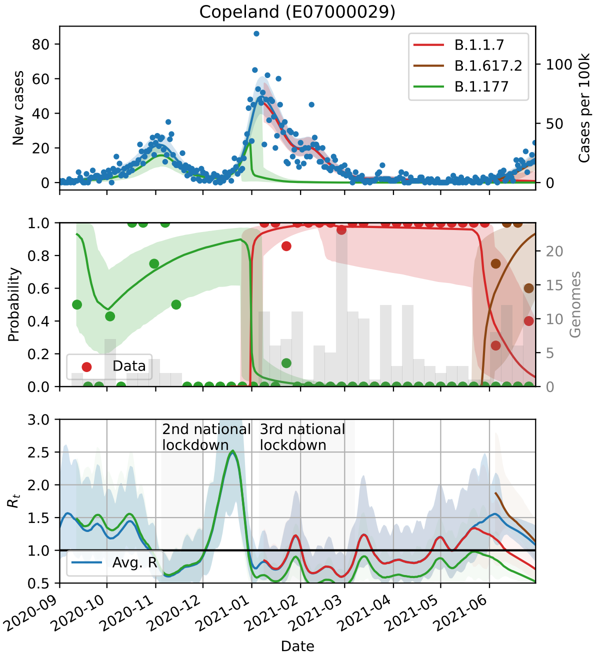
Copeland (E07000029)
B.1.1.7
B.1.617.2
B.1.177
New cases
Cases per 100k
Probability
Genomes
Data
2nd national lockdown
3rd national lockdown
Avg. R
$R_t$
Date

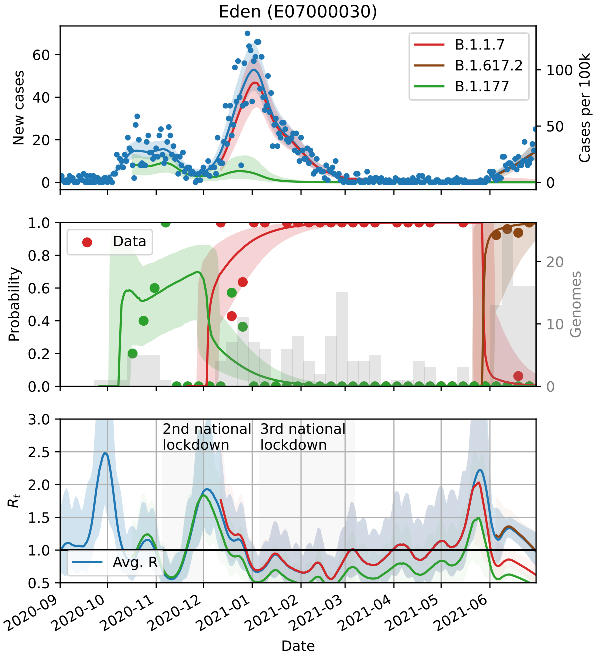
Eden (E07000030)
B.1.1.7
B.1.617.2
B.1.177
New cases
Cases per 100k
Probability
Genomes
Data
2nd national lockdown
3rd national lockdown
Avg. R
$R_t$
Date

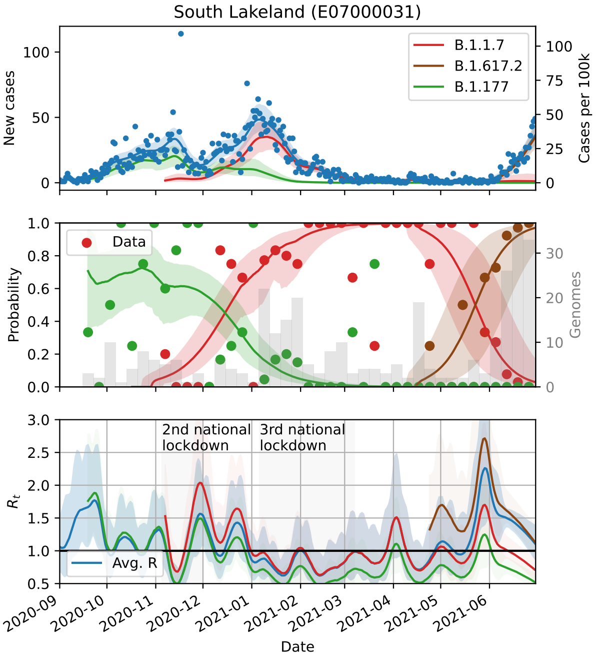
South Lakeland (E07000031)
B.1.1.7
B.1.617.2
B.1.177
New cases
Cases per 100k
Probability
Genomes
Data
2nd national lockdown
3rd national lockdown
Avg. R
$R_t$
Date

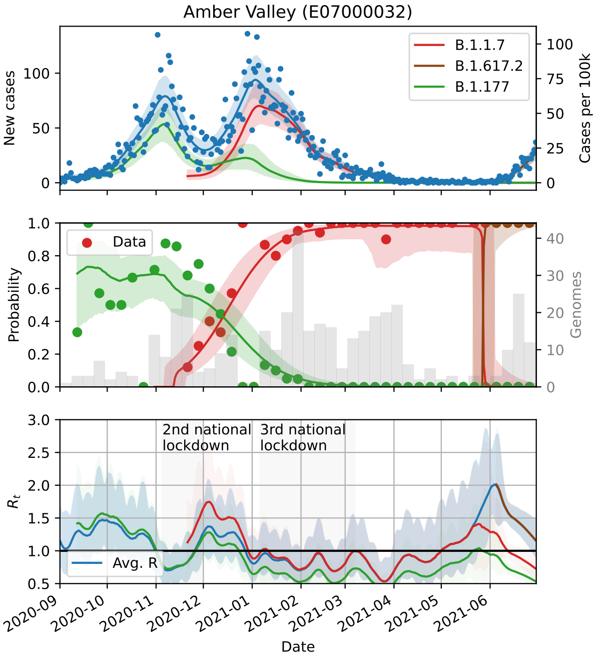
Amber Valley (E07000032)
B.1.1.7
B.1.617.2
B.1.177
New cases
Cases per 100k
Probability
Genomes
Data
2nd national lockdown
3rd national lockdown
Avg. R
$R_t$
Date

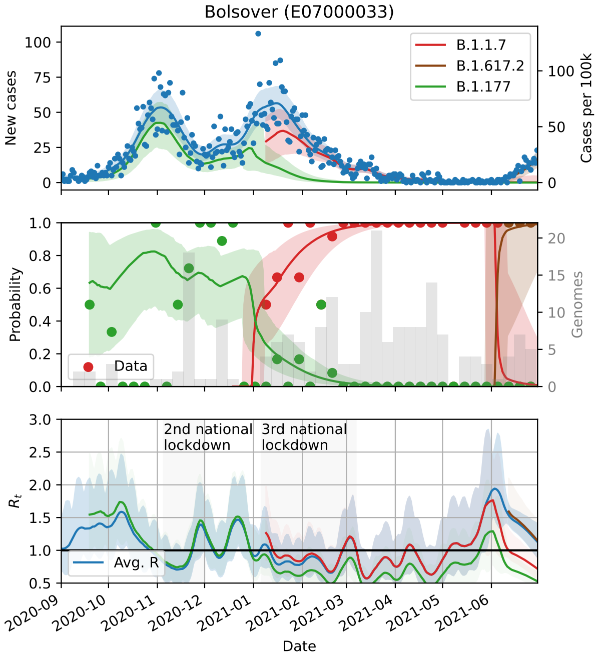
Bolsover (E07000033)
B.1.1.7
B.1.617.2
B.1.177
New cases
Cases per 100k
Probability
Genomes
Data
2nd national lockdown
3rd national lockdown
$R_t$
Avg. R
Date

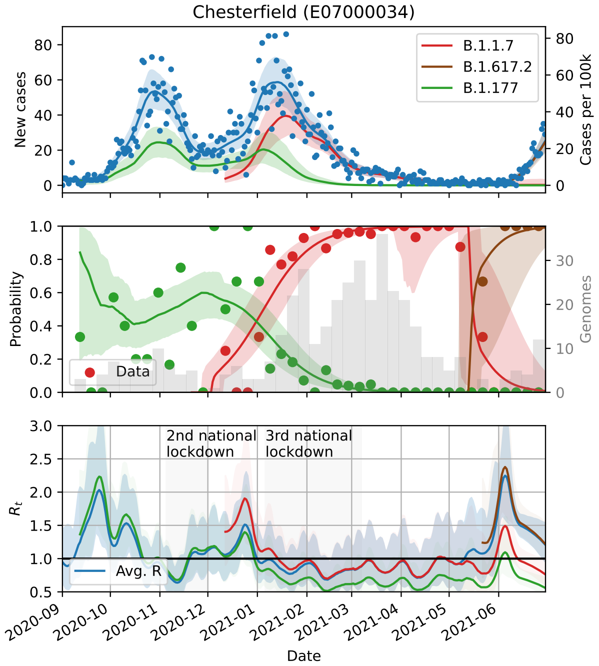
Chesterfield (E07000034)
B.1.1.7
B.1.617.2
B.1.177
New cases
Cases per 100k
Probability
Genomes
Data
2nd national lockdown
3rd national lockdown
$R_t$
Avg. R
Date

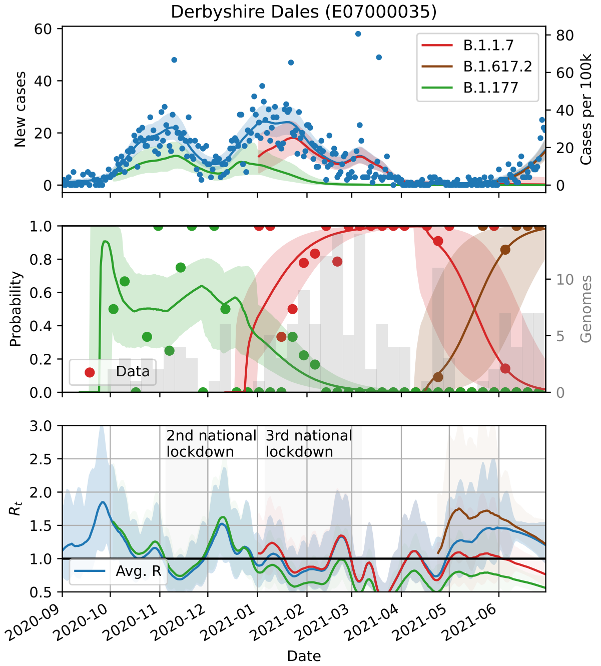
Derbyshire Dales (E07000035)
B.1.1.7
B.1.617.2
B.1.177
New cases
Cases per 100k
Probability
Genomes
Data
2nd national lockdown
3rd national lockdown
$R_t$
Avg. R
Date

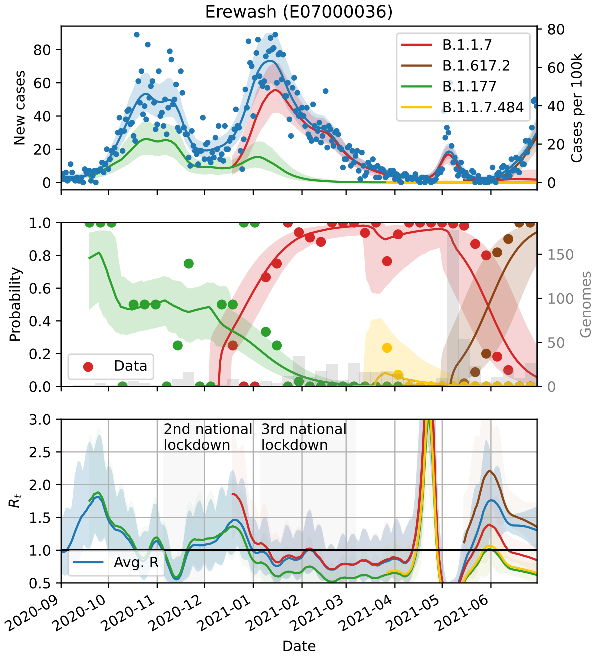
Erewash (E07000036)
B.1.1.7
B.1.617.2
B.1.177
B.1.1.7.484
New cases
Cases per 100k
Probability
Genomes
Data
2nd national lockdown
3rd national lockdown
$R_t$
Avg. R
Date

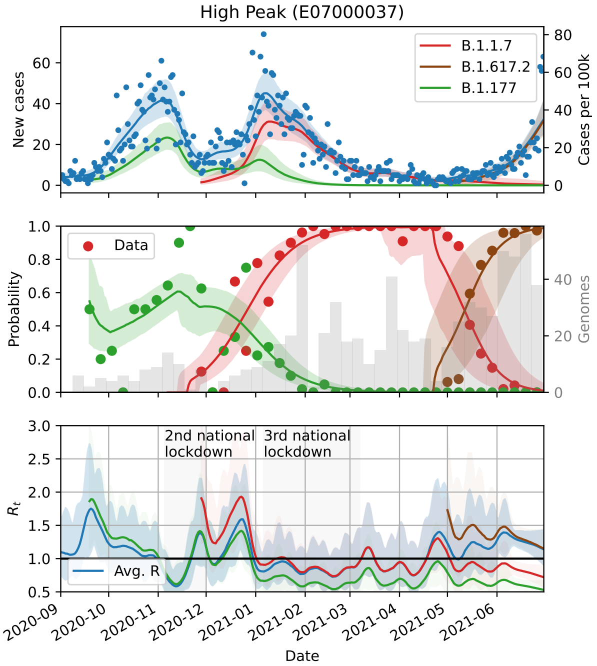
High Peak (E07000037)
New cases
Cases per 100k
B.1.1.7
B.1.617.2
B.1.177
Data
Probability
Genomes
2nd national lockdown
3rd national lockdown
Avg. R
$R_t$
Date

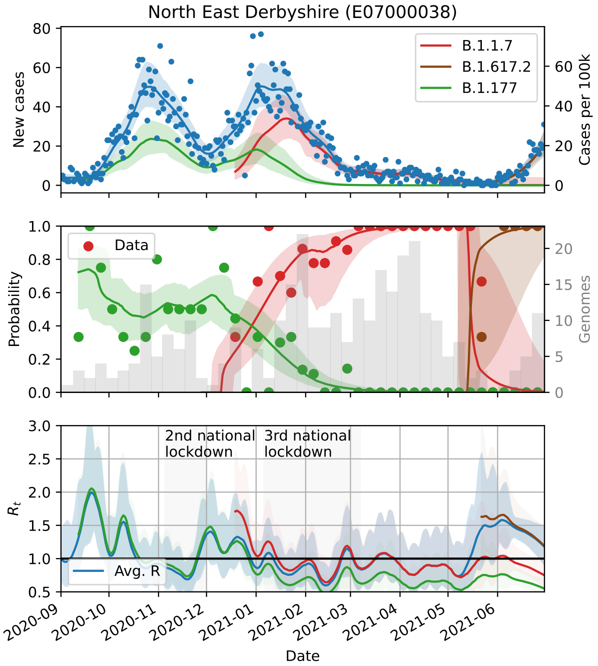
North East Derbyshire (E07000038)
New cases
Cases per 100k
B.1.1.7
B.1.617.2
B.1.177
Data
Probability
Genomes
2nd national lockdown
3rd national lockdown
Avg. R
$R_t$
Date

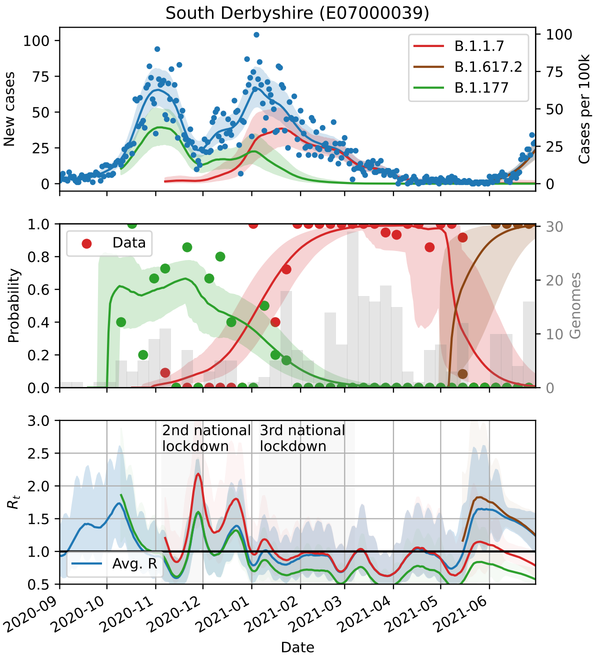
South Derbyshire (E07000039)
New cases
Cases per 100k
B.1.1.7
B.1.617.2
B.1.177
Data
Probability
Genomes
2nd national lockdown
3rd national lockdown
Avg. R
$R_t$
Date

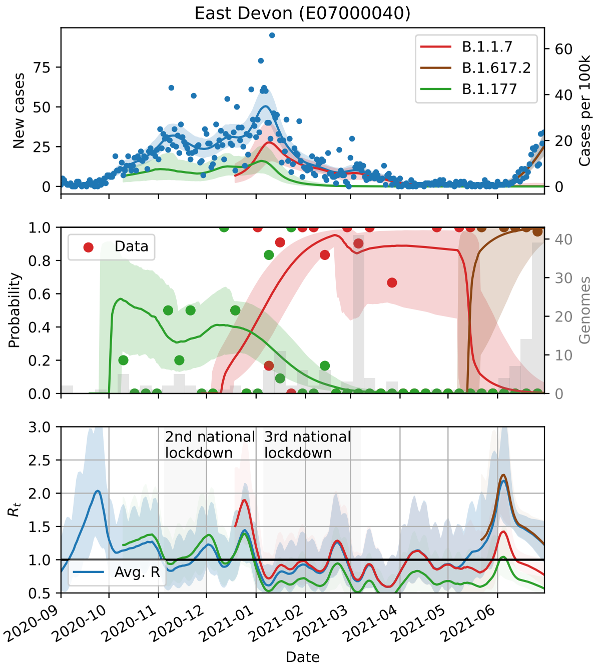
East Devon (E07000040)
New cases
Cases per 100k
B.1.1.7
B.1.617.2
B.1.177
Data
Probability
Genomes
2nd national lockdown
3rd national lockdown
Avg. R
$R_t$
Date

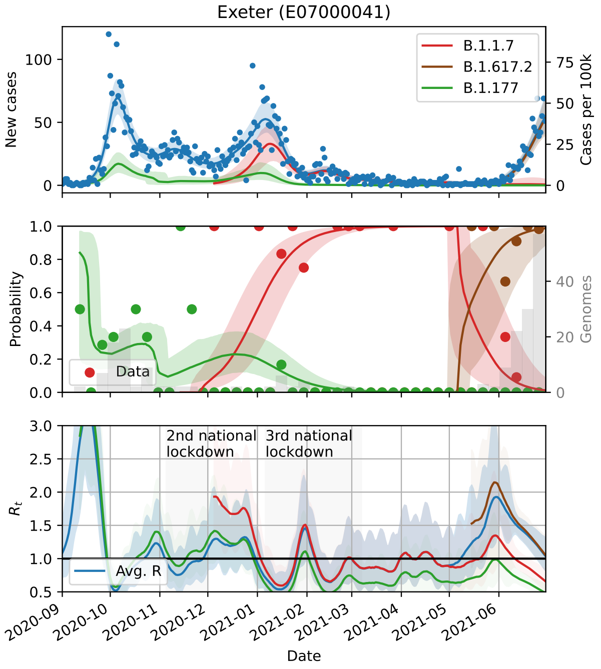
Exeter (E07000041)
B.1.1.7
B.1.617.2
B.1.177
New cases
Cases per 100k
Probability
Genomes
Data
2nd national lockdown
3rd national lockdown
$R_t$
Avg. R
Date

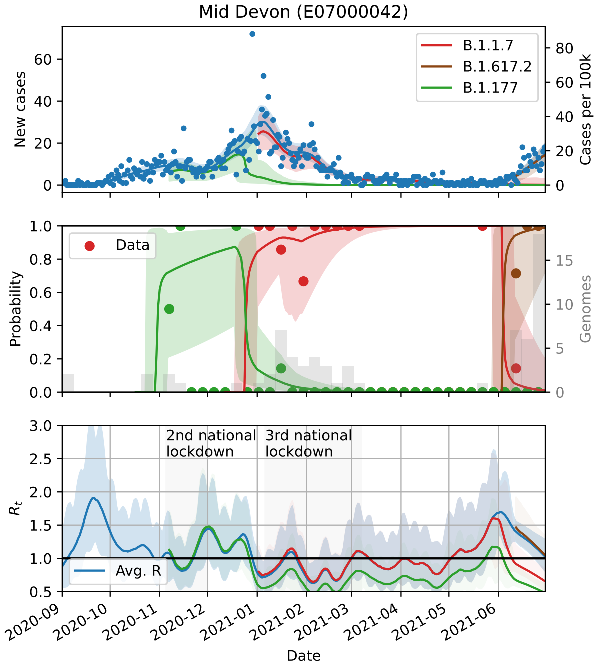
Mid Devon (E07000042)
B.1.1.7
B.1.617.2
B.1.177
New cases
Cases per 100k
Probability
Genomes
Data
2nd national lockdown
3rd national lockdown
$R_t$
Avg. R
Date

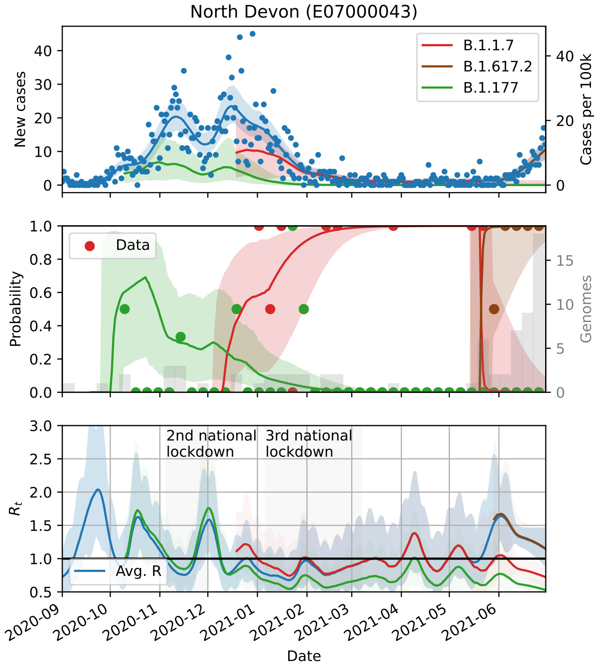
North Devon (E07000043)
B.1.1.7
B.1.617.2
B.1.177
New cases
Cases per 100k
Probability
Genomes
Data
2nd national lockdown
3rd national lockdown
$R_t$
Avg. R
Date

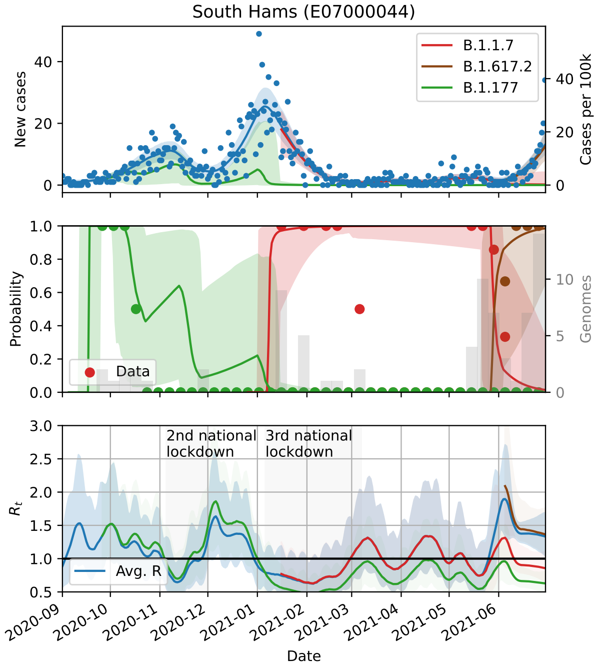
South Hams (E07000044)
B.1.1.7
B.1.617.2
B.1.177
New cases
Cases per 100k
Probability
Genomes
Data
2nd national lockdown
3rd national lockdown
$R_t$
Avg. R
Date

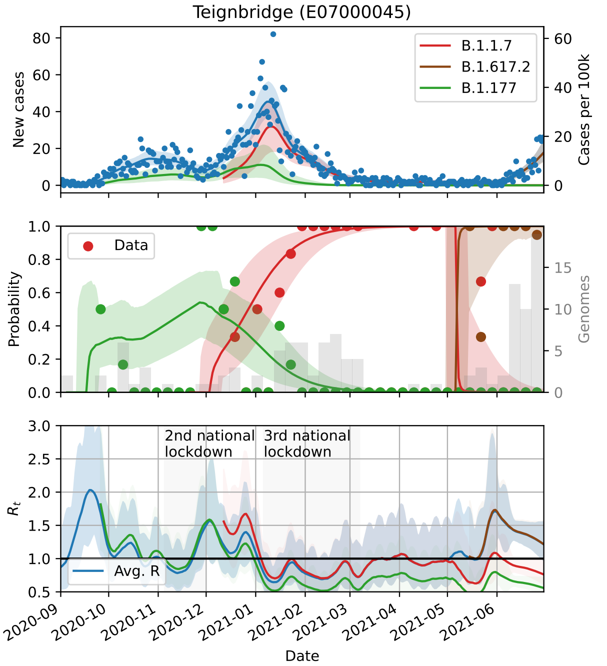
Teignbridge (E07000045)
B.1.1.7
B.1.617.2
B.1.177
New cases
Cases per 100k
Data
Probability
Genomes
2nd national lockdown
3rd national lockdown
Avg. R
$R_t$
Date

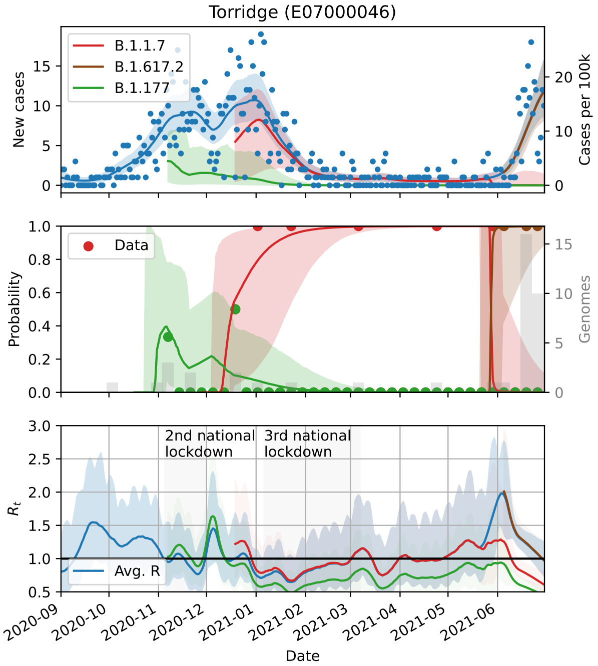
Torridge (E07000046)
B.1.1.7
B.1.617.2
B.1.177
New cases
Cases per 100k
Data
Probability
Genomes
2nd national lockdown
3rd national lockdown
Avg. R
$R_t$
Date

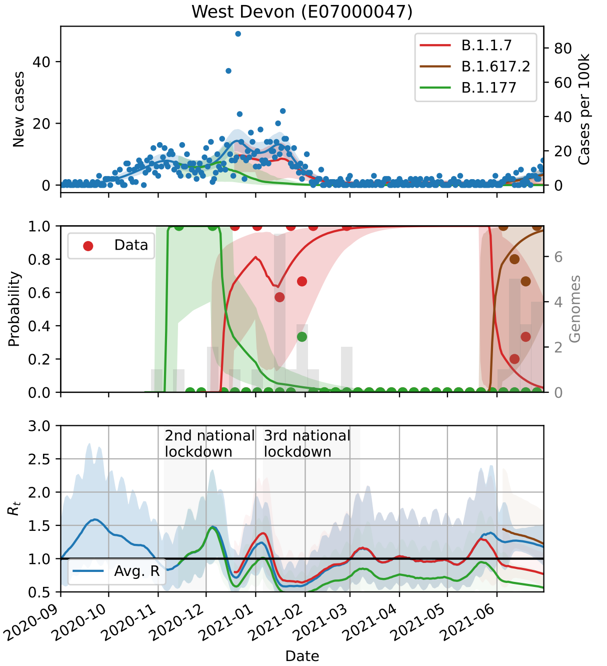
West Devon (E07000047)
B.1.1.7
B.1.617.2
B.1.177
New cases
Cases per 100k
Data
Probability
Genomes
2nd national lockdown
3rd national lockdown
Avg. R
$R_t$
Date

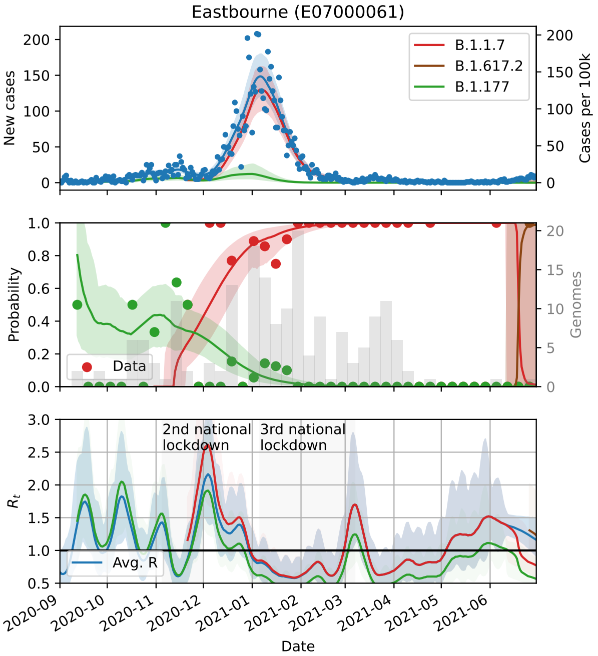
Eastbourne (E07000061)
B.1.1.7
B.1.617.2
B.1.177
New cases
Cases per 100k
Data
Probability
Genomes
2nd national lockdown
3rd national lockdown
Avg. R
$R_t$
Date

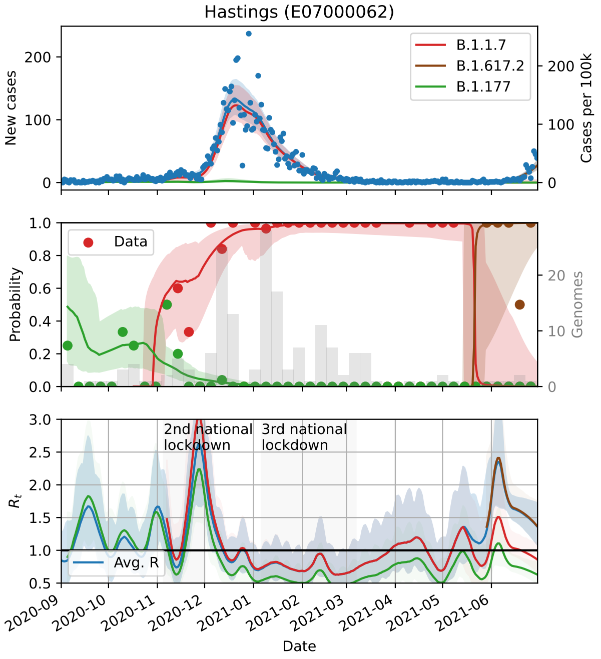
Hastings (E07000062)
New cases
Cases per 100k
B.1.1.7
B.1.617.2
B.1.177
Probability
Genomes
Data
$R_t$
2nd national lockdown
3rd national lockdown
Avg. R
Date

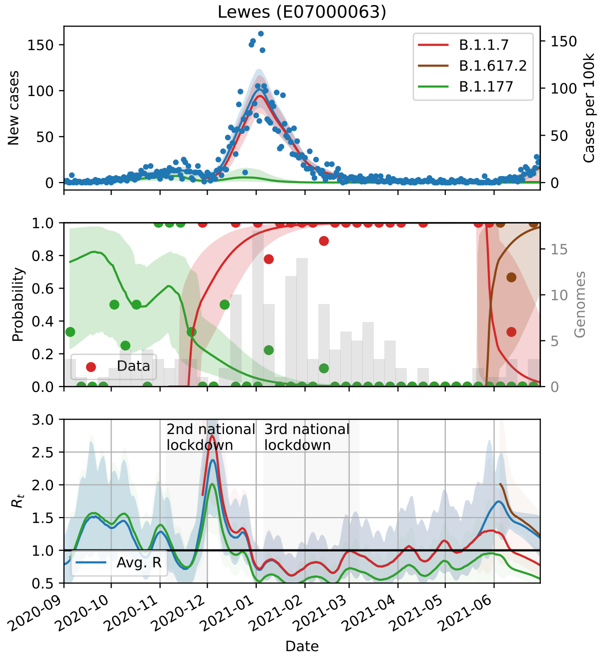
Lewes (E07000063)
New cases
Cases per 100k
B.1.1.7
B.1.617.2
B.1.177
Probability
Genomes
Data
$R_t$
2nd national lockdown
3rd national lockdown
Avg. R
Date

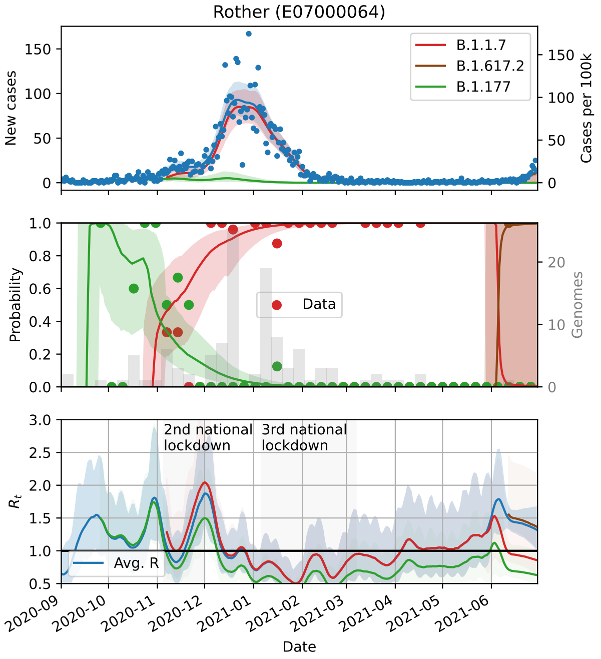
Rother (E07000064)
New cases
Cases per 100k
B.1.1.7
B.1.617.2
B.1.177
Probability
Genomes
Data
$R_t$
2nd national lockdown
3rd national lockdown
Avg. R
Date

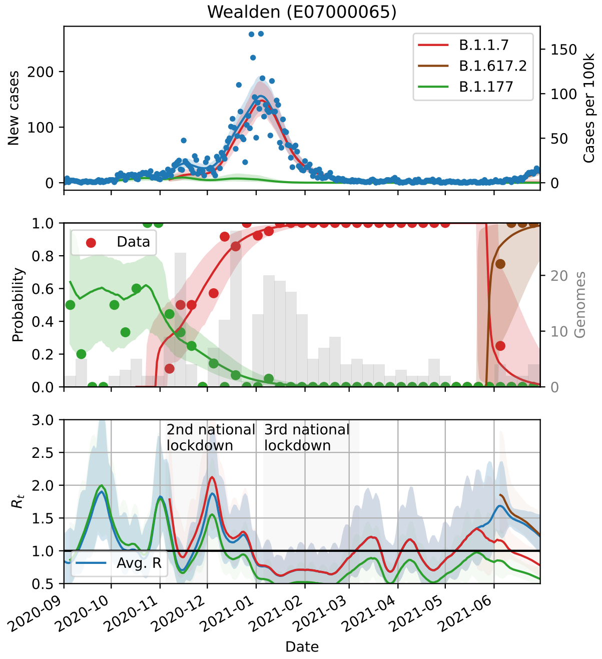
Wealden (E07000065)
New cases
Cases per 100k
B.1.1.7
B.1.617.2
B.1.177
Probability
Genomes
Data
$R_t$
2nd national lockdown
3rd national lockdown
Avg. R
Date

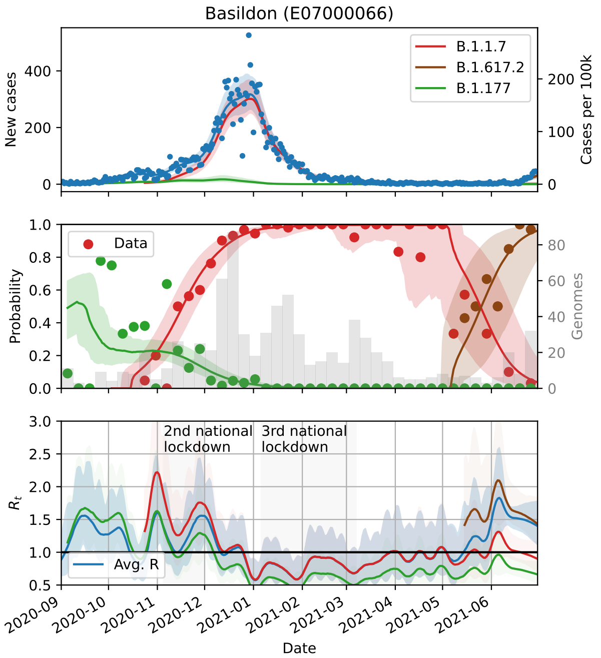
Basildon (E07000066)
New cases
Cases per 100k
B.1.1.7
B.1.617.2
B.1.177
Data
Probability
Genomes
2nd national lockdown
3rd national lockdown
$R_t$
Avg. R
2020-09
2020-10
2020-11
2020-12
2021-01
2021-02
2021-03
2021-04
2021-05
2021-06
Date

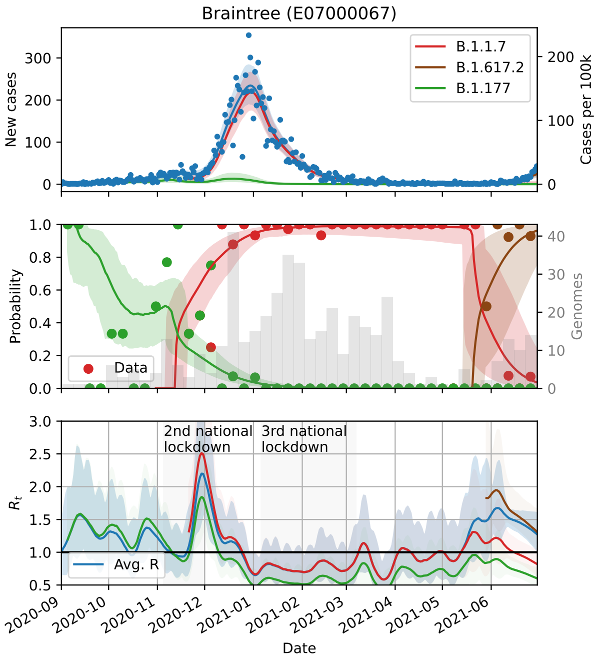
Braintree (E07000067)
New cases
Cases per 100k
B.1.1.7
B.1.617.2
B.1.177
Data
Probability
Genomes
2nd national lockdown
3rd national lockdown
$R_t$
Avg. R
2020-09
2020-10
2020-11
2020-12
2021-01
2021-02
2021-03
2021-04
2021-05
2021-06
Date

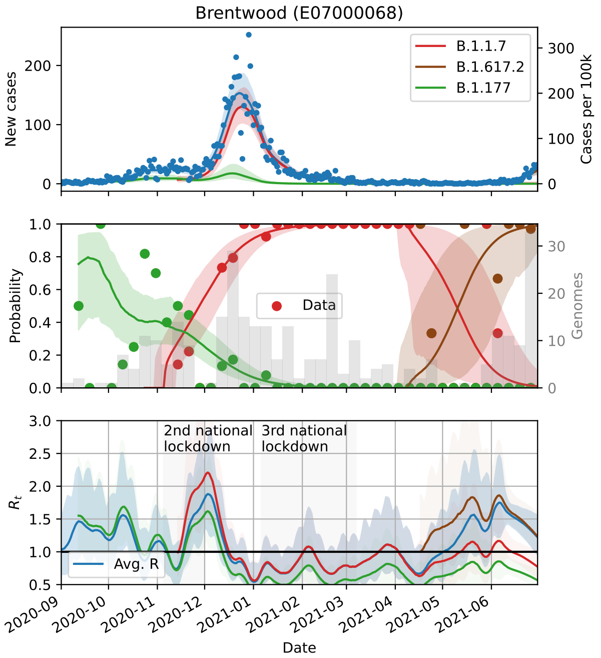
Brentwood (E07000068)
New cases
Cases per 100k
B.1.1.7
B.1.617.2
B.1.177
Data
Probability
Genomes
2nd national lockdown
3rd national lockdown
$R_t$
Avg. R
2020-09
2020-10
2020-11
2020-12
2021-01
2021-02
2021-03
2021-04
2021-05
2021-06
Date

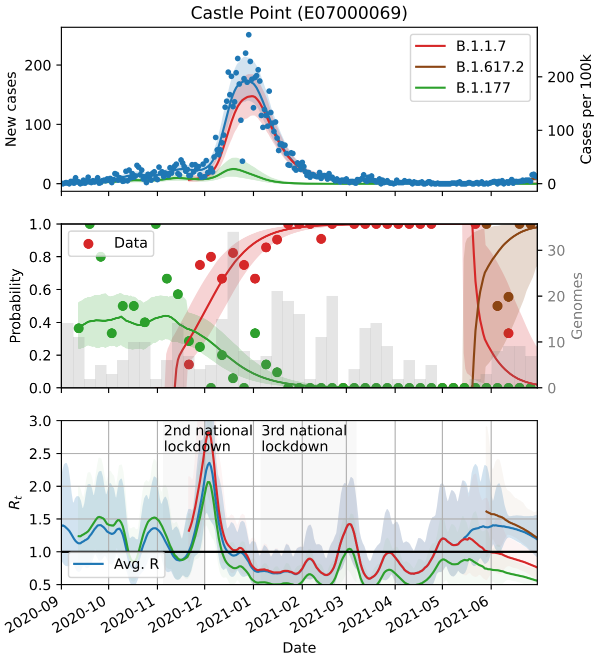
Castle Point (E07000069)
New cases
Cases per 100k
B.1.1.7
B.1.617.2
B.1.177
Data
Probability
Genomes
2nd national lockdown
3rd national lockdown
$R_t$
Avg. R
2020-09
2020-10
2020-11
2020-12
2021-01
2021-02
2021-03
2021-04
2021-05
2021-06
Date

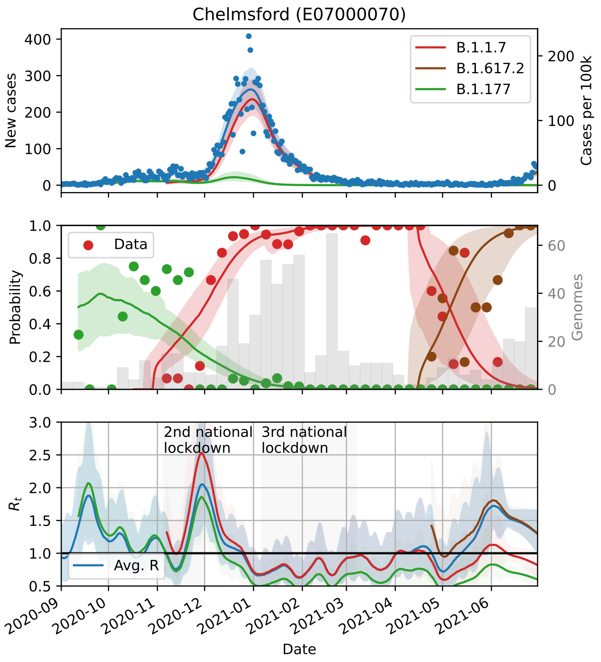
Chelmsford (E07000070)
B.1.1.7
B.1.617.2
B.1.177
New cases
Cases per 100k
Data
Probability
Genomes
2nd national lockdown
3rd national lockdown
Avg. R
$R_t$
Date

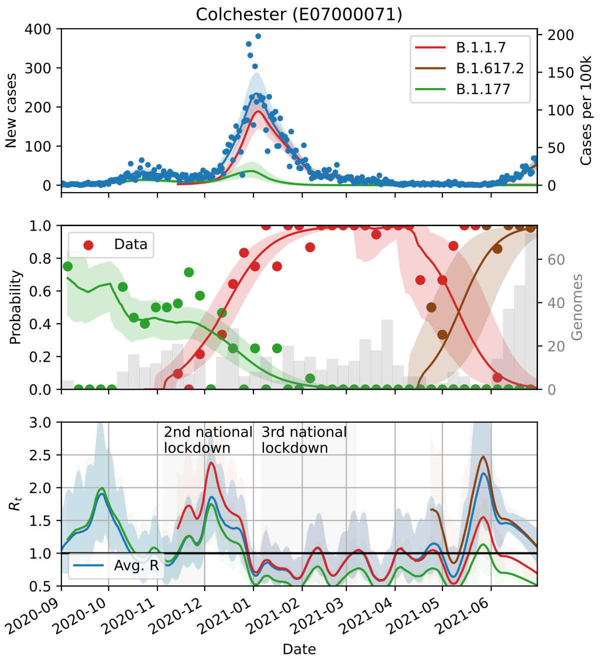
Colchester (E07000071)
B.1.1.7
B.1.617.2
B.1.177
New cases
Cases per 100k
Data
Probability
Genomes
2nd national lockdown
3rd national lockdown
Avg. R
$R_t$
Date

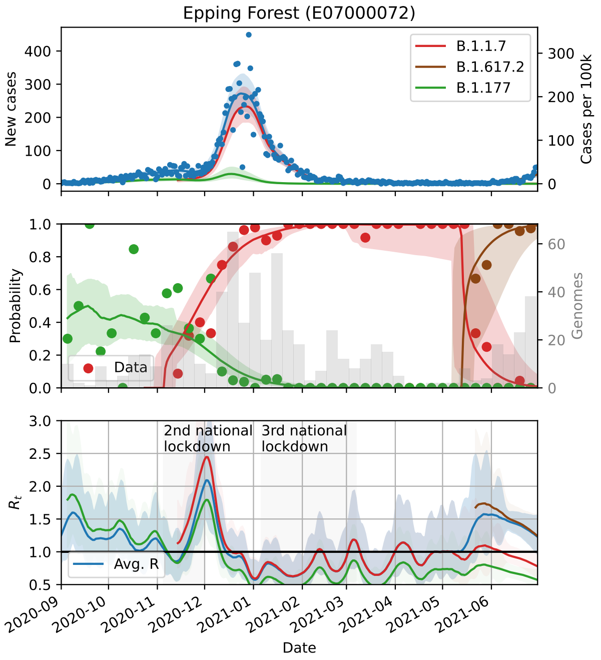
Epping Forest (E07000072)
B.1.1.7
B.1.617.2
B.1.177
New cases
Cases per 100k
Data
Probability
Genomes
2nd national lockdown
3rd national lockdown
Avg. R
$R_t$
Date

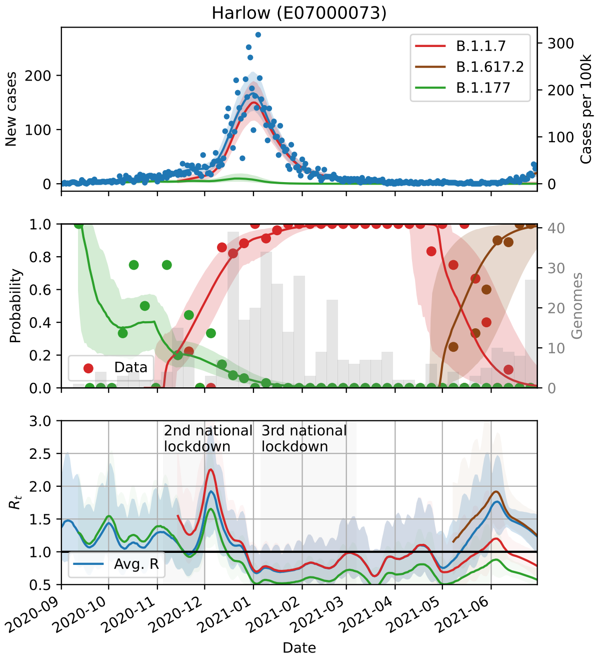
Harlow (E07000073)
B.1.1.7
B.1.617.2
B.1.177
New cases
Cases per 100k
Data
Probability
Genomes
2nd national lockdown
3rd national lockdown
Avg. R
$R_t$
Date

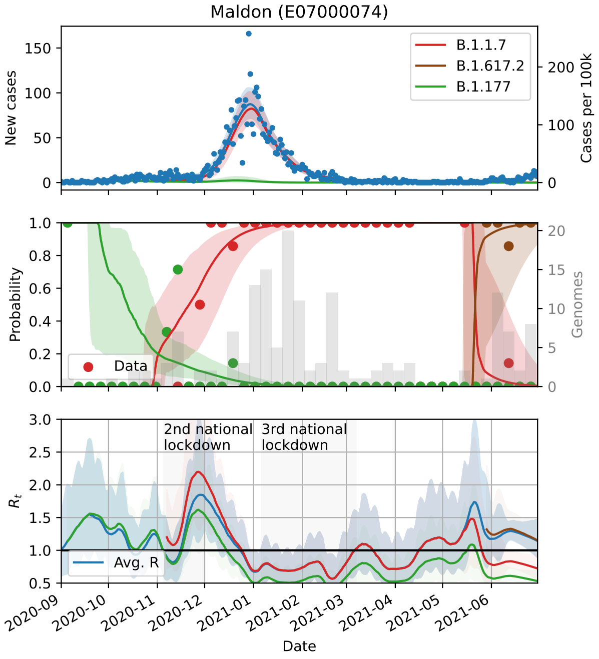
Maldon (E07000074)
B.1.1.7
B.1.617.2
B.1.177
New cases
Cases per 100k
Data
Probability
Genomes
2nd national lockdown
3rd national lockdown
Avg. R
$R_t$
Date

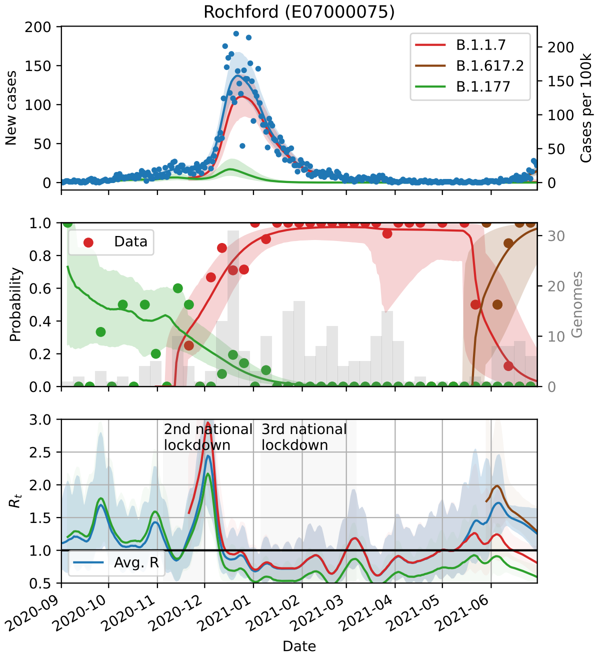
Rochford (E07000075)
B.1.1.7
B.1.617.2
B.1.177
New cases
Cases per 100k
Data
Probability
Genomes
2nd national lockdown
3rd national lockdown
Avg. R
$R_t$
Date

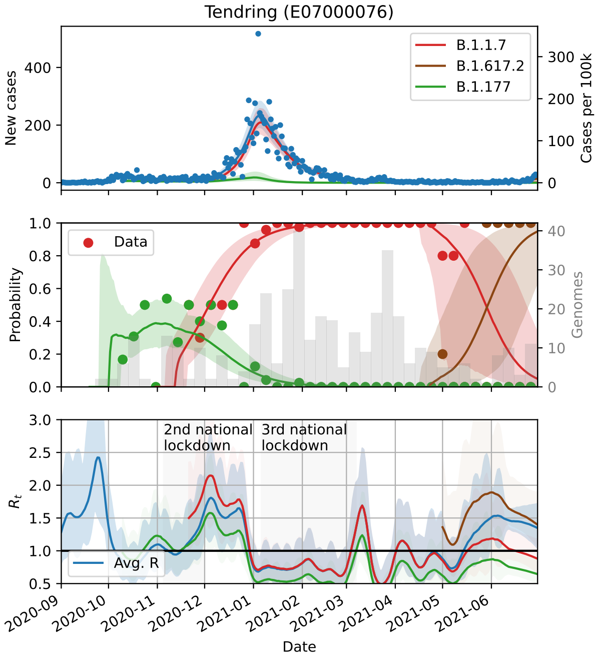
Tendring (E07000076)
B.1.1.7
B.1.617.2
B.1.177
New cases
Cases per 100k
Data
Probability
Genomes
2nd national lockdown
3rd national lockdown
Avg. R
$R_t$
Date

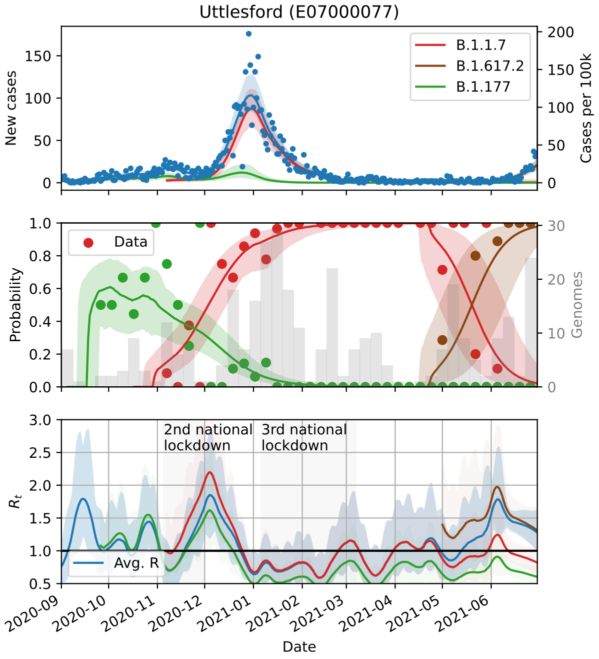
Uttlesford (E07000077)
B.1.1.7
B.1.617.2
B.1.177
New cases
Cases per 100k
Data
Probability
Genomes
2nd national lockdown
3rd national lockdown
Avg. R
$R_t$
Date

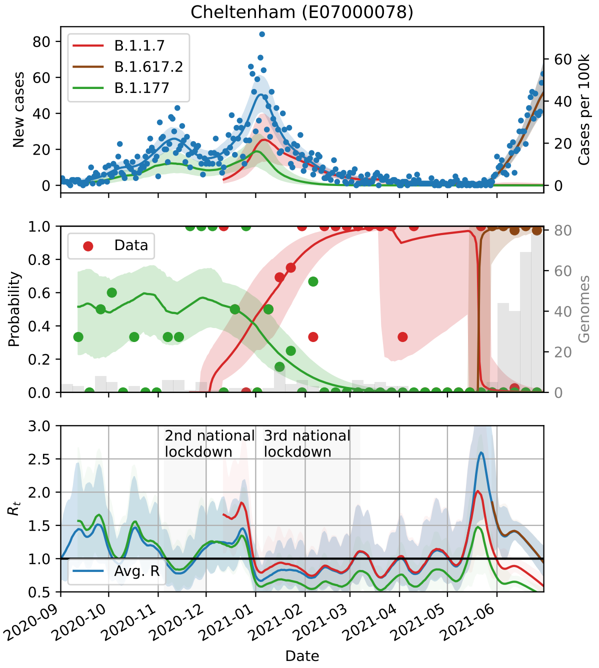
Cheltenham (E07000078)
B.1.1.7
B.1.617.2
B.1.177
New cases
Cases per 100k
Data
Probability
Genomes
2nd national lockdown
3rd national lockdown
$R_t$
Avg. R
Date

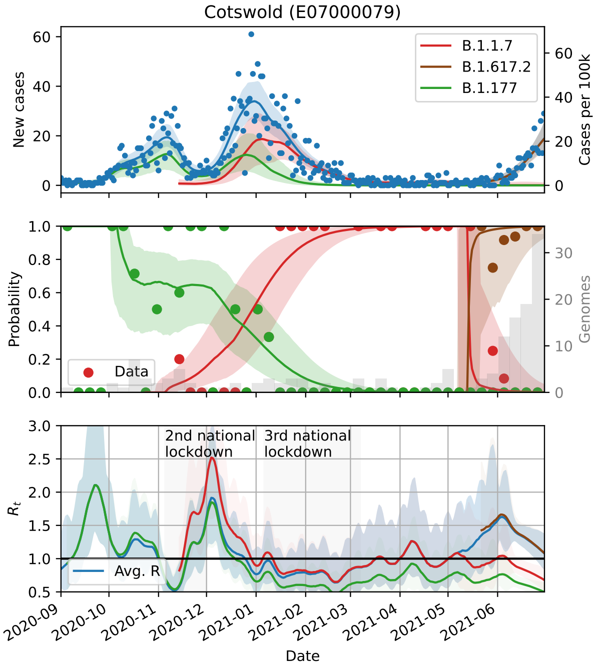
Cotswold (E07000079)
B.1.1.7
B.1.617.2
B.1.177
New cases
Cases per 100k
Data
Probability
Genomes
2nd national lockdown
3rd national lockdown
$R_t$
Avg. R
Date

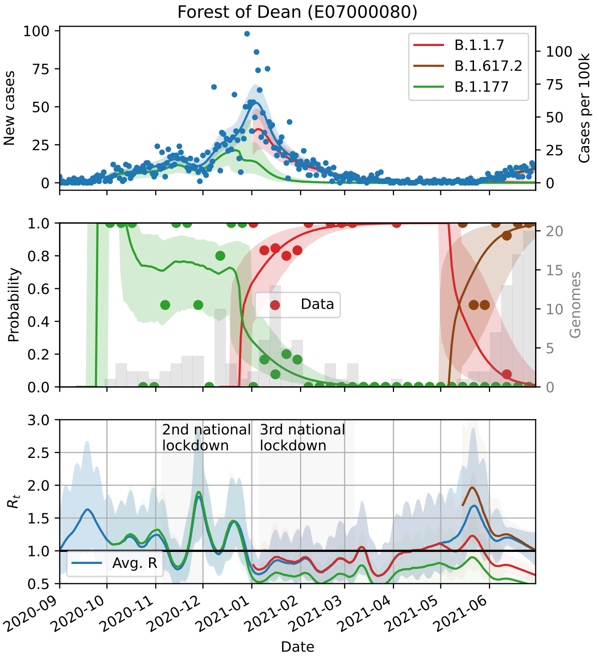
Forest of Dean (E07000080)
B.1.1.7
B.1.617.2
B.1.177
New cases
Cases per 100k
Data
Probability
Genomes
2nd national lockdown
3rd national lockdown
$R_t$
Avg. R
Date

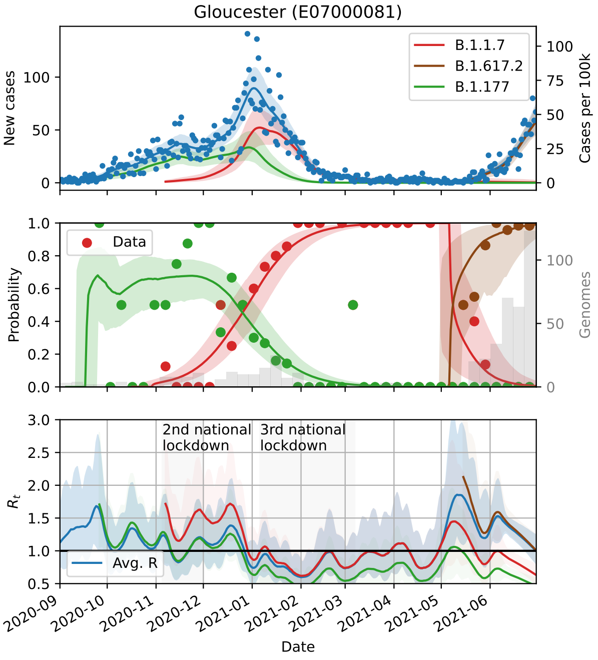
Gloucester (E07000081)
B.1.1.7
B.1.617.2
B.1.177
New cases
Cases per 100k
Data
Probability
Genomes
2nd national lockdown
3rd national lockdown
$R_t$
Avg. R
Date

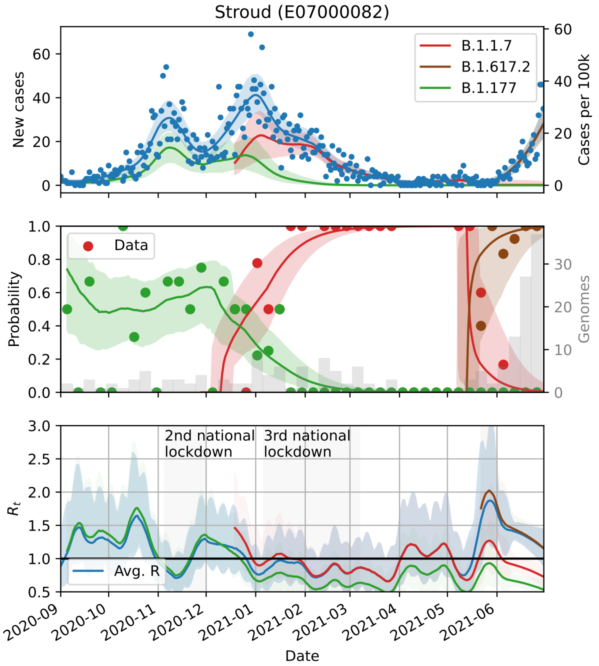
Stroud (E07000082)
B.1.1.7
B.1.617.2
B.1.177
New cases
Cases per 100k
Data
Probability
Genomes
2nd national lockdown
3rd national lockdown
Avg. R
$R_t$
Date

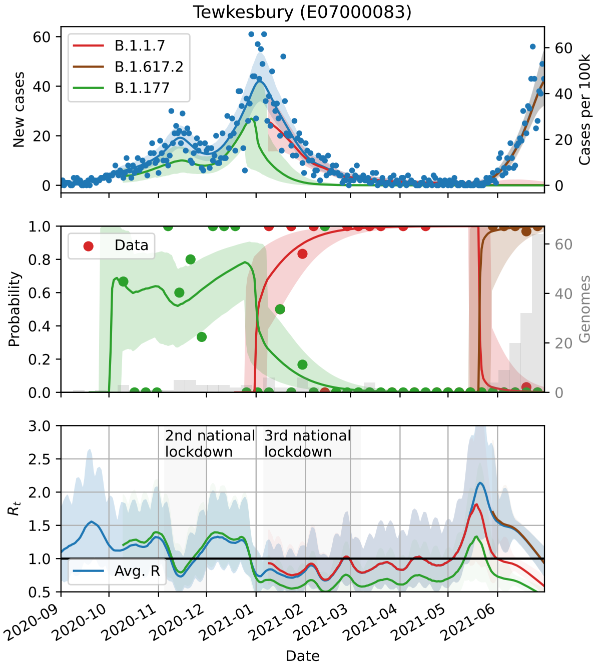
Tewkesbury (E07000083)
B.1.1.7
B.1.617.2
B.1.177
New cases
Cases per 100k
Data
Probability
Genomes
2nd national lockdown
3rd national lockdown
Avg. R
$R_t$
Date

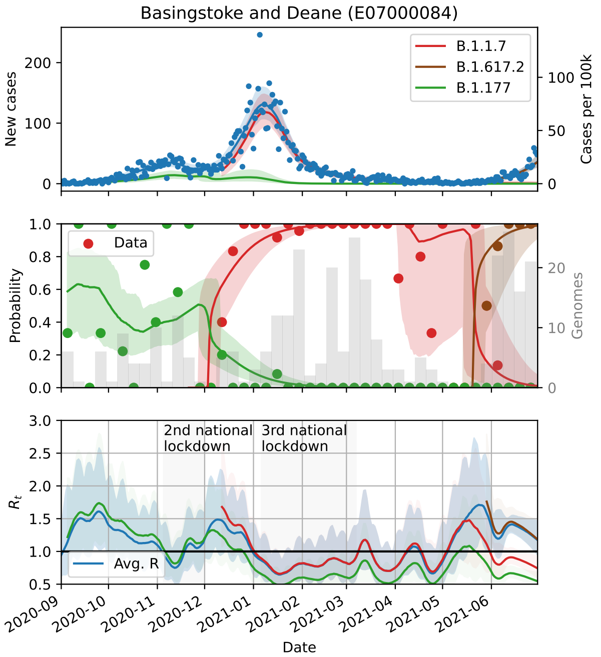
Basingstoke and Deane (E07000084)
B.1.1.7
B.1.617.2
B.1.177
New cases
Cases per 100k
Data
Probability
Genomes
2nd national lockdown
3rd national lockdown
Avg. R
$R_t$
Date

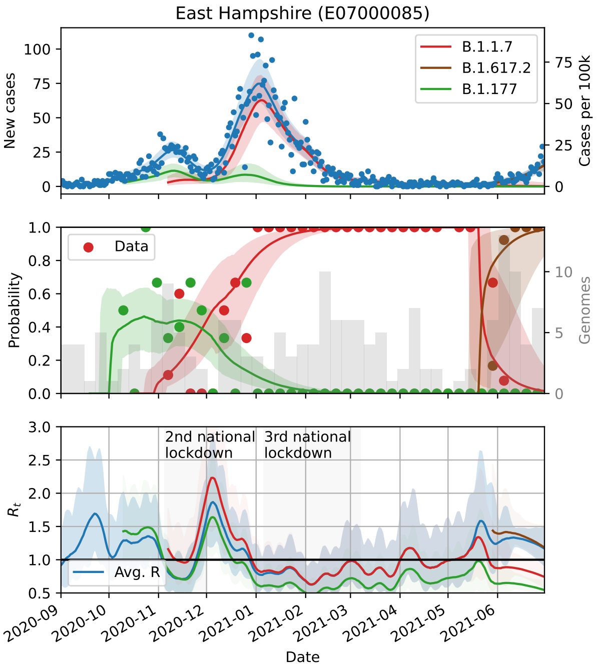
East Hampshire (E07000085)
B.1.1.7
B.1.617.2
B.1.177
New cases
Cases per 100k
Data
Probability
Genomes
2nd national lockdown
3rd national lockdown
Avg. R
$R_t$
Date

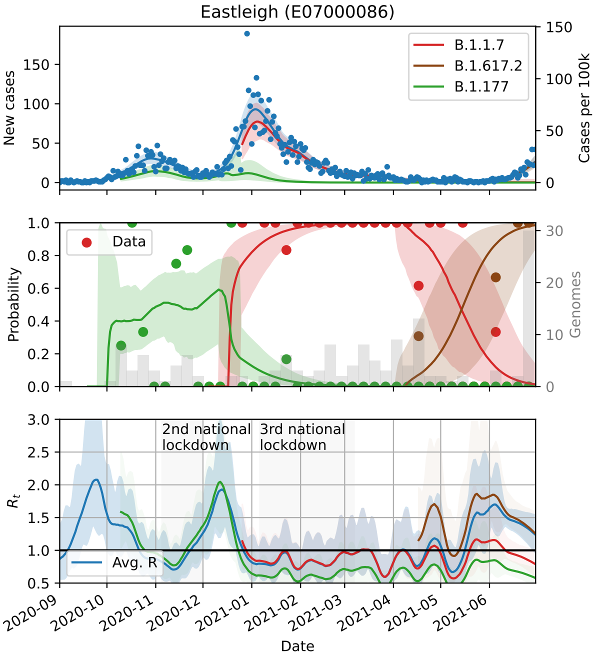
Eastleigh (E07000086)
New cases
Cases per 100k
B.1.1.7
B.1.617.2
B.1.177
Data
Probability
Genomes
2nd national lockdown
3rd national lockdown
Avg. R
$R_t$
Date

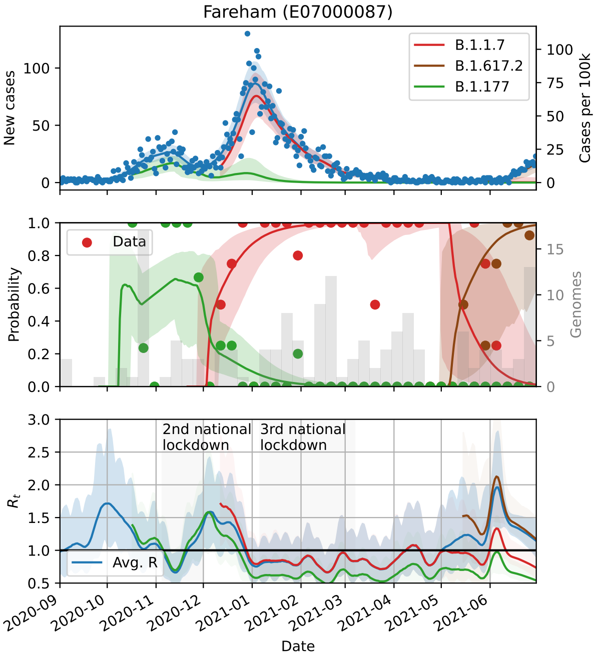
Fareham (E07000087)
New cases
Cases per 100k
B.1.1.7
B.1.617.2
B.1.177
Data
Probability
Genomes
2nd national lockdown
3rd national lockdown
Avg. R
$R_t$
Date

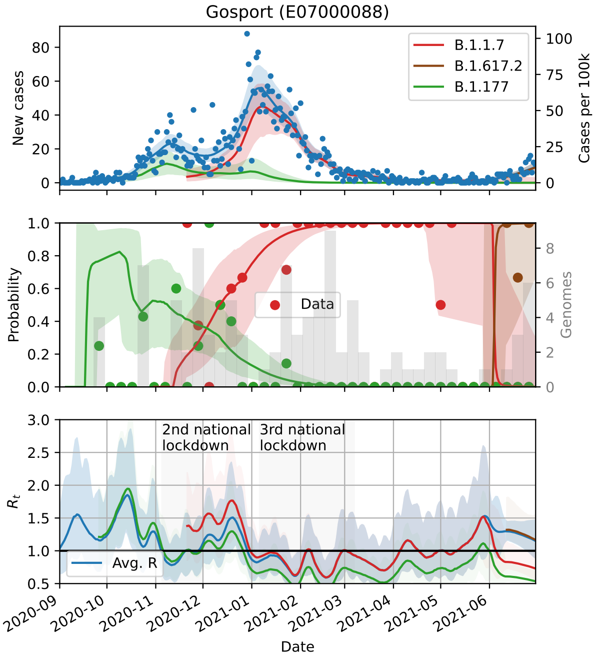
Gosport (E07000088)
New cases
Cases per 100k
B.1.1.7
B.1.617.2
B.1.177
Data
Probability
Genomes
2nd national lockdown
3rd national lockdown
Avg. R
$R_t$
Date

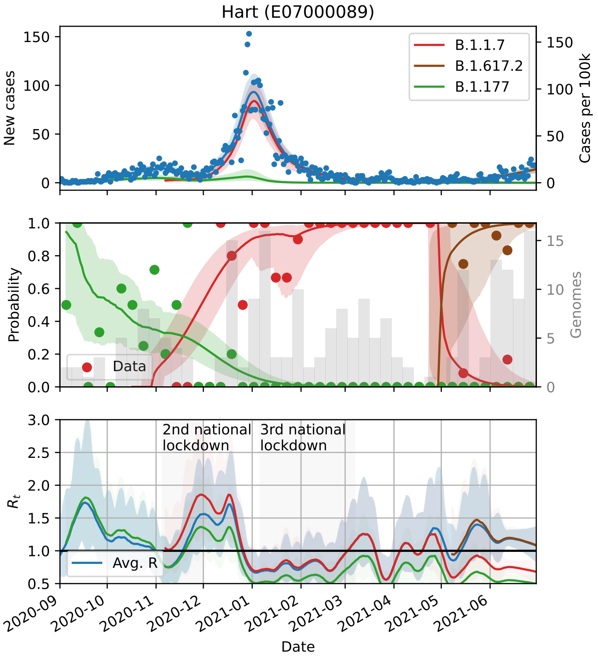
Hart (E07000089)
New cases
Cases per 100k
B.1.1.7
B.1.617.2
B.1.177
Data
Probability
Genomes
2nd national lockdown
3rd national lockdown
Avg. R
$R_t$
Date

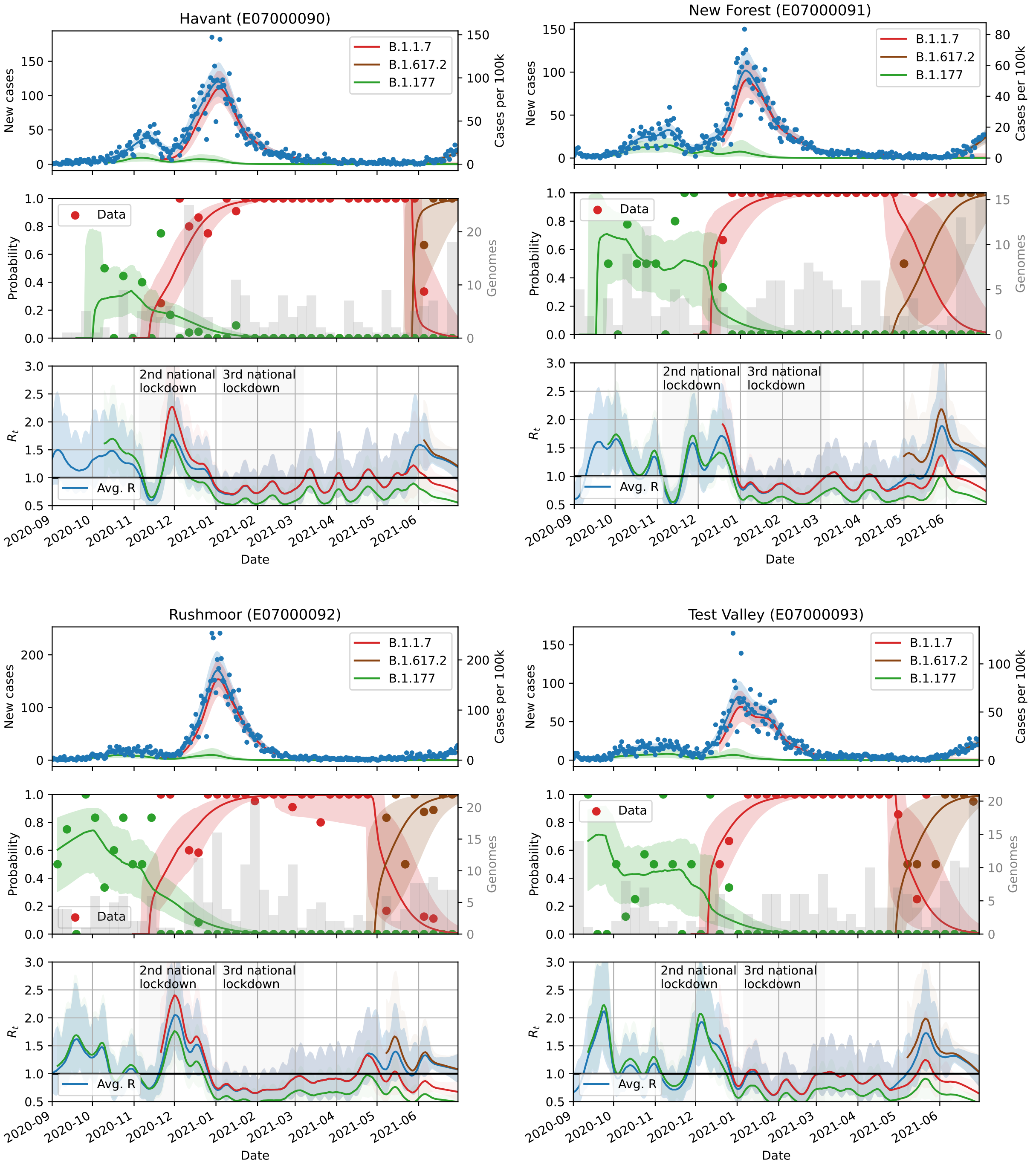
Havant (E07000090)
New Forest (E07000091)
Rushmoor (E07000092)
Test Valley (E07000093)
New cases
Cases per 100k
B.1.1.7
B.1.617.2
B.1.177
Probability
Genomes
Data
$R_t$
Avg. R
2nd national lockdown
3rd national lockdown
Date
2020-09
2020-10
2020-11
2020-12
2021-01
2021-02
2021-03
2021-04
2021-05
2021-06

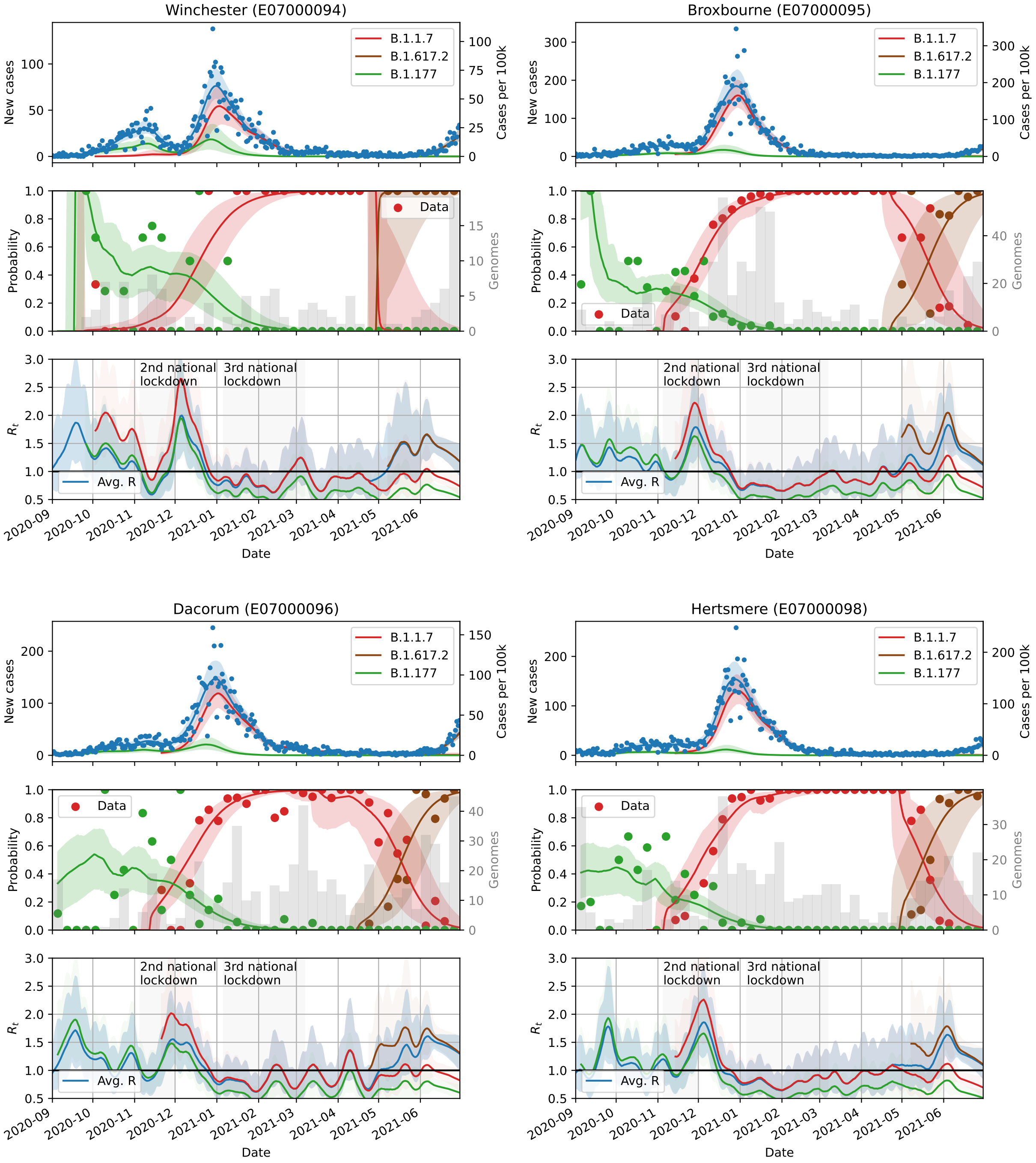
Winchester (E07000094)
Broxbourne (E07000095)
Dacorum (E07000096)
Hertsmere (E07000098)
B.1.1.7
B.1.617.2
B.1.177
New cases
Cases per 100k
Data
Probability
Genomes
2nd national lockdown
3rd national lockdown
Avg. R
$R_t$
2020-09
2020-10
2020-11
2020-12
2021-01
2021-02
2021-03
2021-04
2021-05
2021-06
Date

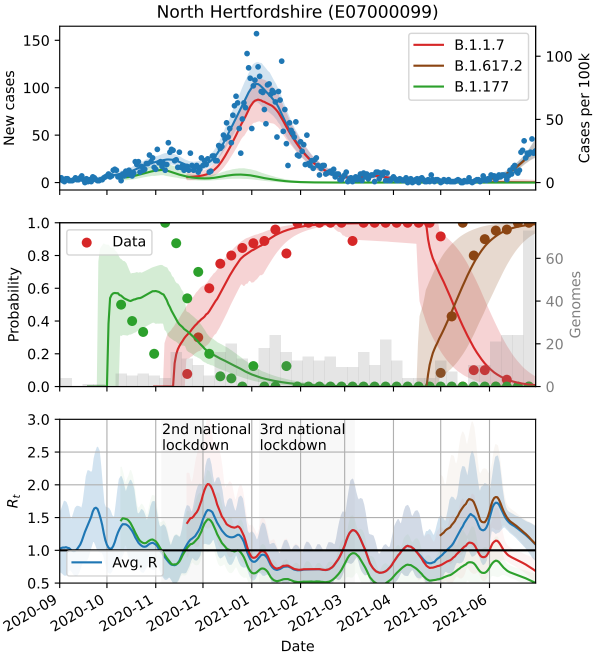
North Hertfordshire (E07000099)
New cases
Cases per 100k
B.1.1.7
B.1.617.2
B.1.177
Probability
Genomes
Data
$R_t$
2nd national lockdown
3rd national lockdown
Avg. R
2020-09
2020-10
2020-11
2020-12
2021-01
2021-02
2021-03
2021-04
2021-05
2021-06
Date

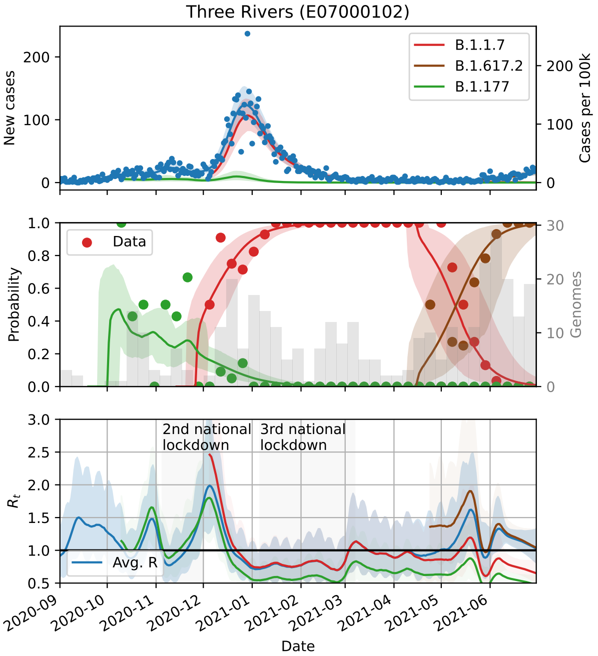
Three Rivers (E07000102)
New cases
Cases per 100k
B.1.1.7
B.1.617.2
B.1.177
Probability
Genomes
Data
$R_t$
2nd national lockdown
3rd national lockdown
Avg. R
2020-09
2020-10
2020-11
2020-12
2021-01
2021-02
2021-03
2021-04
2021-05
2021-06
Date

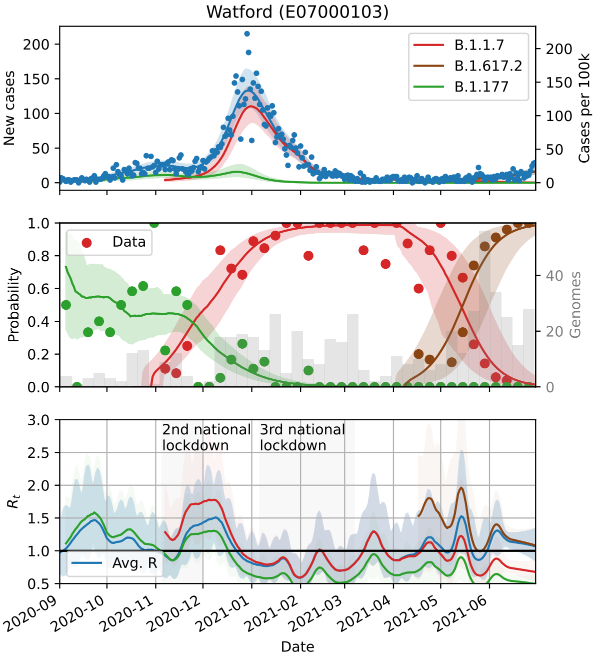
Watford (E07000103)
New cases
Cases per 100k
B.1.1.7
B.1.617.2
B.1.177
Probability
Genomes
Data
$R_t$
2nd national lockdown
3rd national lockdown
Avg. R
2020-09
2020-10
2020-11
2020-12
2021-01
2021-02
2021-03
2021-04
2021-05
2021-06
Date

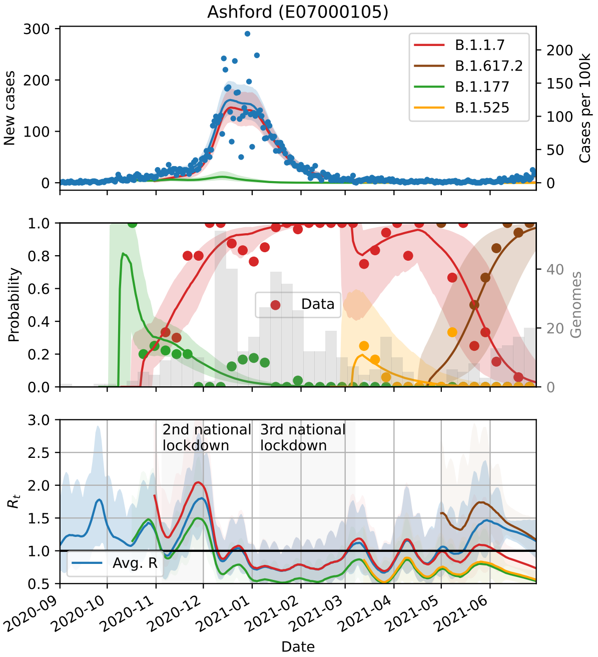
Ashford (E07000105)
New cases
Cases per 100k
B.1.1.7
B.1.617.2
B.1.177
B.1.525
Probability
Genomes
Data
$R_t$
2nd national lockdown
3rd national lockdown
Avg. R
2020-09
2020-10
2020-11
2020-12
2021-01
2021-02
2021-03
2021-04
2021-05
2021-06
Date

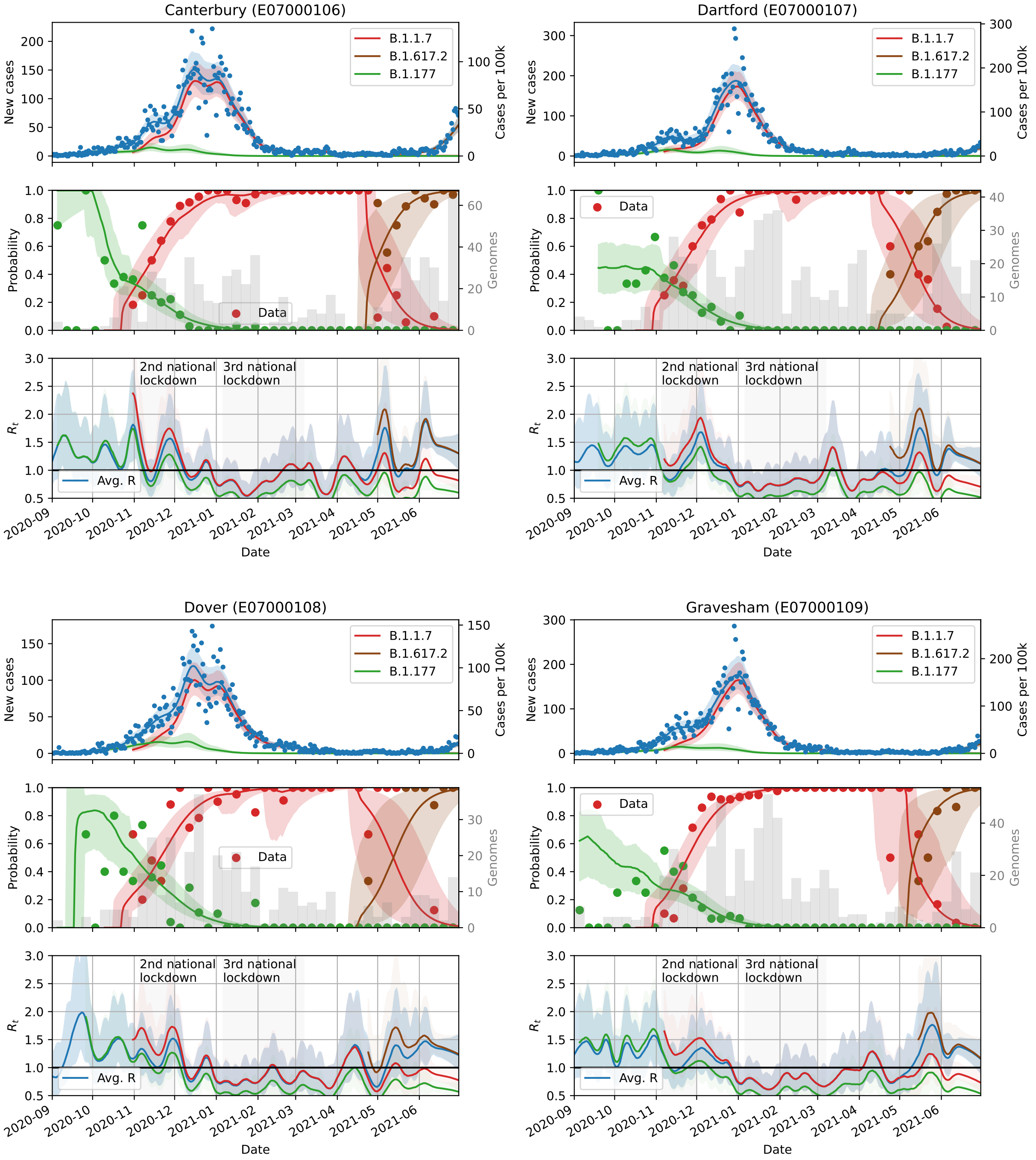

Canterbury (E07000106)
Dartford (E07000107)
Dover (E07000108)
Gravesham (E07000109)
New cases
Cases per 100k
B.1.1.7
B.1.617.2
B.1.177
Probability
Genomes
Data
$R_t$
2nd national lockdown
3rd national lockdown
Avg. R
Date
2020-09
2020-10
2020-11
2020-12
2021-01
2021-02
2021-03
2021-04
2021-05
2021-06

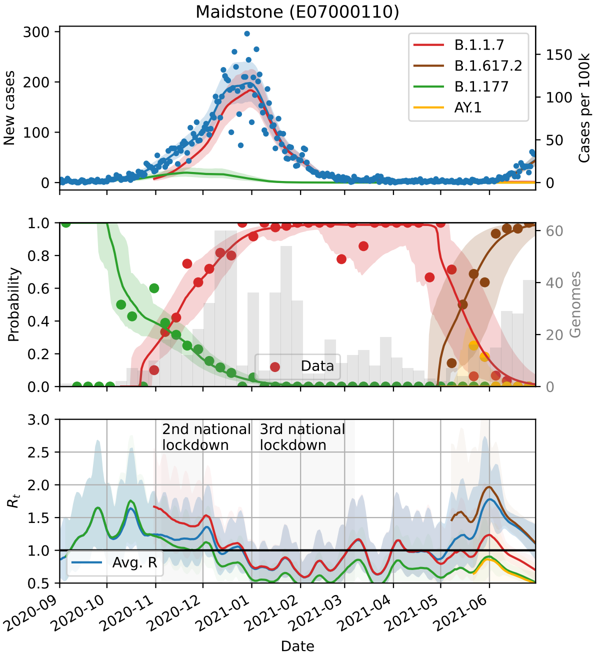
Maidstone (E07000110)
B.1.1.7
B.1.617.2
B.1.177
AY.1
New cases
Cases per 100k
Probability
Genomes
Data
$R_t$
2nd national lockdown
3rd national lockdown
Avg. R
Date

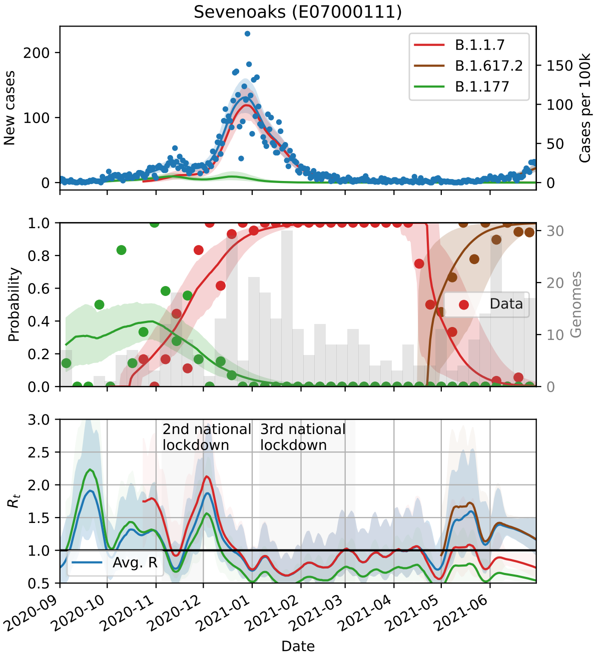
Sevenoaks (E07000111)
B.1.1.7
B.1.617.2
B.1.177
New cases
Cases per 100k
Probability
Genomes
Data
$R_t$
2nd national lockdown
3rd national lockdown
Avg. R
Date

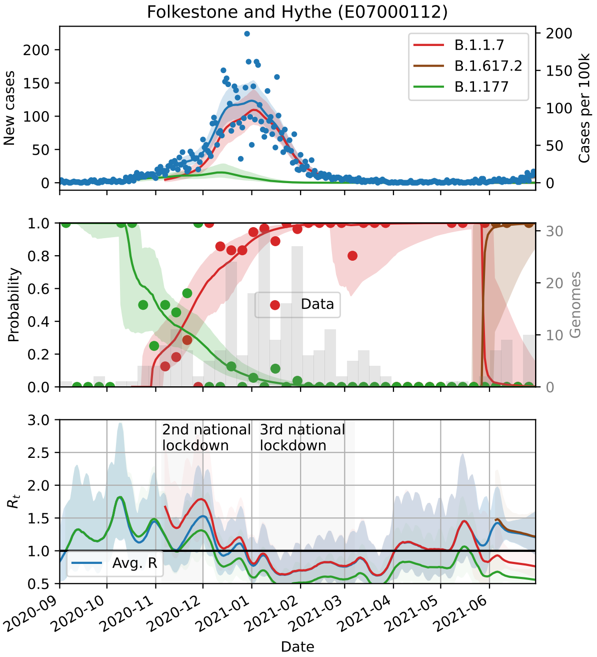
Folkestone and Hythe (E07000112)
B.1.1.7
B.1.617.2
B.1.177
New cases
Cases per 100k
Probability
Genomes
Data
$R_t$
2nd national lockdown
3rd national lockdown
Avg. R
Date

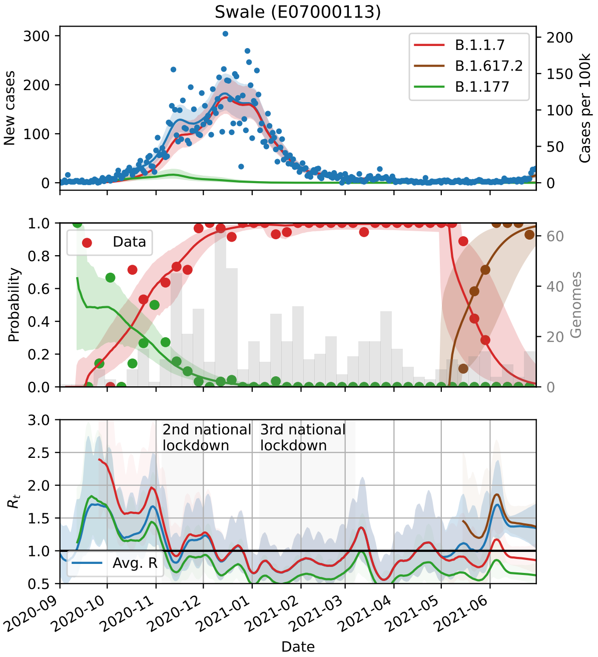
Swale (E07000113)
B.1.1.7
B.1.617.2
B.1.177
New cases
Cases per 100k
Probability
Genomes
Data
$R_t$
2nd national lockdown
3rd national lockdown
Avg. R
Date

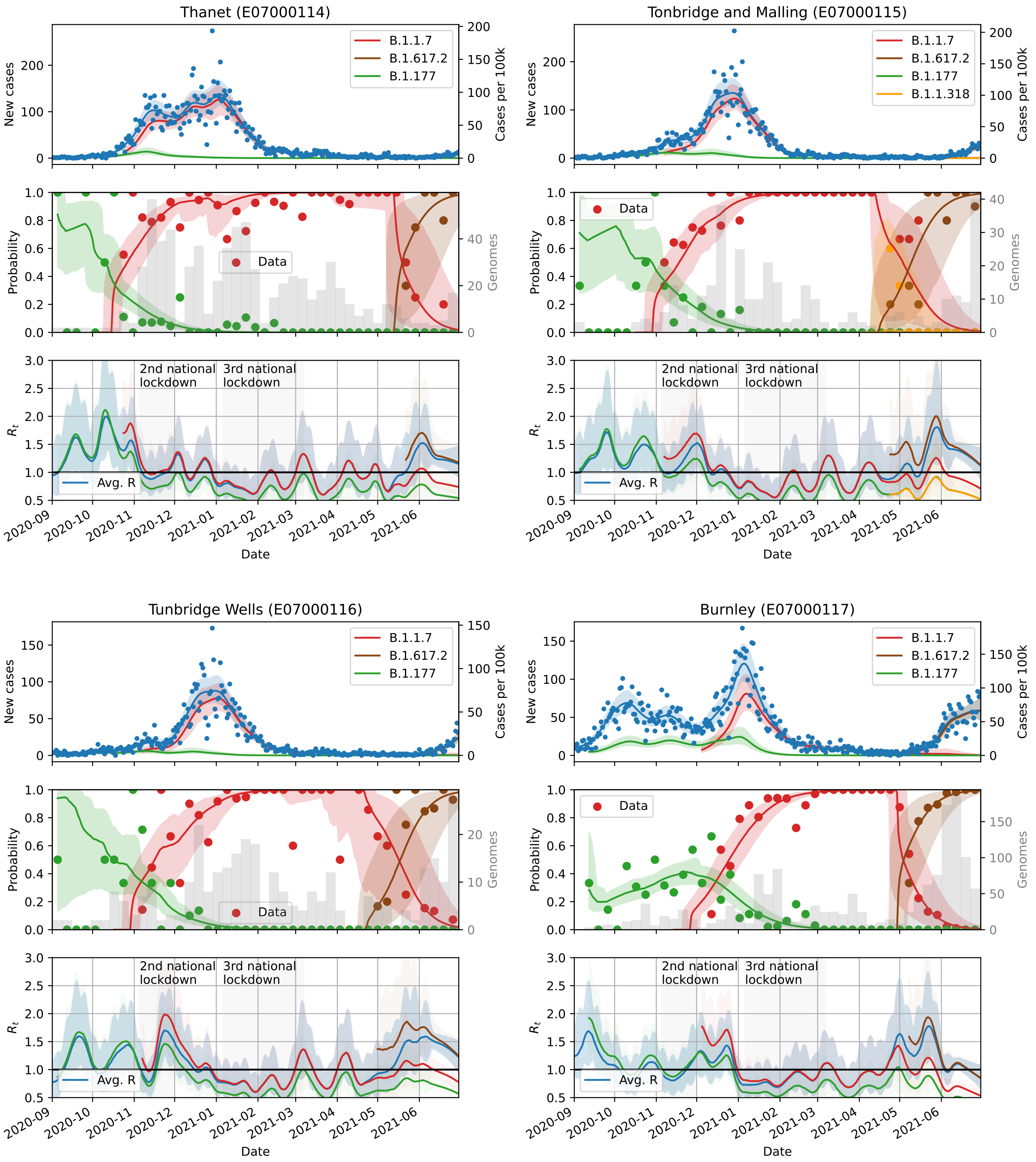
Thanet (E07000114)
Tonbridge and Malling (E07000115)
Tunbridge Wells (E07000116)
Burnley (E07000117)
B.1.1.7
B.1.617.2
B.1.177
B.1.1.318
New cases
Cases per 100k
Probability
Genomes
Data
$R_t$
Avg. R
2nd national lockdown
3rd national lockdown
Date
2020-09
2020-10
2020-11
2020-12
2021-01
2021-02
2021-03
2021-04
2021-05
2021-06

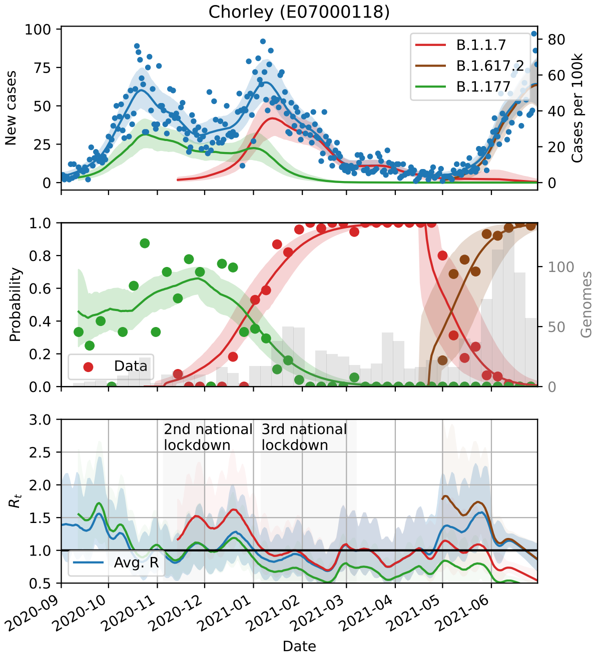
Chorley (E07000118)
New cases
Cases per 100k
B.1.1.7
B.1.617.2
B.1.177
Probability
Genomes
Data
$R_t$
2nd national lockdown
3rd national lockdown
Avg. R
Date

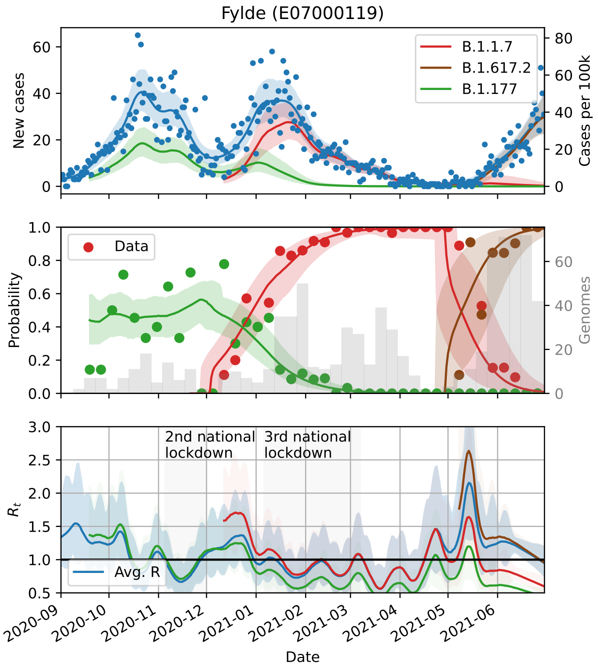
Fylde (E07000119)
New cases
Cases per 100k
B.1.1.7
B.1.617.2
B.1.177
Probability
Genomes
Data
$R_t$
2nd national lockdown
3rd national lockdown
Avg. R
Date

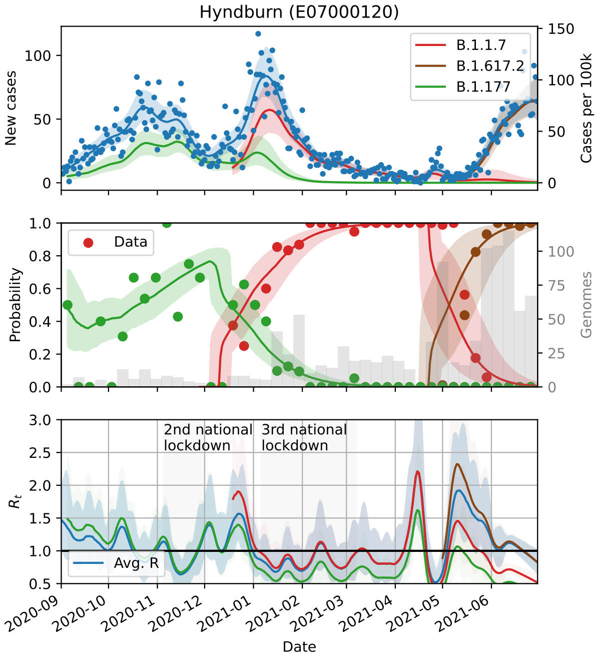
Hyndburn (E07000120)
New cases
Cases per 100k
B.1.1.7
B.1.617.2
B.1.177
Probability
Genomes
Data
$R_t$
2nd national lockdown
3rd national lockdown
Avg. R
Date

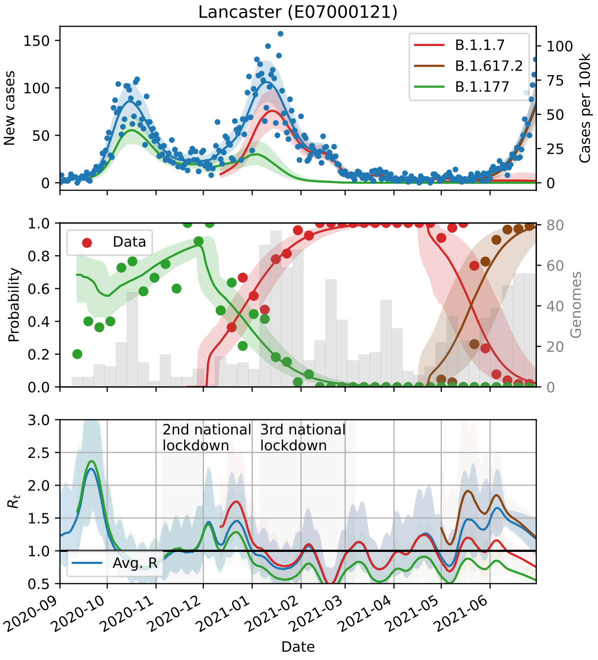
Lancaster (E07000121)
New cases
Cases per 100k
B.1.1.7
B.1.617.2
B.1.177
Probability
Genomes
Data
$R_t$
2nd national lockdown
3rd national lockdown
Avg. R
Date

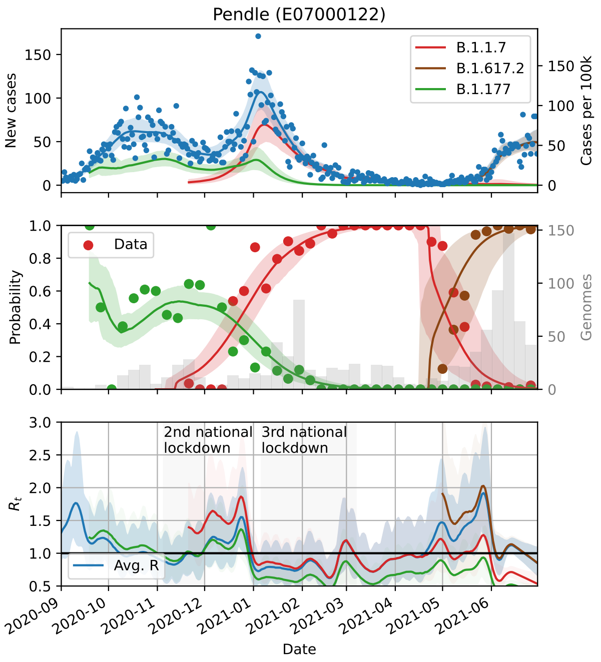
Pendle (E07000122)
New cases
Cases per 100k
B.1.1.7
B.1.617.2
B.1.177
Data
Probability
Genomes
2nd national lockdown
3rd national lockdown
Avg. R
$R_t$
Date

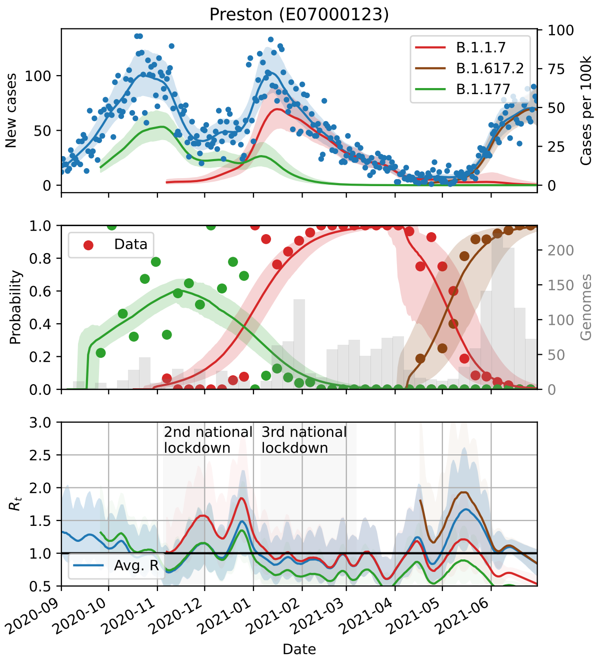
Preston (E07000123)
New cases
Cases per 100k
B.1.1.7
B.1.617.2
B.1.177
Data
Probability
Genomes
2nd national lockdown
3rd national lockdown
Avg. R
$R_t$
Date

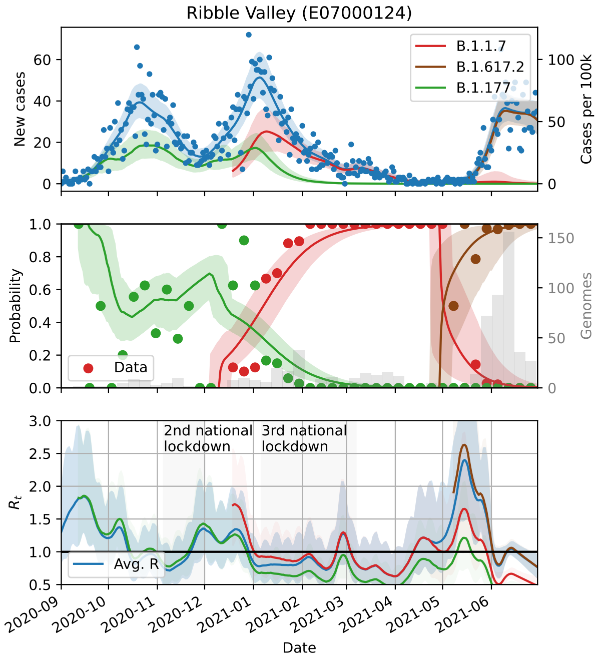
Ribble Valley (E07000124)
New cases
Cases per 100k
B.1.1.7
B.1.617.2
B.1.177
Data
Probability
Genomes
2nd national lockdown
3rd national lockdown
Avg. R
$R_t$
Date

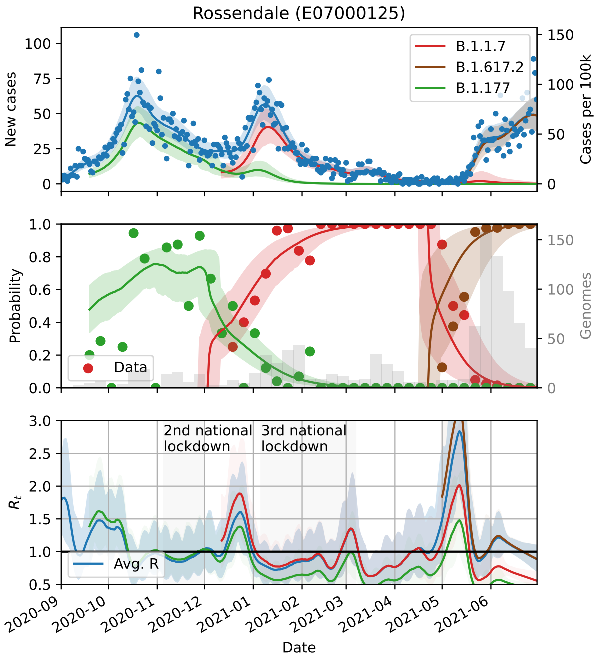
Rossendale (E07000125)
New cases
Cases per 100k
B.1.1.7
B.1.617.2
B.1.177
Data
Probability
Genomes
2nd national lockdown
3rd national lockdown
Avg. R
$R_t$
Date

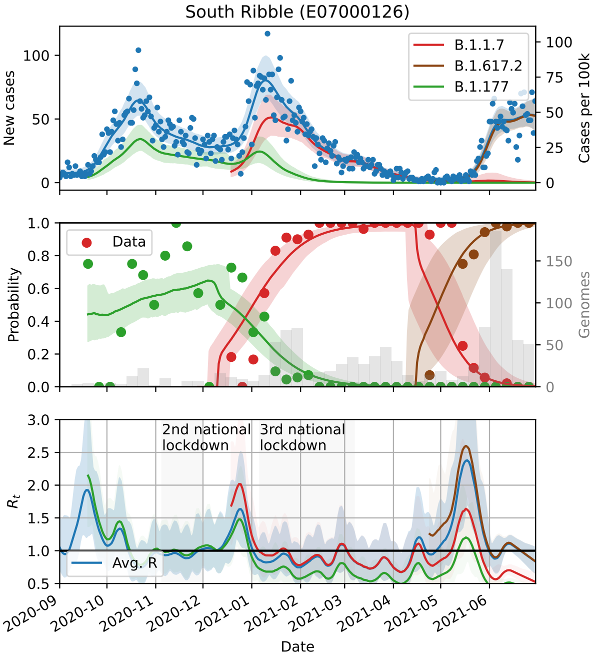
South Ribble (E07000126)
B.1.1.7
B.1.617.2
B.1.177
New cases
Cases per 100k
Data
Probability
Genomes
2nd national lockdown
3rd national lockdown
Avg. R
$R_t$
Date

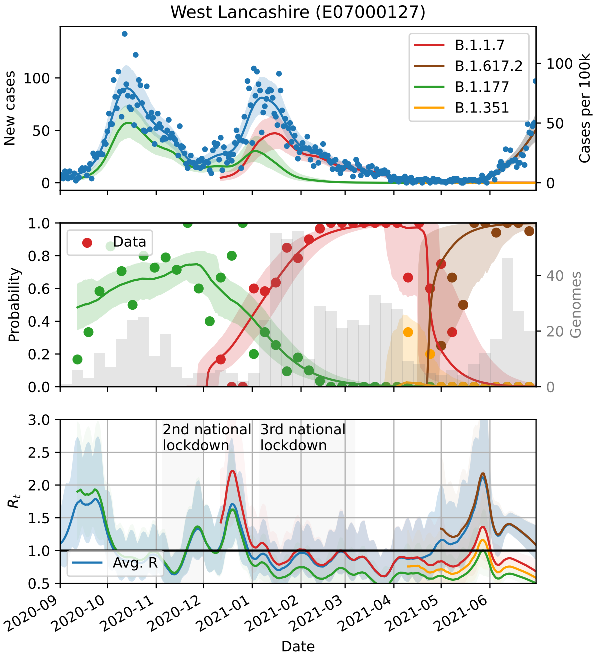
West Lancashire (E07000127)
B.1.1.7
B.1.617.2
B.1.177
B.1.351
New cases
Cases per 100k
Data
Probability
Genomes
2nd national lockdown
3rd national lockdown
Avg. R
$R_t$
Date

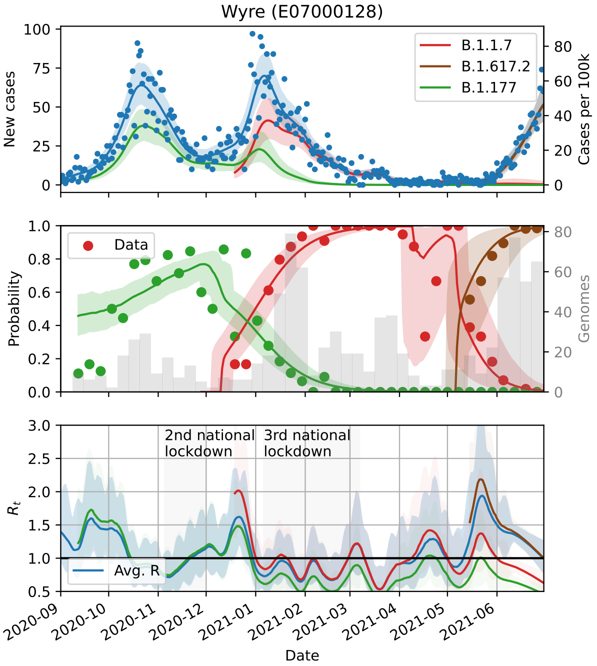
Wyre (E07000128)
B.1.1.7
B.1.617.2
B.1.177
New cases
Cases per 100k
Data
Probability
Genomes
2nd national lockdown
3rd national lockdown
Avg. R
$R_t$
Date

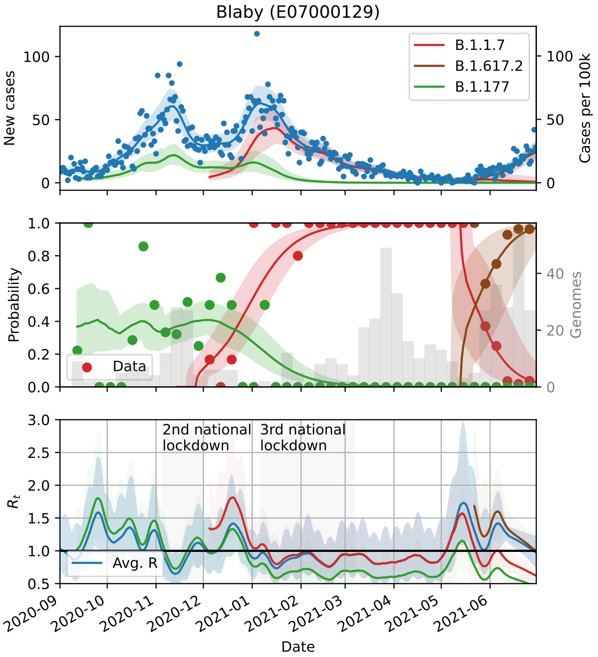
Blaby (E07000129)
B.1.1.7
B.1.617.2
B.1.177
New cases
Cases per 100k
Data
Probability
Genomes
2nd national lockdown
3rd national lockdown
Avg. R
$R_t$
Date

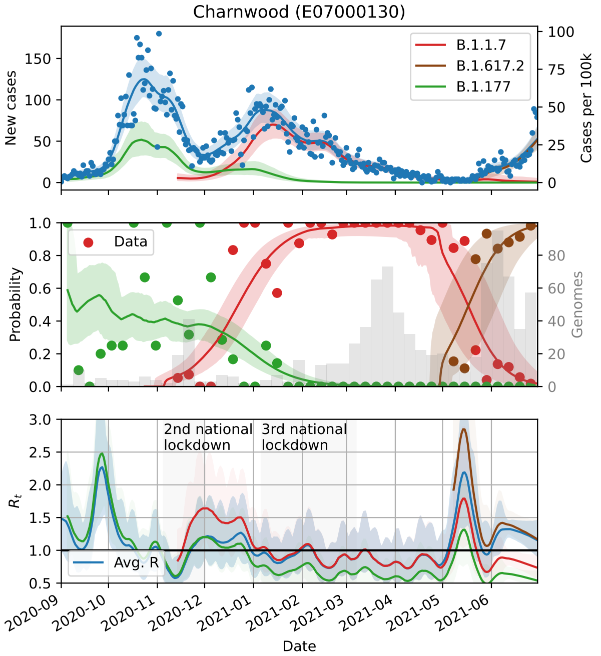
Charnwood (E07000130)
B.1.1.7
B.1.617.2
B.1.177
New cases
Cases per 100k
Data
Probability
Genomes
2nd national lockdown
3rd national lockdown
Avg. R
$R_t$
Date

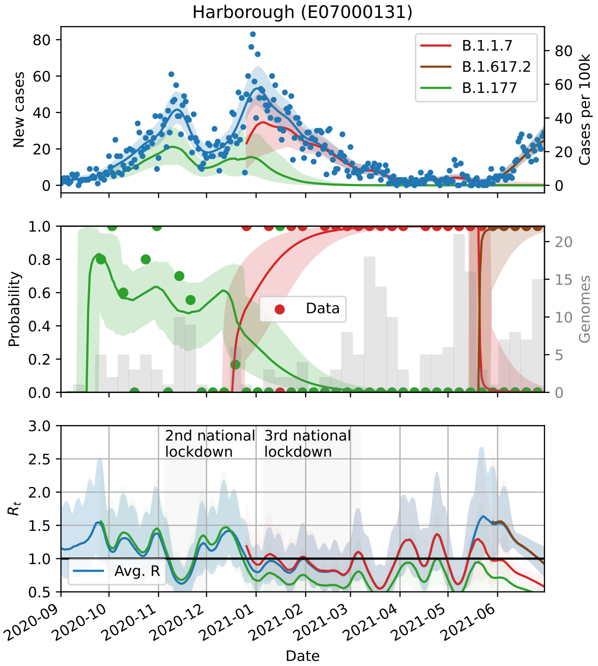
Harborough (E07000131)
B.1.1.7
B.1.617.2
B.1.177
New cases
Cases per 100k
Data
Probability
Genomes
2nd national lockdown
3rd national lockdown
Avg. R
$R_t$
Date

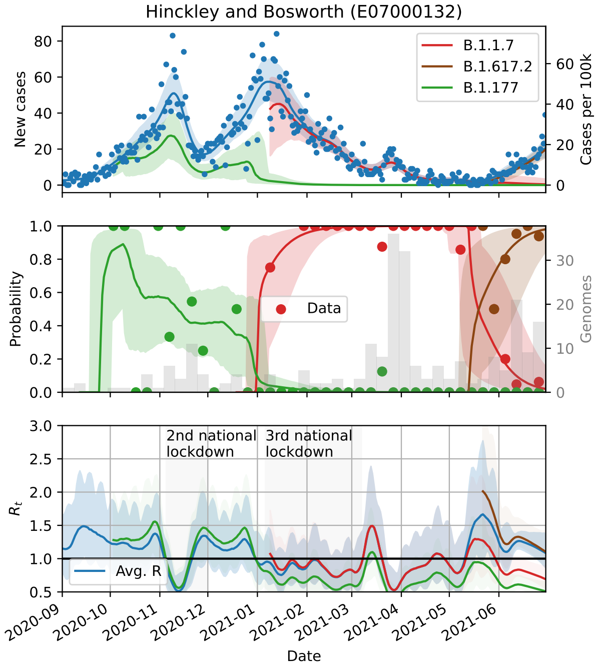
Hinckley and Bosworth (E07000132)
B.1.1.7
B.1.617.2
B.1.177
New cases
Cases per 100k
Data
Probability
Genomes
2nd national lockdown
3rd national lockdown
Avg. R
$R_t$
Date

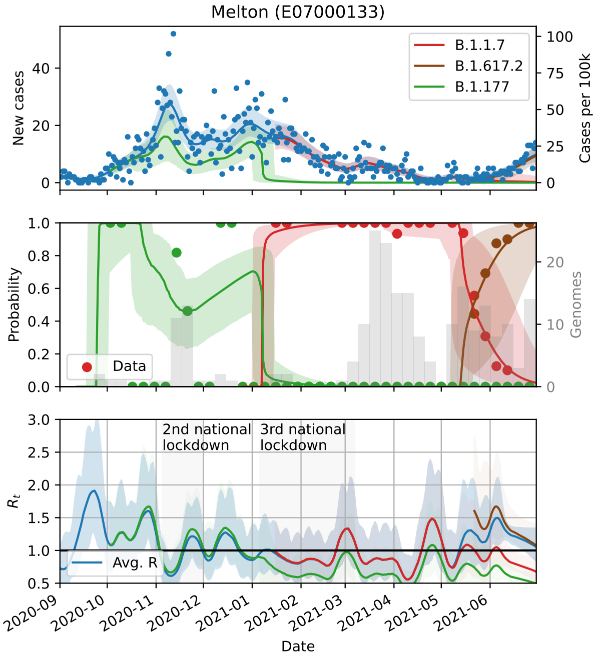
Melton (E07000133)
B.1.1.7
B.1.617.2
B.1.177
New cases
Cases per 100k
Data
Probability
Genomes
2nd national lockdown
3rd national lockdown
Avg. R
$R_t$
Date

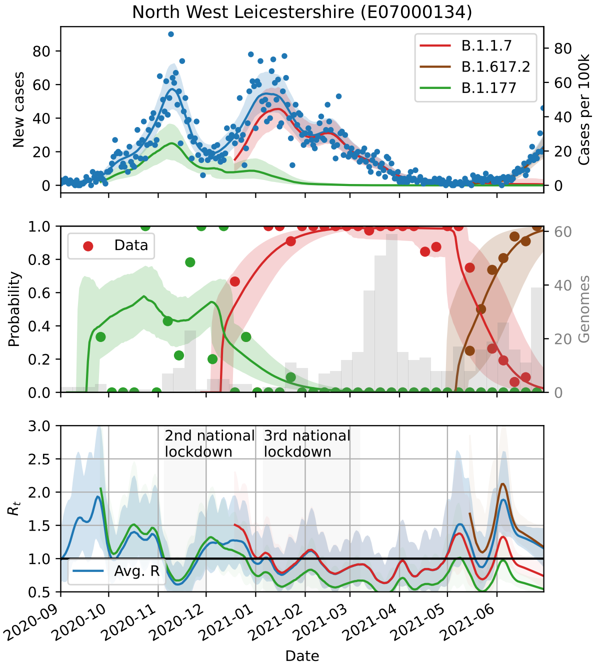
North West Leicestershire (E07000134)
New cases
Cases per 100k
B.1.1.7
B.1.617.2
B.1.177
Data
Probability
Genomes
2nd national lockdown
3rd national lockdown
$R_t$
Avg. R
Date

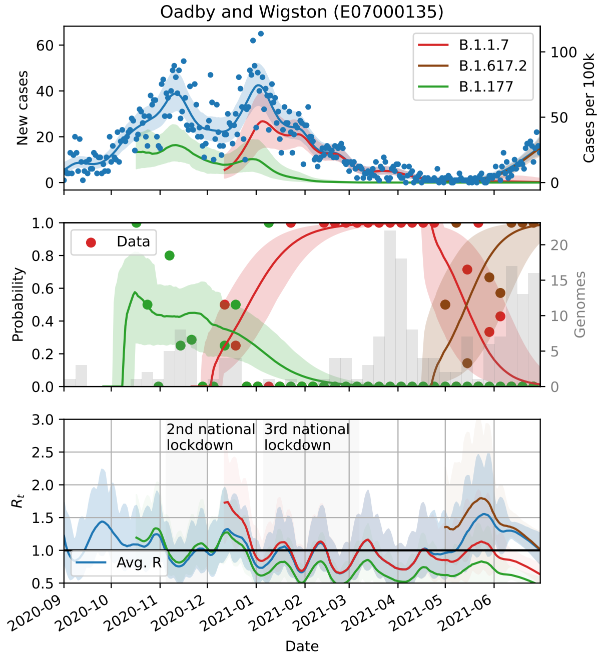
Oadby and Wigston (E07000135)
New cases
Cases per 100k
B.1.1.7
B.1.617.2
B.1.177
Data
Probability
Genomes
2nd national lockdown
3rd national lockdown
$R_t$
Avg. R
Date

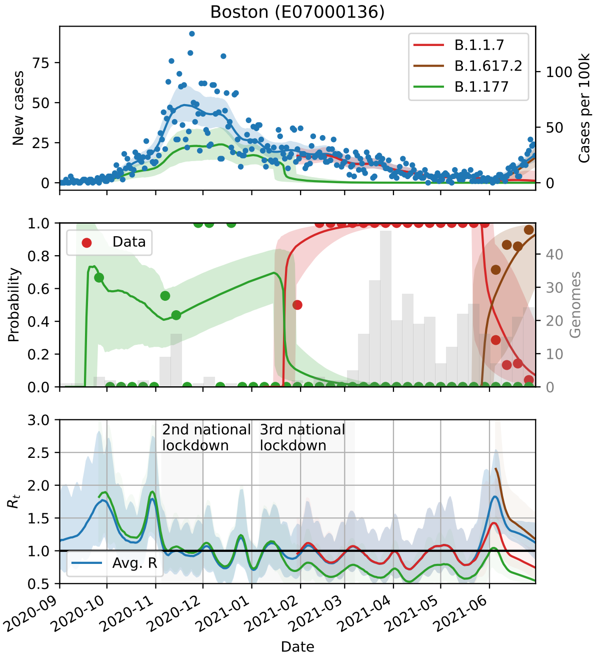
Boston (E07000136)
New cases
Cases per 100k
B.1.1.7
B.1.617.2
B.1.177
Data
Probability
Genomes
2nd national lockdown
3rd national lockdown
$R_t$
Avg. R
Date

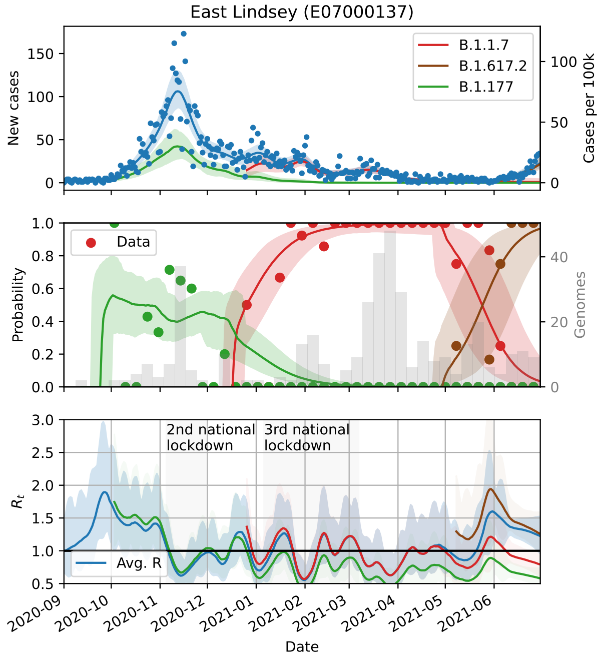
East Lindsey (E07000137)
New cases
Cases per 100k
B.1.1.7
B.1.617.2
B.1.177
Data
Probability
Genomes
2nd national lockdown
3rd national lockdown
$R_t$
Avg. R
Date

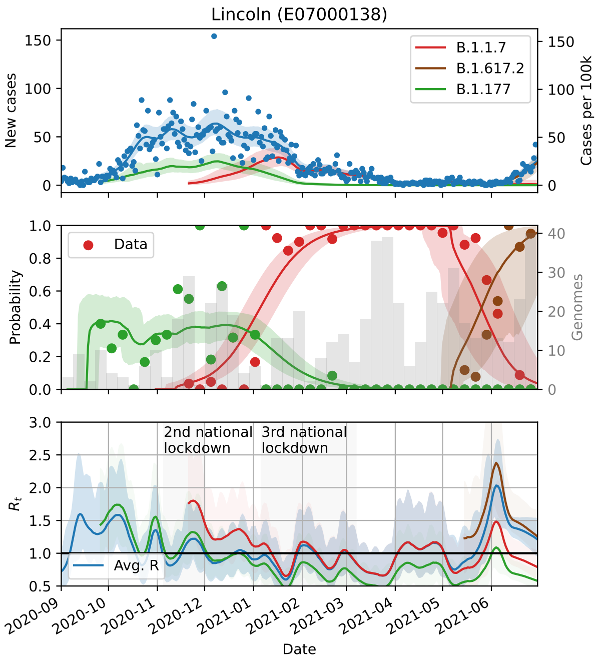
Lincoln (E07000138)
B.1.1.7
B.1.617.2
B.1.177
New cases
Cases per 100k
Data
Probability
Genomes
2nd national lockdown
3rd national lockdown
Avg. R
$R_t$
Date

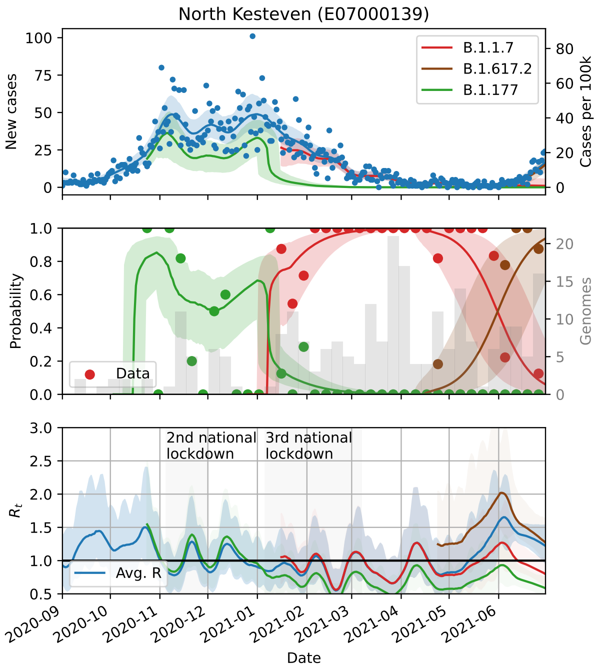
North Kesteven (E07000139)
B.1.1.7
B.1.617.2
B.1.177
New cases
Cases per 100k
Data
Probability
Genomes
2nd national lockdown
3rd national lockdown
Avg. R
$R_t$
Date

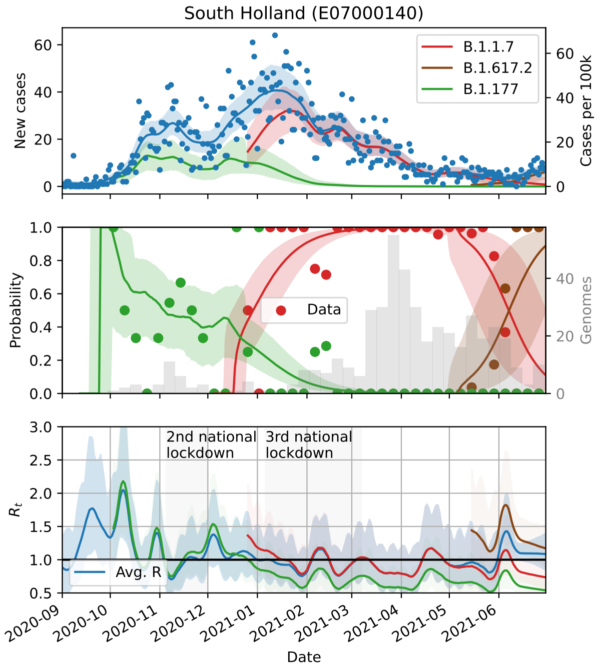
South Holland (E07000140)
B.1.1.7
B.1.617.2
B.1.177
New cases
Cases per 100k
Data
Probability
Genomes
2nd national lockdown
3rd national lockdown
Avg. R
$R_t$
Date

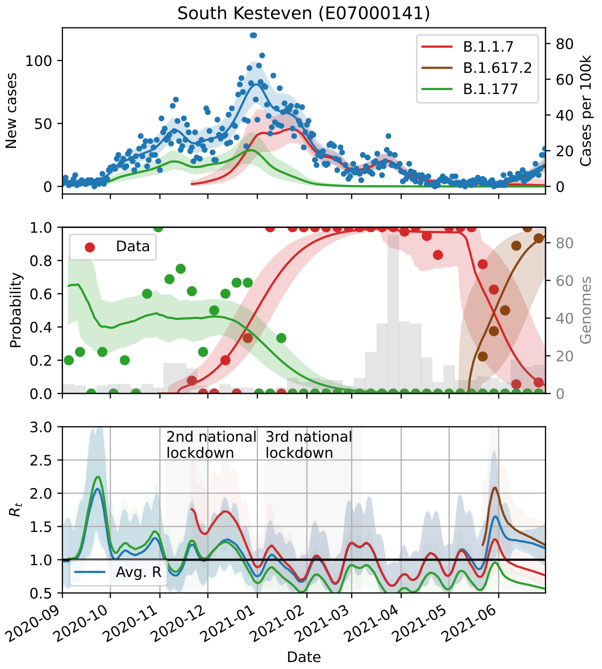
South Kesteven (E07000141)
B.1.1.7
B.1.617.2
B.1.177
New cases
Cases per 100k
Data
Probability
Genomes
2nd national lockdown
3rd national lockdown
Avg. R
$R_t$
Date

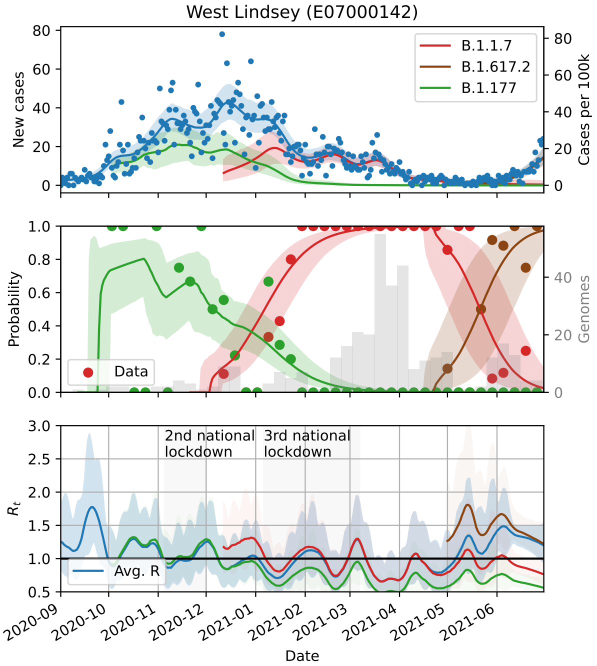
West Lindsey (E07000142)
New cases
Cases per 100k
B.1.1.7
B.1.617.2
B.1.177
Probability
Genomes
Data
$R_t$
2nd national lockdown
3rd national lockdown
Avg. R
Date

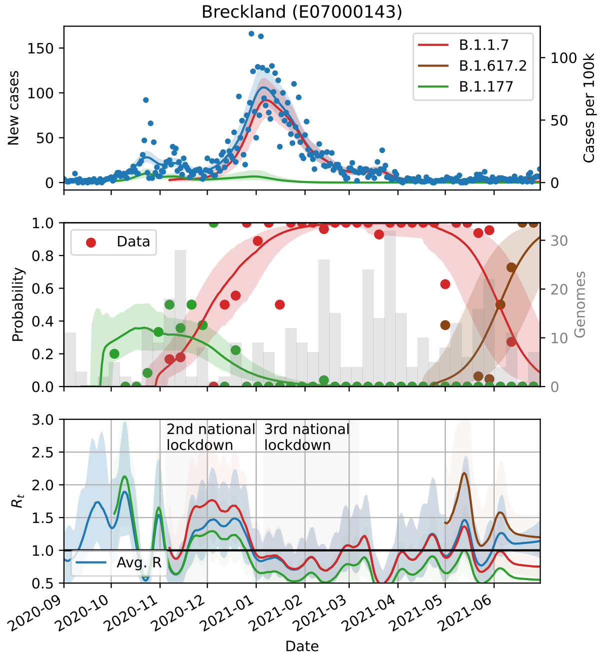
Breckland (E07000143)
New cases
Cases per 100k
B.1.1.7
B.1.617.2
B.1.177
Probability
Genomes
Data
$R_t$
2nd national lockdown
3rd national lockdown
Avg. R
Date

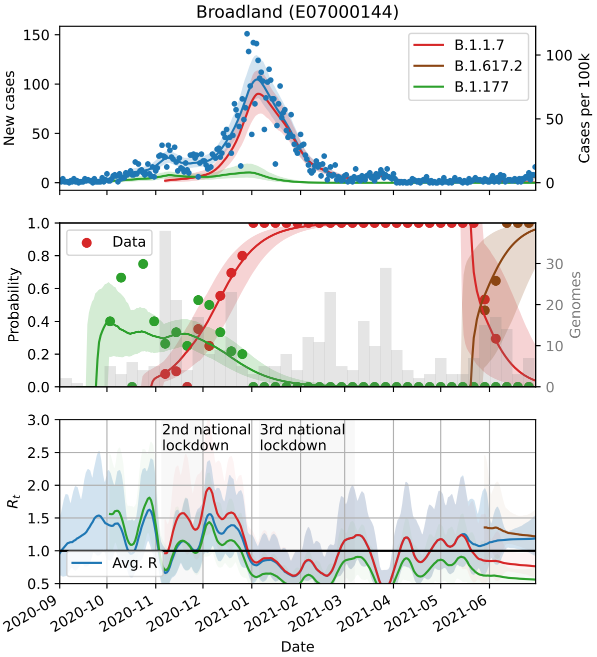
Broadland (E07000144)
New cases
Cases per 100k
B.1.1.7
B.1.617.2
B.1.177
Probability
Genomes
Data
$R_t$
2nd national lockdown
3rd national lockdown
Avg. R
Date

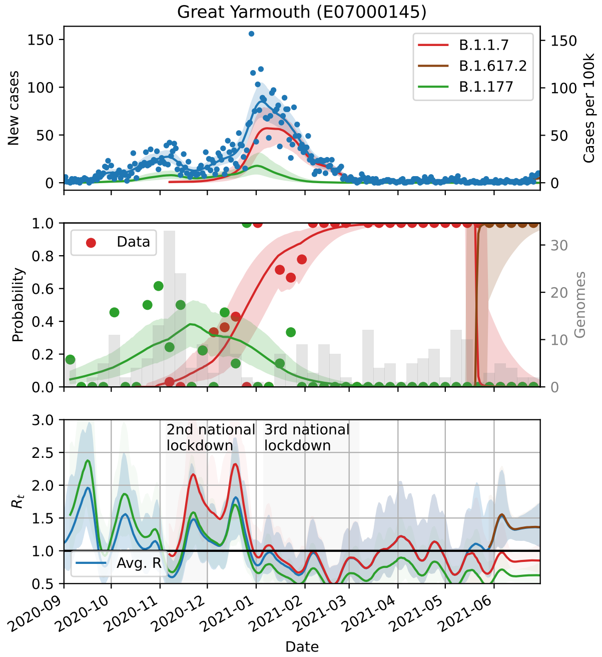
Great Yarmouth (E07000145)
New cases
Cases per 100k
B.1.1.7
B.1.617.2
B.1.177
Probability
Genomes
Data
$R_t$
2nd national lockdown
3rd national lockdown
Avg. R
Date

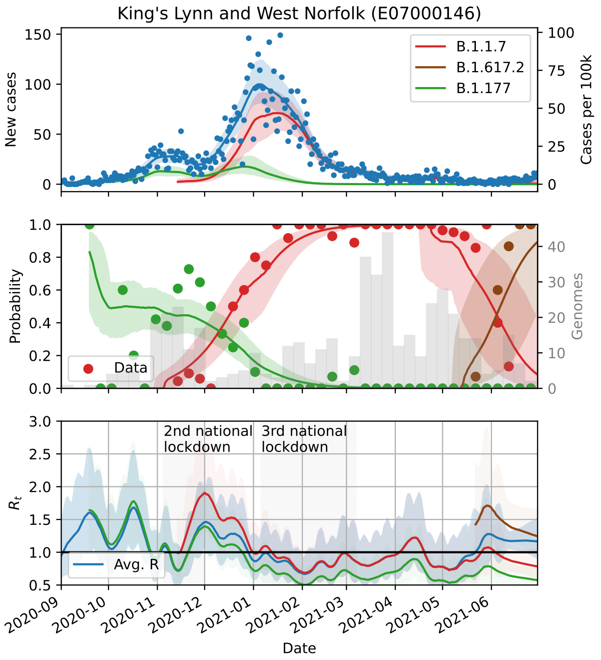
King's Lynn and West Norfolk (E07000146)
New cases
Cases per 100k
B.1.1.7
B.1.617.2
B.1.177
Probability
Genomes
Data
$R_t$
2nd national lockdown
3rd national lockdown
Avg. R
Date

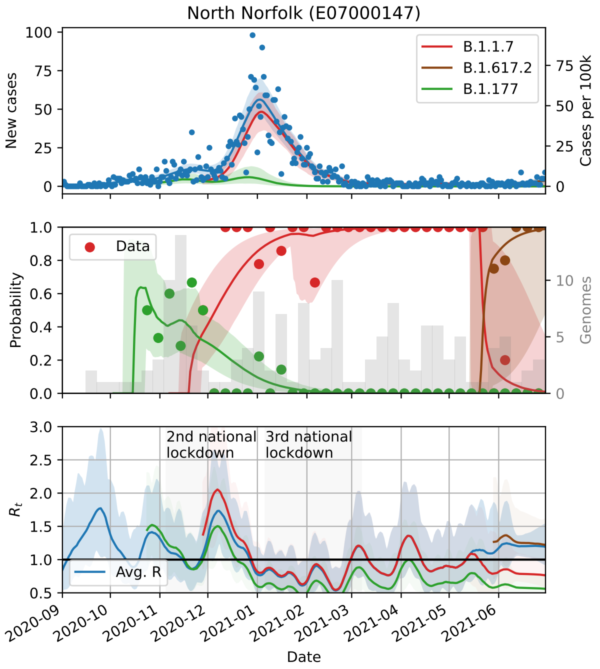
North Norfolk (E07000147)
New cases
Cases per 100k
B.1.1.7
B.1.617.2
B.1.177
Probability
Genomes
Data
$R_t$
2nd national lockdown
3rd national lockdown
Avg. R
Date

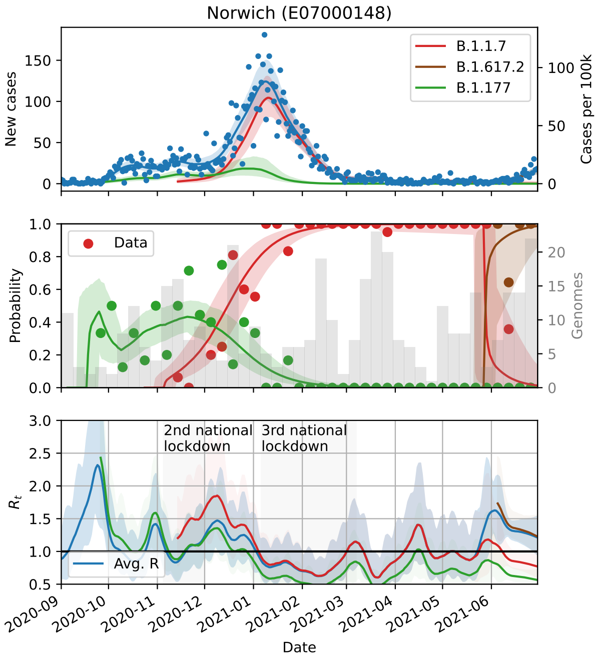
Norwich (E07000148)
New cases
Cases per 100k
B.1.1.7
B.1.617.2
B.1.177
Probability
Genomes
Data
$R_t$
2nd national lockdown
3rd national lockdown
Avg. R
Date

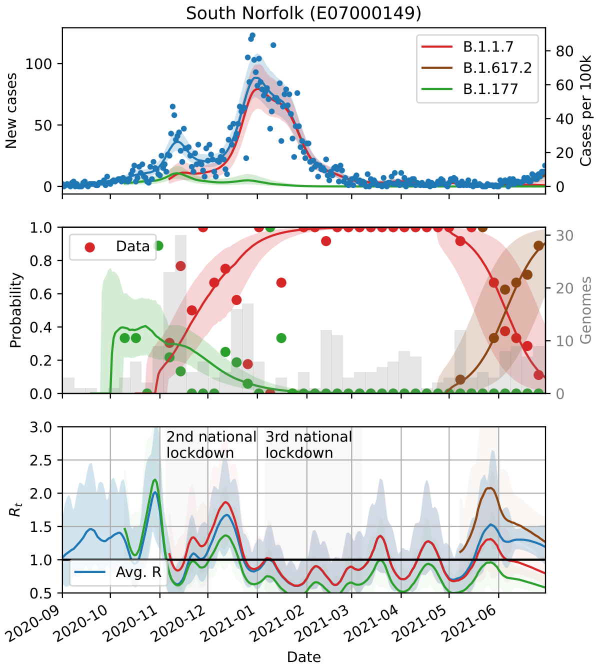
South Norfolk (E07000149)
New cases
Cases per 100k
B.1.1.7
B.1.617.2
B.1.177
Probability
Genomes
Data
$R_t$
2nd national lockdown
3rd national lockdown
Avg. R
Date

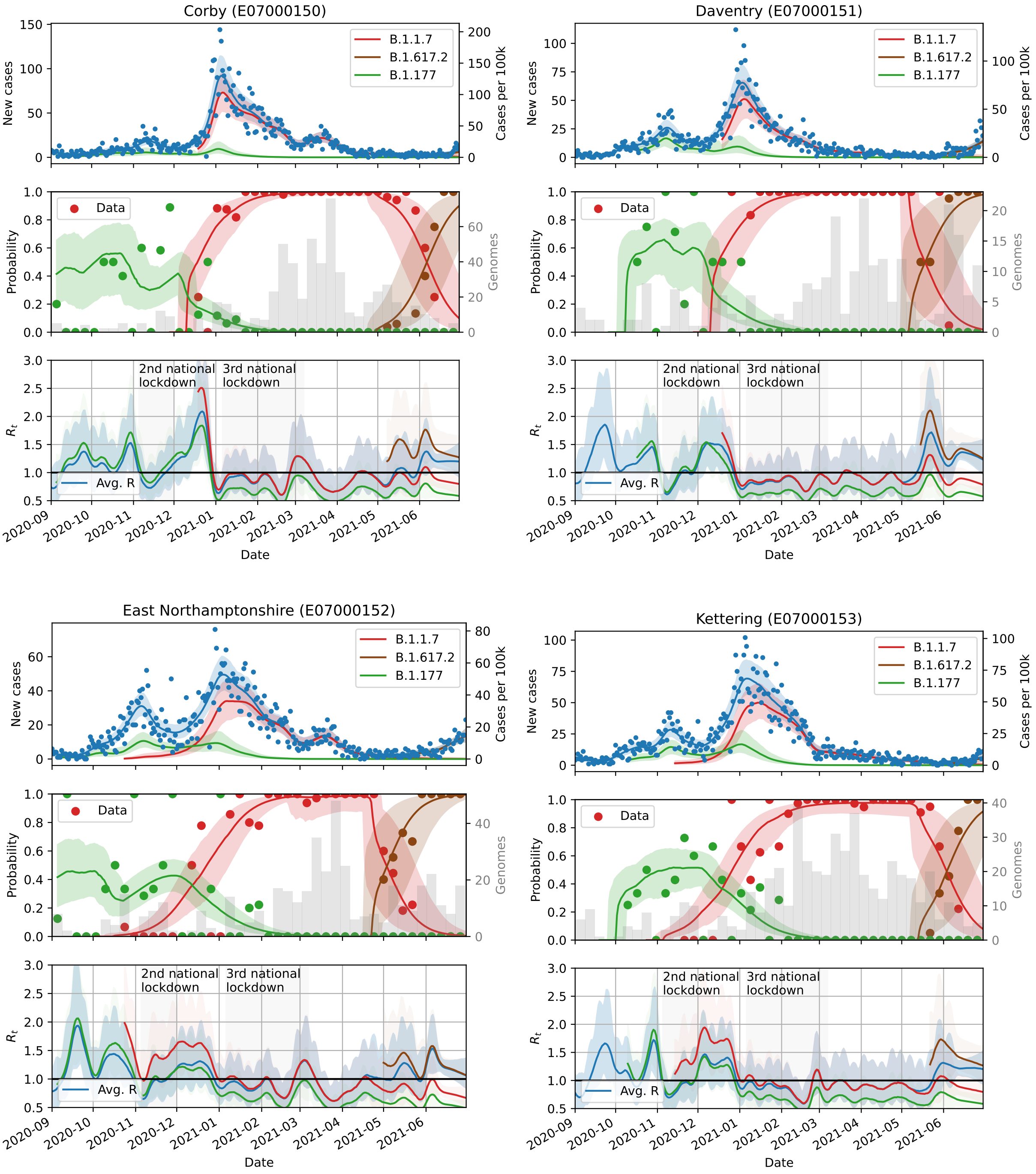

Corby (E07000150)
Daventry (E07000151)
East Northamptonshire (E07000152)
Kettering (E07000153)
New cases
Cases per 100k
B.1.1.7
B.1.617.2
B.1.177
Probability
Genomes
Data
$R_t$
Avg. R
2nd national lockdown
3rd national lockdown
Date
2020-09
2020-10
2020-11
2020-12
2021-01
2021-02
2021-03
2021-04
2021-05
2021-06

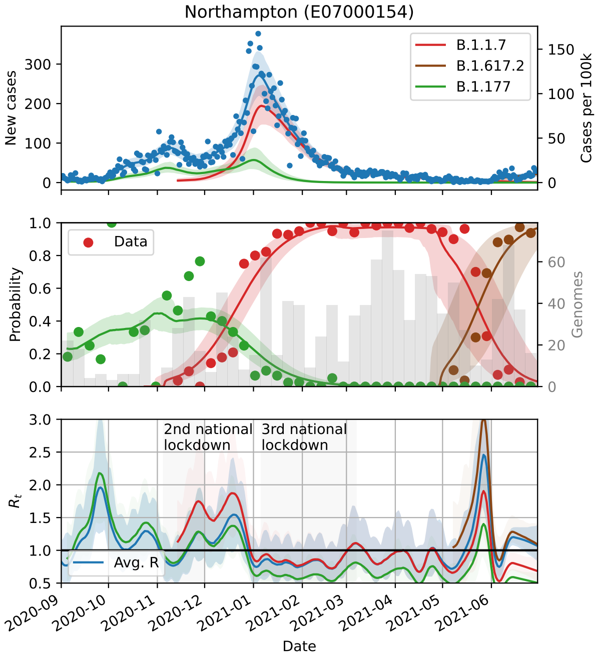
Northampton (E07000154)
B.1.1.7
B.1.617.2
B.1.177
New cases
Cases per 100k
Data
Probability
Genomes
2nd national lockdown
3rd national lockdown
Avg. R
$R_t$
Date

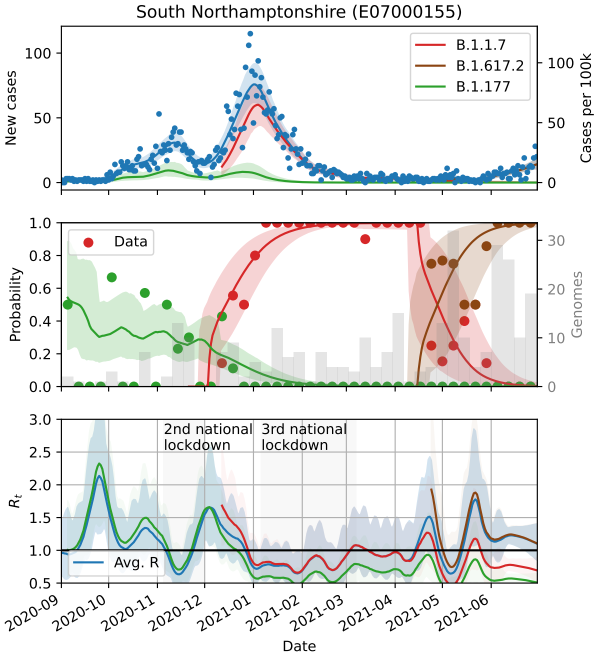
South Northamptonshire (E07000155)
B.1.1.7
B.1.617.2
B.1.177
New cases
Cases per 100k
Data
Probability
Genomes
2nd national lockdown
3rd national lockdown
Avg. R
$R_t$
Date

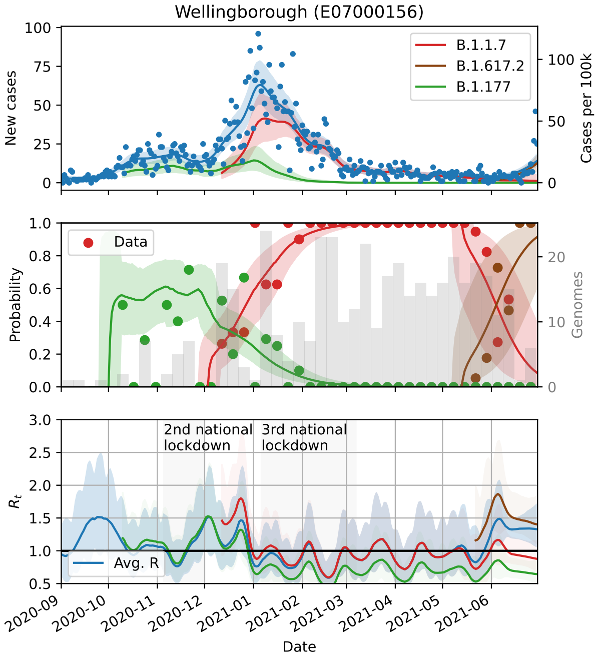
Wellingborough (E07000156)
B.1.1.7
B.1.617.2
B.1.177
New cases
Cases per 100k
Data
Probability
Genomes
2nd national lockdown
3rd national lockdown
Avg. R
$R_t$
Date

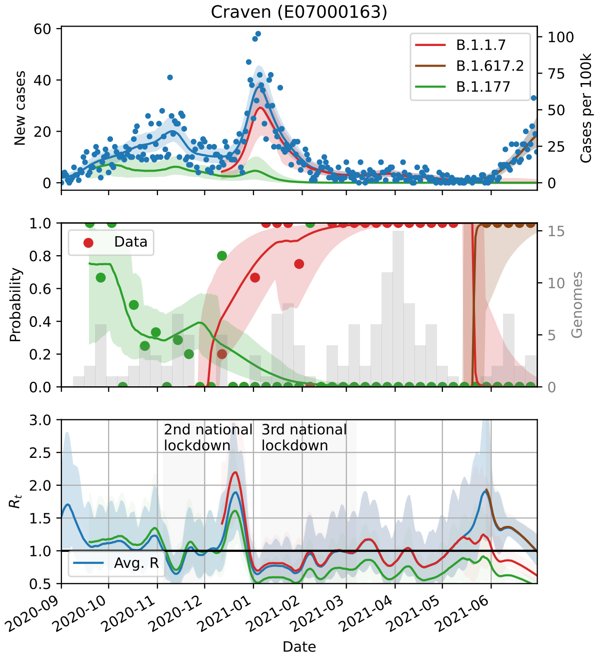
Craven (E07000163)
B.1.1.7
B.1.617.2
B.1.177
New cases
Cases per 100k
Data
Probability
Genomes
2nd national lockdown
3rd national lockdown
Avg. R
$R_t$
Date

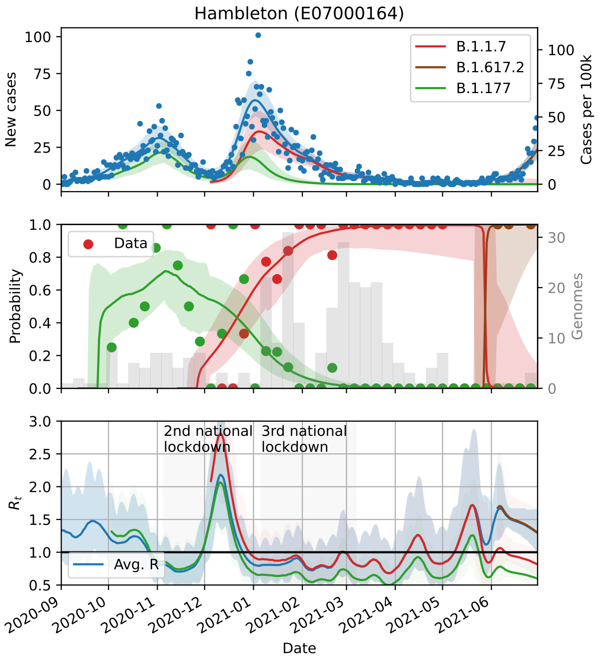
Hambleton (E07000164)
B.1.1.7
B.1.617.2
B.1.177
New cases
Cases per 100k
Data
Probability
Genomes
2nd national lockdown
3rd national lockdown
Avg. R
$R_t$
Date

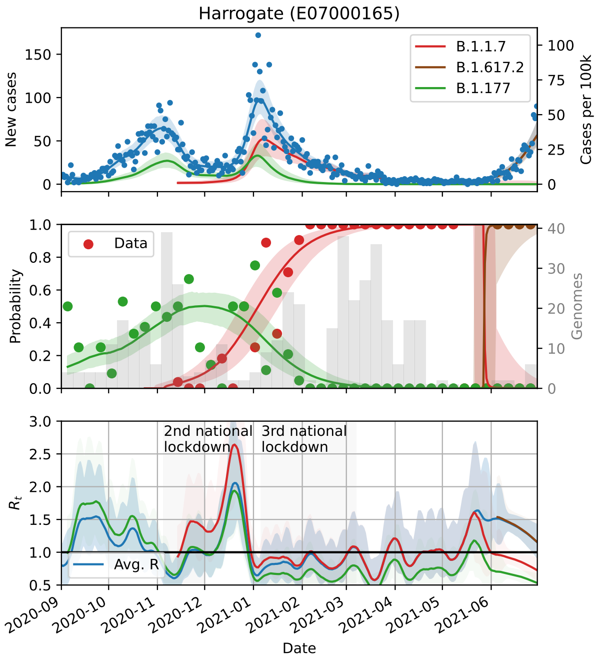
Harrogate (E07000165)
B.1.1.7
B.1.617.2
B.1.177
New cases
Cases per 100k
Data
Probability
Genomes
2nd national lockdown
3rd national lockdown
Avg. R
$R_t$
Date

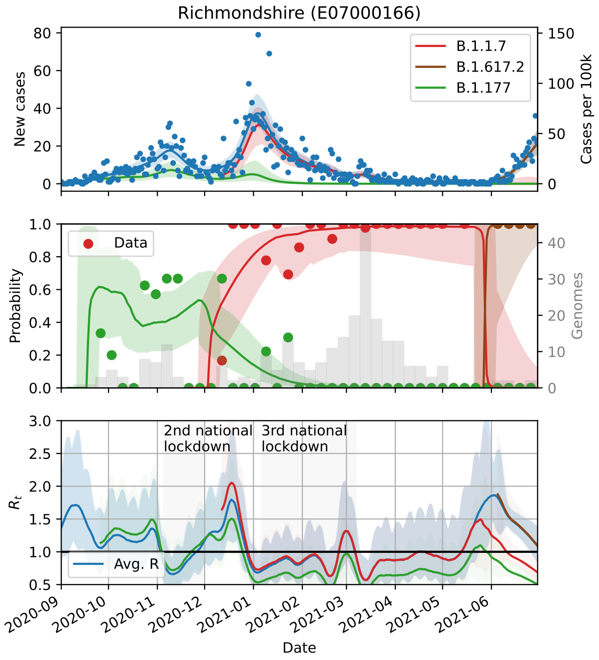
Richmondshire (E07000166)
B.1.1.7
B.1.617.2
B.1.177
New cases
Cases per 100k
Data
Probability
Genomes
2nd national lockdown
3rd national lockdown
Avg. R
$R_t$
Date

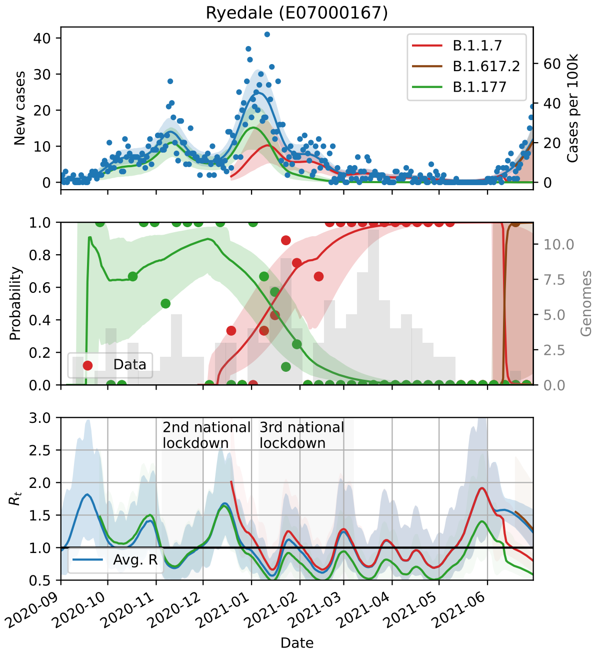
Ryedale (E07000167)
B.1.1.7
B.1.617.2
B.1.177
New cases
Cases per 100k
Data
Probability
Genomes
2nd national lockdown
3rd national lockdown
Avg. R
$R_t$
Date

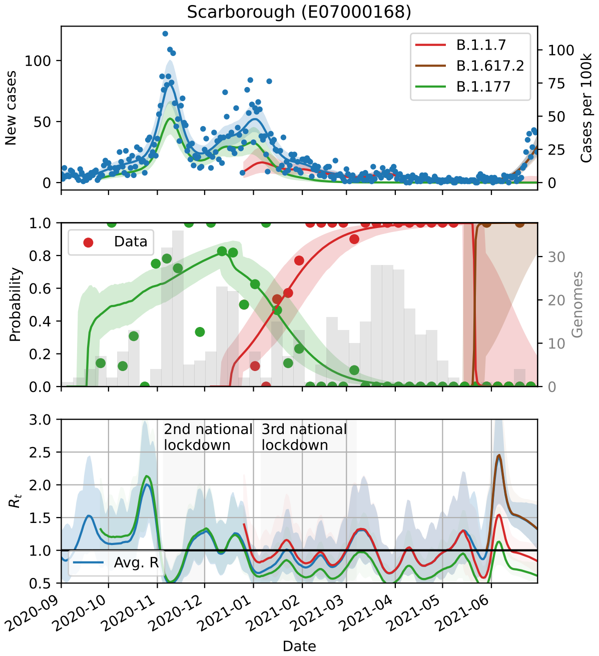
Scarborough (E07000168)
New cases
Cases per 100k
B.1.1.7
B.1.617.2
B.1.177
Probability
Genomes
Data
$R_t$
2nd national lockdown
3rd national lockdown
Avg. R
Date

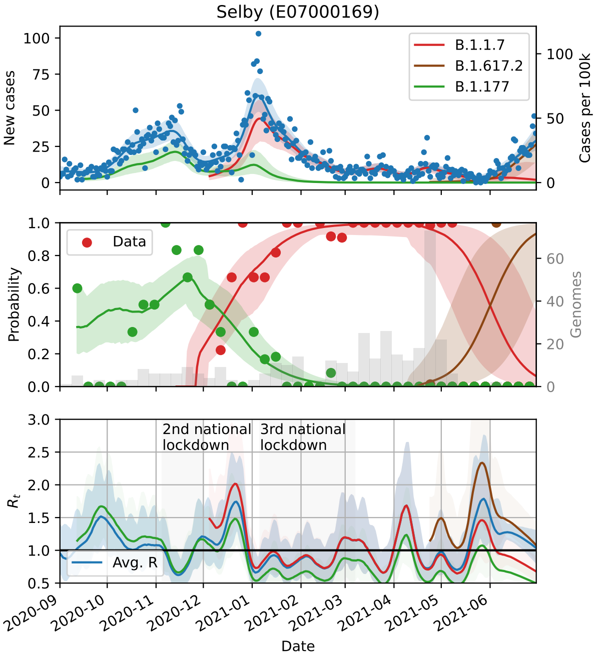
Selby (E07000169)
New cases
Cases per 100k
B.1.1.7
B.1.617.2
B.1.177
Probability
Genomes
Data
$R_t$
2nd national lockdown
3rd national lockdown
Avg. R
Date

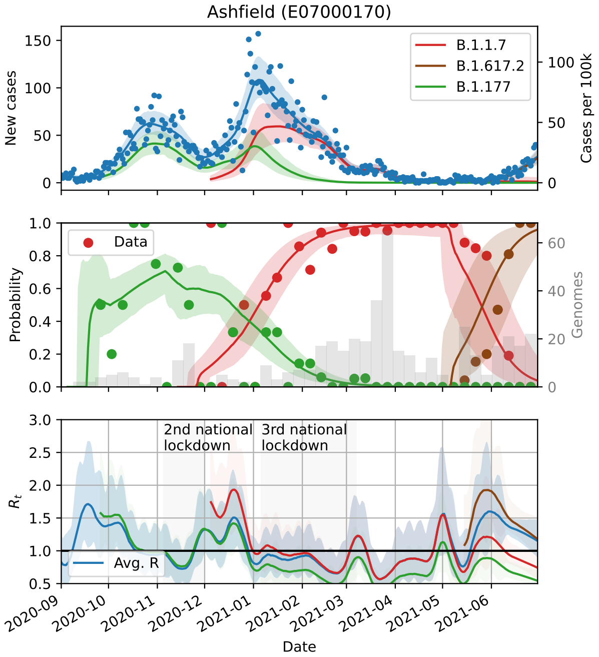
Ashfield (E07000170)
New cases
Cases per 100k
B.1.1.7
B.1.617.2
B.1.177
Probability
Genomes
Data
$R_t$
2nd national lockdown
3rd national lockdown
Avg. R
Date

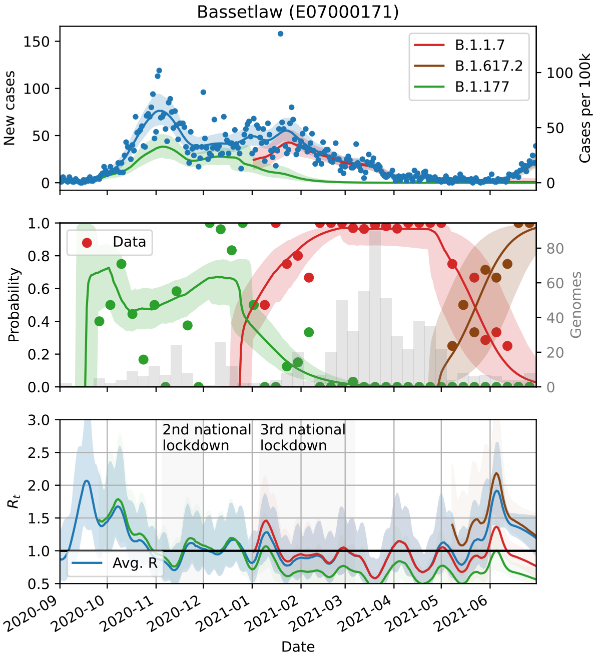
Bassetlaw (E07000171)
New cases
Cases per 100k
B.1.1.7
B.1.617.2
B.1.177
Probability
Genomes
Data
$R_t$
2nd national lockdown
3rd national lockdown
Avg. R
Date

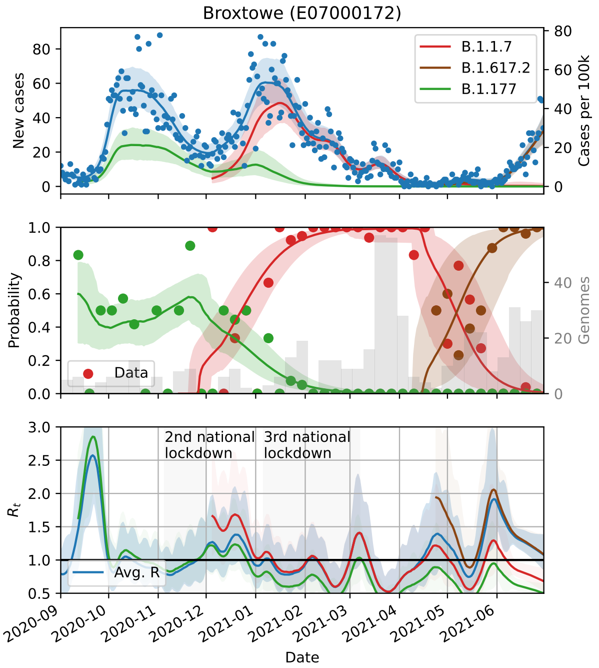
Broxtowe (E07000172)
B.1.1.7
B.1.617.2
B.1.177
New cases
Cases per 100k
Data
Probability
Genomes
2nd national lockdown
3rd national lockdown
Avg. R
$R_t$
Date

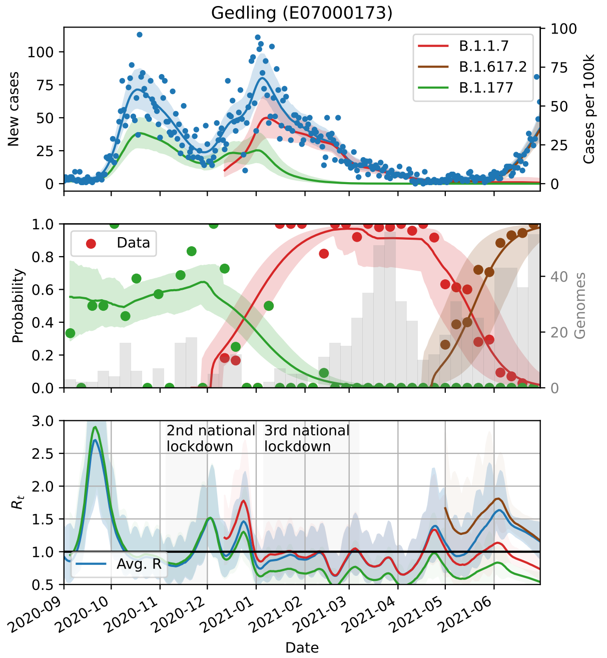
Gedling (E07000173)
B.1.1.7
B.1.617.2
B.1.177
New cases
Cases per 100k
Data
Probability
Genomes
2nd national lockdown
3rd national lockdown
Avg. R
$R_t$
Date

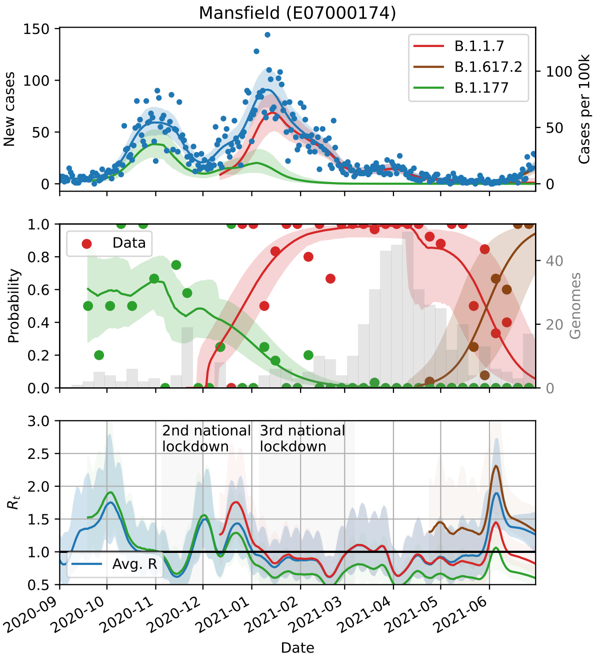
Mansfield (E07000174)
B.1.1.7
B.1.617.2
B.1.177
New cases
Cases per 100k
Data
Probability
Genomes
2nd national lockdown
3rd national lockdown
Avg. R
$R_t$
Date

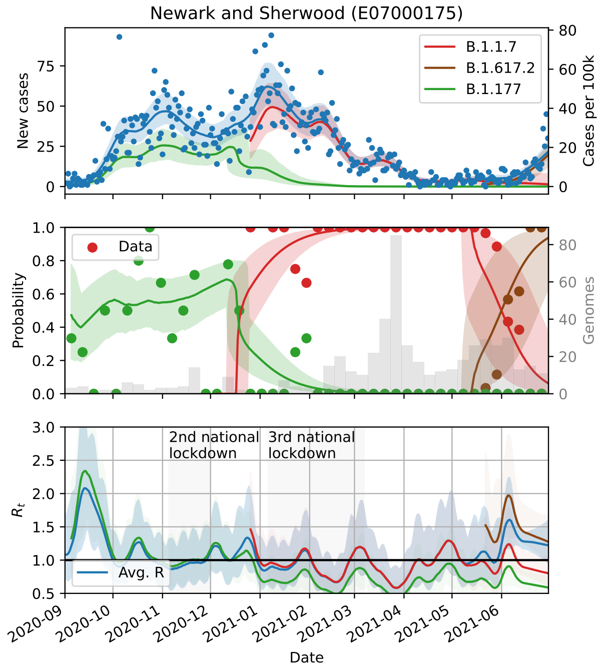
Newark and Sherwood (E07000175)
B.1.1.7
B.1.617.2
B.1.177
New cases
Cases per 100k
Data
Probability
Genomes
2nd national lockdown
3rd national lockdown
Avg. R
$R_t$
Date

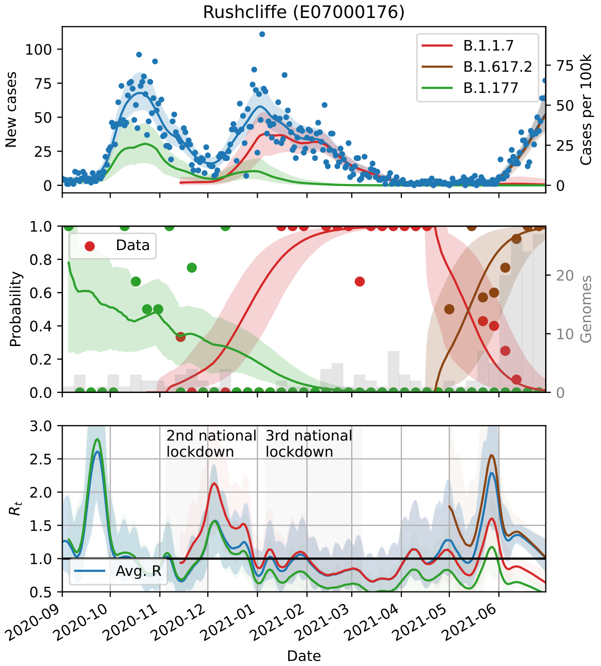
Rushcliffe (E07000176)
New cases
Cases per 100k
B.1.1.7
B.1.617.2
B.1.177
Probability
Genomes
Data
$R_t$
Avg. R
2nd national lockdown
3rd national lockdown
Date

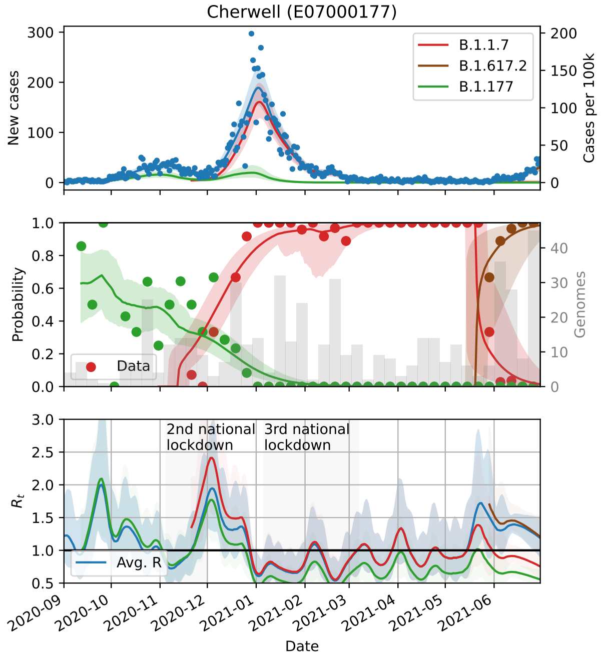
Cherwell (E07000177)
New cases
Cases per 100k
B.1.1.7
B.1.617.2
B.1.177
Probability
Genomes
Data
$R_t$
Avg. R
2nd national lockdown
3rd national lockdown
Date

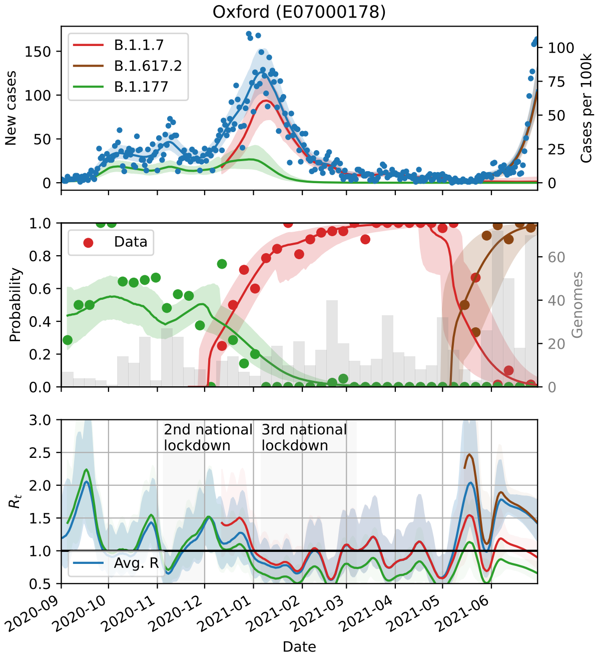
Oxford (E07000178)
New cases
Cases per 100k
B.1.1.7
B.1.617.2
B.1.177
Probability
Genomes
Data
$R_t$
Avg. R
2nd national lockdown
3rd national lockdown
Date

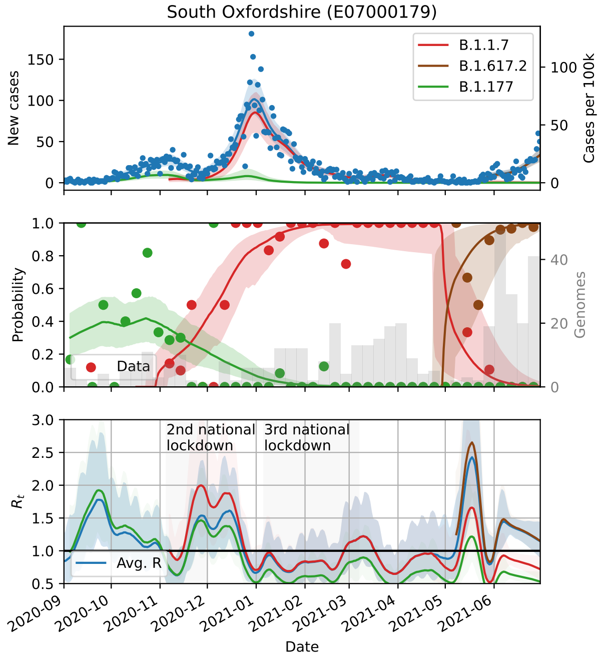
South Oxfordshire (E07000179)
New cases
Cases per 100k
B.1.1.7
B.1.617.2
B.1.177
Probability
Genomes
Data
$R_t$
Avg. R
2nd national lockdown
3rd national lockdown
Date

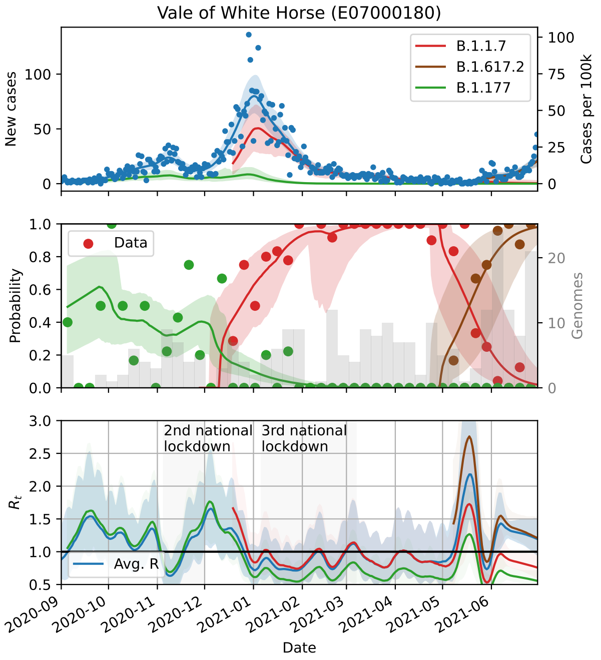
Vale of White Horse (E07000180)
New cases
Cases per 100k
B.1.1.7
B.1.617.2
B.1.177
Data
Probability
Genomes
2nd national lockdown
3rd national lockdown
$R_t$
Avg. R
Date

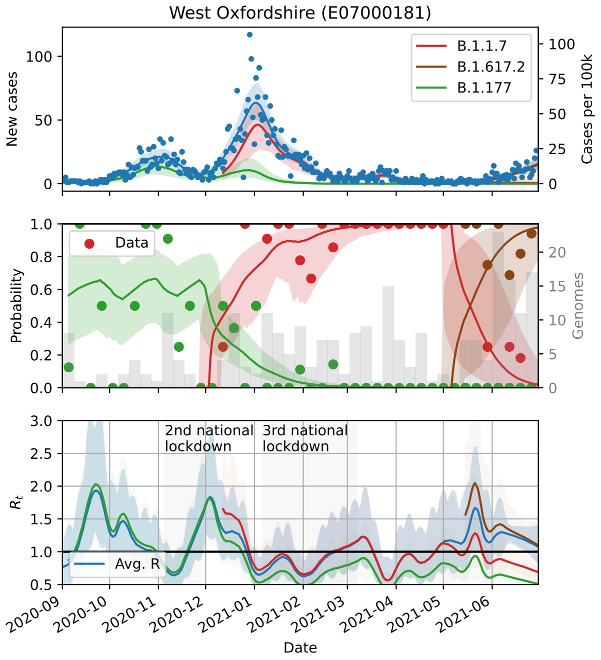
West Oxfordshire (E07000181)
New cases
Cases per 100k
B.1.1.7
B.1.617.2
B.1.177
Data
Probability
Genomes
2nd national lockdown
3rd national lockdown
$R_t$
Avg. R
Date

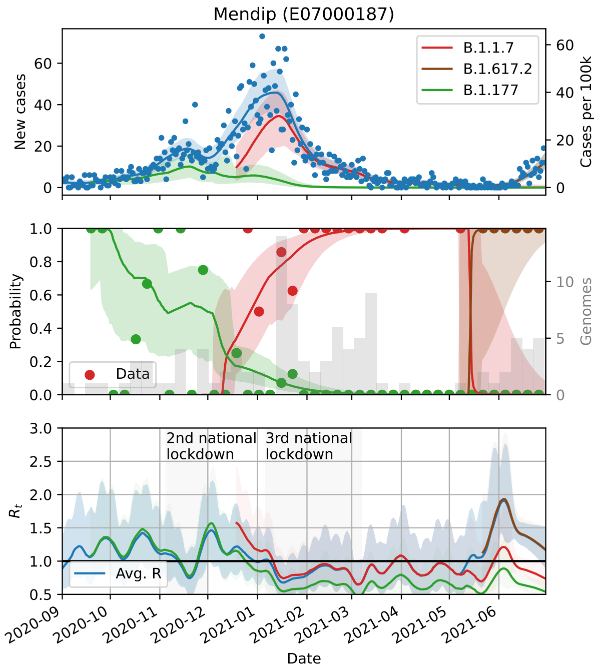
Mendip (E07000187)
New cases
Cases per 100k
B.1.1.7
B.1.617.2
B.1.177
Data
Probability
Genomes
2nd national lockdown
3rd national lockdown
$R_t$
Avg. R
Date

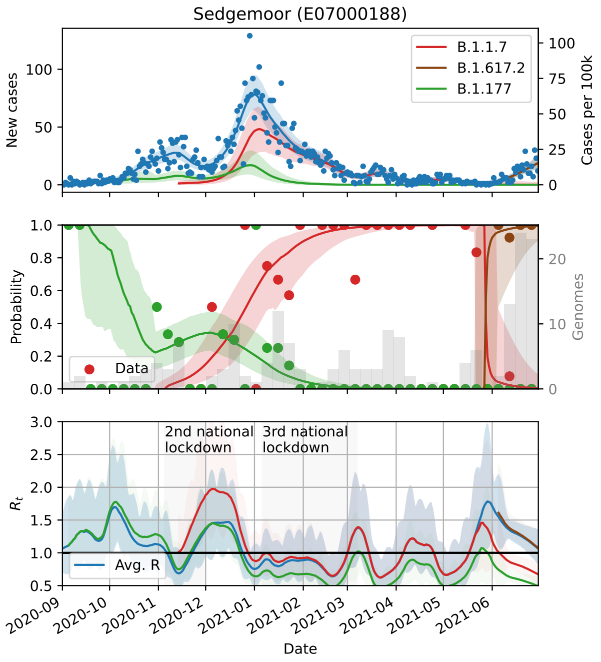
Sedgemoor (E07000188)
New cases
Cases per 100k
B.1.1.7
B.1.617.2
B.1.177
Data
Probability
Genomes
2nd national lockdown
3rd national lockdown
$R_t$
Avg. R
Date

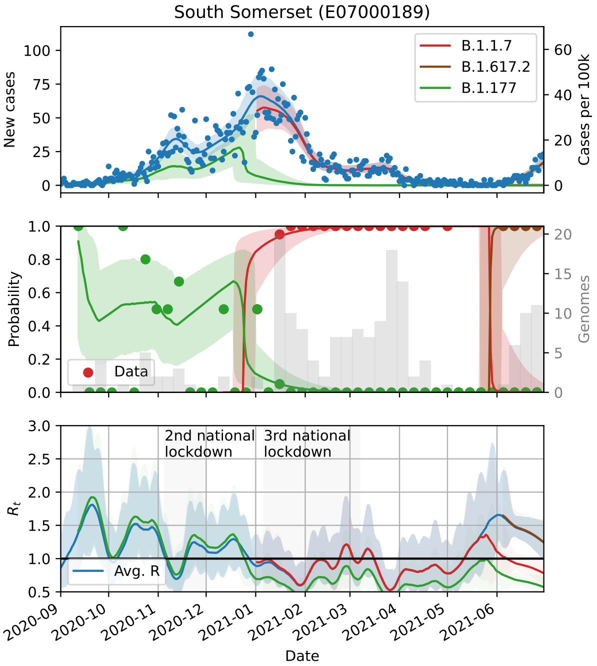
South Somerset (E07000189)
B.1.1.7
B.1.617.2
B.1.177
New cases
Cases per 100k
Probability
Genomes
Data
2nd national lockdown
3rd national lockdown
$R_t$
Avg. R
Date

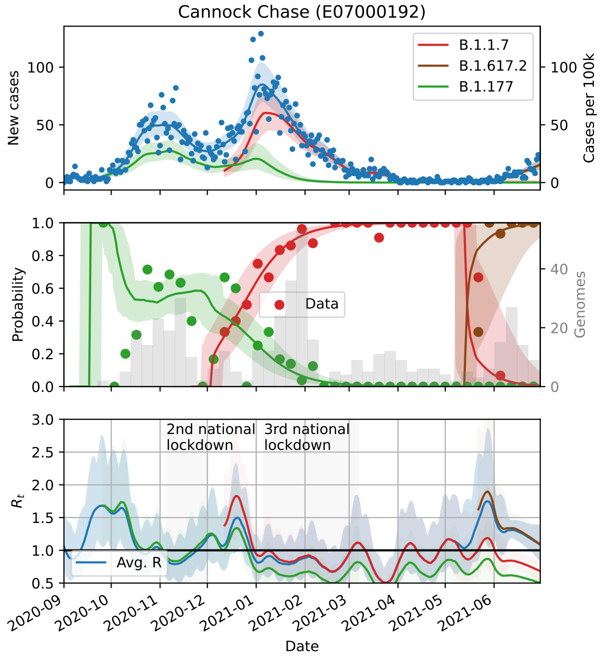
Cannock Chase (E07000192)
B.1.1.7
B.1.617.2
B.1.177
New cases
Cases per 100k
Probability
Genomes
Data
2nd national lockdown
3rd national lockdown
$R_t$
Avg. R
Date

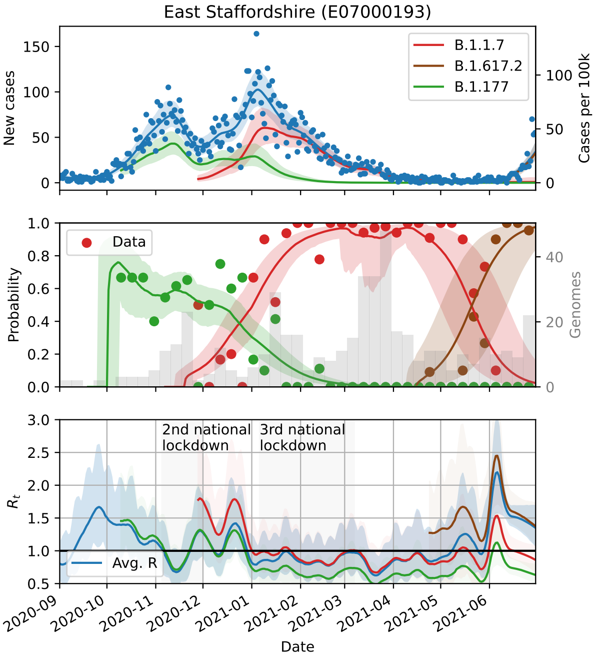
East Staffordshire (E07000193)
B.1.1.7
B.1.617.2
B.1.177
New cases
Cases per 100k
Probability
Genomes
Data
2nd national lockdown
3rd national lockdown
$R_t$
Avg. R
Date

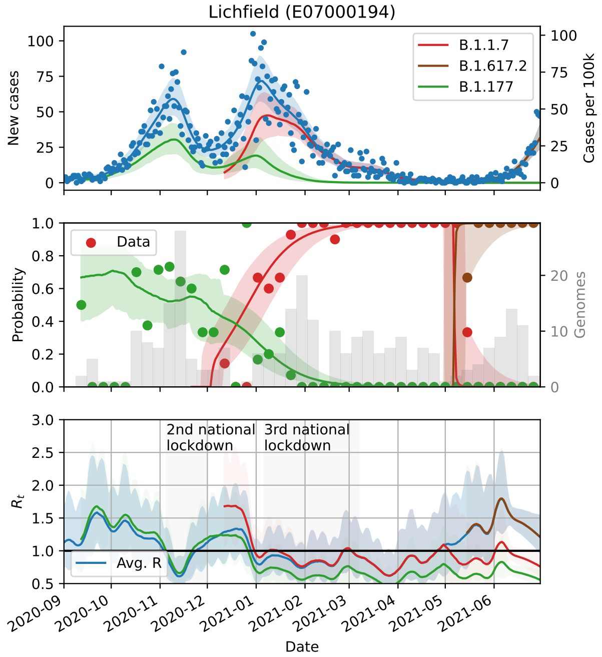
Lichfield (E07000194)
B.1.1.7
B.1.617.2
B.1.177
New cases
Cases per 100k
Probability
Genomes
Data
2nd national lockdown
3rd national lockdown
$R_t$
Avg. R
Date

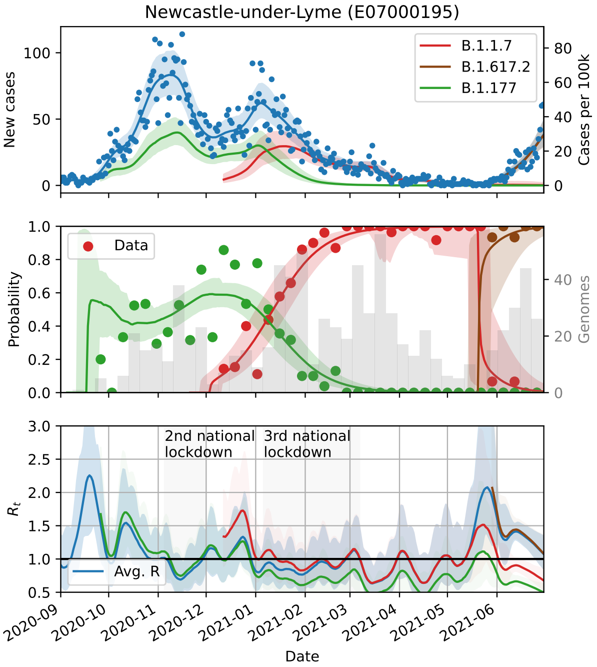
Newcastle-under-Lyme (E07000195)
New cases
Cases per 100k
B.1.1.7
B.1.617.2
B.1.177
Data
Probability
Genomes
2nd national lockdown
3rd national lockdown
$R_t$
Avg. R
Date

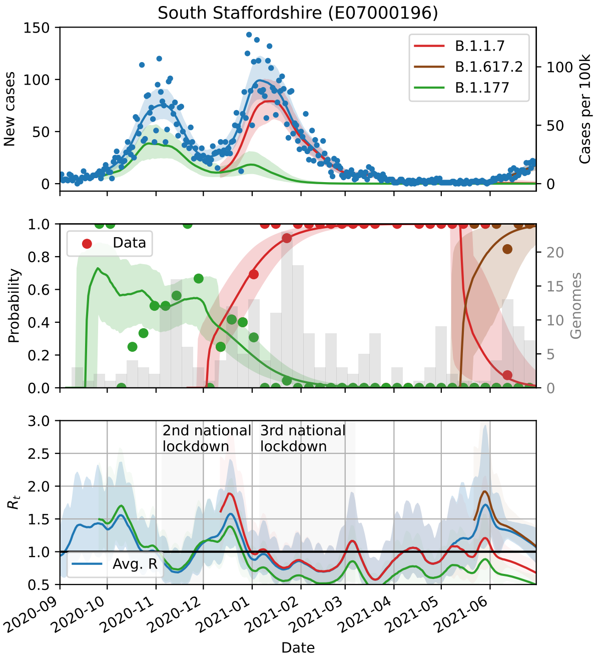
South Staffordshire (E07000196)
New cases
Cases per 100k
B.1.1.7
B.1.617.2
B.1.177
Data
Probability
Genomes
2nd national lockdown
3rd national lockdown
$R_t$
Avg. R
Date

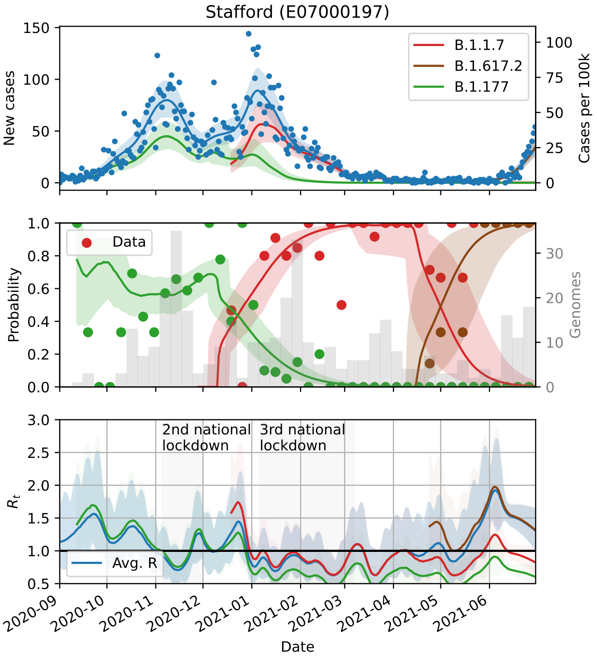
Stafford (E07000197)
New cases
Cases per 100k
B.1.1.7
B.1.617.2
B.1.177
Data
Probability
Genomes
2nd national lockdown
3rd national lockdown
$R_t$
Avg. R
Date

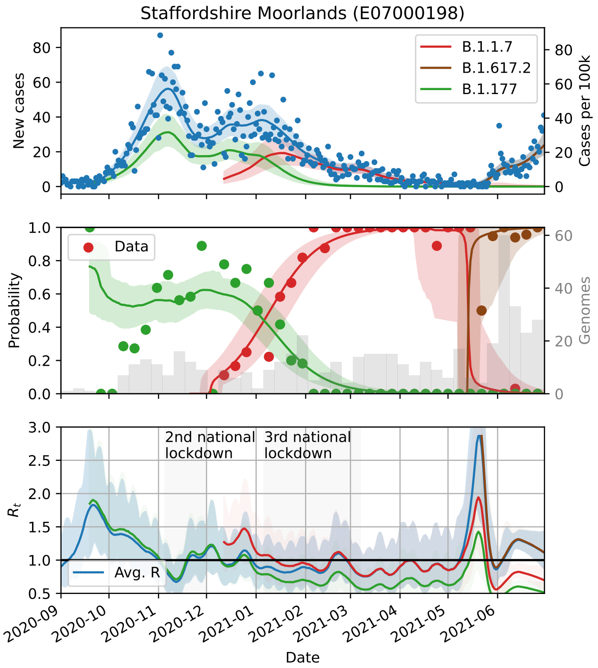
Staffordshire Moorlands (E07000198)
New cases
Cases per 100k
B.1.1.7
B.1.617.2
B.1.177
Data
Probability
Genomes
2nd national lockdown
3rd national lockdown
$R_t$
Avg. R
Date

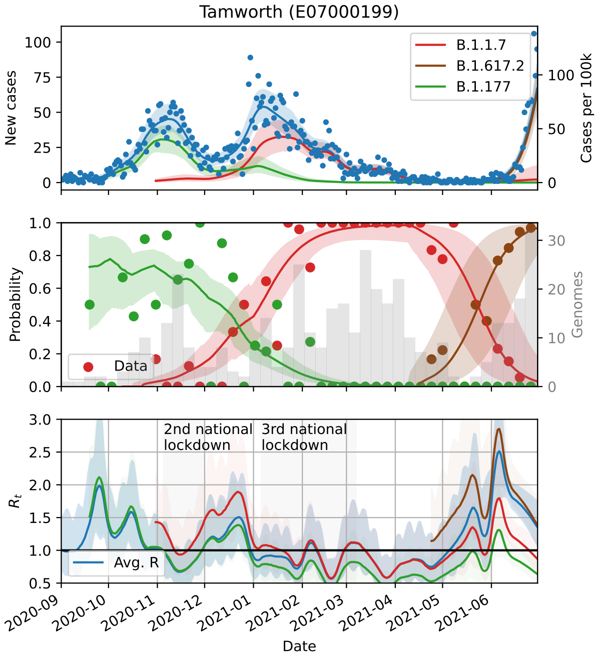
Tamworth (E07000199)
B.1.1.7
B.1.617.2
B.1.177
New cases
Cases per 100k
Data
Probability
Genomes
2nd national lockdown
3rd national lockdown
Avg. R
$R_t$
Date

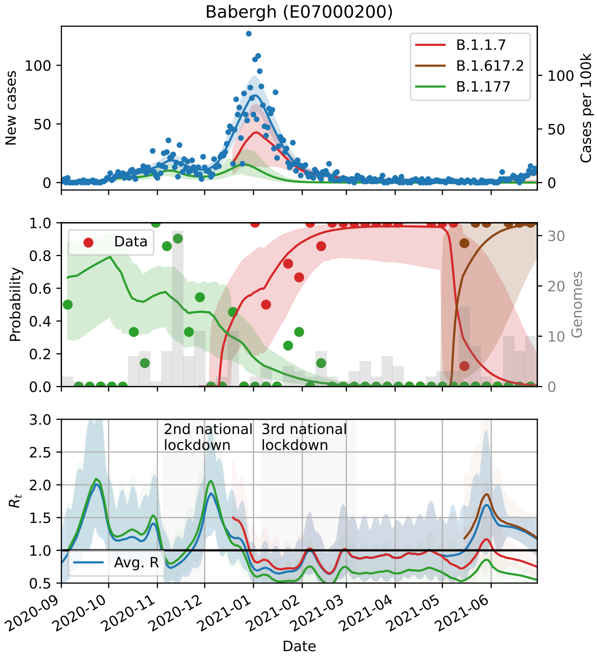
Babergh (E07000200)
B.1.1.7
B.1.617.2
B.1.177
New cases
Cases per 100k
Data
Probability
Genomes
2nd national lockdown
3rd national lockdown
Avg. R
$R_t$
Date

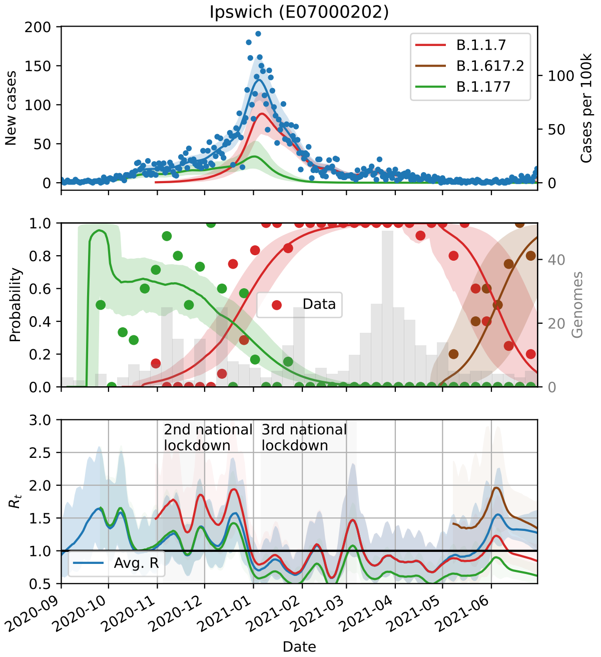
Ipswich (E07000202)
B.1.1.7
B.1.617.2
B.1.177
New cases
Cases per 100k
Data
Probability
Genomes
2nd national lockdown
3rd national lockdown
Avg. R
$R_t$
Date

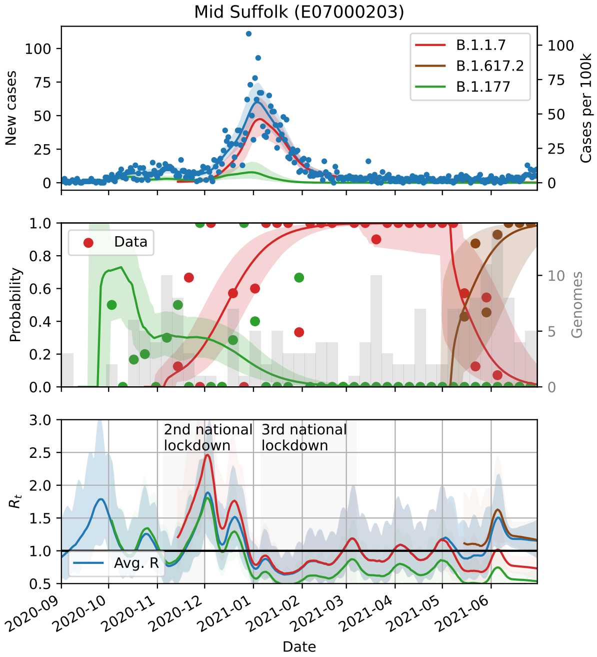
Mid Suffolk (E07000203)
B.1.1.7
B.1.617.2
B.1.177
New cases
Cases per 100k
Data
Probability
Genomes
2nd national lockdown
3rd national lockdown
Avg. R
$R_t$
Date

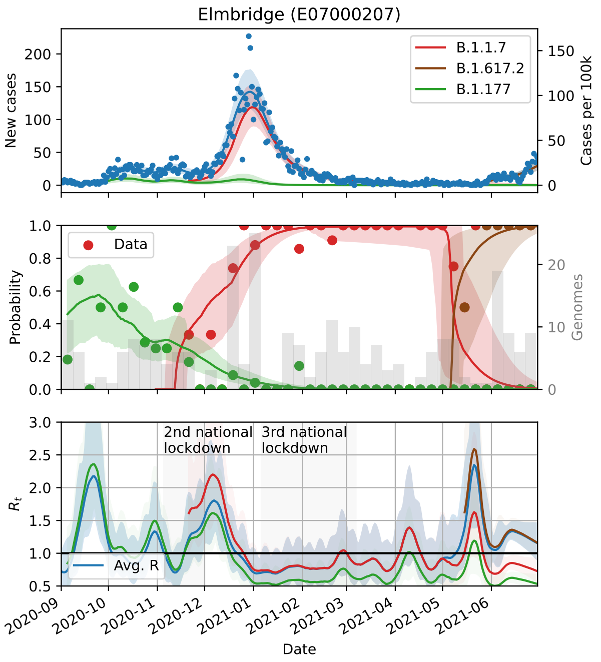
Elmbridge (E07000207)
B.1.1.7
B.1.617.2
B.1.177
New cases
Cases per 100k
Data
Probability
Genomes
2nd national lockdown
3rd national lockdown
Avg. R
$R_t$
Date

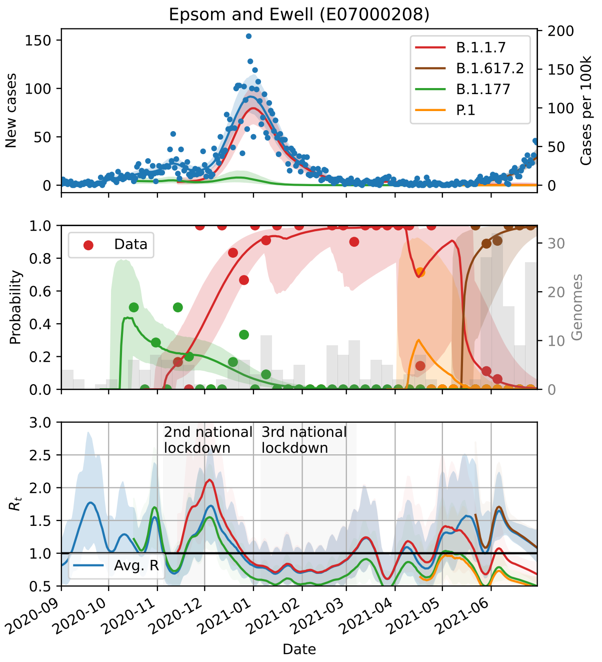
Epsom and Ewell (E07000208)
B.1.1.7
B.1.617.2
B.1.177
P.1
New cases
Cases per 100k
Data
Probability
Genomes
2nd national lockdown
3rd national lockdown
Avg. R
$R_t$
Date

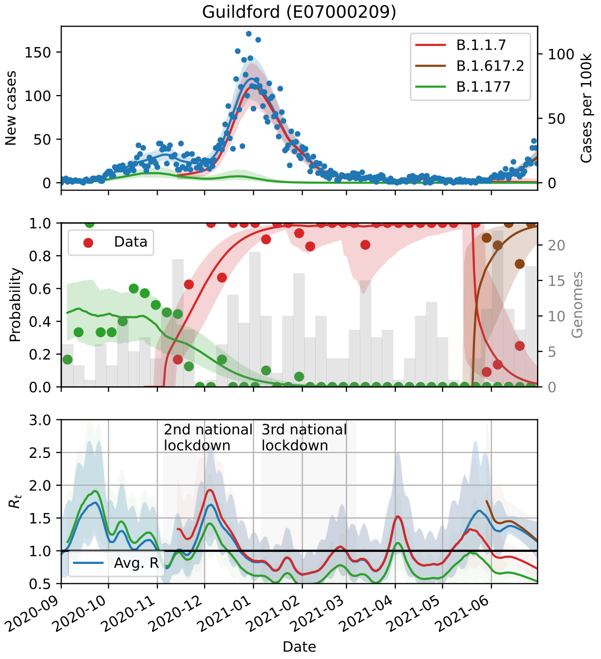
Guildford (E07000209)
B.1.1.7
B.1.617.2
B.1.177
New cases
Cases per 100k
Data
Probability
Genomes
2nd national lockdown
3rd national lockdown
Avg. R
$R_t$
Date

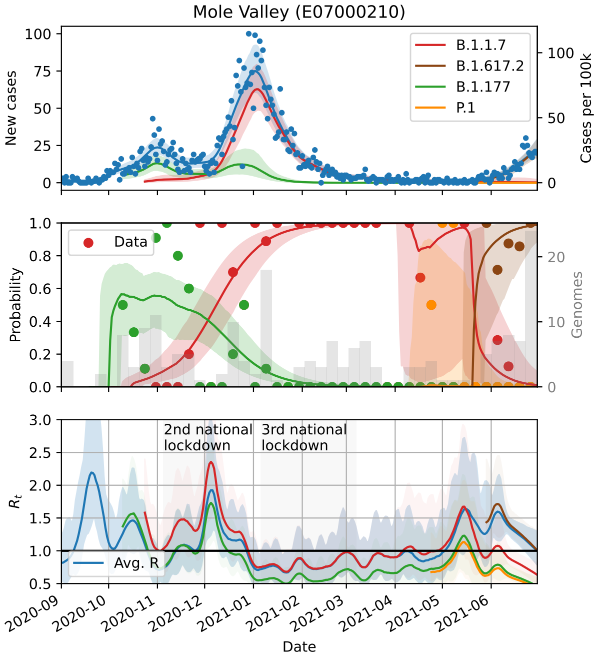
Mole Valley (E07000210)
B.1.1.7
B.1.617.2
B.1.177
P.1
New cases
Cases per 100k
Data
Probability
Genomes
2nd national lockdown
3rd national lockdown
Avg. R
$R_t$
Date

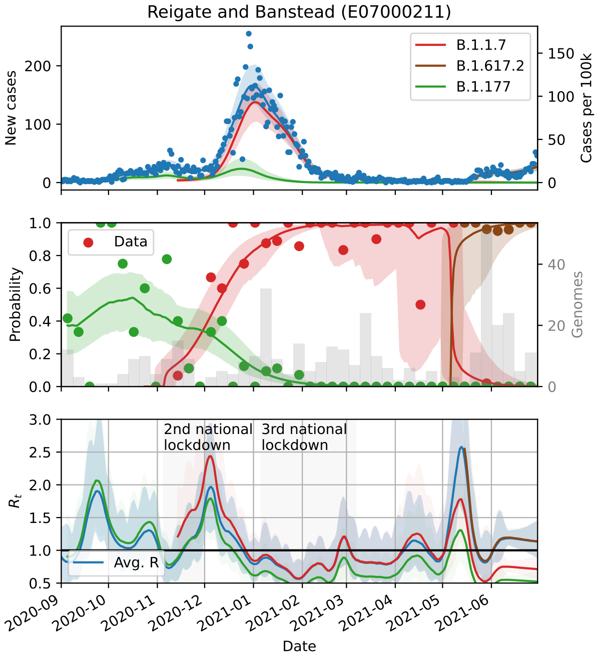
Reigate and Banstead (E07000211)
B.1.1.7
B.1.617.2
B.1.177
New cases
Cases per 100k
Data
Probability
Genomes
2nd national lockdown
3rd national lockdown
Avg. R
$R_t$
2020-09
2020-10
2020-11
2020-12
2021-01
2021-02
2021-03
2021-04
2021-05
2021-06
Date

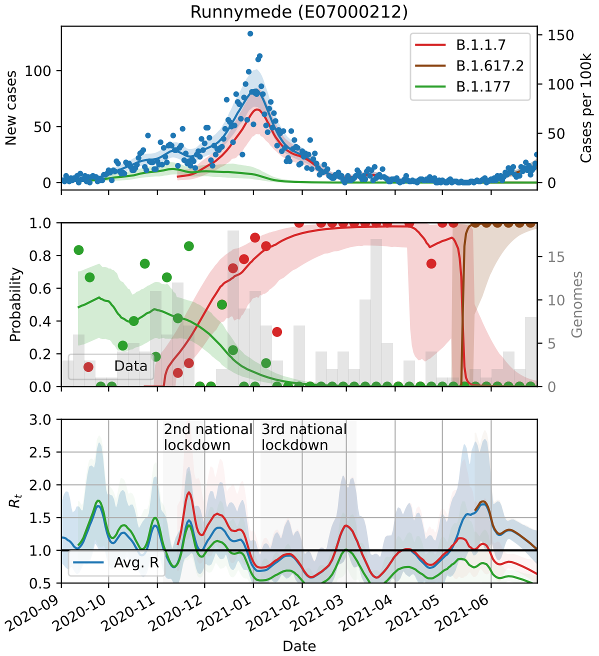
Runnymede (E07000212)
B.1.1.7
B.1.617.2
B.1.177
New cases
Cases per 100k
Data
Probability
Genomes
2nd national lockdown
3rd national lockdown
Avg. R
$R_t$
2020-09
2020-10
2020-11
2020-12
2021-01
2021-02
2021-03
2021-04
2021-05
2021-06
Date

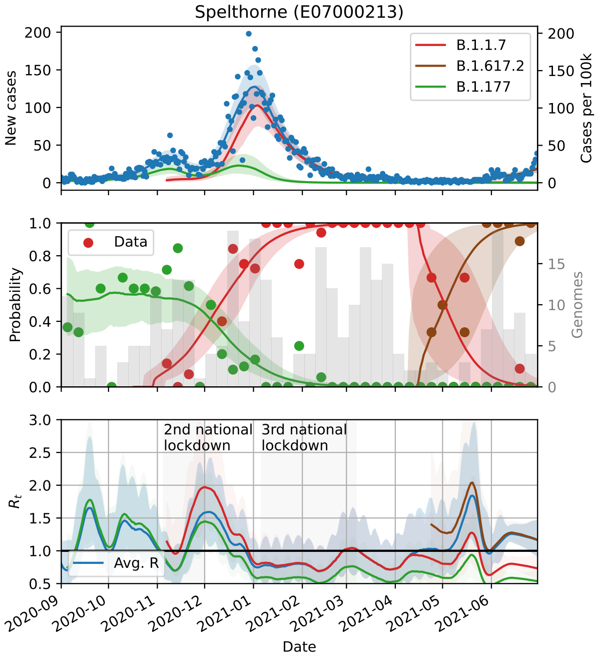
Spelthorne (E07000213)
B.1.1.7
B.1.617.2
B.1.177
New cases
Cases per 100k
Data
Probability
Genomes
2nd national lockdown
3rd national lockdown
Avg. R
$R_t$
2020-09
2020-10
2020-11
2020-12
2021-01
2021-02
2021-03
2021-04
2021-05
2021-06
Date

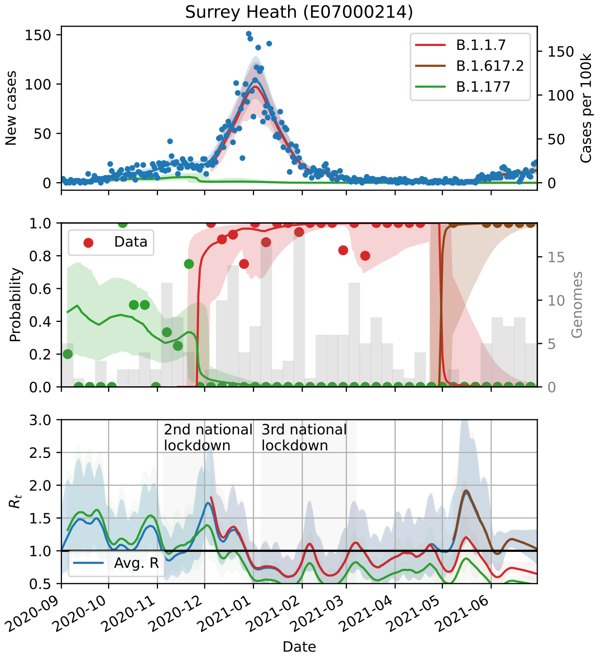
Surrey Heath (E07000214)
B.1.1.7
B.1.617.2
B.1.177
New cases
Cases per 100k
Data
Probability
Genomes
2nd national lockdown
3rd national lockdown
Avg. R
$R_t$
2020-09
2020-10
2020-11
2020-12
2021-01
2021-02
2021-03
2021-04
2021-05
2021-06
Date

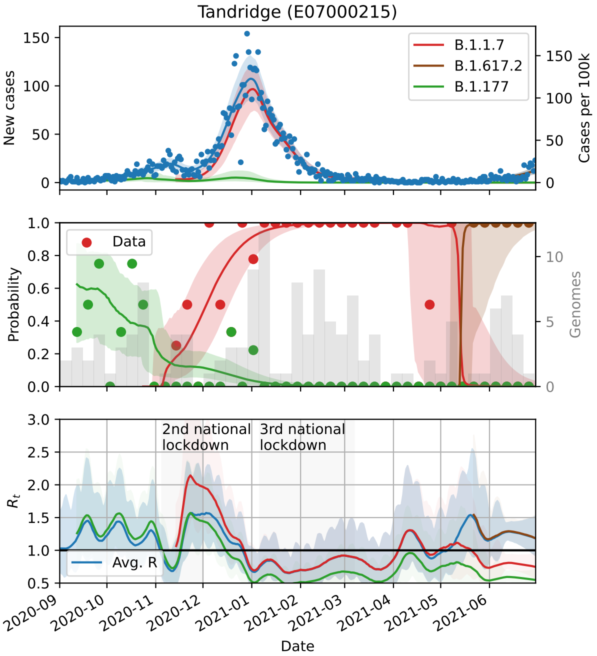
Tandridge (E07000215)
B.1.1.7
B.1.617.2
B.1.177
New cases
Cases per 100k
Data
Probability
Genomes
2nd national lockdown
3rd national lockdown
Avg. R
$R_t$
Date

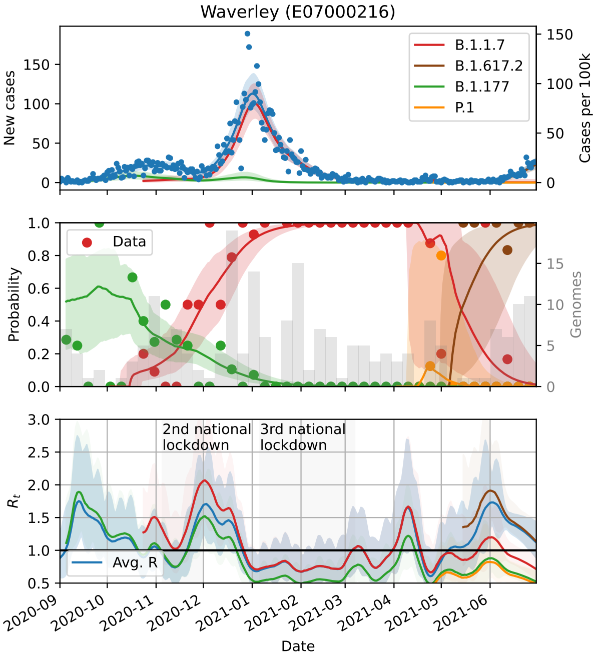
Waverley (E07000216)
B.1.1.7
B.1.617.2
B.1.177
P.1
New cases
Cases per 100k
Data
Probability
Genomes
2nd national lockdown
3rd national lockdown
Avg. R
$R_t$
Date

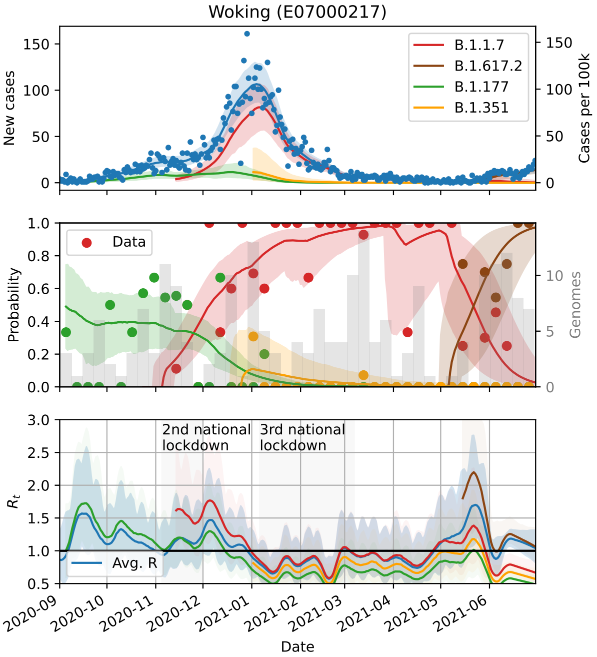
Woking (E07000217)
B.1.1.7
B.1.617.2
B.1.177
B.1.351
New cases
Cases per 100k
Data
Probability
Genomes
2nd national lockdown
3rd national lockdown
Avg. R
$R_t$
Date

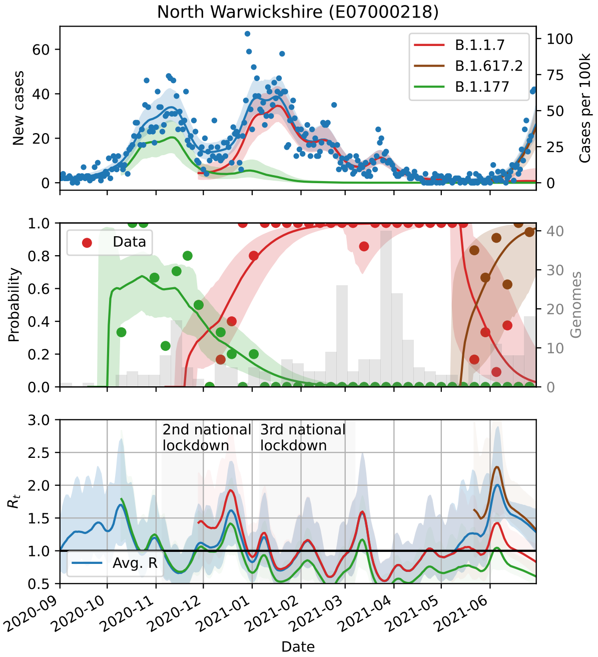
North Warwickshire (E07000218)
B.1.1.7
B.1.617.2
B.1.177
New cases
Cases per 100k
Data
Probability
Genomes
2nd national lockdown
3rd national lockdown
Avg. R
$R_t$
Date

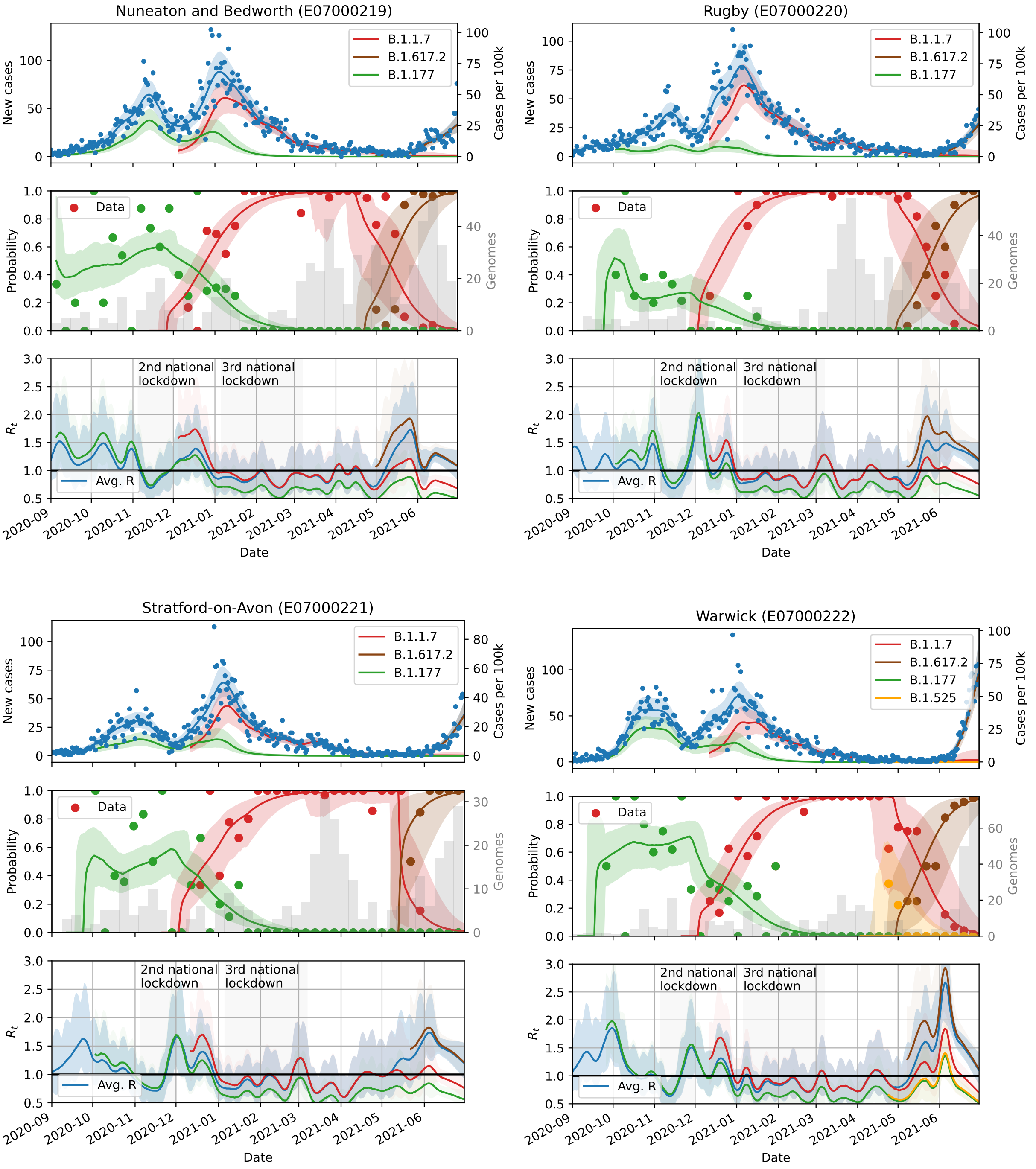
Nuneaton and Bedworth (E07000219)
Rugby (E07000220)
Stratford-on-Avon (E07000221)
Warwick (E07000222)
B.1.1.7
B.1.617.2
B.1.177
B.1.525
New cases
Cases per 100k
Data
Probability
Genomes
2nd national lockdown
3rd national lockdown
Avg. R
$R_t$
Date
2020-09
2020-10
2020-11
2020-12
2021-01
2021-02
2021-03
2021-04
2021-05
2021-06

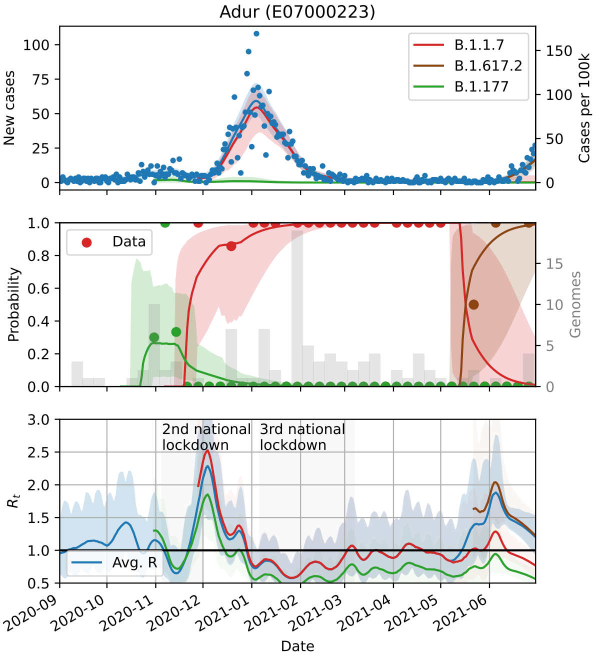
Adur (E07000223)
B.1.1.7
B.1.617.2
B.1.177
New cases
Cases per 100k
Data
Probability
Genomes
2nd national lockdown
3rd national lockdown
Avg. R
$R_t$
Date

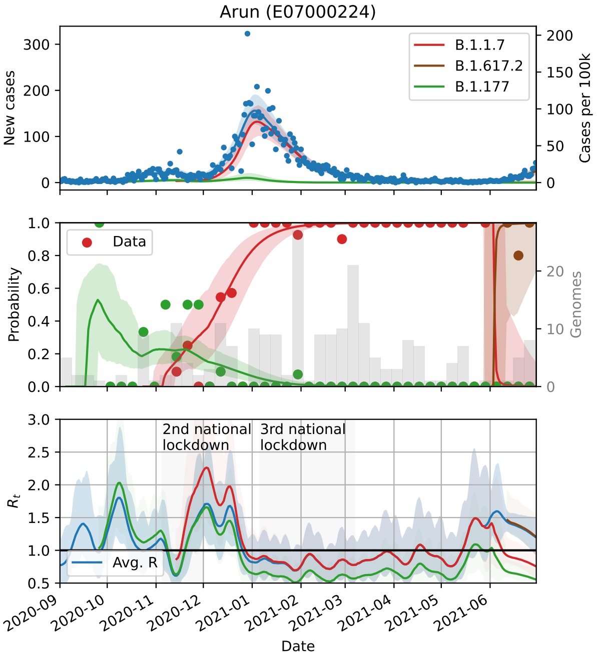
Arun (E07000224)
B.1.1.7
B.1.617.2
B.1.177
New cases
Cases per 100k
Data
Probability
Genomes
2nd national lockdown
3rd national lockdown
Avg. R
$R_t$
Date

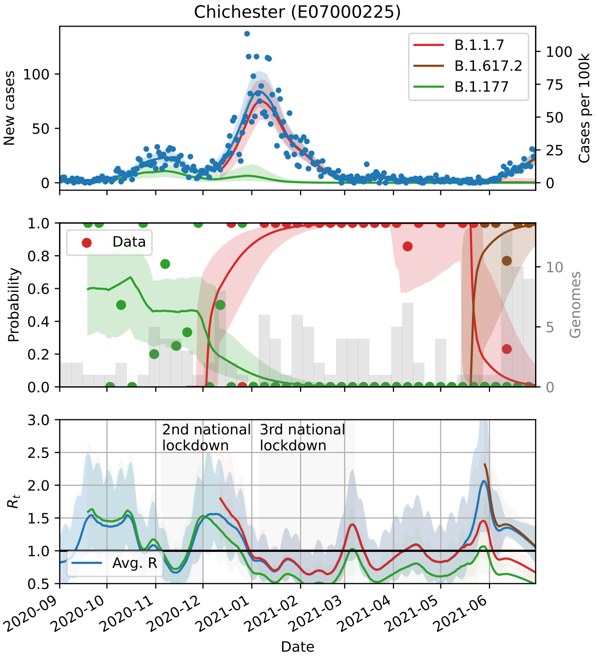
Chichester (E07000225)
B.1.1.7
B.1.617.2
B.1.177
New cases
Cases per 100k
Data
Probability
Genomes
2nd national lockdown
3rd national lockdown
Avg. R
$R_t$
Date

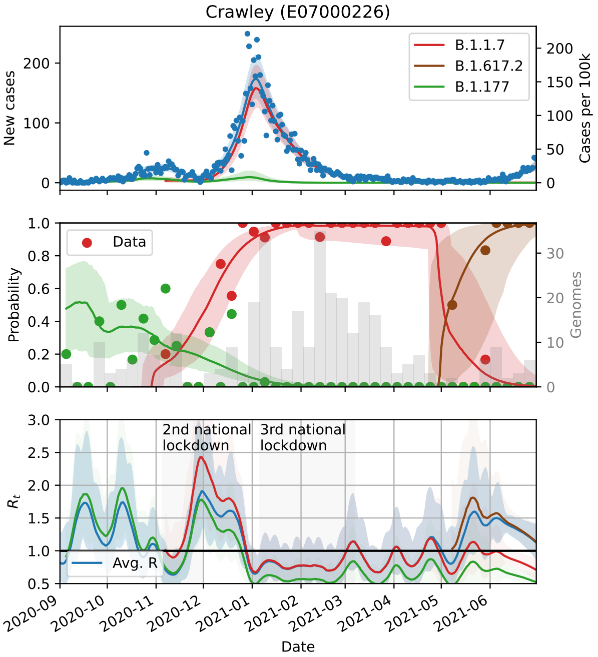
Crawley (E07000226)
B.1.1.7
B.1.617.2
B.1.177
New cases
Cases per 100k
Data
Probability
Genomes
2nd national lockdown
3rd national lockdown
Avg. R
$R_t$
Date

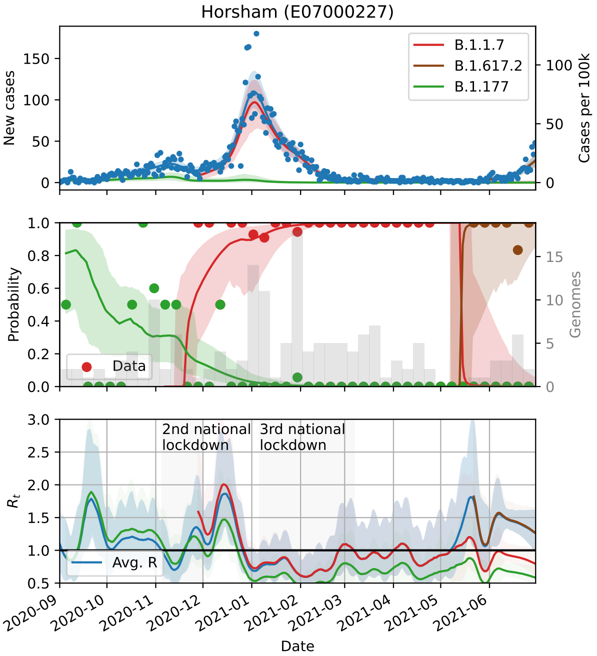

Horsham (E07000227)
New cases
Cases per 100k
B.1.1.7
B.1.617.2
B.1.177
Probability
Genomes
Data
$R_t$
2nd national lockdown
3rd national lockdown
Avg. R
Date


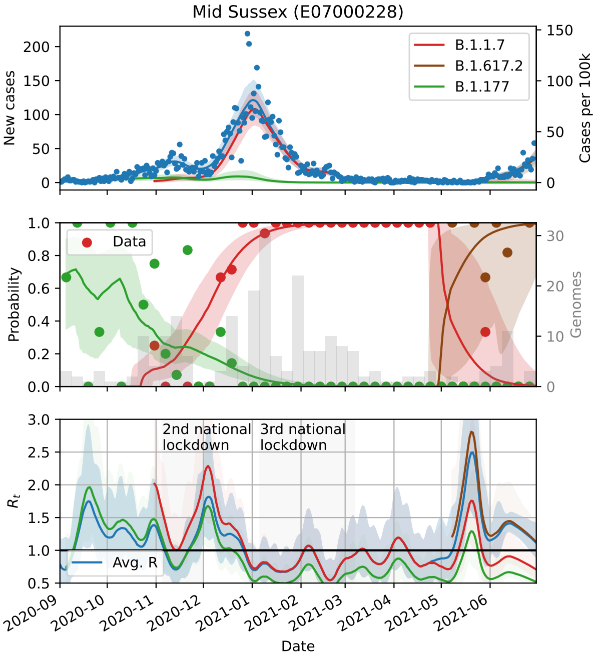

Mid Sussex (E07000228)
New cases
Cases per 100k
B.1.1.7
B.1.617.2
B.1.177
Probability
Genomes
Data
$R_t$
2nd national lockdown
3rd national lockdown
Avg. R
Date


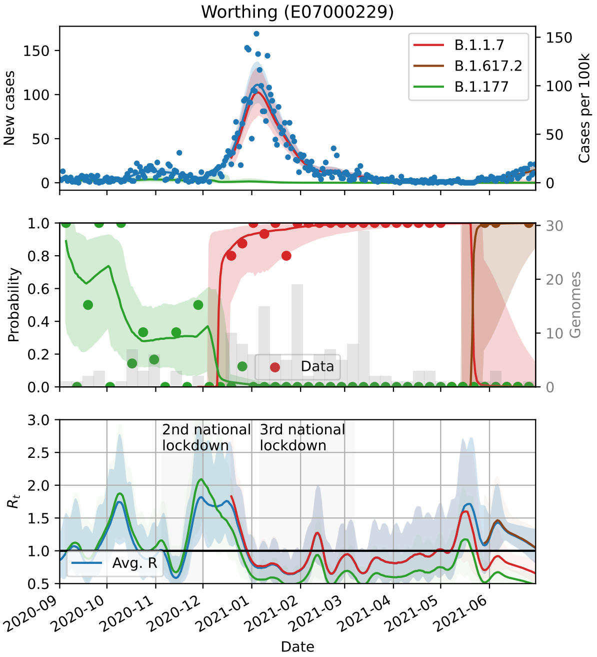

Worthing (E07000229)
New cases
Cases per 100k
B.1.1.7
B.1.617.2
B.1.177
Probability
Genomes
Data
$R_t$
2nd national lockdown
3rd national lockdown
Avg. R
Date


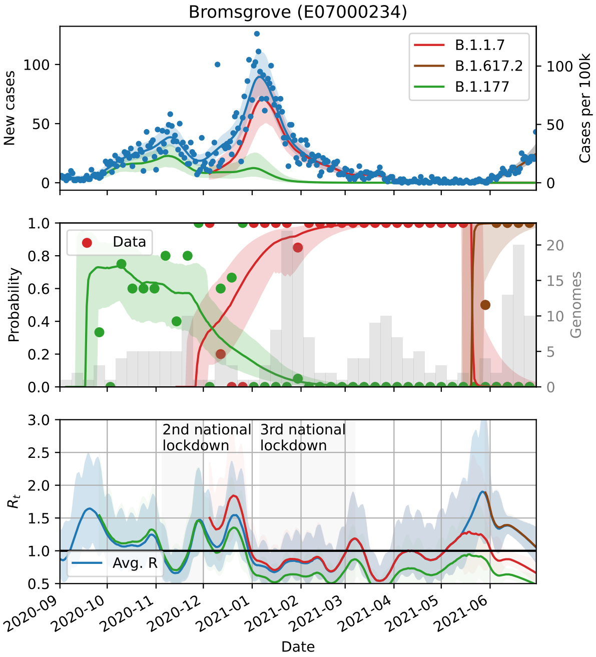

Bromsgrove (E07000234)
New cases
Cases per 100k
B.1.1.7
B.1.617.2
B.1.177
Probability
Genomes
Data
$R_t$
2nd national lockdown
3rd national lockdown
Avg. R
Date

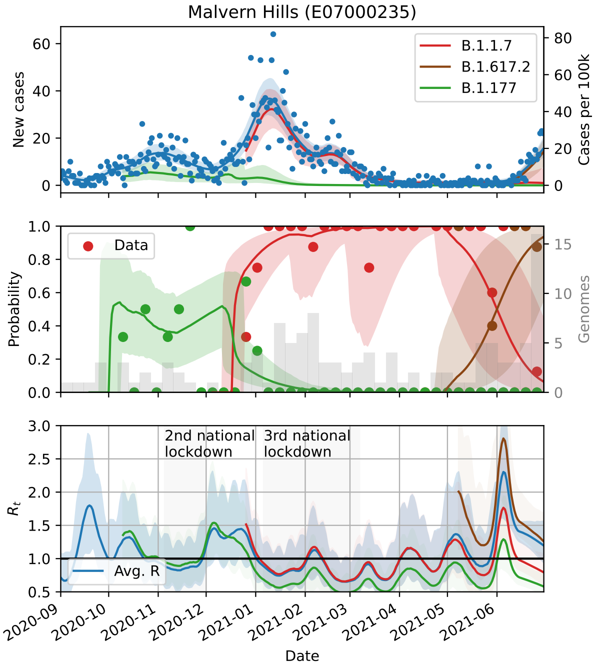
Malvern Hills (E07000235)
B.1.1.7
B.1.617.2
B.1.177
New cases
Cases per 100k
Data
Probability
Genomes
2nd national lockdown
3rd national lockdown
Avg. R
$R_t$
Date

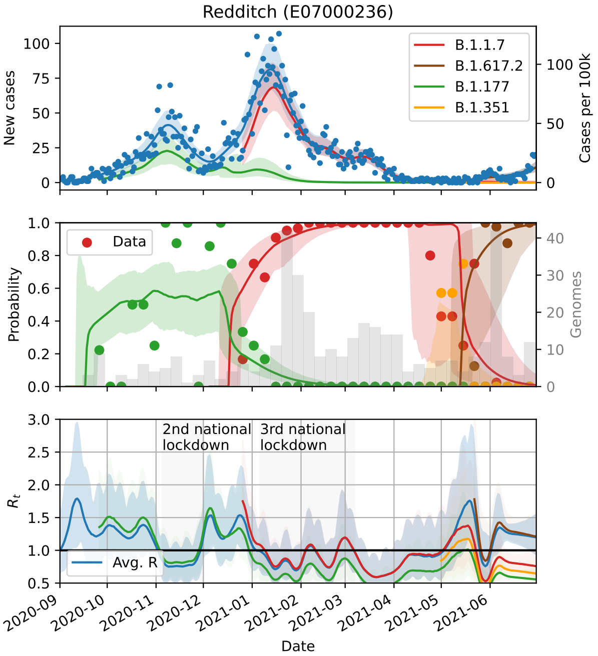
Redditch (E07000236)
B.1.1.7
B.1.617.2
B.1.177
B.1.351
New cases
Cases per 100k
Data
Probability
Genomes
2nd national lockdown
3rd national lockdown
Avg. R
$R_t$
Date

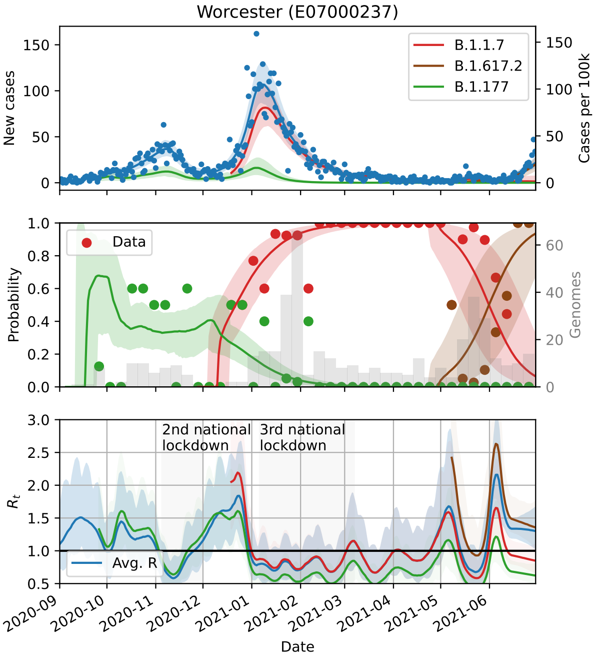
Worcester (E07000237)
B.1.1.7
B.1.617.2
B.1.177
New cases
Cases per 100k
Data
Probability
Genomes
2nd national lockdown
3rd national lockdown
Avg. R
$R_t$
Date

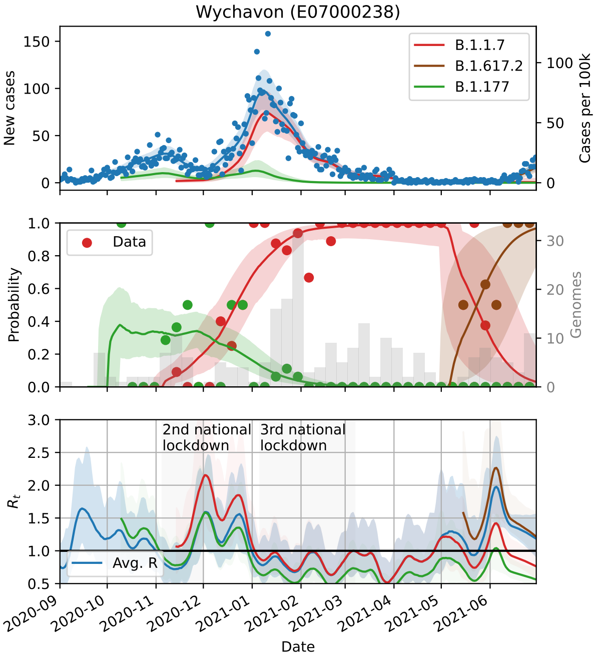
Wychavon (E07000238)
B.1.1.7
B.1.617.2
B.1.177
New cases
Cases per 100k
Data
Probability
Genomes
2nd national lockdown
3rd national lockdown
Avg. R
$R_t$
Date

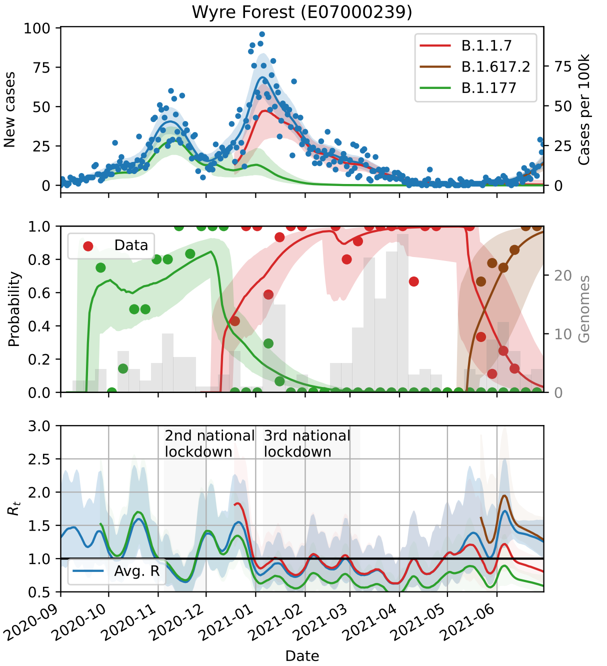
Wyre Forest (E07000239)
New cases
Cases per 100k
B.1.1.7
B.1.617.2
B.1.177
Data
Probability
Genomes
2nd national lockdown
3rd national lockdown
$R_t$
Avg. R
2020-09
2020-10
2020-11
2020-12
2021-01
2021-02
2021-03
2021-04
2021-05
2021-06
Date

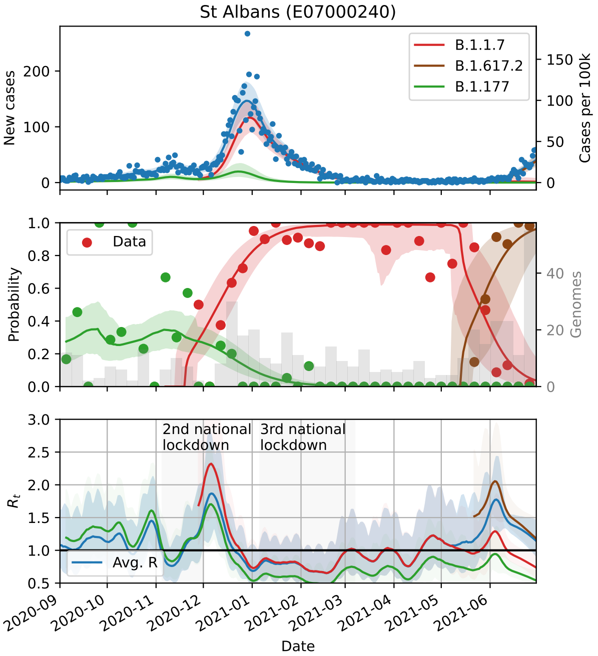
St Albans (E07000240)
New cases
Cases per 100k
B.1.1.7
B.1.617.2
B.1.177
Data
Probability
Genomes
2nd national lockdown
3rd national lockdown
$R_t$
Avg. R
2020-09
2020-10
2020-11
2020-12
2021-01
2021-02
2021-03
2021-04
2021-05
2021-06
Date

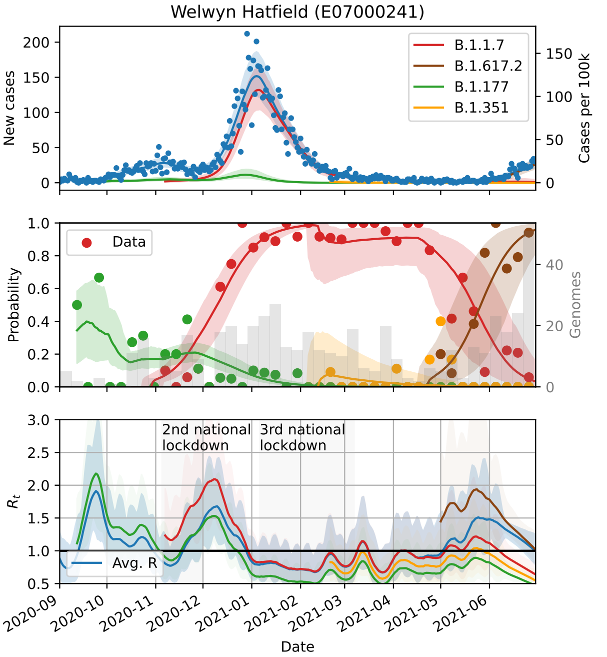
Welwyn Hatfield (E07000241)
New cases
Cases per 100k
B.1.1.7
B.1.617.2
B.1.177
B.1.351
Data
Probability
Genomes
2nd national lockdown
3rd national lockdown
$R_t$
Avg. R
2020-09
2020-10
2020-11
2020-12
2021-01
2021-02
2021-03
2021-04
2021-05
2021-06
Date

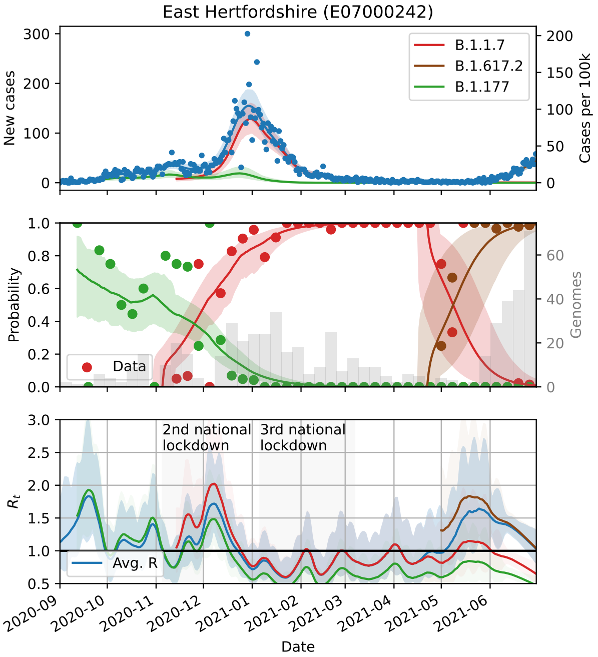
East Hertfordshire (E07000242)
New cases
Cases per 100k
B.1.1.7
B.1.617.2
B.1.177
Data
Probability
Genomes
2nd national lockdown
3rd national lockdown
$R_t$
Avg. R
2020-09
2020-10
2020-11
2020-12
2021-01
2021-02
2021-03
2021-04
2021-05
2021-06
Date

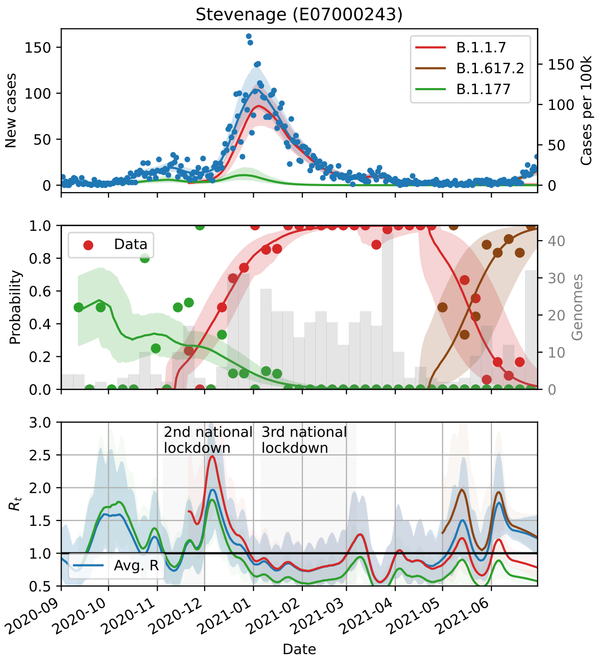
Stevenage (E07000243)
B.1.1.7
B.1.617.2
B.1.177
New cases
Cases per 100k
Data
Probability
Genomes
2nd national lockdown
3rd national lockdown
Avg. R
$R_t$
Date

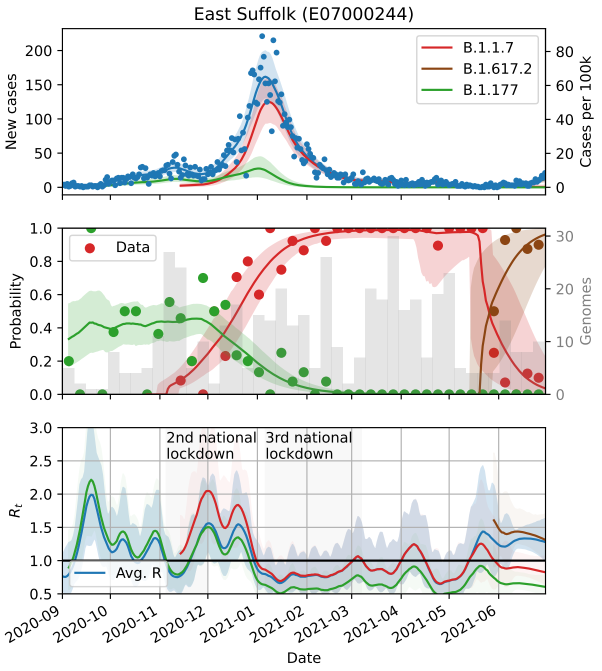
East Suffolk (E07000244)
B.1.1.7
B.1.617.2
B.1.177
New cases
Cases per 100k
Data
Probability
Genomes
2nd national lockdown
3rd national lockdown
Avg. R
$R_t$
Date

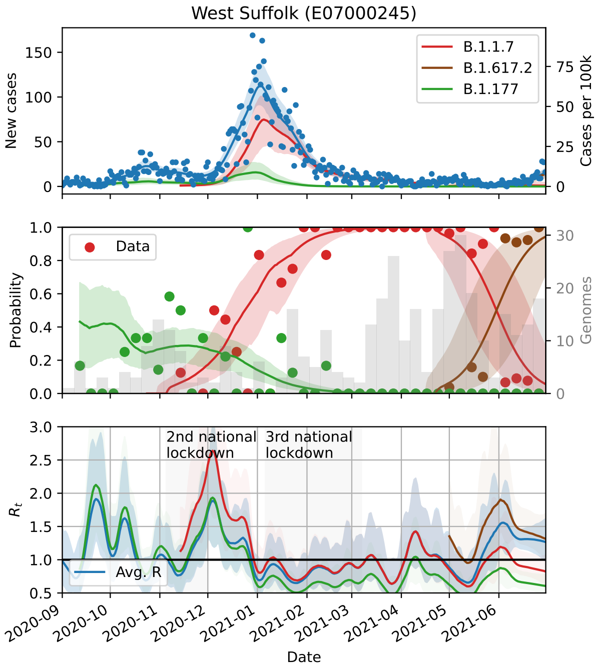
West Suffolk (E07000245)
B.1.1.7
B.1.617.2
B.1.177
New cases
Cases per 100k
Data
Probability
Genomes
2nd national lockdown
3rd national lockdown
Avg. R
$R_t$
Date

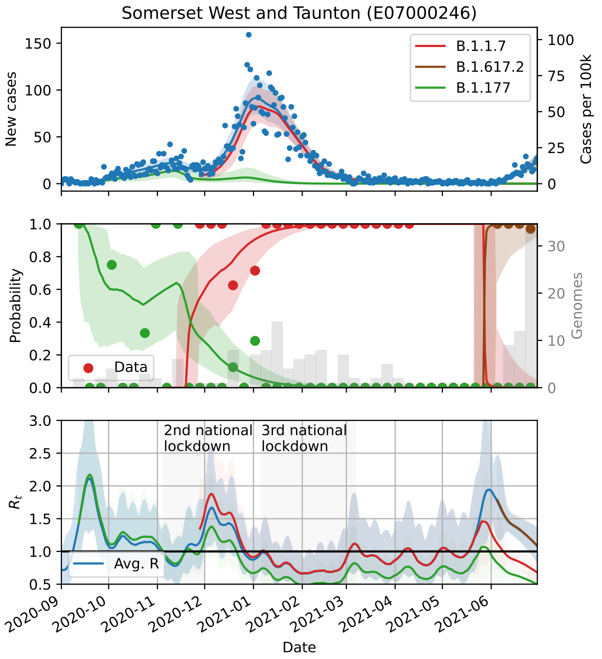
Somerset West and Taunton (E07000246)
B.1.1.7
B.1.617.2
B.1.177
New cases
Cases per 100k
Data
Probability
Genomes
2nd national lockdown
3rd national lockdown
Avg. R
$R_t$
Date

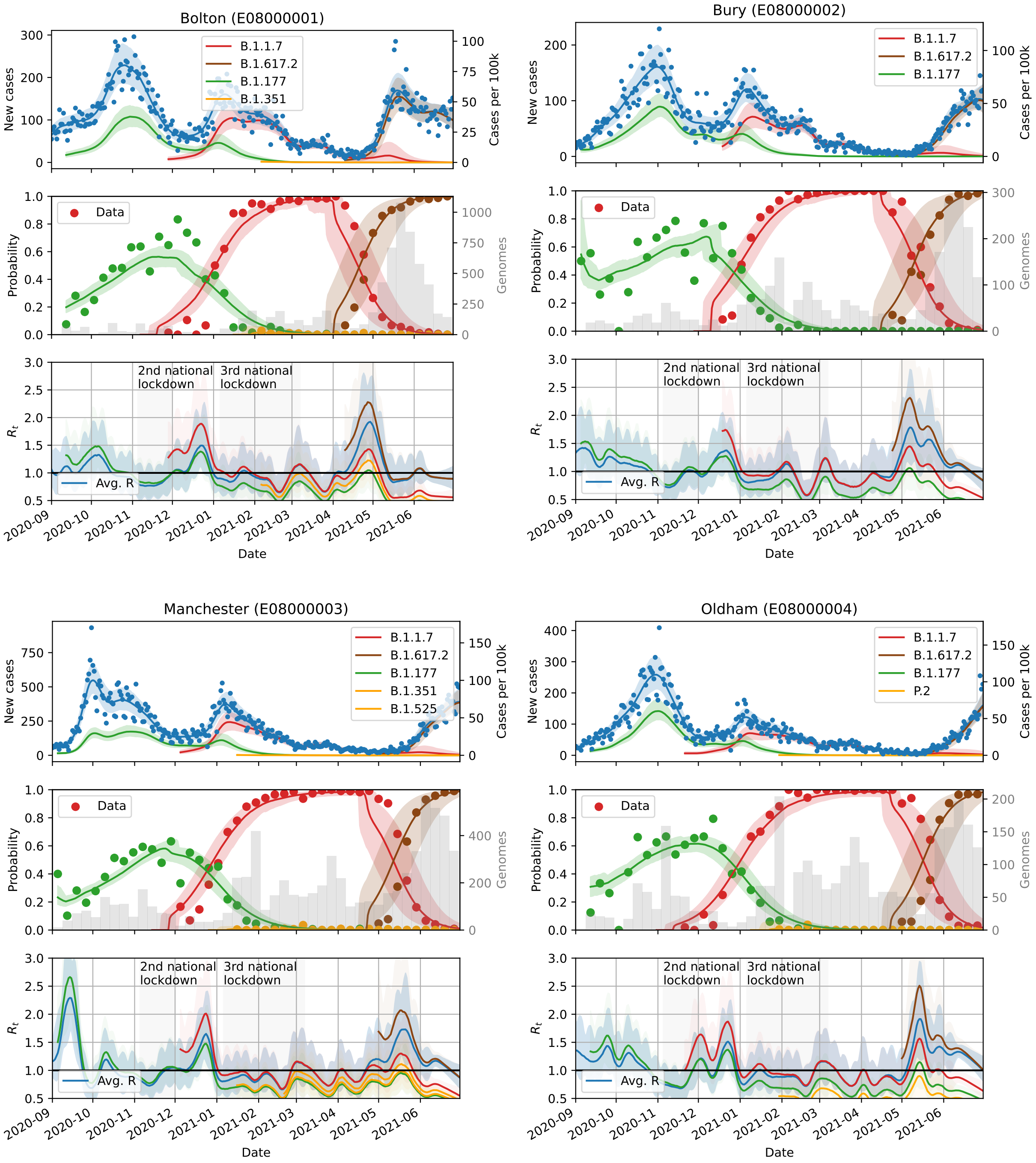
Bolton (E08000001)
B.1.1.7
B.1.617.2
B.1.177
B.1.351
New cases
Cases per 100k
Data
Probability
Genomes
2nd national lockdown
3rd national lockdown
Avg. R
$R_t$
Date
Bury (E08000002)
B.1.1.7
B.1.617.2
B.1.177
New cases
Cases per 100k
Data
Probability
Genomes
2nd national lockdown
3rd national lockdown
Avg. R
$R_t$
Date
Manchester (E08000003)
B.1.1.7
B.1.617.2
B.1.177
B.1.351
B.1.525
New cases
Cases per 100k
Data
Probability
Genomes
2nd national lockdown
3rd national lockdown
Avg. R
$R_t$
Date
Oldham (E08000004)
B.1.1.7
B.1.617.2
B.1.177
P.2
New cases
Cases per 100k
Data
Probability
Genomes
2nd national lockdown
3rd national lockdown
Avg. R
$R_t$
Date

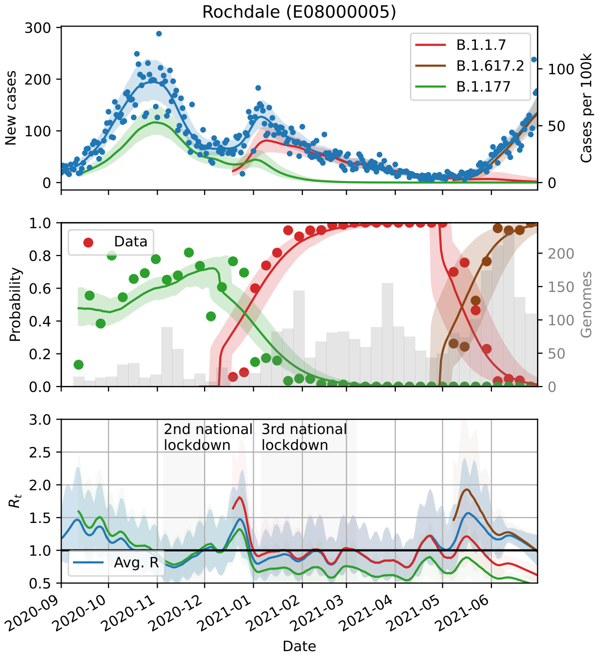
Rochdale (E08000005)
B.1.1.7
B.1.617.2
B.1.177
New cases
Cases per 100k
Data
Probability
Genomes
2nd national lockdown
3rd national lockdown
Avg. R
$R_t$
Date

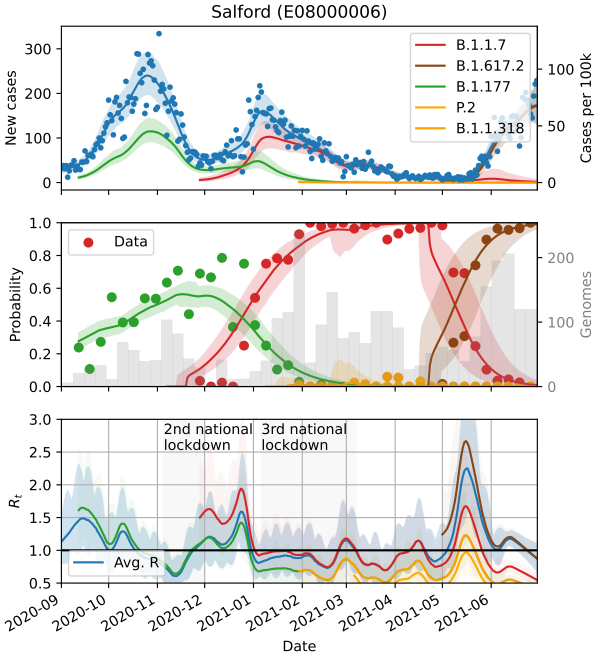
Salford (E08000006)
B.1.1.7
B.1.617.2
B.1.177
P.2
B.1.1.318
New cases
Cases per 100k
Data
Probability
Genomes
2nd national lockdown
3rd national lockdown
Avg. R
$R_t$
Date

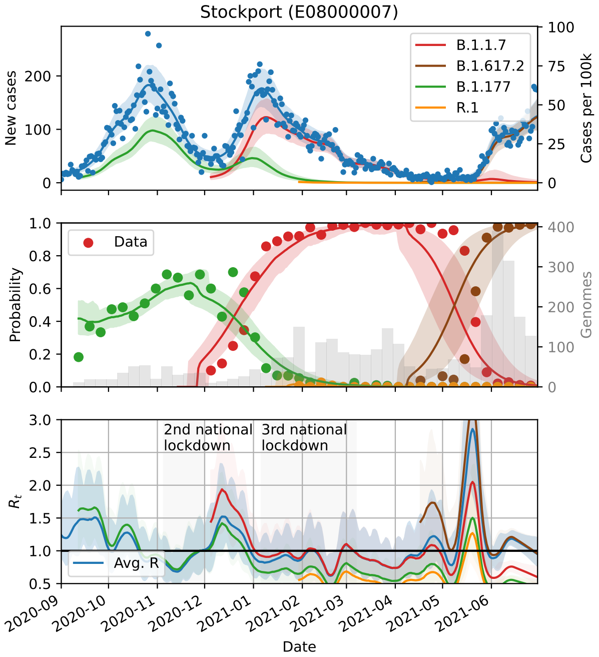
Stockport (E08000007)
B.1.1.7
B.1.617.2
B.1.177
R.1
New cases
Cases per 100k
Data
Probability
Genomes
2nd national lockdown
3rd national lockdown
Avg. R
$R_t$
Date

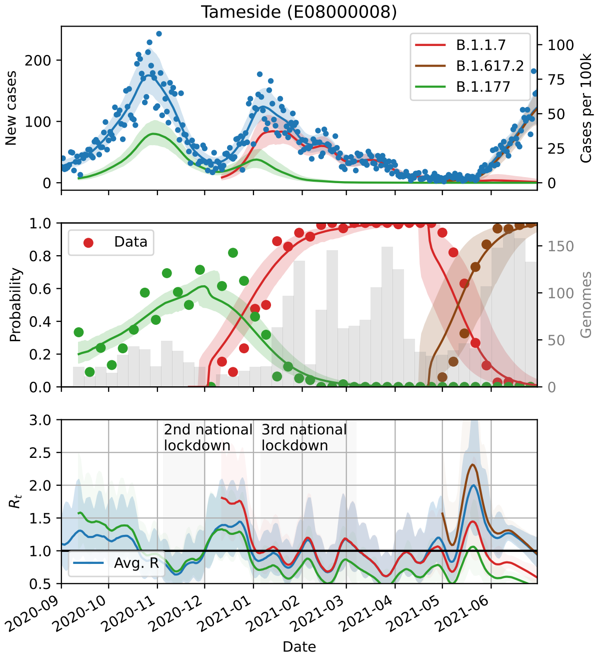
Tameside (E08000008)
B.1.1.7
B.1.617.2
B.1.177
New cases
Cases per 100k
Data
Probability
Genomes
2nd national lockdown
3rd national lockdown
Avg. R
$R_t$
Date

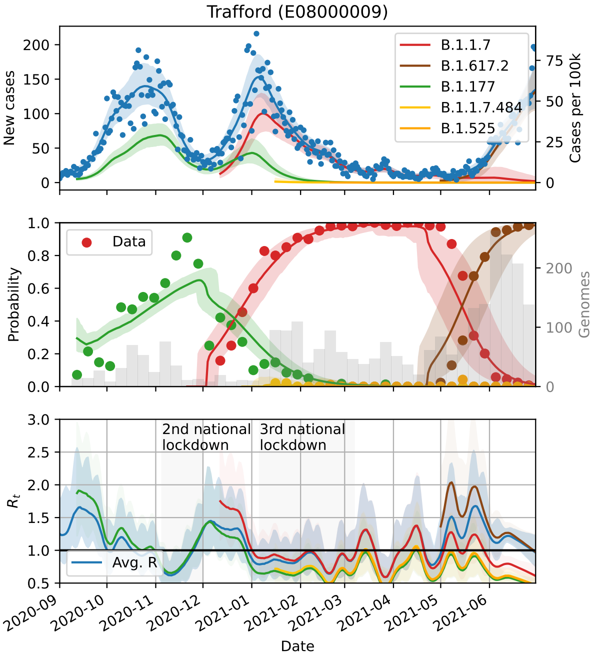
Trafford (E08000009)
New cases
Cases per 100k
B.1.1.7
B.1.617.2
B.1.177
B.1.1.7.484
B.1.525
Probability
Genomes
Data
$R_t$
2nd national lockdown
3rd national lockdown
Avg. R
Date

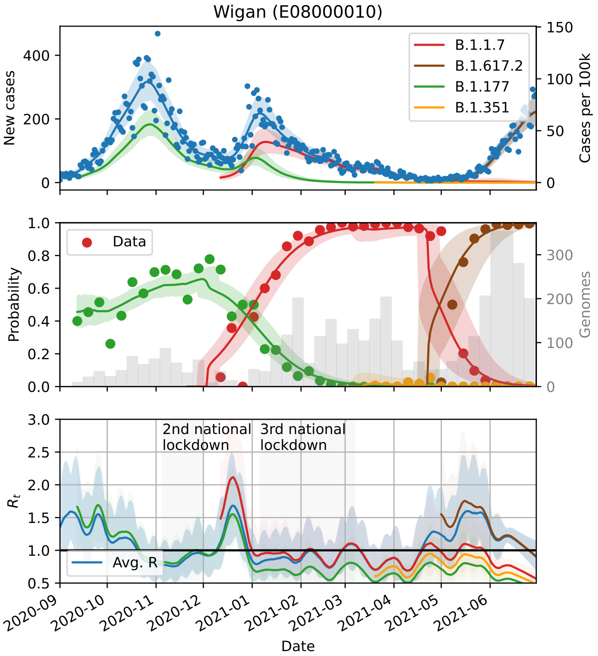
Wigan (E08000010)
New cases
Cases per 100k
B.1.1.7
B.1.617.2
B.1.177
B.1.351
Probability
Genomes
Data
$R_t$
2nd national lockdown
3rd national lockdown
Avg. R
Date

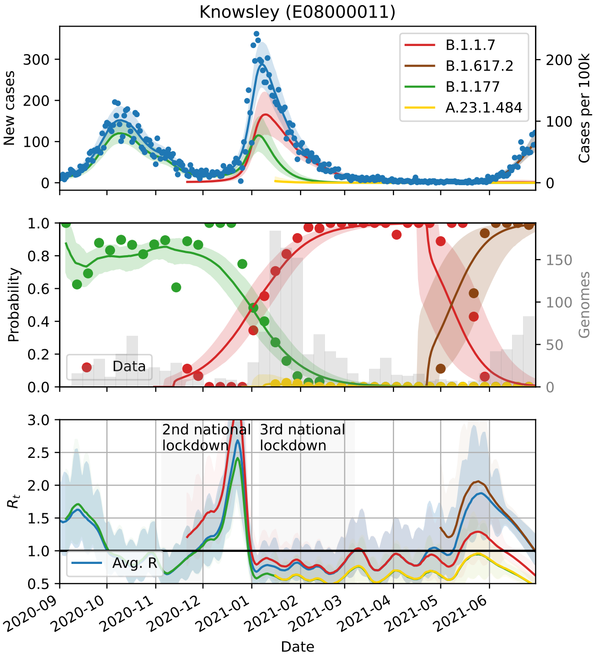
Knowsley (E08000011)
New cases
Cases per 100k
B.1.1.7
B.1.617.2
B.1.177
A.23.1.484
Probability
Genomes
Data
$R_t$
2nd national lockdown
3rd national lockdown
Avg. R
Date

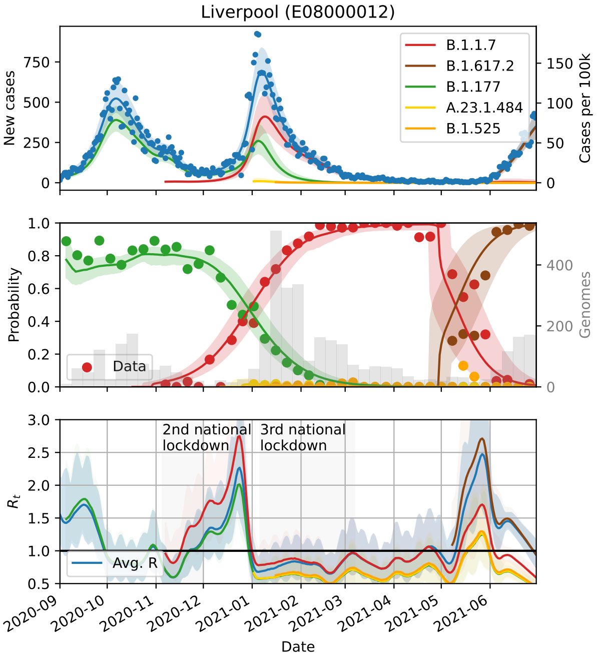
Liverpool (E08000012)
New cases
Cases per 100k
B.1.1.7
B.1.617.2
B.1.177
A.23.1.484
B.1.525
Probability
Genomes
Data
$R_t$
2nd national lockdown
3rd national lockdown
Avg. R
Date

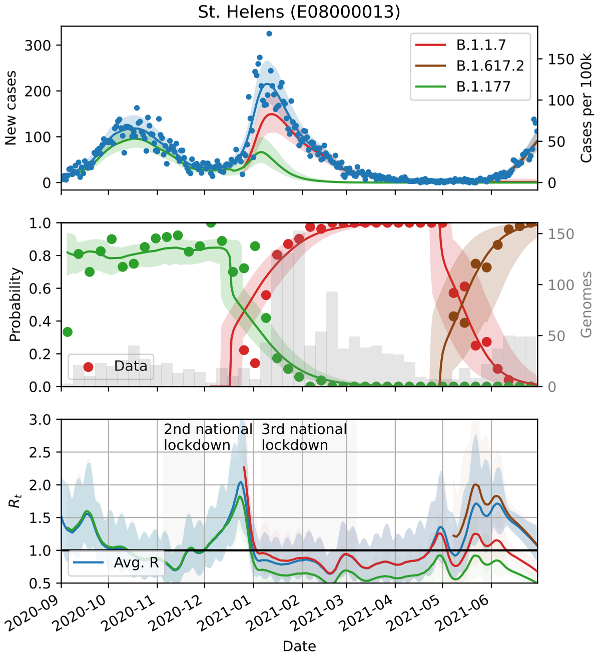
St. Helens (E08000013)
New cases
Cases per 100k
B.1.1.7
B.1.617.2
B.1.177
Probability
Genomes
Data
2nd national lockdown
3rd national lockdown
$R_t$
Avg. R
Date

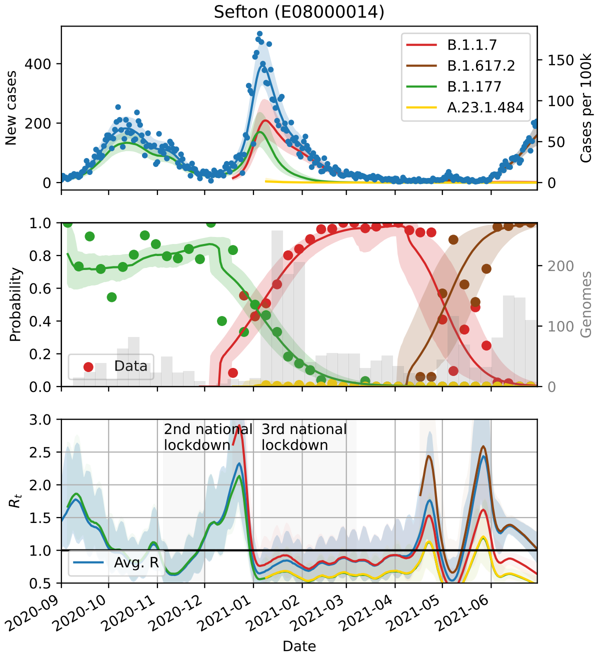
Sefton (E08000014)
New cases
Cases per 100k
B.1.1.7
B.1.617.2
B.1.177
A.23.1.484
Probability
Genomes
Data
2nd national lockdown
3rd national lockdown
$R_t$
Avg. R
Date

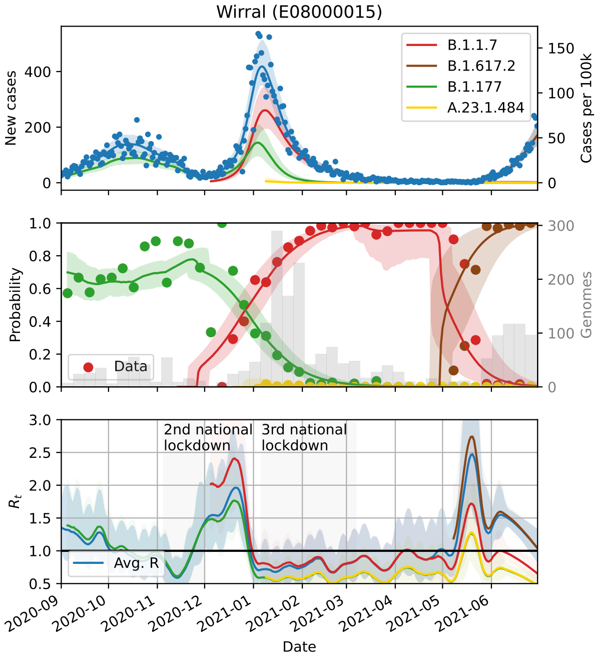
Wirral (E08000015)
New cases
Cases per 100k
B.1.1.7
B.1.617.2
B.1.177
A.23.1.484
Probability
Genomes
Data
2nd national lockdown
3rd national lockdown
$R_t$
Avg. R
Date

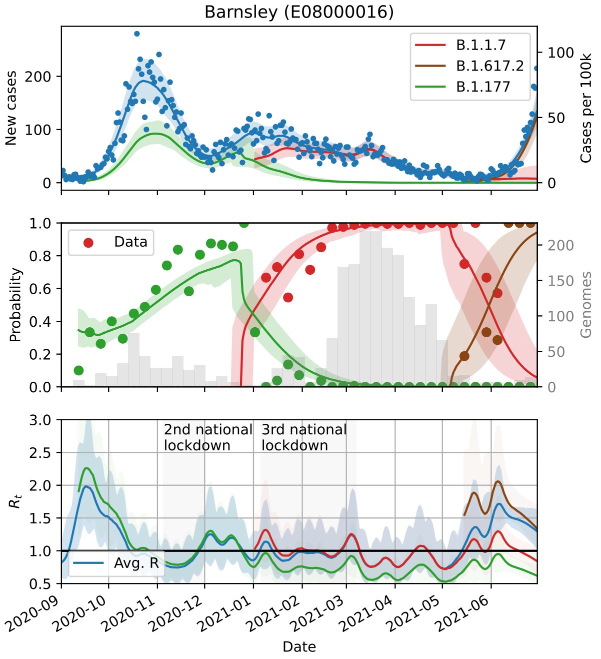
Barnsley (E08000016)
New cases
Cases per 100k
B.1.1.7
B.1.617.2
B.1.177
Probability
Genomes
Data
2nd national lockdown
3rd national lockdown
$R_t$
Avg. R
Date

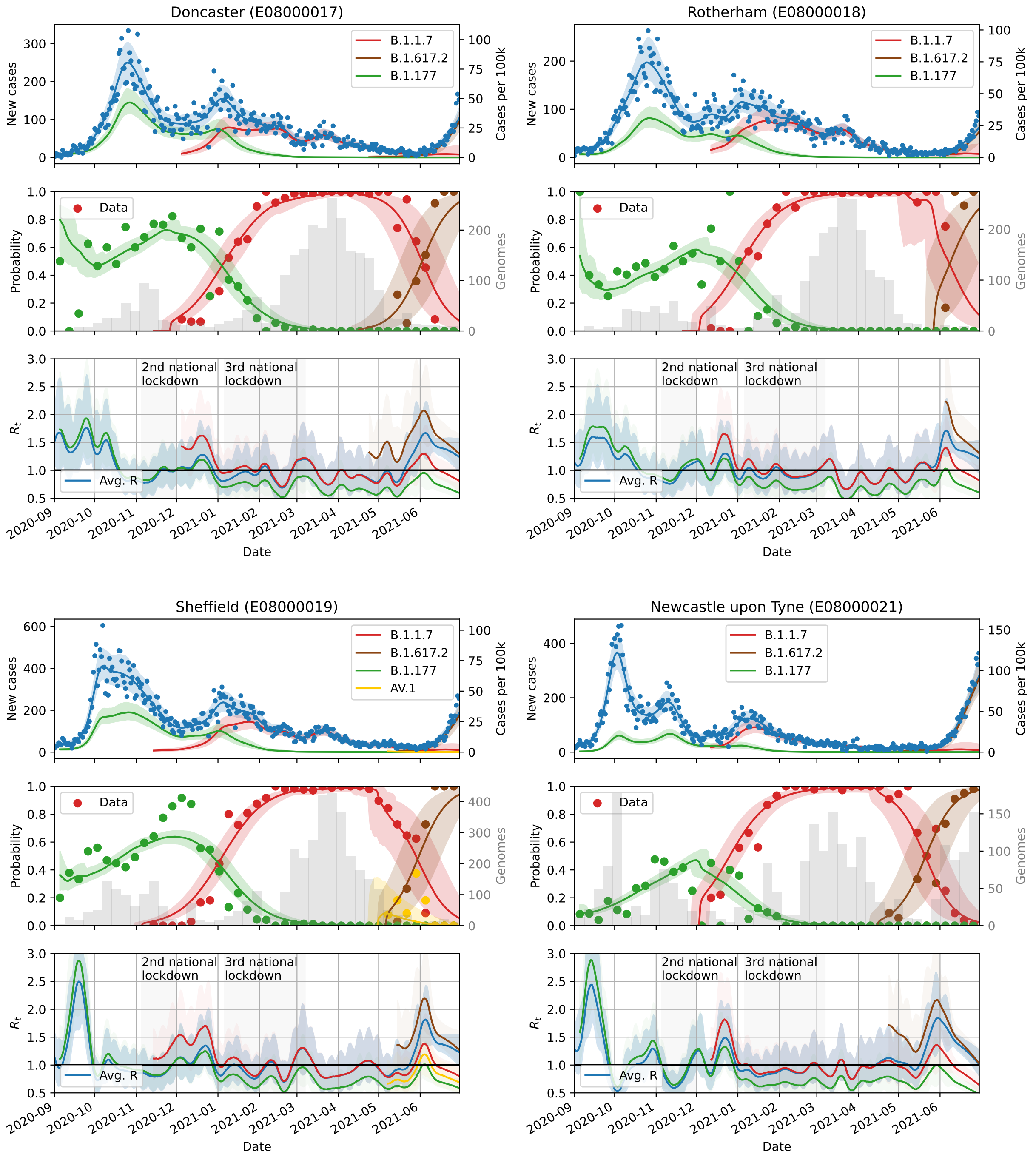
Doncaster (E08000017)
Rotherham (E08000018)
Sheffield (E08000019)
Newcastle upon Tyne (E08000021)
New cases
Cases per 100k
B.1.1.7
B.1.617.2
B.1.177
AV.1
Data
Probability
Genomes
2nd national lockdown
3rd national lockdown
Avg. R
$R_t$
Date
2020-09
2020-10
2020-11
2020-12
2021-01
2021-02
2021-03
2021-04
2021-05
2021-06

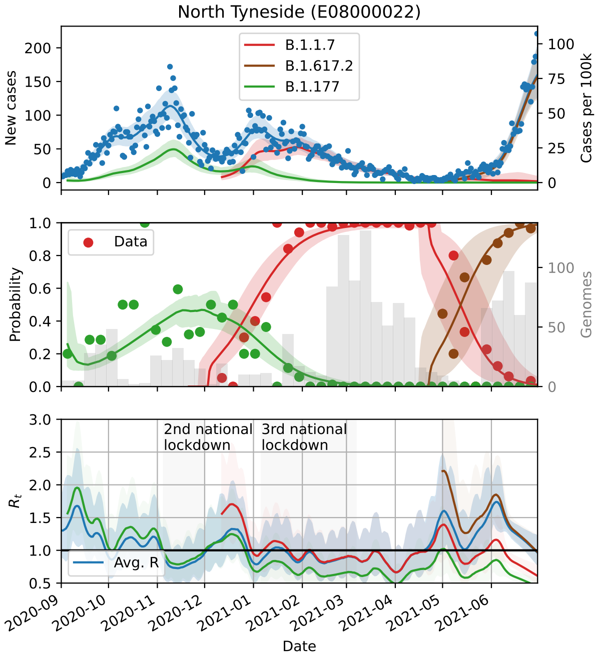
North Tyneside (E08000022)
New cases
Cases per 100k
B.1.1.7
B.1.617.2
B.1.177
Data
Probability
Genomes
2nd national lockdown
3rd national lockdown
$R_t$
Avg. R
Date

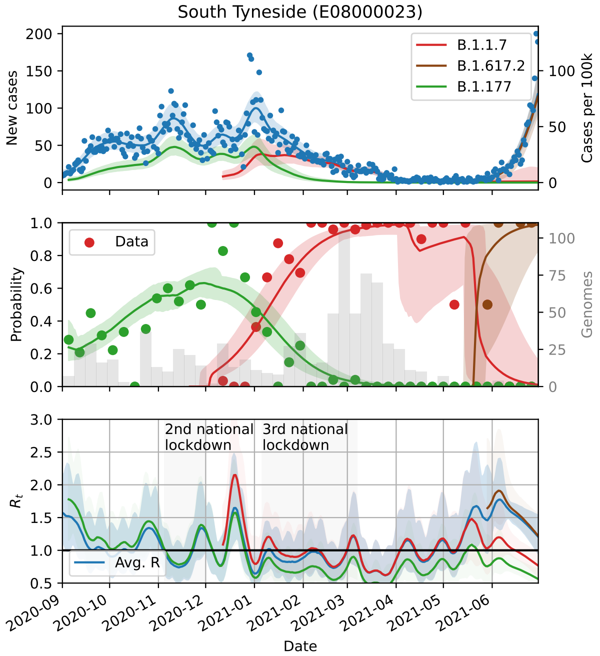
South Tyneside (E08000023)
New cases
Cases per 100k
B.1.1.7
B.1.617.2
B.1.177
Data
Probability
Genomes
2nd national lockdown
3rd national lockdown
$R_t$
Avg. R
Date

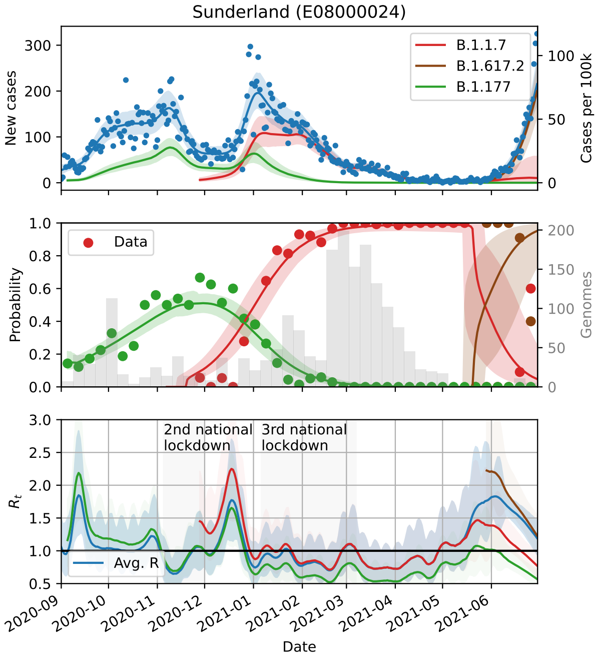
Sunderland (E08000024)
New cases
Cases per 100k
B.1.1.7
B.1.617.2
B.1.177
Data
Probability
Genomes
2nd national lockdown
3rd national lockdown
$R_t$
Avg. R
Date

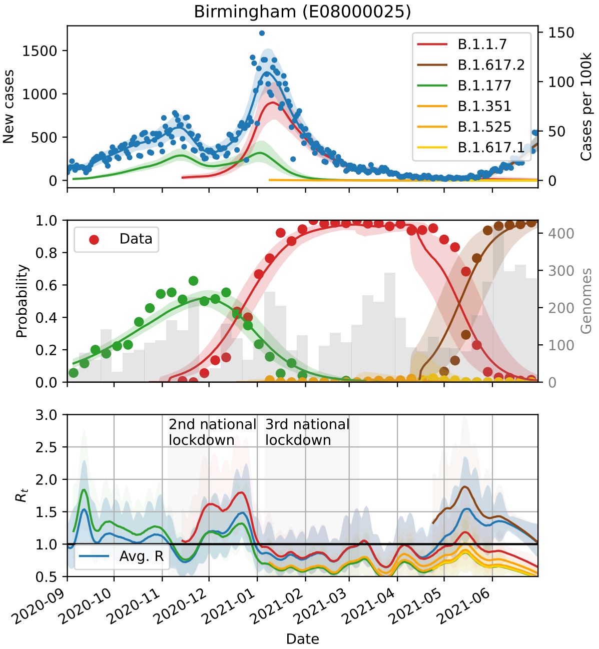
Birmingham (E08000025)
New cases
Cases per 100k
B.1.1.7
B.1.617.2
B.1.177
B.1.351
B.1.525
B.1.617.1
Data
Probability
Genomes
2nd national lockdown
3rd national lockdown
$R_t$
Avg. R
Date

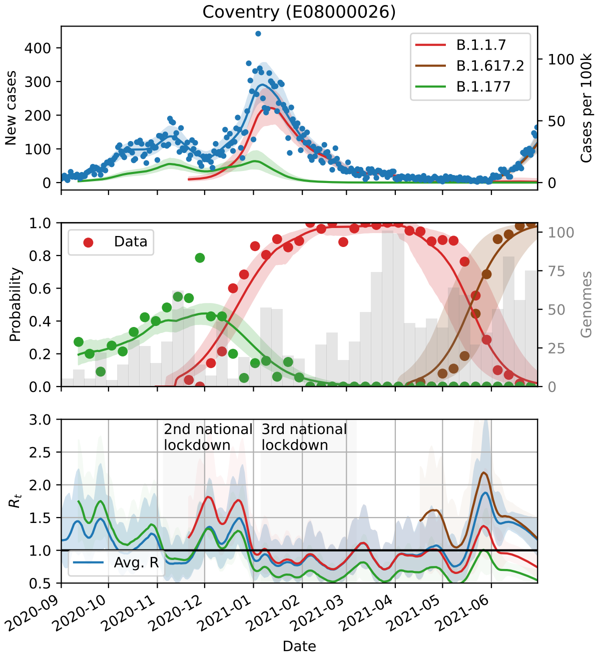
Coventry (E08000026)
B.1.1.7
B.1.617.2
B.1.177
New cases
Cases per 100k
Data
Probability
Genomes
2nd national lockdown
3rd national lockdown
Avg. R
$R_t$
Date

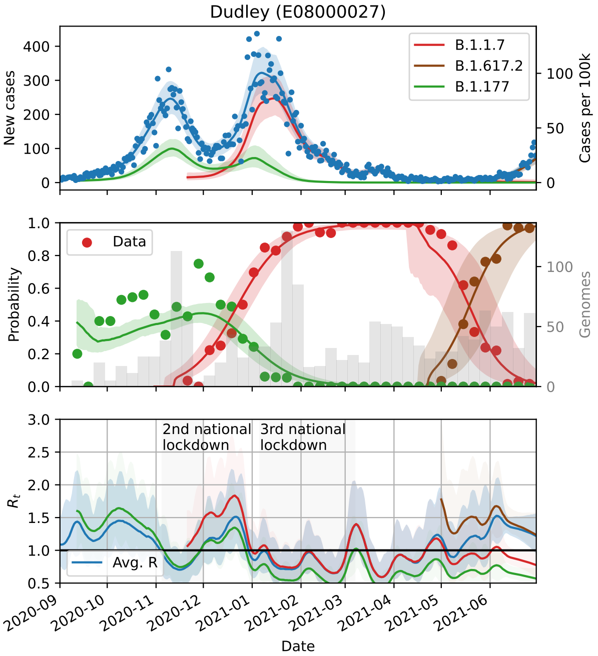
Dudley (E08000027)
B.1.1.7
B.1.617.2
B.1.177
New cases
Cases per 100k
Data
Probability
Genomes
2nd national lockdown
3rd national lockdown
Avg. R
$R_t$
Date

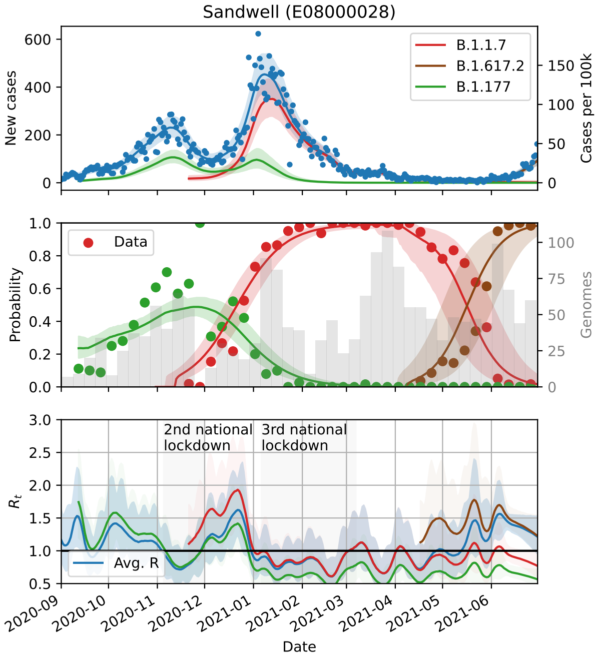
Sandwell (E08000028)
B.1.1.7
B.1.617.2
B.1.177
New cases
Cases per 100k
Data
Probability
Genomes
2nd national lockdown
3rd national lockdown
Avg. R
$R_t$
Date

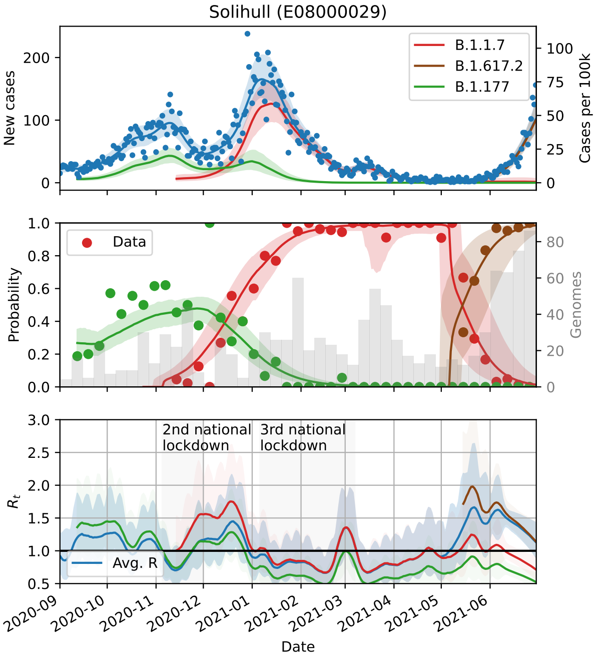
Solihull (E08000029)
B.1.1.7
B.1.617.2
B.1.177
New cases
Cases per 100k
Data
Probability
Genomes
2nd national lockdown
3rd national lockdown
Avg. R
$R_t$
Date

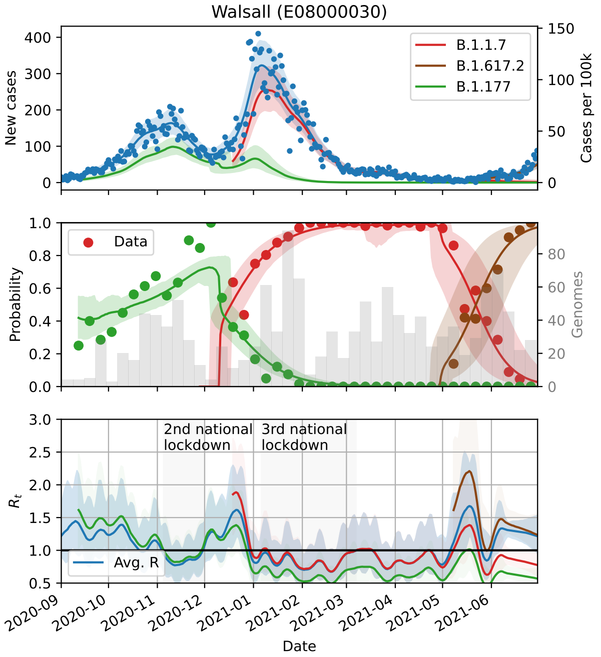
Walsall (E08000030)
B.1.1.7
B.1.617.2
B.1.177
New cases
Cases per 100k
Data
Probability
Genomes
2nd national lockdown
3rd national lockdown
Avg. R
$R_t$
Date

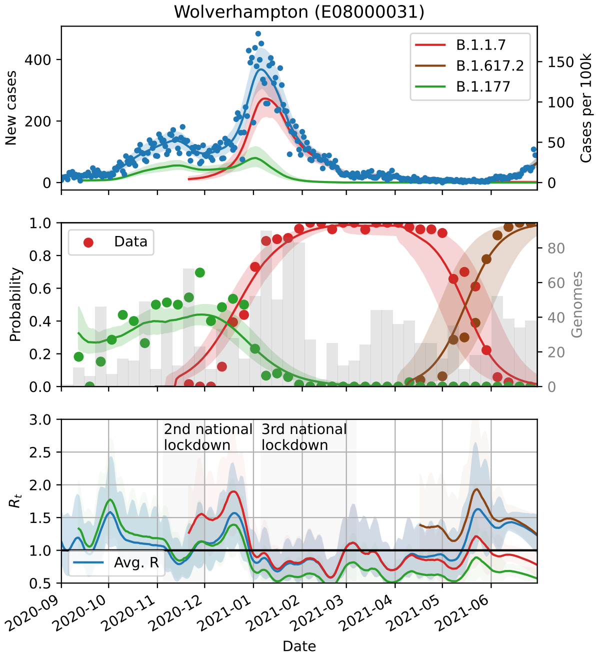
Wolverhampton (E08000031)
B.1.1.7
B.1.617.2
B.1.177
New cases
Cases per 100k
Data
Probability
Genomes
2nd national lockdown
3rd national lockdown
Avg. R
$R_t$
Date

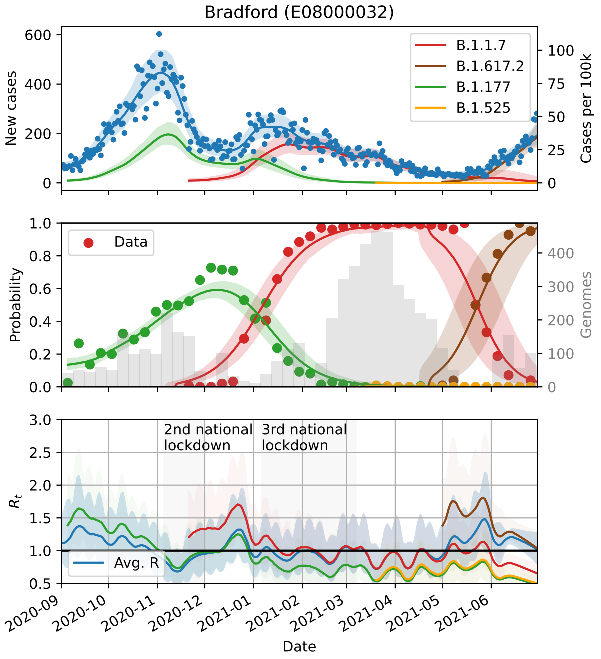
Bradford (E08000032)
B.1.1.7
B.1.617.2
B.1.177
B.1.525
New cases
Cases per 100k
Data
Probability
Genomes
2nd national lockdown
3rd national lockdown
Avg. R
$R_t$
Date

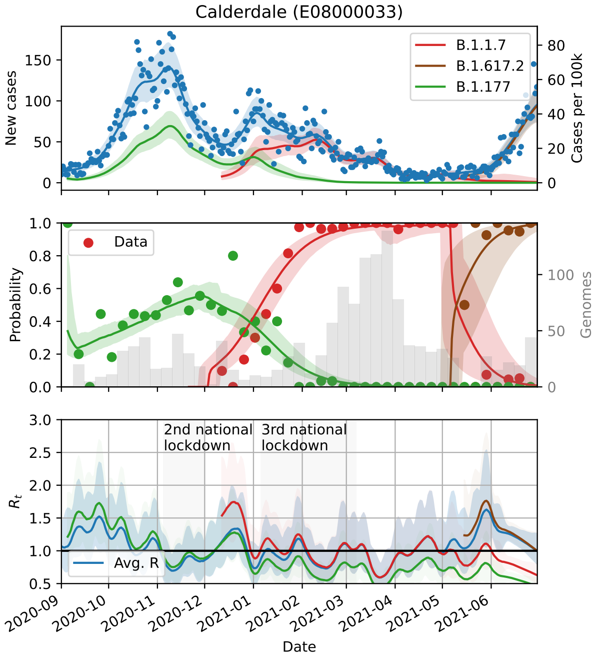
Calderdale (E08000033)
B.1.1.7
B.1.617.2
B.1.177
New cases
Cases per 100k
Data
Probability
Genomes
2nd national lockdown
3rd national lockdown
Avg. R
$R_t$
Date

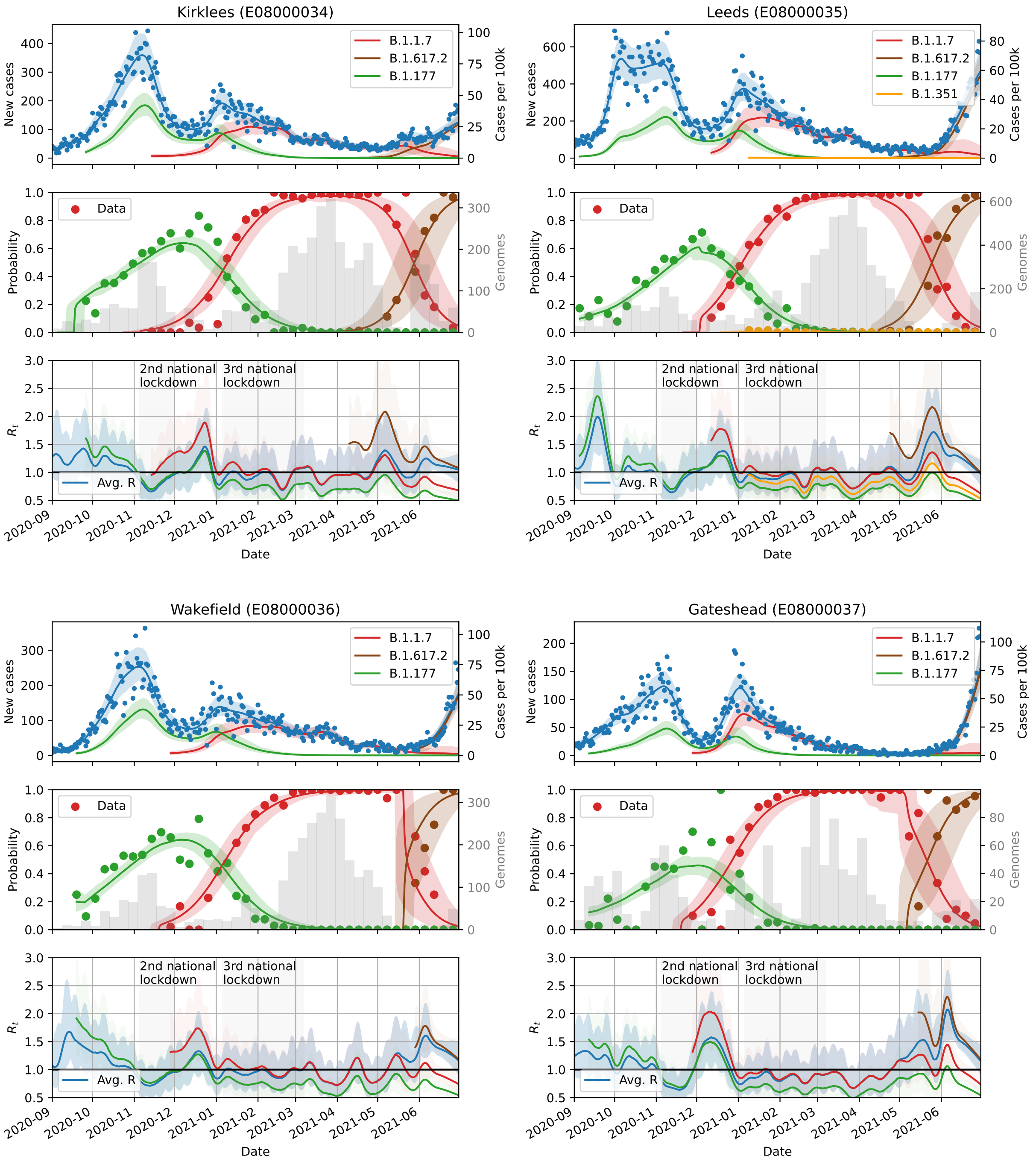
Kirklees (E08000034)
Leeds (E08000035)
Wakefield (E08000036)
Gateshead (E08000037)
New cases
Cases per 100k
B.1.1.7
B.1.617.2
B.1.177
B.1.351
Data
Probability
Genomes
$R_t$
Avg. R
2nd national lockdown
3rd national lockdown
2020-09
2020-10
2020-11
2020-12
2021-01
2021-02
2021-03
2021-04
2021-05
2021-06
Date

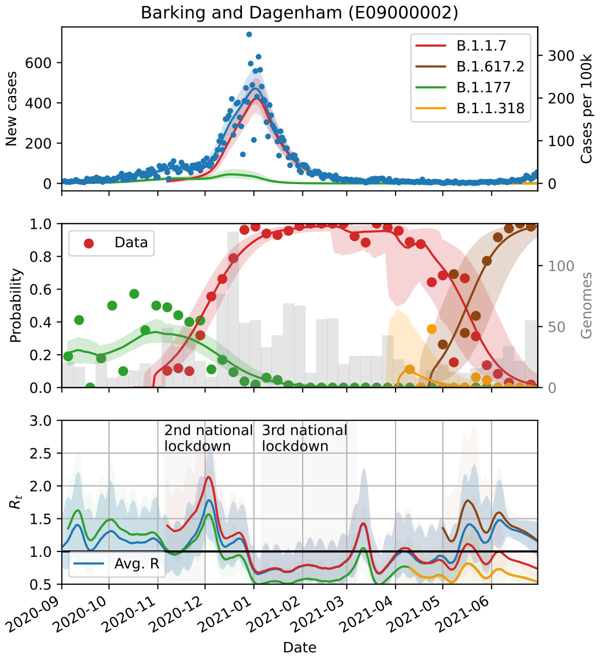
Barking and Dagenham (E09000002)
B.1.1.7
B.1.617.2
B.1.177
B.1.1.318
New cases
Cases per 100k
Data
Probability
Genomes
2nd national lockdown
3rd national lockdown
$R_t$
Avg. R
Date

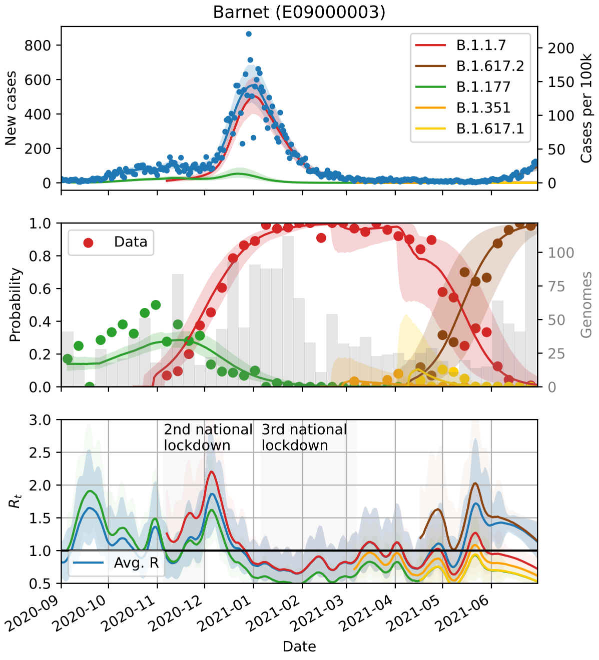
Barnet (E09000003)
B.1.1.7
B.1.617.2
B.1.177
B.1.351
B.1.617.1
New cases
Cases per 100k
Data
Probability
Genomes
2nd national lockdown
3rd national lockdown
$R_t$
Avg. R
Date

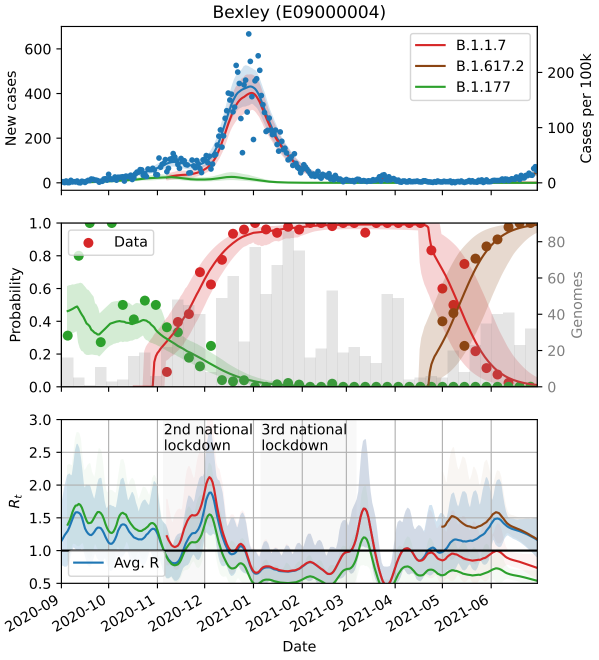
Bexley (E09000004)
B.1.1.7
B.1.617.2
B.1.177
New cases
Cases per 100k
Data
Probability
Genomes
2nd national lockdown
3rd national lockdown
$R_t$
Avg. R
Date

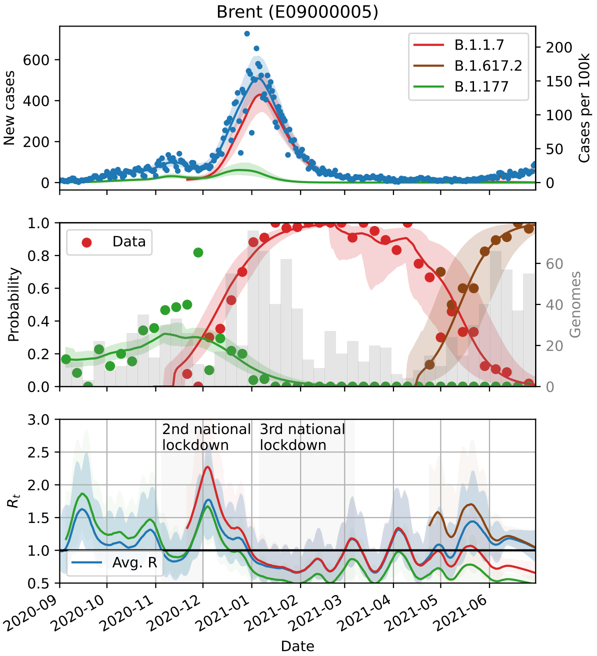
Brent (E09000005)
New cases
Cases per 100k
B.1.1.7
B.1.617.2
B.1.177
Data
Probability
Genomes
2nd national lockdown
3rd national lockdown
$R_t$
Avg. R
Date

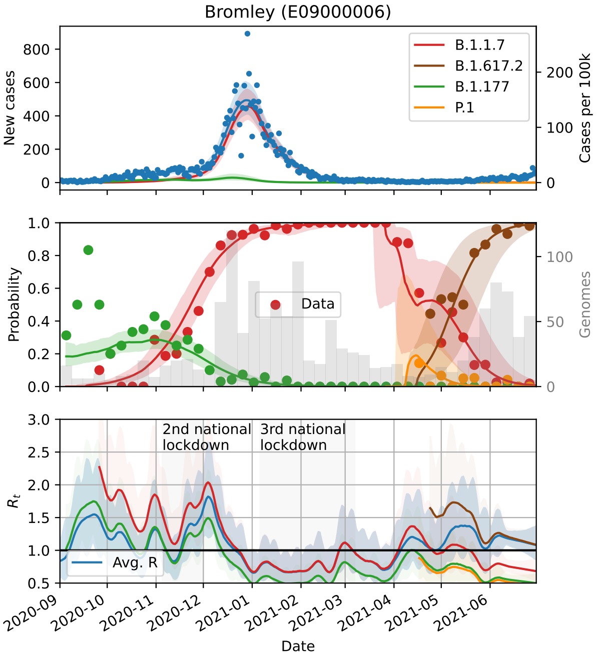
Bromley (E09000006)
New cases
Cases per 100k
B.1.1.7
B.1.617.2
B.1.177
P.1
Data
Probability
Genomes
2nd national lockdown
3rd national lockdown
$R_t$
Avg. R
Date

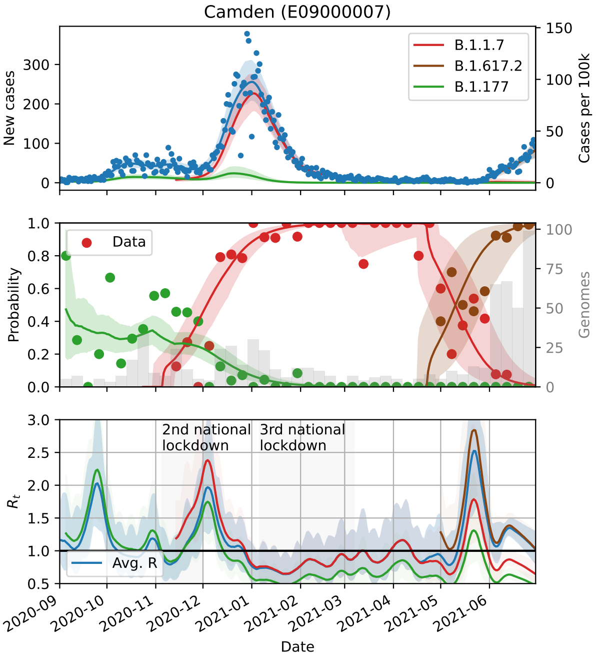
Camden (E09000007)
New cases
Cases per 100k
B.1.1.7
B.1.617.2
B.1.177
Data
Probability
Genomes
2nd national lockdown
3rd national lockdown
$R_t$
Avg. R
Date

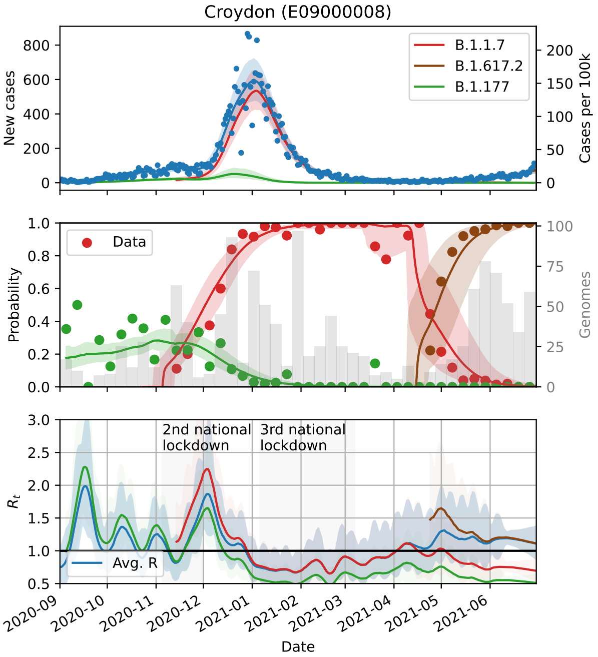
Croydon (E09000008)
New cases
Cases per 100k
B.1.1.7
B.1.617.2
B.1.177
Data
Probability
Genomes
2nd national lockdown
3rd national lockdown
$R_t$
Avg. R
Date

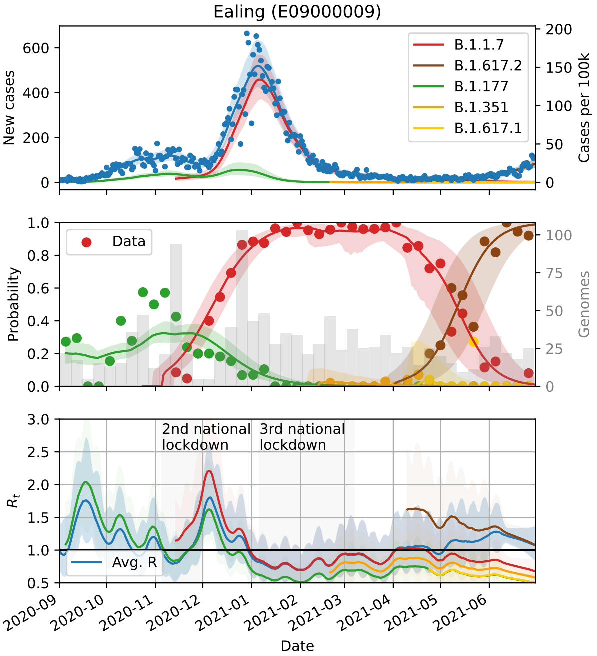
Ealing (E09000009)
B.1.1.7
B.1.617.2
B.1.177
B.1.351
B.1.617.1
New cases
Cases per 100k
Data
Probability
Genomes
2nd national lockdown
3rd national lockdown
$R_t$
Avg. R
Date

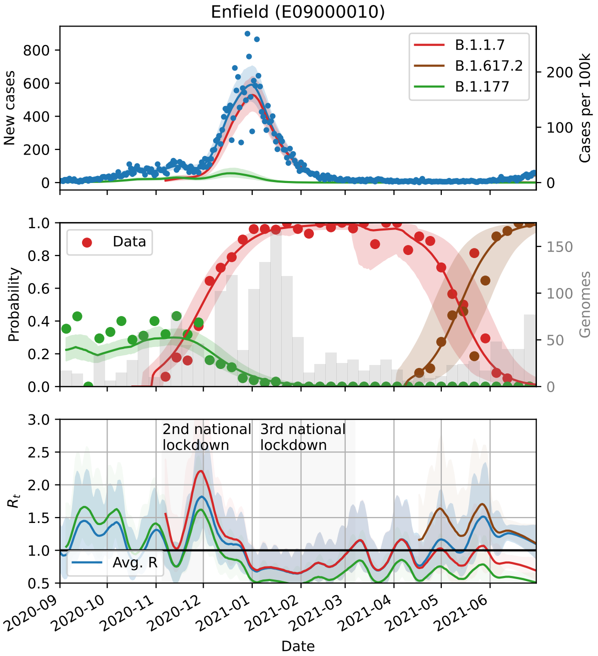
Enfield (E09000010)
B.1.1.7
B.1.617.2
B.1.177
New cases
Cases per 100k
Data
Probability
Genomes
2nd national lockdown
3rd national lockdown
$R_t$
Avg. R
Date

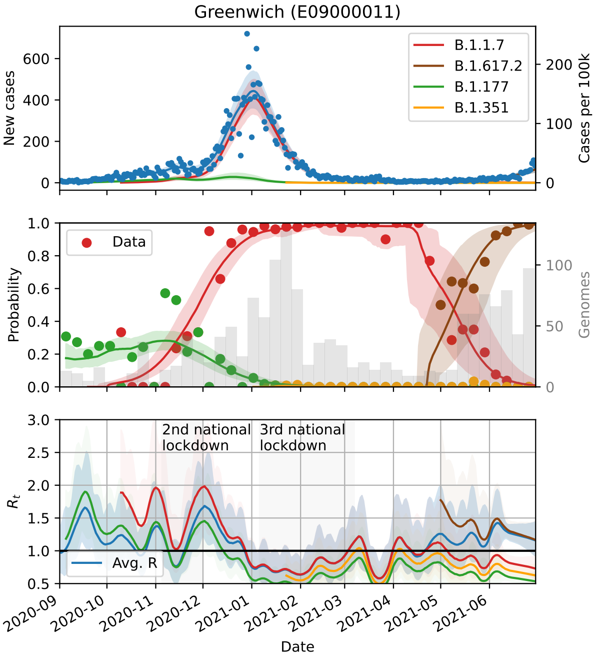
Greenwich (E09000011)
B.1.1.7
B.1.617.2
B.1.177
B.1.351
New cases
Cases per 100k
Data
Probability
Genomes
2nd national lockdown
3rd national lockdown
$R_t$
Avg. R
Date

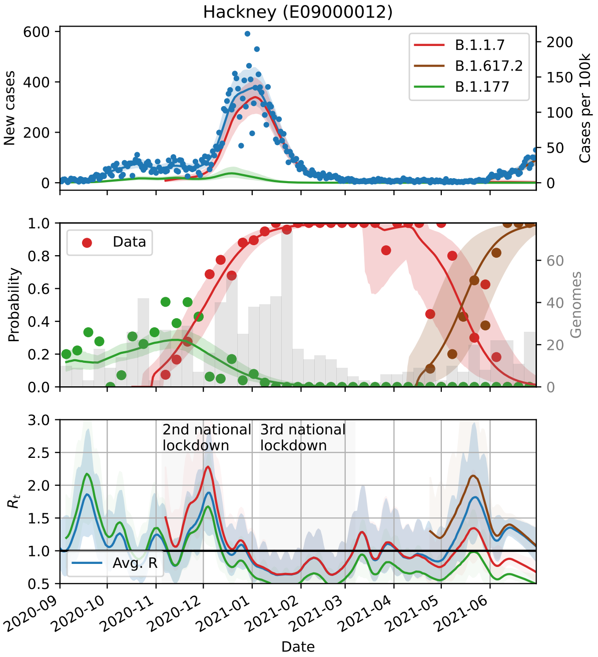
Hackney (E09000012)
B.1.1.7
B.1.617.2
B.1.177
New cases
Cases per 100k
Data
Probability
Genomes
2nd national lockdown
3rd national lockdown
$R_t$
Avg. R
Date

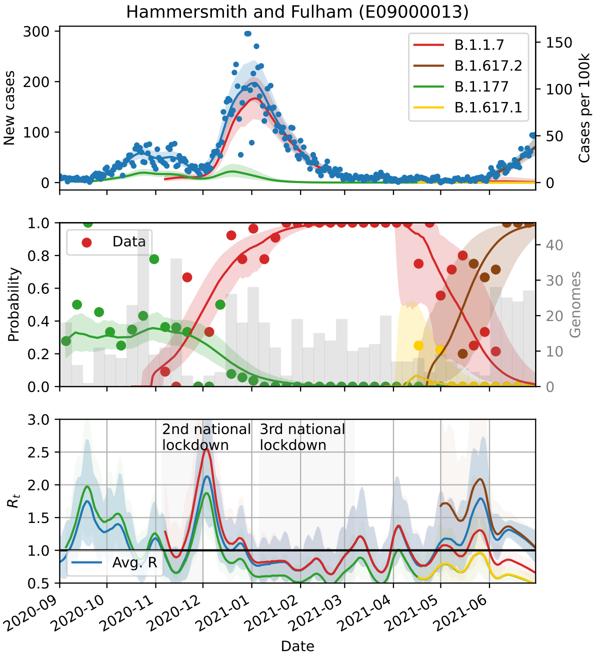
Hammersmith and Fulham (E09000013)
New cases
Cases per 100k
B.1.1.7
B.1.617.2
B.1.177
B.1.617.1
Data
Probability
Genomes
2nd national lockdown
3rd national lockdown
$R_t$
Avg. R
Date

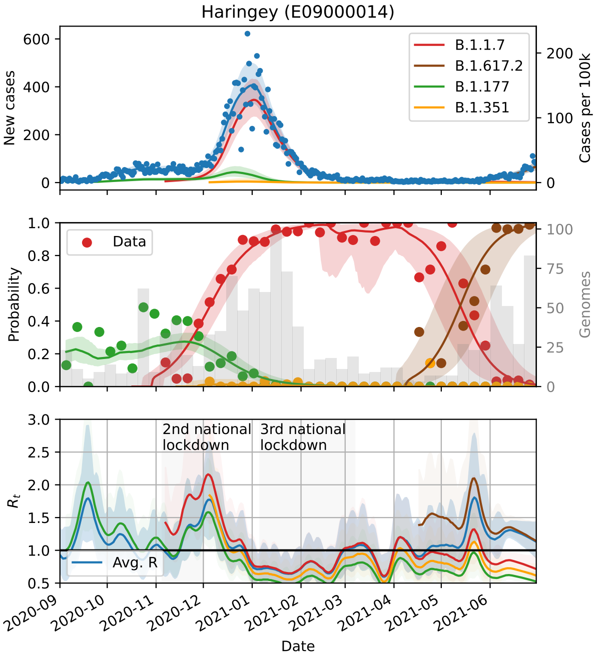
Haringey (E09000014)
New cases
Cases per 100k
B.1.1.7
B.1.617.2
B.1.177
B.1.351
Data
Probability
Genomes
2nd national lockdown
3rd national lockdown
$R_t$
Avg. R
Date

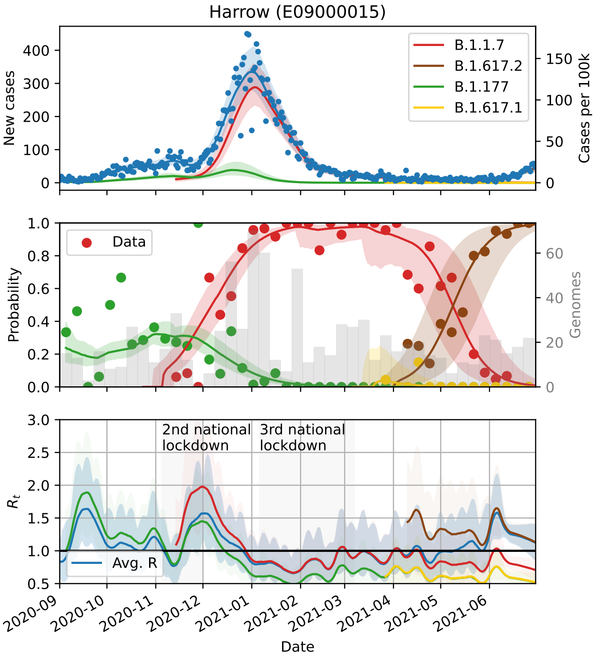
Harrow (E09000015)
New cases
Cases per 100k
B.1.1.7
B.1.617.2
B.1.177
B.1.617.1
Data
Probability
Genomes
2nd national lockdown
3rd national lockdown
$R_t$
Avg. R
Date

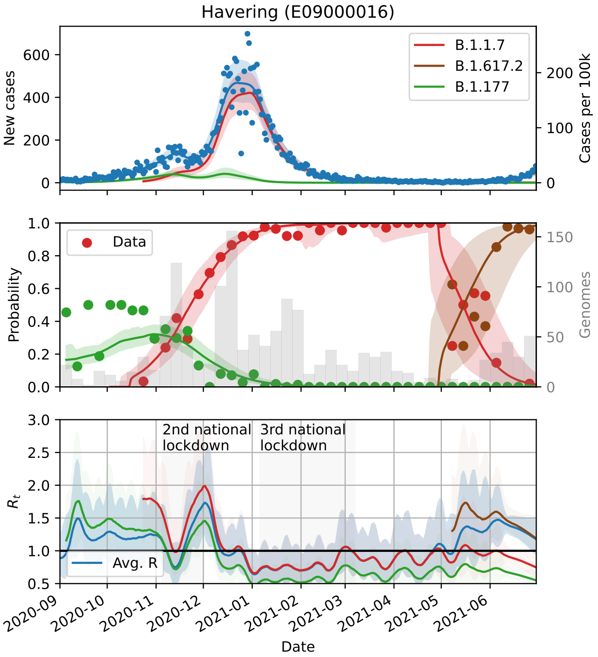
Havering (E09000016)
New cases
Cases per 100k
B.1.1.7
B.1.617.2
B.1.177
Data
Probability
Genomes
2nd national lockdown
3rd national lockdown
$R_t$
Avg. R
Date

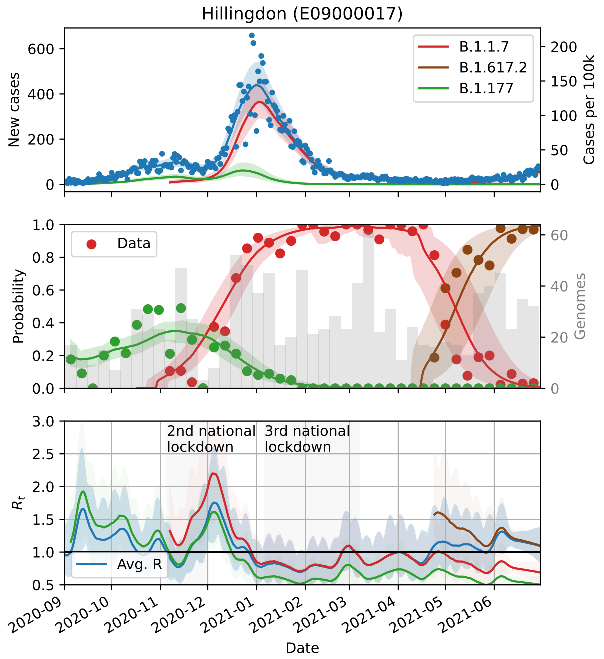
Hillingdon (E09000017)
New cases
Cases per 100k
B.1.1.7
B.1.617.2
B.1.177
Data
Probability
Genomes
2nd national lockdown
3rd national lockdown
Avg. R
$R_t$
Date

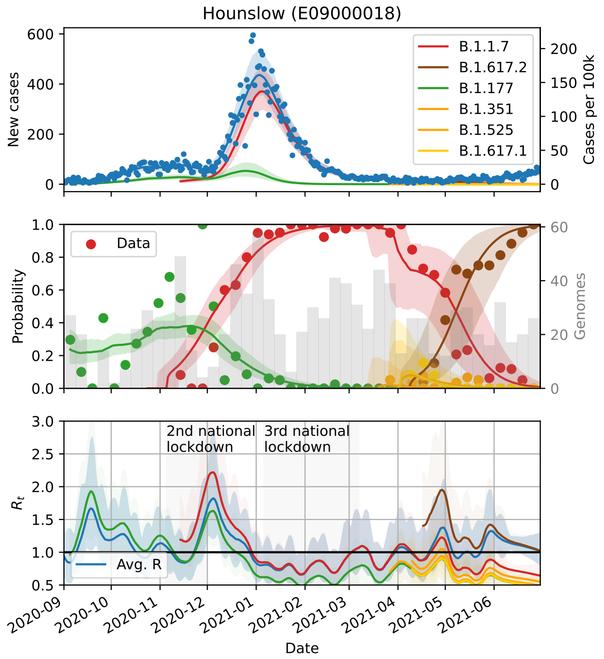
Hounslow (E09000018)
New cases
Cases per 100k
B.1.1.7
B.1.617.2
B.1.177
B.1.351
B.1.525
B.1.617.1
Data
Probability
Genomes
2nd national lockdown
3rd national lockdown
Avg. R
$R_t$
Date

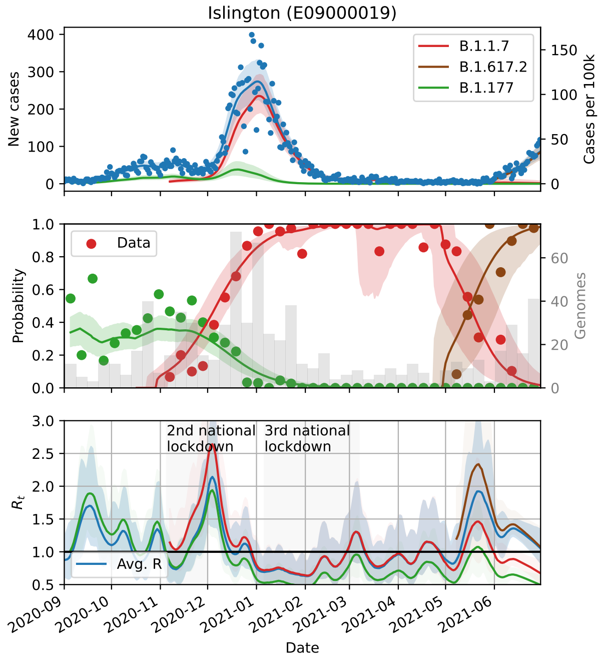
Islington (E09000019)
New cases
Cases per 100k
B.1.1.7
B.1.617.2
B.1.177
Data
Probability
Genomes
2nd national lockdown
3rd national lockdown
Avg. R
$R_t$
Date

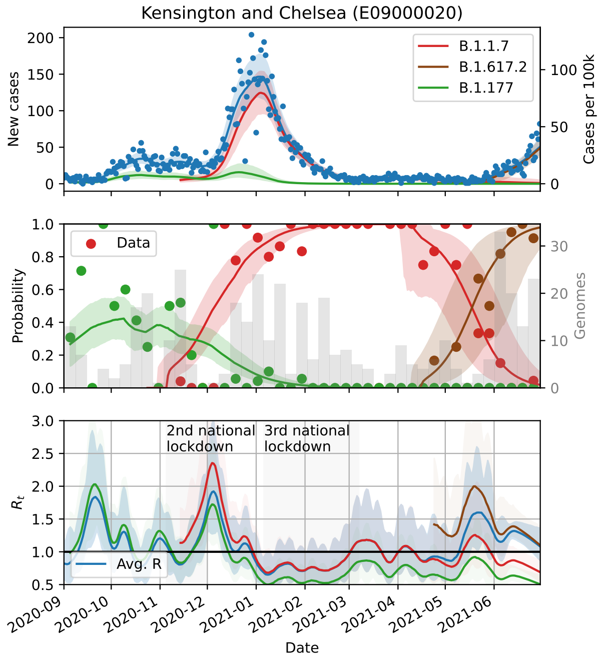
Kensington and Chelsea (E09000020)
New cases
Cases per 100k
B.1.1.7
B.1.617.2
B.1.177
Data
Probability
Genomes
2nd national lockdown
3rd national lockdown
Avg. R
$R_t$
Date

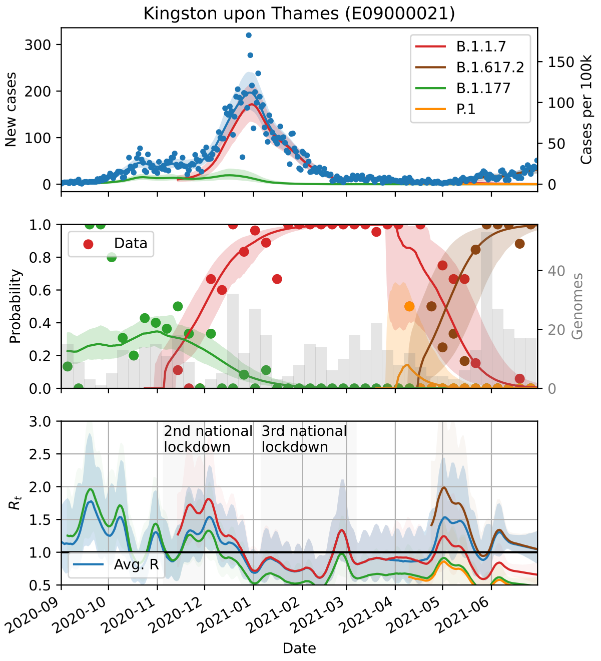
Kingston upon Thames (E09000021)
B.1.1.7
B.1.617.2
B.1.177
P.1
New cases
Cases per 100k
Data
Probability
Genomes
2nd national lockdown
3rd national lockdown
Avg. R
$R_t$
Date

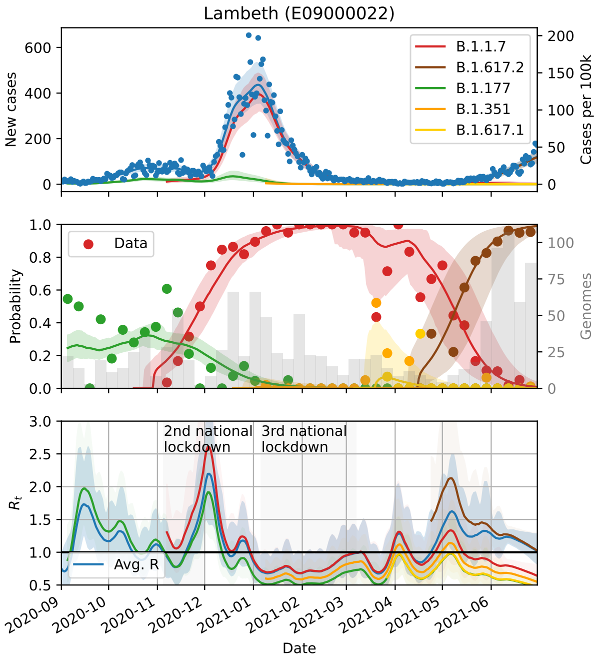
Lambeth (E09000022)
B.1.1.7
B.1.617.2
B.1.177
B.1.351
B.1.617.1
New cases
Cases per 100k
Data
Probability
Genomes
2nd national lockdown
3rd national lockdown
Avg. R
$R_t$
Date

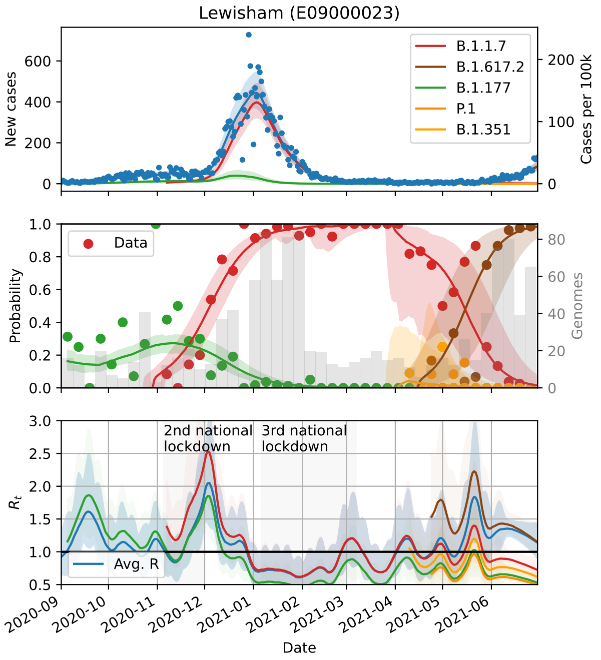
Lewisham (E09000023)
B.1.1.7
B.1.617.2
B.1.177
P.1
B.1.351
New cases
Cases per 100k
Data
Probability
Genomes
2nd national lockdown
3rd national lockdown
Avg. R
$R_t$
Date

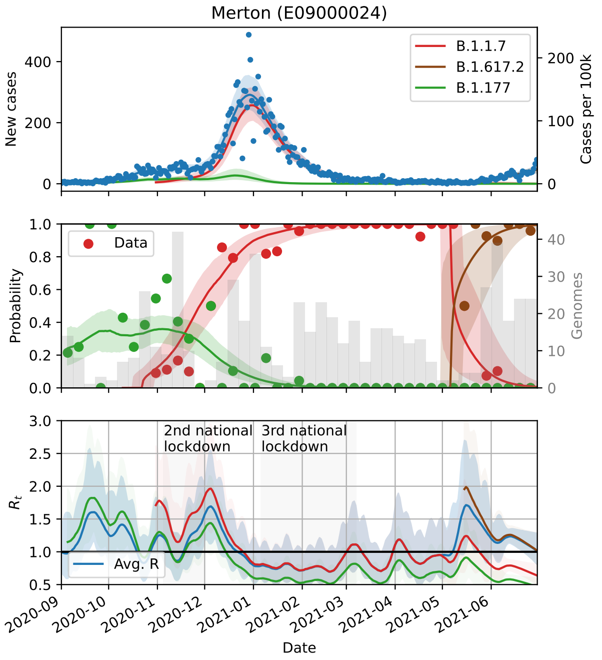
Merton (E09000024)
B.1.1.7
B.1.617.2
B.1.177
New cases
Cases per 100k
Data
Probability
Genomes
2nd national lockdown
3rd national lockdown
Avg. R
$R_t$
Date

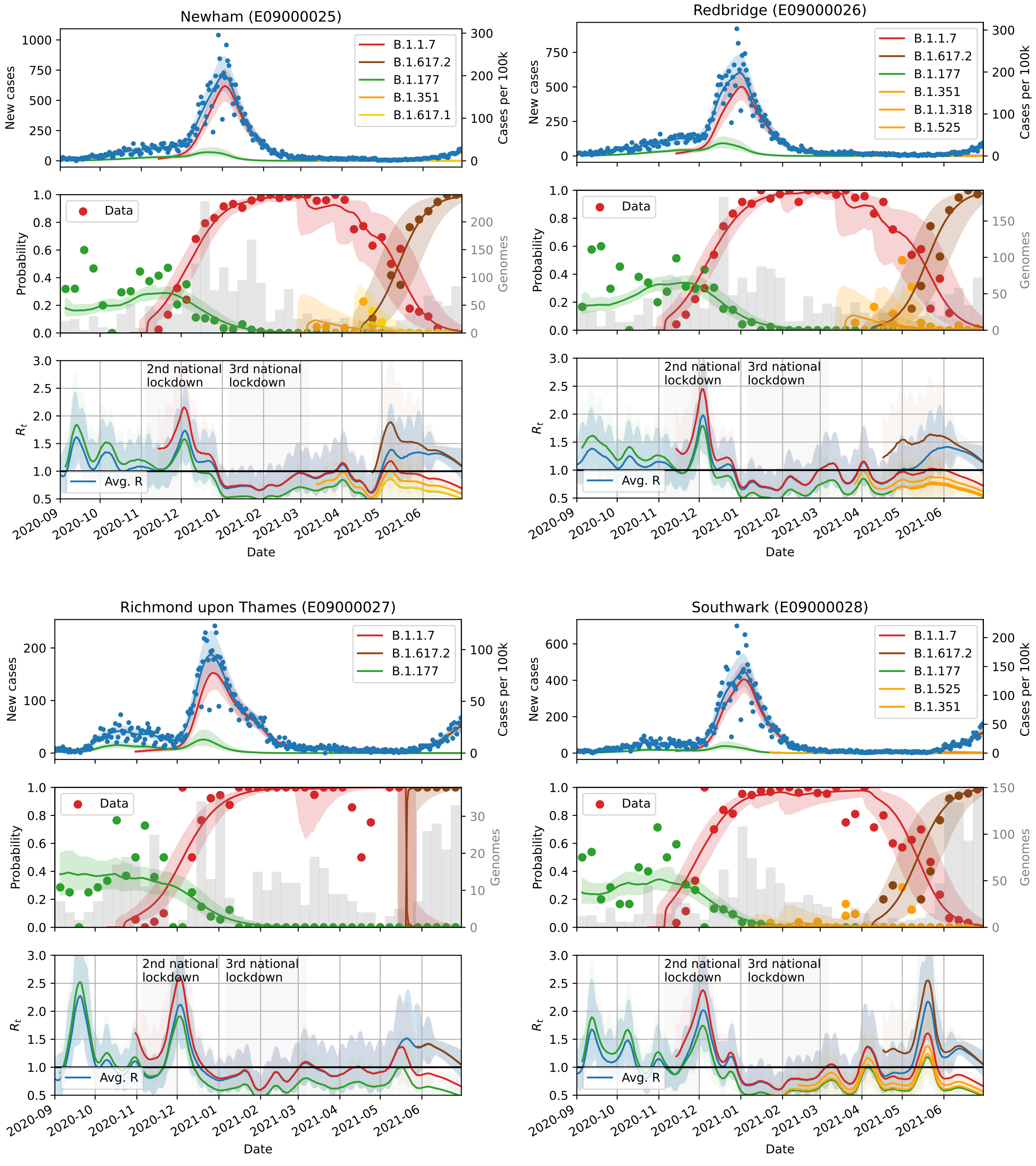
Newham (E09000025)
B.1.1.7
B.1.617.2
B.1.177
B.1.351
B.1.617.1
New cases
Cases per 100k
Data
Probability
Genomes
2nd national lockdown
3rd national lockdown
Avg. R
$R_t$
Date
Redbridge (E09000026)
B.1.1.7
B.1.617.2
B.1.177
B.1.351
B.1.1.318
B.1.525
Richmond upon Thames (E09000027)
B.1.1.7
B.1.617.2
B.1.177
Southwark (E09000028)
B.1.1.7
B.1.617.2
B.1.177
B.1.525
B.1.351

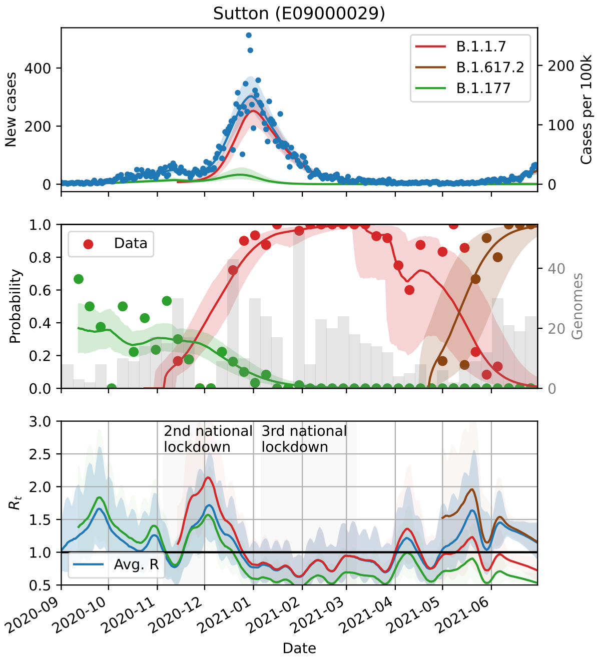
Sutton (E09000029)
B.1.1.7
B.1.617.2
B.1.177
New cases
Cases per 100k
Data
Probability
Genomes
2nd national lockdown
3rd national lockdown
Avg. R
$R_t$
Date

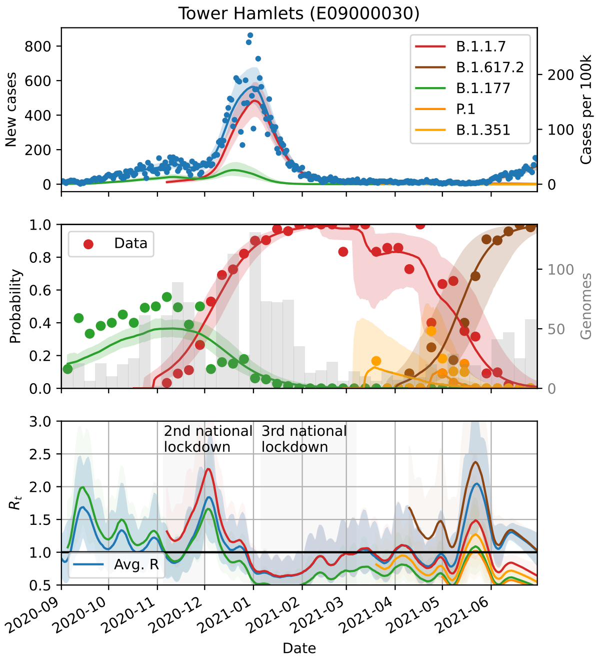
Tower Hamlets (E09000030)
B.1.1.7
B.1.617.2
B.1.177
P.1
B.1.351
New cases
Cases per 100k
Data
Probability
Genomes
2nd national lockdown
3rd national lockdown
Avg. R
$R_t$
Date

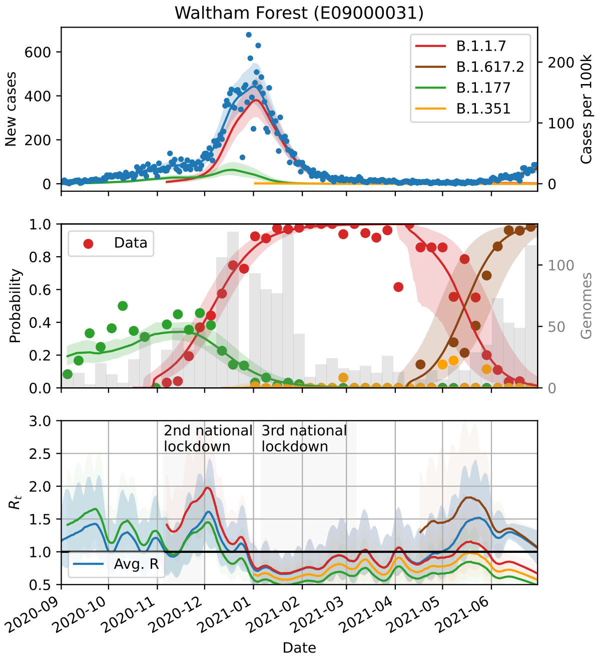
Waltham Forest (E09000031)
B.1.1.7
B.1.617.2
B.1.177
B.1.351
New cases
Cases per 100k
Data
Probability
Genomes
2nd national lockdown
3rd national lockdown
Avg. R
$R_t$
Date

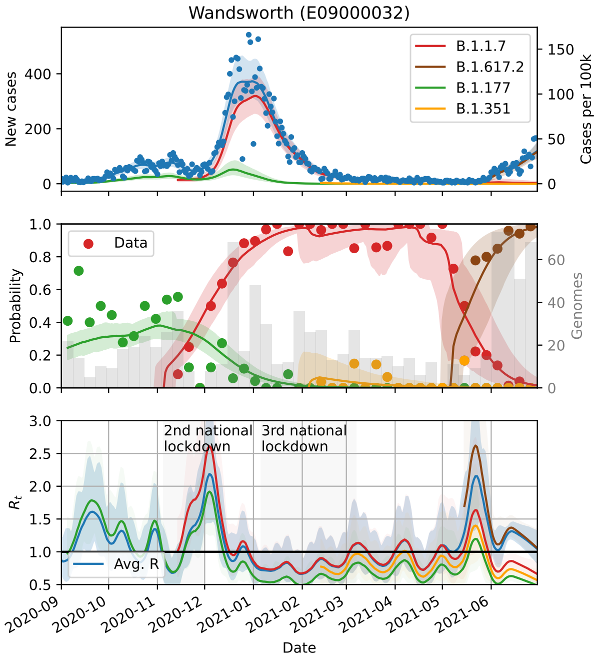
Wandsworth (E09000032)
B.1.1.7
B.1.617.2
B.1.177
B.1.351
New cases
Cases per 100k
Data
Probability
Genomes
2nd national lockdown
3rd national lockdown
Avg. R
$R_t$
Date

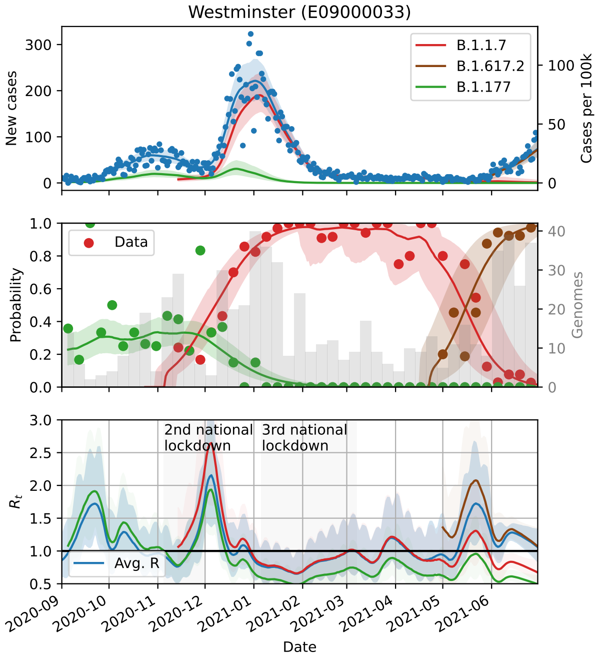

Westminster (E09000033)
New cases
Cases per 100k
B.1.1.7
B.1.617.2
B.1.177
Probability
Data
Genomes
2nd national lockdown
3rd national lockdown
$R_t$
Avg. R
Date

### 3 Supplementary Note 3

Supplementary Note 3 Tab. 1: **Lower Tier Local Authorities (LTLA) lookup codes and names.**

| | LTLA look up code | LTLA name |
|---|---|---|
| 0 | E06000001 | Hartlepool |
| 1 | E06000002 | Middlesbrough |
| 2 | E06000003 | Redcar and Cleveland |
| 3 | E06000004 | Stockton-on-Tees |
| 4 | E06000005 | Darlington |
| 5 | E06000006 | Halton |
| 6 | E06000007 | Warrington |
| 7 | E06000008 | Blackburn with Darwen |
| 8 | E06000009 | Blackpool |
| 9 | E06000010 | Kingston upon Hull, City of |
| 10 | E06000011 | East Riding of Yorkshire |
| 11 | E06000012 | North East Lincolnshire |
| 12 | E06000013 | North Lincolnshire |
| 13 | E06000014 | York |
| 14 | E06000015 | Derby |
| 15 | E06000016 | Leicester |
| 16 | E06000017 | Rutland |
| 17 | E06000018 | Nottingham |
| 18 | E06000019 | Herefordshire, County of |
| 19 | E06000020 | Telford and Wrekin |
| 20 | E06000021 | Stoke-on-Trent |
| 21 | E06000022 | Bath and North East Somerset |
| 22 | E06000023 | Bristol, City of |
| 23 | E06000024 | North Somerset |
| 24 | E06000025 | South Gloucestershire |
| 25 | E06000026 | Plymouth |
| 26 | E06000027 | Torbay |
| 27 | E06000030 | Swindon |
| 28 | E06000031 | Peterborough |
| 29 | E06000032 | Luton |
| 30 | E06000033 | Southend-on-Sea |

Continued on next page

| | LTLA look up code | LTLA name |
|---|---|---|
| 31 | E06000034 | Thurrock |
| 32 | E06000035 | Medway |
| 33 | E06000036 | Bracknell Forest |
| 34 | E06000037 | West Berkshire |
| 35 | E06000038 | Reading |
| 36 | E06000039 | Slough |
| 37 | E06000040 | Windsor and Maidenhead |
| 38 | E06000041 | Wokingham |
| 39 | E06000042 | Milton Keynes |
| 40 | E06000043 | Brighton and Hove |
| 41 | E06000044 | Portsmouth |
| 42 | E06000045 | Southampton |
| 43 | E06000046 | Isle of Wight |
| 44 | E06000047 | County Durham |
| 45 | E06000049 | Cheshire East |
| 46 | E06000050 | Cheshire West and Chester |
| 47 | E06000051 | Shropshire |
| 48 | E06000052 | Cornwall |
| 49 | E06000053 | Isles of Scilly |
| 50 | E06000054 | Wiltshire |
| 51 | E06000055 | Bedford |
| 52 | E06000056 | Central Bedfordshire |
| 53 | E06000057 | Northumberland |
| 54 | E06000058 | Bournemouth, Christchurch and Poole |
| 55 | E06000059 | Dorset |
| 56 | E07000004 | Aylesbury Vale |
| 57 | E07000005 | Chiltern |
| 58 | E07000006 | South Bucks |
| 59 | E07000007 | Wycombe |
| 60 | E07000008 | Cambridge |
| 61 | E07000009 | East Cambridgeshire |
| 62 | E07000010 | Fenland |
| 63 | E07000011 | Huntingdonshire |
| 64 | E07000012 | South Cambridgeshire |
| 65 | E07000026 | Allerdale |

Continued on next page

| | LTLA look up code | LTLA name |
|---|---|---|
| 66 | E07000027 | Barrow-in-Furness |
| 67 | E07000028 | Carlisle |
| 68 | E07000029 | Copeland |
| 69 | E07000030 | Eden |
| 70 | E07000031 | South Lakeland |
| 71 | E07000032 | Amber Valley |
| 72 | E07000033 | Bolsover |
| 73 | E07000034 | Chesterfield |
| 74 | E07000035 | Derbyshire Dales |
| 75 | E07000036 | Erewash |
| 76 | E07000037 | High Peak |
| 77 | E07000038 | North East Derbyshire |
| 78 | E07000039 | South Derbyshire |
| 79 | E07000040 | East Devon |
| 80 | E07000041 | Exeter |
| 81 | E07000042 | Mid Devon |
| 82 | E07000043 | North Devon |
| 83 | E07000044 | South Hams |
| 84 | E07000045 | Teignbridge |
| 85 | E07000046 | Torridge |
| 86 | E07000047 | West Devon |
| 87 | E07000061 | Eastbourne |
| 88 | E07000062 | Hastings |
| 89 | E07000063 | Lewes |
| 90 | E07000064 | Rother |
| 91 | E07000065 | Wealden |
| 92 | E07000066 | Basildon |
| 93 | E07000067 | Braintree |
| 94 | E07000068 | Brentwood |
| 95 | E07000069 | Castle Point |
| 96 | E07000070 | Chelmsford |
| 97 | E07000071 | Colchester |
| 98 | E07000072 | Epping Forest |
| 99 | E07000073 | Harlow |
| 100 | E07000074 | Maldon |

Continued on next page

| | LTLA look up code | LTLA name |
|---|---|---|
| 101 | E07000075 | Rochford |
| 102 | E07000076 | Tendring |
| 103 | E07000077 | Uttlesford |
| 104 | E07000078 | Cheltenham |
| 105 | E07000079 | Cotswold |
| 106 | E07000080 | Forest of Dean |
| 107 | E07000081 | Gloucester |
| 108 | E07000082 | Stroud |
| 109 | E07000083 | Tewkesbury |
| 110 | E07000084 | Basingstoke and Deane |
| 111 | E07000085 | East Hampshire |
| 112 | E07000086 | Eastleigh |
| 113 | E07000087 | Fareham |
| 114 | E07000088 | Gosport |
| 115 | E07000089 | Hart |
| 116 | E07000090 | Havant |
| 117 | E07000091 | New Forest |
| 118 | E07000092 | Rushmoor |
| 119 | E07000093 | Test Valley |
| 120 | E07000094 | Winchester |
| 121 | E07000095 | Broxbourne |
| 122 | E07000096 | Dacorum |
| 123 | E07000098 | Hertsmere |
| 124 | E07000099 | North Hertfordshire |
| 125 | E07000102 | Three Rivers |
| 126 | E07000103 | Watford |
| 127 | E07000105 | Ashford |
| 128 | E07000106 | Canterbury |
| 129 | E07000107 | Dartford |
| 130 | E07000108 | Dover |
| 131 | E07000109 | Gravesham |
| 132 | E07000110 | Maidstone |
| 133 | E07000111 | Sevenoaks |
| 134 | E07000112 | Folkestone and Hythe |
| 135 | E07000113 | Swale |

Continued on next page

| | LTLA look up code | LTLA name |
|---|---|---|
| 136 | E07000114 | Thanet |
| 137 | E07000115 | Tonbridge and Malling |
| 138 | E07000116 | Tunbridge Wells |
| 139 | E07000117 | Burnley |
| 140 | E07000118 | Chorley |
| 141 | E07000119 | Fylde |
| 142 | E07000120 | Hyndburn |
| 143 | E07000121 | Lancaster |
| 144 | E07000122 | Pendle |
| 145 | E07000123 | Preston |
| 146 | E07000124 | Ribble Valley |
| 147 | E07000125 | Rossendale |
| 148 | E07000126 | South Ribble |
| 149 | E07000127 | West Lancashire |
| 150 | E07000128 | Wyre |
| 151 | E07000129 | Blaby |
| 152 | E07000130 | Charnwood |
| 153 | E07000131 | Harborough |
| 154 | E07000132 | Hinckley and Bosworth |
| 155 | E07000133 | Melton |
| 156 | E07000134 | North West Leicestershire |
| 157 | E07000135 | Oadby and Wigston |
| 158 | E07000136 | Boston |
| 159 | E07000137 | East Lindsey |
| 160 | E07000138 | Lincoln |
| 161 | E07000139 | North Kesteven |
| 162 | E07000140 | South Holland |
| 163 | E07000141 | South Kesteven |
| 164 | E07000142 | West Lindsey |
| 165 | E07000143 | Breckland |
| 166 | E07000144 | Broadland |
| 167 | E07000145 | Great Yarmouth |
| 168 | E07000146 | King's Lynn and West Norfolk |
| 169 | E07000147 | North Norfolk |
| 170 | E07000148 | Norwich |

Continued on next page

| | LTLA look up code | LTLA name |
|---|---|---|
| 171 | E07000149 | South Norfolk |
| 172 | E07000150 | Corby |
| 173 | E07000151 | Daventry |
| 174 | E07000152 | East Northamptonshire |
| 175 | E07000153 | Kettering |
| 176 | E07000154 | Northampton |
| 177 | E07000155 | South Northamptonshire |
| 178 | E07000156 | Wellingborough |
| 179 | E07000163 | Craven |
| 180 | E07000164 | Hambleton |
| 181 | E07000165 | Harrogate |
| 182 | E07000166 | Richmondshire |
| 183 | E07000167 | Ryedale |
| 184 | E07000168 | Scarborough |
| 185 | E07000169 | Selby |
| 186 | E07000170 | Ashfield |
| 187 | E07000171 | Bassetlaw |
| 188 | E07000172 | Broxtowe |
| 189 | E07000173 | Gedling |
| 190 | E07000174 | Mansfield |
| 191 | E07000175 | Newark and Sherwood |
| 192 | E07000176 | Rushcliffe |
| 193 | E07000177 | Cherwell |
| 194 | E07000178 | Oxford |
| 195 | E07000179 | South Oxfordshire |
| 196 | E07000180 | Vale of White Horse |
| 197 | E07000181 | West Oxfordshire |
| 198 | E07000187 | Mendip |
| 199 | E07000188 | Sedgemoor |
| 200 | E07000189 | South Somerset |
| 201 | E07000192 | Cannock Chase |
| 202 | E07000193 | East Staffordshire |
| 203 | E07000194 | Lichfield |
| 204 | E07000195 | Newcastle-under-Lyme |
| 205 | E07000196 | South Staffordshire |

Continued on next page

| | LTLA look up code | LTLA name |
|---|---|---|
| 206 | E07000197 | Stafford |
| 207 | E07000198 | Staffordshire Moorlands |
| 208 | E07000199 | Tamworth |
| 209 | E07000200 | Babergh |
| 210 | E07000202 | Ipswich |
| 211 | E07000203 | Mid Suffolk |
| 212 | E07000207 | Elmbridge |
| 213 | E07000208 | Epsom and Ewell |
| 214 | E07000209 | Guildford |
| 215 | E07000210 | Mole Valley |
| 216 | E07000211 | Reigate and Banstead |
| 217 | E07000212 | Runnymede |
| 218 | E07000213 | Spelthorne |
| 219 | E07000214 | Surrey Heath |
| 220 | E07000215 | Tandridge |
| 221 | E07000216 | Waverley |
| 222 | E07000217 | Woking |
| 223 | E07000218 | North Warwickshire |
| 224 | E07000219 | Nuneaton and Bedworth |
| 225 | E07000220 | Rugby |
| 226 | E07000221 | Stratford-on-Avon |
| 227 | E07000222 | Warwick |
| 228 | E07000223 | Adur |
| 229 | E07000224 | Arun |
| 230 | E07000225 | Chichester |
| 231 | E07000226 | Crawley |
| 232 | E07000227 | Horsham |
| 233 | E07000228 | Mid Sussex |
| 234 | E07000229 | Worthing |
| 235 | E07000234 | Bromsgrove |
| 236 | E07000235 | Malvern Hills |
| 237 | E07000236 | Redditch |
| 238 | E07000237 | Worcester |
| 239 | E07000238 | Wychavon |
| 240 | E07000239 | Wyre Forest |

Continued on next page

| | LTLA look up code | LTLA name |
|---|---|---|
| 241 | E07000240 | St Albans |
| 242 | E07000241 | Welwyn Hatfield |
| 243 | E07000242 | East Hertfordshire |
| 244 | E07000243 | Stevenage |
| 245 | E07000244 | East Suffolk |
| 246 | E07000245 | West Suffolk |
| 247 | E07000246 | Somerset West and Taunton |
| 248 | E08000001 | Bolton |
| 249 | E08000002 | Bury |
| 250 | E08000003 | Manchester |
| 251 | E08000004 | Oldham |
| 252 | E08000005 | Rochdale |
| 253 | E08000006 | Salford |
| 254 | E08000007 | Stockport |
| 255 | E08000008 | Tameside |
| 256 | E08000009 | Trafford |
| 257 | E08000010 | Wigan |
| 258 | E08000011 | Knowsley |
| 259 | E08000012 | Liverpool |
| 260 | E08000013 | St. Helens |
| 261 | E08000014 | Sefton |
| 262 | E08000015 | Wirral |
| 263 | E08000016 | Barnsley |
| 264 | E08000017 | Doncaster |
| 265 | E08000018 | Rotherham |
| 266 | E08000019 | Sheffield |
| 267 | E08000021 | Newcastle upon Tyne |
| 268 | E08000022 | North Tyneside |
| 269 | E08000023 | South Tyneside |
| 270 | E08000024 | Sunderland |
| 271 | E08000025 | Birmingham |
| 272 | E08000026 | Coventry |
| 273 | E08000027 | Dudley |
| 274 | E08000028 | Sandwell |
| 275 | E08000029 | Solihull |

Continued on next page

| | LTLA look up code | LTLA name |
|---|---|---|
| 276 | E08000030 | Walsall |
| 277 | E08000031 | Wolverhampton |
| 278 | E08000032 | Bradford |
| 279 | E08000033 | Calderdale |
| 280 | E08000034 | Kirklees |
| 281 | E08000035 | Leeds |
| 282 | E08000036 | Wakefield |
| 283 | E08000037 | Gateshead |
| 284 | E09000001 | City of London |
| 285 | E09000002 | Barking and Dagenham |
| 286 | E09000003 | Barnet |
| 287 | E09000004 | Bexley |
| 288 | E09000005 | Brent |
| 289 | E09000006 | Bromley |
| 290 | E09000007 | Camden |
| 291 | E09000008 | Croydon |
| 292 | E09000009 | Ealing |
| 293 | E09000010 | Enfield |
| 294 | E09000011 | Greenwich |
| 295 | E09000012 | Hackney |
| 296 | E09000013 | Hammersmith and Fulham |
| 297 | E09000014 | Haringey |
| 298 | E09000015 | Harrow |
| 299 | E09000016 | Havering |
| 300 | E09000017 | Hillingdon |
| 301 | E09000018 | Hounslow |
| 302 | E09000019 | Islington |
| 303 | E09000020 | Kensington and Chelsea |
| 304 | E09000021 | Kingston upon Thames |
| 305 | E09000022 | Lambeth |
| 306 | E09000023 | Lewisham |
| 307 | E09000024 | Merton |
| 308 | E09000025 | Newham |
| 309 | E09000026 | Redbridge |
| 310 | E09000027 | Richmond upon Thames |

Continued on next page

| | LTLA look up code | LTLA name |
|---|---|---|
| 311 | E09000028 | Southwark |
| 312 | E09000029 | Sutton |
| 313 | E09000030 | Tower Hamlets |
| 314 | E09000031 | Waltham Forest |
| 315 | E09000032 | Wandsworth |
| 316 | E09000033 | Westminster |
